# Sex-specific skeletal aging trajectories and reference centiles across vertebrae and ribs in over 78,000 patients

**DOI:** 10.64898/2026.10.30.26363202

**Authors:** Yosuke Yamagishi, Tomohiro Kikuchi, Kohei Yamamoto, Shouhei Hanaoka, Takeharu Yoshikawa, Osamu Abe

**Author notes:** Corresponding author: Tomohiro Kikuchi, Data Science Center, Jichi Medical University, 3311-1 Yakushi-ji, Shimotsuke, Tochigi 329-0498, Japan.

## Abstract

Skeletal aging is usually assessed at one site, so how bone loss differs across the skeleton and between sexes is unclear. We analyzed non-contrast torso CT from 2024 in 78,620 patients aged 15–100 years (41,056 men, 37,564 women) at nine Japanese hospitals, measuring mean attenuation of 25 vertebrae and 24 ribs from TotalSegmentator masks. We fitted sex-specific mean curves and quantile-regression centiles, and assessed each site as an L1 surrogate by age-adjusted correlation, errors-in-variables conversion and discrimination of low-for-age L1.

Attenuation fell with age at all 49 sites in women and 46 of 49 in men, with a sex-dependent shape everywhere (all FDR-adjusted *P* < 0.0001). Female decline was steepest at a median of 52.0 years. L2 reproduced L1 closely (limits of agreement ±45–46 HU); ribs did not (±68 to ±82 HU). These centiles place an automated measurement at any of 49 sites against age- and sex-matched peers.

## Introduction

Age-related bone loss underlies a growing burden of fragility fracture in aging populations.^1–3^ Yet how the skeleton ages is known mainly from a few sites: dual-energy X-ray absorptiometry, the diagnostic reference standard, is a two-dimensional projection, is not universally available, and is performed only when osteoporosis is already suspected.^4,5^ Whether bones that differ in size, composition and mechanical loading lose density at the same ages, and how that timing differs between women and men, has therefore been difficult to examine at population scale. Routine CT offers a way to do so. Every non-contrast CT of the torso contains three-dimensional information about every bone in the field of view, acquired for an unrelated indication and then left unused; attenuation in Hounsfield units measured on such scans predicts low bone mineral density and incident vertebral fracture, and opportunistic screening built on this observation is now an active area of radiological practice,^6–9^ adding no radiation, no scanner time and no cost.

Much of the established clinical literature rests on a single vertebra, usually L1, and on normative series predominantly of European ancestry,^8,9^ although recent automated work has extended trabecular measurements across the thoracic and lumbar spine in a large, diverse population.^10^ Two consequences follow. First, when L1 is unavailable — because of instrumentation, a compression fracture, cement augmentation, or exclusion from the field of view — other levels can be measured, but there is no well-established basis for treating them as interchangeable with L1. Studies addressing this report correlations between sites,^11^ but a correlation between two variables that both decline steeply with age is inflated by that shared trend, and a high correlation is compatible with poor agreement on the Hounsfield scale. Second, reference data are usually reported as means with standard deviations, whereas interpreting an individual’s result requires knowing where it falls among age- and sex-matched peers — that is, centiles.

Deep-learning segmentation has removed the practical obstacle: one model delineates every vertebra and rib without operator input,^12^ so the measurement is no longer confined to wherever a reader places a region of interest, and national imaging repositories supply the numbers needed to build a reference.^13^ Whole-body charts of this kind now exist for European and North American cohorts.^14,15^ We therefore used routine CT from a large Japanese cohort to describe skeletal aging across 25 vertebrae and 24 ribs: to construct age- and sex-specific reference centiles for each site, to characterize when and how steeply attenuation declines in women and in men, and to establish whether that timing is shared across the skeleton. As practical corollaries, we determined — using agreement rather than correlation alone — which sites can stand in for L1, and quantified how far the same measurement drifts between hospitals.

## Results

### Study cohort

J-MID held 239,766 non-contrast CT examinations from 135,425 patients during 2024. Of these, 123,507 covered the torso, and retaining each patient’s first such examination left 79,482; a further 862 were excluded for missing or out-of-range age or unrecognized sex (Supplementary Fig. 1). The cohort comprised 78,620 patients: 41,056 men (52.2%) and 37,564 women (47.8%), mean age 65.3 years (SD 15.8). Body-mass index was available for 51,046 (64.9%), and centers contributed 281 to 20,296 patients each (Table 1). Of 3,288,175 segmentations, 3,069,679 (93.4%) passed quality control, leaving 26,288 (C2) to 74,681 (right rib 11) analysable patients per site, median 70,264 (Supplementary Table 1).

**Table 1.** Characteristics of the study cohort.

| Characteristic | All | Men | Women |
| --- | --- | --- | --- |
| Patients, n | 78,620 | 41,056 | 37,564 |
| Age, years, mean (SD) | 65.3 (15.8) | 66.5 (15.2) | 63.9 (16.2) |
| Age, years, median (IQR) | 68 (55–77) | 70 (58–77) | 66 (53–76) |
| Age, years, range | 15–100 | 15–100 | 15–100 |
| Body-mass index, kg/m<sup>2</sup>, mean (SD) | 23.1 (4.3) | 23.4 (3.9) | 22.6 (4.6) |
| Body-mass index available, n (%) | 51,046 (64.9) | 27,093 (66.0) | 23,953 (63.8) |
| Sex, n (%) |  |  |  |
| Men | 41,056 (52.2) | — | — |
| Women | 37,564 (47.8) | — | — |
| Age group, n (%) |  |  |  |
| 15–19 | 599 (0.8) | 311 (0.8) | 288 (0.8) |
| 20–29 | 1,963 (2.5) | 940 (2.3) | 1,023 (2.7) |
| 30–39 | 3,159 (4.0) | 1,397 (3.4) | 1,762 (4.7) |
| 40–49 | 6,885 (8.8) | 2,859 (7.0) | 4,026 (10.7) |
| 50–59 | 12,688 (16.1) | 5,727 (13.9) | 6,961 (18.5) |
| 60–69 | 15,998 (20.3) | 8,683 (21.1) | 7,315 (19.5) |
| 70–79 | 23,241 (29.6) | 13,508 (32.9) | 9,733 (25.9) |
| 80–89 | 12,574 (16.0) | 6,961 (17.0) | 5,613 (14.9) |
| 90–100 | 1,513 (1.9) | 670 (1.6) | 843 (2.2) |
| Contributing centers, n | 9 | — | — |
| Patients per center, median (range) | 7,619 (281–20,296) | — | — |
| Analysable patients per site, median (range) | 70,264 (26,288–74,681) | 36,632 (12,939–38,989) | 33,600 (13,349–35,692) |

Data are n (%) unless stated. One examination per patient identifier was retained; because a J-MID identifier denotes an individual, each row of the cohort is a distinct patient. Contributing centers are reported under pseudonyms ordered by decreasing number of patients.

### Age trajectories across 49 skeletal sites

Attenuation decreased with age at all 49 sites in women and at 46 of 49 in men (Fig. 1); at C1, C4 and C6 in men the fitted 40–80-year change was slightly positive (+1 to +3 HU), with a CI including zero. At age 50 years, fitted attenuation ranged from 231 to 446 HU in men and from 247 to 469 HU in women, highest in the cervical spine and lowest in the thoracolumbar spine, and within the rib cage rose from rib 3 to rib 11 (Supplementary Figs. 3–5, Supplementary Table 2).

**Fig. 1.**
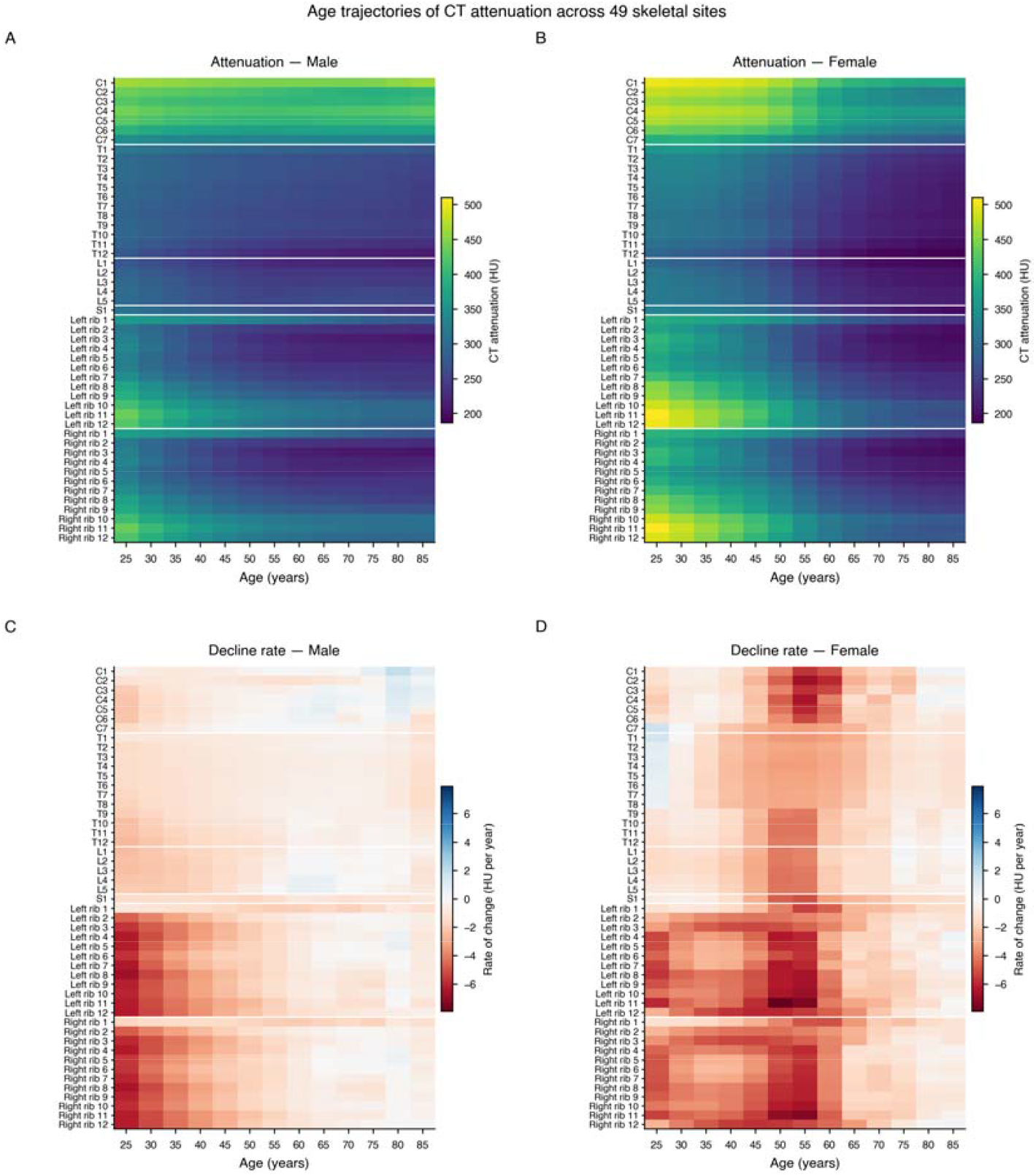
Age trajectories of CT attenuation across 49 skeletal sites. Fitted mean attenuation (**a**, men; **b**, women) and its rate of change (**c**, men; **d**, women) for each of 25 vertebrae and 24 ribs, evaluated at 5-year intervals from 25 to 85 years. Rows are ordered anatomically and separated by white lines into cervical, thoracic and lumbar vertebrae, sacrum, left ribs and right ribs. In **c** and **d** red indicates loss of attenuation and blue indicates gain; the band of rapid loss confined to ages 45–60 years is present in women at every site and absent in men. Rates of change are cross-sectional slopes with age and do not represent within-person change over time.

### Sex differences in trajectory shape and rate of decline

The shape of the age trajectory differed between the sexes at all 49 sites (joint Wald test, all FDR-adjusted *P* < 0.0001) (Supplementary Table 2B). Women had higher attenuation at younger ages and lower at older ages: the female-minus-male difference had a median of +49 HU at 30 years (range +24 to +90) and −25 HU at 70 years (−56 to +1), and the curves crossed at every site, at a median age of 57 years (IQR 55–64).

Decline rates diverged accordingly (Fig. 2, Table 2). Between 40 and 50 years the median rate across sites was −1.54 HU per year in men and −3.00 in women; between 50 and 60 years it was −0.98 in men and −4.81 in women. In women, decline accelerated to an interior maximum at all 49 sites, at a median age of 52.0 years (IQR 51.8–52.5; range across sites 50.5–55.5), whereas in men 46 of 49 sites showed no interior maximum, decline being steepest at the lower limit of the search window. Averaged over 45–60 years women declined faster at all 49 sites, by a median of 3.32 HU per year (range 2.34–4.87; all FDR-adjusted *P* < 0.0001).

**Fig. 2.**
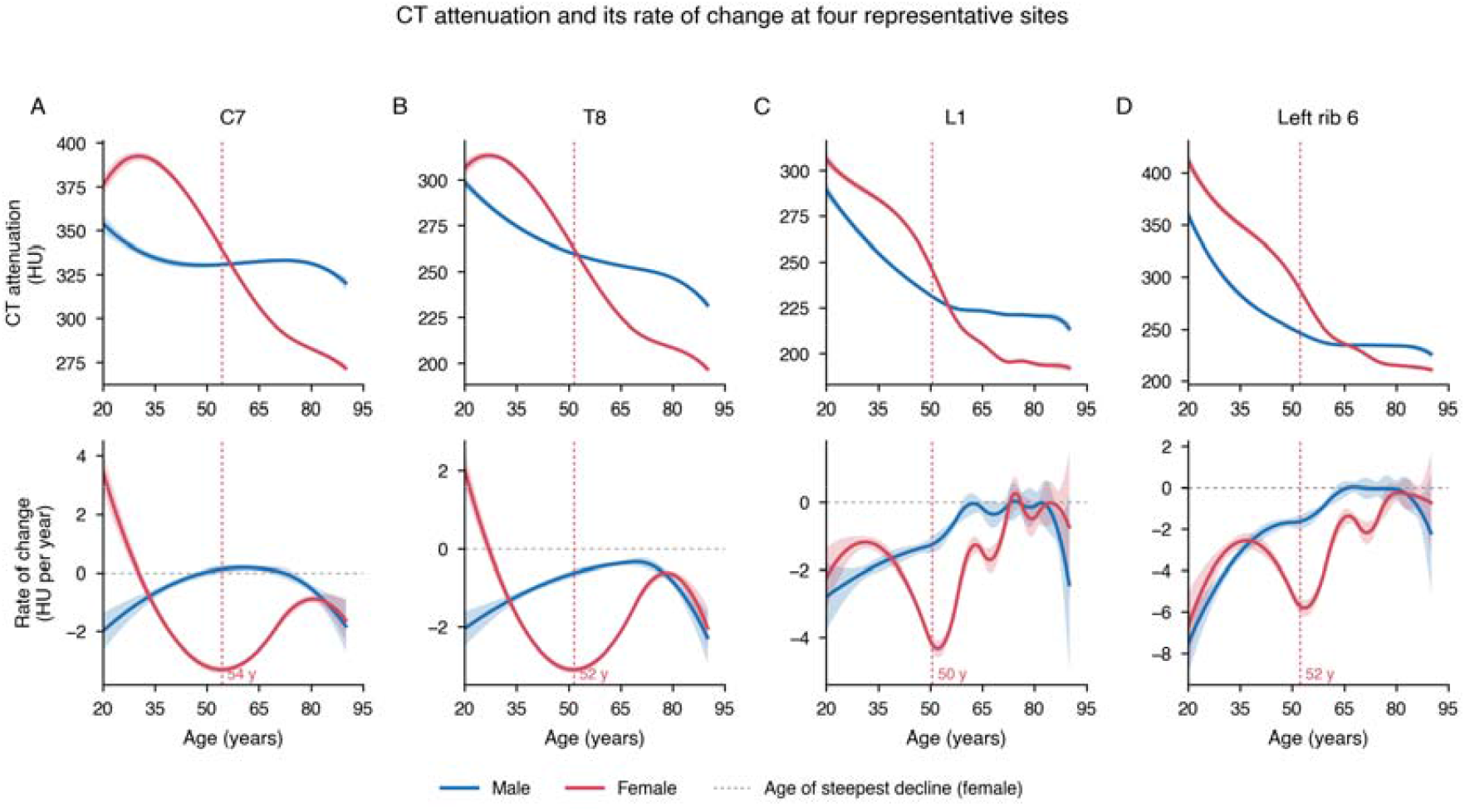
CT attenuation and its rate of change at four representative sites. Upper row, fitted mean attenuation with 95% CI; lower row, the rate of change with 95% CI, obtained analytically from the same fitted spline. Blue, men; red, women. The vertical dashed line marks the age of steepest decline in women where an interior maximum exists. **a** C7, **b** T8, **c** L1, **d** left rib 6. The rate of change is the analytic derivative of the fitted cross-sectional curve and is not a longitudinal within-person change.

**Table 2.**
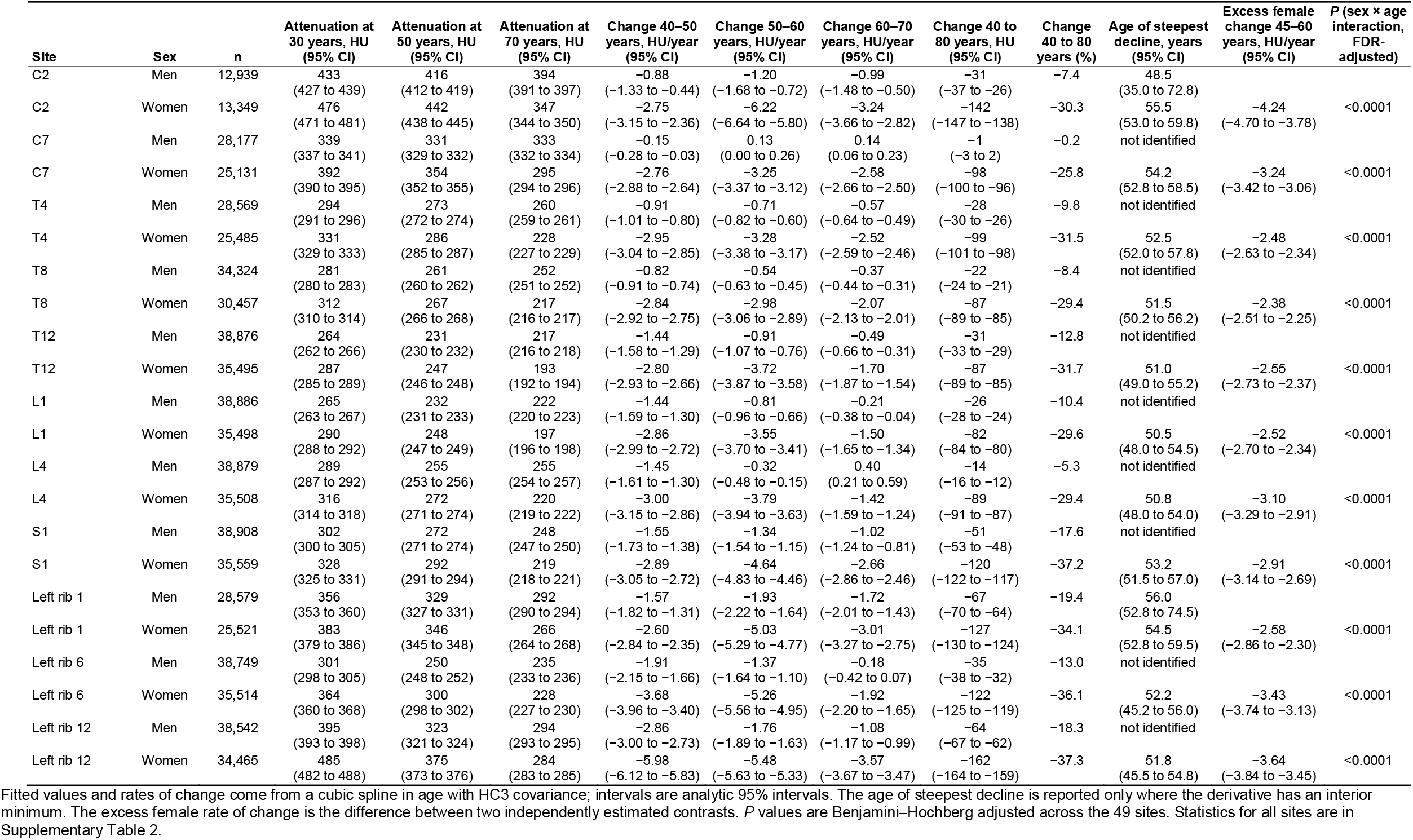
CT attenuation and its rate of change at representative skeletal sites.

### Reference centiles and prevalence of low attenuation

Centile curves for all 49 sites and both sexes are in the Supplementary Figs. 9–12 and Supplementary Tables 3 and 3B; no crossings required repair. For L1 in women the P3–P97 interval was 217–383 HU at 30 years, 172–332 HU at 50 years and 138– 284 HU at 70 years; in men it was 191–341, 163–311 and 145–325 HU (Fig. 3).

**Fig. 3.**
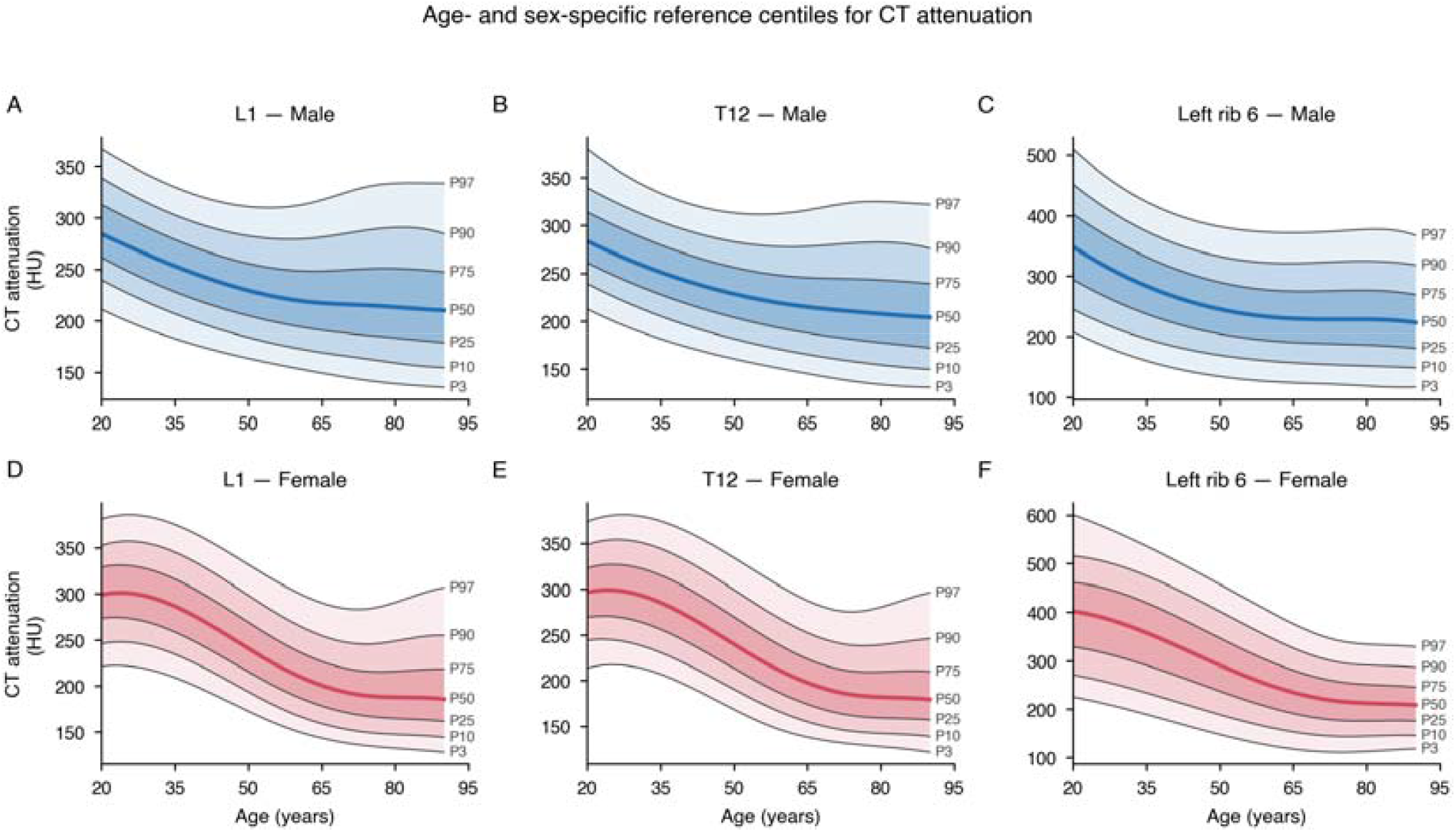
Age- and sex-specific reference centiles for CT attenuation. Reference centiles (P3, P10, P25, P50, P75, P90, P97) estimated by quantile regression on a cubic spline in age, for L1 (**a, d**), T12 (**b, e**) and left rib 6 (**c, f**) in men (upper row) and women (lower row). The heavy line is the median; shading marks the P25–P75, P10– P90 and P3–P97 intervals. Curves for all 49 sites are in Supplementary Figs. 9–12.

Only 22 of 74,384 analysable L1 measurements (0.03%) fell below 100 HU and 3681 (4.9%) below 150 HU, the thresholds used with trabecular region-of-interest measurements (Supplementary Table 4). Using a young-adult-referenced score below −2.5 instead, prevalence rose from 0.6% in men and 0.4% in women aged 20– 29 years to 15.1% and 50.8% at 80–89 years (Supplementary Table 6).

### Other skeletal sites as surrogates for L1

Correlation with L1 attenuation decreased with anatomical distance (Fig. 4a). Among vertebrae and the sacrum, unadjusted correlations were 0.63–0.88 in men and 0.71– 0.90 in women, and after adjustment for age 0.64–0.88 and 0.60–0.85. Among ribs they fell from 0.54–0.65 and 0.63–0.74 to 0.53–0.63 and 0.45–0.59 after adjustment (all FDR-adjusted *P* < 0.0001).

**Fig. 4.**
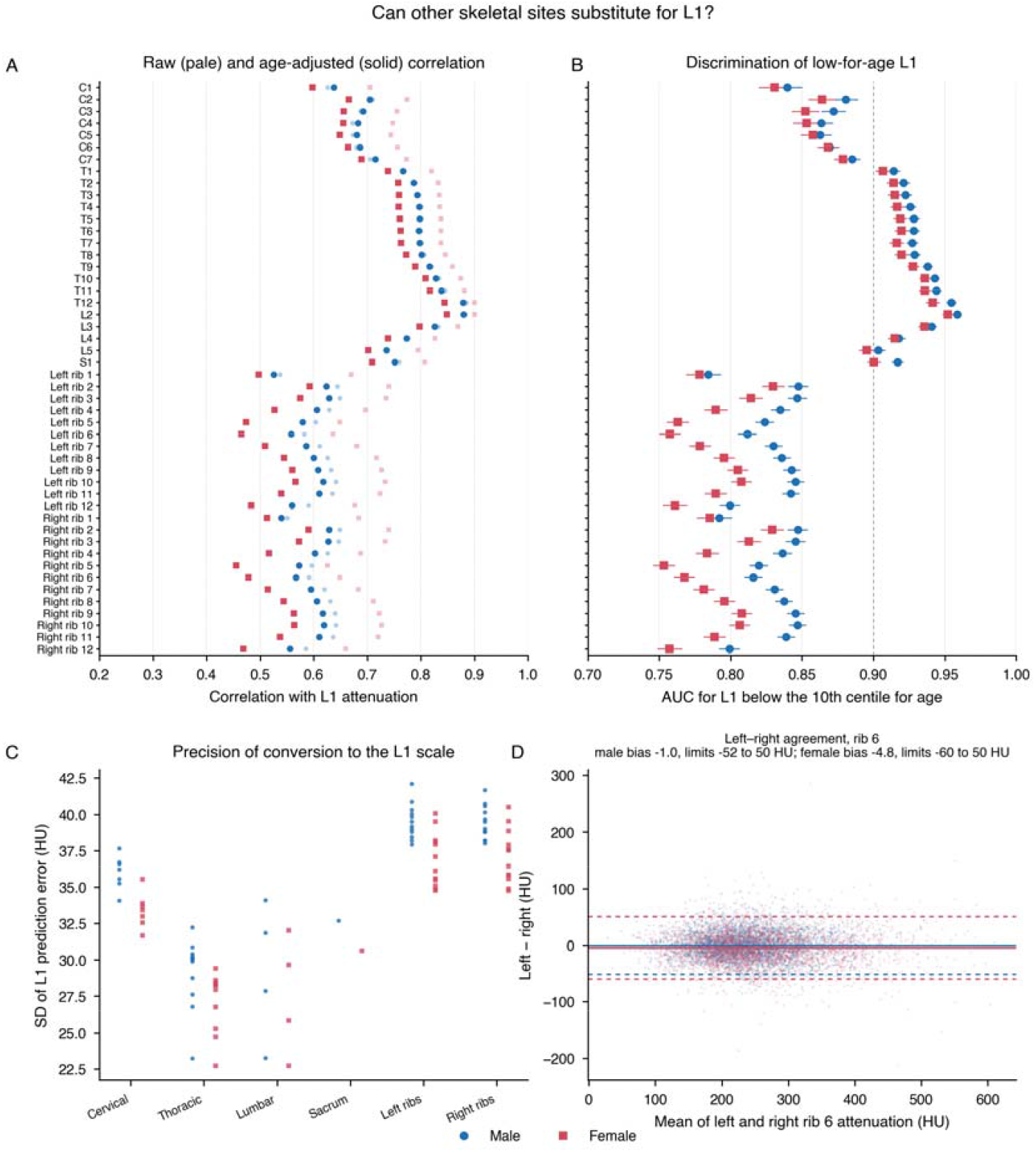
Can other skeletal sites substitute for L1? a. Correlation of each site’s attenuation with L1: pale symbols, unadjusted Pearson correlation; solid symbols, correlation after removing a spline in age from both variables. **b** Area under the ROC curve, with 95% CI, for identifying an L1 value below the 10th reference centile for age and sex, using each site’s own age- and sex-standardized score; the dashed line marks an AUC of 0.90. **c** SD of the error when a site is converted to the L1 scale by Deming regression, grouped anatomically. **d** Bland–Altman plot of left minus right rib 6 attenuation (random sample of 8000 patients shown); bias (solid) and 95% limits of agreement (dashed) are drawn separately for each sex in the corresponding color, matching Supplementary Table 13. Circles, men; squares, women.

Discrimination of an L1 value below the 10th centile for age and sex (by construction 10.0% of analysable patients: 3888 men, 3574 women) followed the same ordering (Fig. 4b, Table 3). AUC was 0.959 (95% CI 0.956–0.962) for L2 in men and 0.952 (0.948–0.956) in women; 17 of 48 sites in men and 16 of 48 in women reached at least 0.90, all vertebrae. For ribs AUC was 0.78–0.85 in men and 0.75–0.83 in women, and for cervical vertebrae 0.84–0.89 and 0.83–0.88.

**Table 3.** Performance of other skeletal sites as surrogates for L1.

| Site | Sex | n | Correlation with L1, unadjusted (95% CI) | Correlation with L1, age-adjusted (95% CI) | Conversion to L1 (Deming) | SD of L1 prediction error, HU | 95% limits of agreement, HU | AUC for L1 below the 10th centile for age and sex (95% CI) | P (age-adjusted correlation, FDR-adjusted) |
| --- | --- | --- | --- | --- | --- | --- | --- | --- | --- |
| L2 | Men | 38,069 | 0.88 (0.88 to 0.89) | 0.88 (0.88 to 0.88) | $L1 = -1.8 + 0.94 \times \text{site}$ | 23.3 | -46 to 46 | 0.959 (0.956 to 0.962) | <0.0001 |
| T12 | Men | 38,052 | 0.88 (0.88 to 0.89) | 0.88 (0.88 to 0.88) | $L1 = 4.0 + 1.00 \times \text{site}$ | 23.2 | -46 to 46 | 0.955 (0.951 to 0.958) | <0.0001 |
| T11 | Men | 37,844 | 0.85 (0.84 to 0.85) | 0.84 (0.83 to 0.84) | $L1 = 3.9 + 0.93 \times \text{site}$ | 26.8 | -52 to 52 | 0.944 (0.941 to 0.948) | <0.0001 |
| T10 | Men | 37,231 | 0.83 (0.83 to 0.84) | 0.83 (0.82 to 0.83) | $L1 = 0.4 + 0.89 \times \text{site}$ | 27.6 | -54 to 54 | 0.943 (0.939 to 0.946) | <0.0001 |
| L3 | Men | 37,809 | 0.83 (0.83 to 0.84) | 0.83 (0.82 to 0.83) | $L1 = -2.1 + 0.92 \times \text{site}$ | 27.9 | -55 to 55 | 0.941 (0.937 to 0.944) | <0.0001 |
| T9 | Men | 35,315 | 0.82 (0.82 to 0.82) | 0.82 (0.81 to 0.82) | $L1 = -0.2 + 0.88 \times \text{site}$ | 28.8 | -56 to 56 | 0.938 (0.934 to 0.942) | <0.0001 |
| Right rib 2 | Men | 27,479 | 0.65 (0.64 to 0.66) | 0.63 (0.62 to 0.64) | $L1 = 45.3 + 0.76 \times \text{site}$ | 38.8 | -76 to 76 | 0.847 (0.840 to 0.854) | <0.0001 |
| Left rib 3 | Men | 27,937 | 0.65 (0.64 to 0.66) | 0.63 (0.62 to 0.64) | $L1 = 60.9 + 0.75 \times \text{site}$ | 38.8 | -76 to 76 | 0.847 (0.840 to 0.853) | <0.0001 |
| Left rib 1 | Men | 27,448 | 0.54 (0.53 to 0.55) | 0.53 (0.52 to 0.53) | $L1 = 60.4 + 0.55 \times \text{site}$ | 42.1 | -82 to 82 | 0.784 (0.776 to 0.793) | <0.0001 |
| L2 | Women | 34,673 | 0.90 (0.90 to 0.90) | 0.85 (0.85 to 0.85) | $L1 = -3.8 + 0.96 \times \text{site}$ | 22.7 | -45 to 45 | 0.952 (0.948 to 0.956) | <0.0001 |
| T12 | Women | 34,530 | 0.90 (0.90 to 0.90) | 0.84 (0.84 to 0.85) | $L1 = 8.8 + 0.98 \times \text{site}$ | 22.7 | -45 to 45 | 0.941 (0.937 to 0.946) | <0.0001 |
| T11 | Women | 34,403 | 0.88 (0.88 to 0.88) | 0.82 (0.81 to 0.82) | $L1 = 7.5 + 0.92 \times \text{site}$ | 24.7 | -48 to 48 | 0.936 (0.932 to 0.940) | <0.0001 |
| T10 | Women | 33,715 | 0.87 (0.87 to 0.88) | 0.81 (0.81 to 0.81) | $L1 = 1.9 + 0.91 \times \text{site}$ | 25.3 | -50 to 50 | 0.936 (0.932 to 0.940) | <0.0001 |
| L3 | Women | 34,386 | 0.87 (0.87 to 0.87) | 0.80 (0.79 to 0.80) | $L1 = -1.5 + 0.94 \times \text{site}$ | 25.9 | -51 to 51 | 0.936 (0.931 to 0.940) | <0.0001 |
| T9 | Women | 31,583 | 0.86 (0.86 to 0.86) | 0.79 (0.79 to 0.79) | $L1 = -0.5 + 0.91 \times \text{site}$ | 26.8 | -52 to 52 | 0.928 (0.923 to 0.932) | <0.0001 |
| Left rib 2 | Women | 24,393 | 0.74 (0.73 to 0.75) | 0.59 (0.58 to 0.60) | $L1 = 53.0 + 0.68 \times \text{site}$ | 34.8 | -68 to 68 | 0.830 (0.822 to 0.838) | <0.0001 |
| Right rib 2 | Women | 24,438 | 0.74 (0.73 to 0.75) | 0.59 (0.58 to 0.60) | $L1 = 54.9 + 0.67 \times \text{site}$ | 34.8 | -68 to 68 | 0.829 (0.821 to 0.837) | <0.0001 |
| Right rib 5 | Women | 31,756 | 0.63 (0.62 to 0.63) | 0.45 (0.45 to 0.46) | $L1 = 99.4 + 0.48 \times \text{site}$ | 40.5 | -79 to 79 | 0.753 (0.745 to 0.761) | <0.0001 |

Conversion to the L1 scale was near-identity for the immediately adjacent levels (in men, L2: L1 = −1.8 + 0.94 × L2; T12: L1 = 4.0 + 1.00 × T12), with a prediction error of 23 HU and limits of agreement of ±45 to ±46 HU (Fig. 4c, Table 3). The prediction error was 25–34 HU for the remaining thoracolumbar levels and the sacrum, 32–38 HU for cervical vertebrae and 35–42 HU for ribs, the last corresponding to limits of ±68 to ±82 HU.

### Left–right rib agreement

Left and right ribs of the same level were strongly associated (ICC(A,1) 0.77–0.96) with a small systematic difference (bias −7.4 to +3.2 HU), but the 95% limits of agreement were narrowest at ribs 2–3 (±35 to ±40 HU) and widened to ±109 HU at rib 12 (Fig. 4d; Supplementary Fig. 15, Supplementary Table 13).

### Between-center variation

Attenuation differed systematically between the nine centers after adjustment for age and sex (Supplementary Fig. 17, Supplementary Tables 8 and 14). The range of adjusted center deviations had a median of 21.6 HU across the 49 sites and depended strongly on the site: smallest in the lumbar (median 14.8 HU) and thoracic spine (15.8 HU) and largest in the cervical spine (35.8 HU) and the ribs (27.7 HU left, 31.2 HU right), with L1 showing the smallest between-center spread of all 49 sites (13.3 HU). Sites with a wide between-center range were also the poorer surrogates for L1: across sites the range correlated with the SD of the L1 conversion error (Spearman ρ +0.55 in men, +0.65 in women) and inversely with the age-adjusted correlation with L1 and with discrimination of a low-for-age L1 (ρ −0.62 to −0.64; all *P* < 0.0001).

### Sensitivity analyses

The stricter quality-control rule changed fitted attenuation at 50 years by a median of 3.8 HU (maximum 12.0) and the 45–60-year rate of change by at most 0.79 HU per year, and omitting each center in turn changed them by at most 6.2 HU and 0.77 HU per year. Attenuation rose with body-mass index (median 1.22 HU per kg/m^2^), but adjustment changed the rate of change by at most 0.23 HU per year, and the association between attenuation and segmented volume after quality control was near zero (median age-adjusted r −0.01), though it reached 0.38 at right rib 5 in women (Supplementary Fig. 16, Supplementary Tables 7–11).

## Discussion

In 78,620 patients undergoing routine CT we described skeletal aging at 49 individually resolved sites, constructing age- and sex-specific reference centiles for CT attenuation, and quantified how far each site can substitute for L1. Four results have practical consequences.

The first is the crossover between the sexes. Women had higher attenuation than men until the sixth decade and lower thereafter, the curves crossing at a median age of 57 years. Sex-pooled reference values, and comparisons between cohorts of differing age structure, will therefore mislead: averaging over a curve rather than at a fixed age conceals the crossover entirely. The finding is consistent with previous CT studies showing higher trabecular density in younger women and accelerated subsequent loss,^9,16^ and means a woman of 45 and a woman of 65 with the same attenuation occupy very different positions relative to their peers.

The second is the timing and generality of accelerated loss in women. Decline reached an interior maximum at all 49 sites, within a narrow range of 50.5–55.5 years, despite substantial differences among sites in size, cortical-to-trabecular ratio and mechanical loading. The median age of natural menopause in Japanese women is 52.1 years,^17^ and this concordance is consistent with a systemic hormonal contribution around the menopausal transition acting near-simultaneously throughout the axial skeleton, although menopausal status was not recorded. In men, 46 of 49 sites showed no interior maximum: decline was greatest at the youngest ages examined and attenuated thereafter. That apparent deceleration may partly reflect cross-sectional cohort or survival effects and degenerative change, but the sex difference is unlikely to be a modeling artifact because both sexes were fitted with identical knots and flexibility.

The third concerns surrogacy, and it revises a claim previously made on the basis of correlation alone. Adjacent thoracolumbar vertebrae are the closest substitutes for L1, convertible to its scale with a stated precision: the conversion is near-identity, the prediction error about 23 HU, the limits of agreement about ±45 HU, and discrimination of a low-for-age L1 reaches an AUC of 0.94–0.96. Ribs are not: their correlation falls further after age adjustment, showing that much of the raw association reflects the shared age trend, and conversion carries limits approaching ±80 HU, spanning most of the clinically relevant range. The same applies within a rib pair, whose limits reach ±109 HU at rib 12 despite a high intraclass correlation. Correlation quantifies ranking, agreement quantifies substitutability,^18^ and only the latter licenses replacing one measurement by another.

Recent work by Westerhoff and colleagues established scanner- and protocol-adjusted trabecular attenuation values across the thoracic and lumbar spine in more than 280,000 patients,^10^ and Wachinger and colleagues modeled volume and attenuation across 106 anatomical structures in more than 350,000 European and North American examinations.^14,15^ Our study complements both: it extends coverage to the cervical spine and to individual ribs, uses whole-bone rather than trabecular attenuation, resolves every level and rib separately rather than merging them, and directly assesses interchangeability with L1 — a question neither design addresses. Restricting acquisition to non-contrast CT within one calendar year across nine independent Japanese academic centers also reduces the technical heterogeneity inherent to very large retrospective datasets. The difference in measurement definition matters clinically: our low prevalence below 100 and 150 HU is not evidence of a healthy population but a consequence of averaging over an entire vertebral mask including the cortical shell, whereas published thresholds derive from a trabecular region of interest.^8,9,19^ Transporting cut-offs between measurement definitions, scanners or populations without recalibration will misclassify;^10,20^ centiles conditional on age and sex, and the young-adult-referenced score, are internal to the measurement system and avoid this.^21^

A fourth result concerns where these measurements can be transported. Nine independent centers imaging within one year let the center term be estimated cleanly, and it is not negligible: for L1 the adjusted spread between centers was 13.3 HU, of the order of nine years of male aging at the rate we measured between 40 and 50 years, or a quarter of the interquartile range of L1 attenuation in women at 50 years — enough to displace an individual by a centile band. The pattern is informative rather than merely cautionary: the thoracolumbar spine varied least and L1 least of all, whereas the thin, cortical-rich, obliquely oriented cervical vertebrae and ribs — those most exposed to differences in reconstruction and partial-volume averaging — varied most, and across sites center sensitivity tracked poor substitutability for L1. That L1 also varied least of all 49 sites supports its robustness as a reference site; because L1 was the reference target throughout, we cannot claim it represents the rest of the skeleton better than other sites.

The main strengths are scale, breadth and reproducibility: 49 sites measured by one published segmentation model, restriction to non-contrast examinations within a single calendar year, nine centers, and a pipeline that regenerates every reported number from the raw export in one command. Derivatives and their intervals are exact, both sexes receive identical smoothing, and multiplicity across 49 sites is controlled.

Several limitations remain. The design is cross-sectional, so the curves describe differences between people of different ages, not within-person change. The cohort is one of patients imaged for a clinical indication rather than of healthy volunteers, so the outer centiles are empirical reference limits for routine practice rather than thresholds of normality, and it is skewed toward older adults, with fewer than 3000 patients below 30 years, so the ascending limb and the age of peak attenuation are poorly determined and we do not report them. J-MID supplies no diagnoses, medications, menopausal status or fracture outcomes, so the curves are unadjusted for determinants of bone metabolism and attenuation could not be related to fracture. Tube voltage and reconstruction were not recorded per examination, so their effects are absorbed into the center term.^20^ Segmentation was not manually reviewed, the cohort is Japanese, race and ethnicity were not recorded in J-MID, and administrative sex was the only classification available.

In conclusion, we provide reference centiles for CT attenuation at 49 individually resolved skeletal sites in a Japanese patient population, with the conversion equations, agreement limits and between-center variation needed to use them. The sexes differ not only in rate but in ordering, crossing in the sixth decade, with a sharply localized acceleration in women at about 52 years. Sites adjacent to L1 may stand in for it with a stated precision; ribs, cervical vertebrae and the contralateral rib may not.

## Methods

### Study design and participants

This was a retrospective, multicenter, cross-sectional study of the Japan Medical Image Database (J-MID), which aggregates anonymized routine clinical CT from nine Japanese academic hospitals under an opt-out consent model.^13^ We included non-contrast torso CT acquired between Jan 1 and Dec 31, 2024 in which the field of view covered the abdomen and pelvis; for patients with more than one eligible examination the first was retained. A single calendar year limits confounding by secular change in scanner hardware, reconstruction and body composition. The scan range was not recorded per examination and varied, so the number of analysable patients differs between sites and is lowest in the cervical spine. Eligible patients were aged 15–100 years with recorded sex, taken from the DICOM header, which records administrative sex rather than self-reported gender. A J-MID patient identifier denotes an individual, confirmed with the data custodian, so one examination per identifier yields one independent observation per patient.

### Ethics approval and consent

The study was approved by the Institutional Review Board of Jichi Medical University (approval number J24-017), which waived written informed consent because the analysis was retrospective and used anonymized data.

### Measurements

Segmentation masks for 25 vertebrae (C1–C7, T1–T12, L1–L5, S1) and 24 ribs (left and right, 1–12) were generated with TotalSegmentator v2.10.0.^12^ For each structure we recorded the mean attenuation over all voxels inside its mask and the mask volume. This whole-mask definition includes cortical as well as trabecular bone and is not numerically interchangeable with the trabecular region-of-interest measurements on which published L1 cut-offs are based. Height and weight were taken from the DICOM header where present, to derive body-mass index.

### Quality control

Structures that TotalSegmentator did not delineate are exported with zero attenuation and zero volume and were treated as missing. Three further rules were applied per structure and are reported separately (Supplementary Table 1). First, a vertebra was analysable only if its immediate within-region neighbors were also segmented, which guards against levels clipped by the field of view and against off-by-one labeling.

Second, segmentations whose volume fell outside the 5th to 99.9th percentile of the site- and sex-specific volume distribution were excluded, because partially covered structures have a small volume and a strongly biased mean attenuation. Third, attenuation more than five scaled median absolute deviations from the median of its site, sex and 5-year age cell was excluded as artifact. The third rule is deliberately mild because the centiles are estimated from these data and the physiological spread must be preserved; a stricter rule trimming the 5th–95th percentile within each 1-year age stratum discards about a fifth of observations and was retained only as a sensitivity analysis.

### Statistical analysis

Analyses were done separately by sex. For each site we fitted the mean trajectory as a cubic B-spline in age by ordinary least squares with heteroscedasticity-consistent (HC3) standard errors, with knots at quantiles of that site’s pooled age distribution and the number of basis functions (5–10) chosen by minimizing the summed Bayesian information criterion over both sexes, so that male and female curves for a site have identical knots and identical flexibility. Because the model is linear in the basis, the fitted value, the derivative, the average rate of change over an age window and the total change between two ages are exact linear functionals of the coefficient vector, each with a confidence interval from the coefficient covariance.^22^ The age at which decline is steepest was searched between 35 and 80 years and counted as an interior maximum only if it lay more than 1 year inside that range; its intervals were obtained by pooling coefficient draws across candidate spline dimensions, so that they reflect uncertainty about the smoother as well as the coefficients. Curves were evaluated between 20 and 90 years. Reference centiles (P3, P10, P25, P50, P75, P90, P97) were estimated by quantile regression on the same basis;^23–25^ crossings were repaired by a running maximum. Full specifications are in the Supplementary Information and in the code.

Whether the trajectory shape differed by sex was tested at each site by a joint Wald test of the sex-by-spline interaction in a pooled model allowing a separate level for each sex. The female-minus-male difference in the average rate of change between 45 and 60 years was estimated as the difference of two independent contrasts.

For each site other than L1 we report the Pearson correlation with L1 attenuation and the partial correlation after removing a spline in age from both variables; an errors-in-variables (Deming) equation converting the site to the L1 scale, fitted with an assumed error-variance ratio of one,^26^ with the SD of the prediction error and 95% limits of agreement; and the area under the receiver-operating-characteristic curve, with a DeLong interval,^27^ for identifying patients whose L1 attenuation lay below the 10th reference centile for their age and sex, using the site’s own age- and sex-standardized score. Left–right rib agreement was assessed with ICC(A,1)^28^ and Bland–Altman limits^18^ rather than correlation, because interchangeability requires agreement. The Benjamini–Hochberg procedure controlled the false discovery rate at 5% within each family of tests.^29^ Because published absolute L1 cut-offs are not transferable to whole-mask attenuation, we also defined a young-adult-referenced score (patients aged 20–34 of the same sex), mirroring the T-score construction,^30^ and report the prevalence below −2.5.

### Outcomes, study size and missing data

The main outcome was the age- and sex-specific distribution of mean attenuation at each of the 49 sites, summarized as a fitted mean curve and as reference centiles; secondary outcomes were its rate of change with age and the agreement of each site with L1. No formal sample-size calculation was performed: all eligible examinations in the period were analyzed. Analyses were complete-case within each site, so a patient contributes to a site only when that site passed quality control, and the number analyzed per site is given in Supplementary Table 1. Body-mass index, available for 64.9% of patients, entered only a sensitivity analysis restricted to those patients; no values were imputed.

### Sensitivity analyses and software

Sensitivity analyses examined the quality-control rule, between-center variation, adjustment for body-mass index, and the dependence of attenuation on segmented volume. Analyses used Python 3.13; the pipeline runs from the raw export to every figure and table in one command and records package versions and all parameters in a run manifest.

### Use of large language models

During the preparation of this work the authors used ChatGPT (OpenAI; GPT-5.6 Sol) and Claude (Anthropic; Claude 5 Opus) to assist with language editing, restructuring and refinement of the manuscript text. The tools were not used to generate scientific content, to analyze or interpret data, or to draw conclusions. All AI-assisted text was reviewed and edited by the authors, who take full responsibility for the content of the publication.

## Data Availability

The data analysed in this study are not publicly available because they are subject to access restrictions imposed by the data provider. Access to the data may be obtained from the Japan Medical Image Database subject to its eligibility requirements and approval procedures.

https://www.radiology.jp/j-mid/

## Data availability

Individual participant data were obtained from the Japan Medical Image Database (J-MID), established by the Japan Radiological Society with support from the Japan Agency for Medical Research and Development. J-MID data are collected under an opt-out consent model, are restricted to approved academic research, and cannot be redistributed by the authors, so no individual participant data, and no data dictionary describing them, can be released by us; both are held by J-MID. Researchers may apply to the J-MID office through the Japan Radiological Society, which reviews each proposal and requires a data use agreement with the contributing institutions; access is available on the same terms to any approved academic investigator, for any purpose within the approved scope, from the date of publication and without a fixed end date. Contributing centers are reported under pseudonyms and the mapping to real identifiers is not released.

## Code availability

There was no separate study protocol or statistical analysis plan registered before the analysis; the analysis plan is defined by the code. The complete analysis code — the quality-control rules, the modeling, and the code that generates every figure and table in this report, together with a run manifest recording every parameter and package version — is available at https://github.com/yamagishi0824/jmid-bone-ct-reference under an MIT license, with effect from publication. The reference centile tables for all 49 sites and both sexes are released with it under a CC BY 4.0 license.

## Acknowledgements

This research was partially supported by the Cross-ministerial Strategic Innovation Promotion Program (SIP) on “Integrated Health Care System” (grant number JPJ012425) and by JSPS KAKENHI (grant number JP23K17234). We thank the departments of radiology that provided the J-MID database, including Juntendo University, Kyushu University, Keio University, The University of Tokyo, Okayama University, Kyoto University, Osaka University, Tokyo Medical and Dental University, Hokkaido University, Ehime University, and Tokushima University. The present analysis used examinations from nine of these contributing centers. The funders had no role in study design, data collection, data analysis, data interpretation, the writing of the manuscript, or the decision to submit for publication.

## Author contributions

YY conceived and designed the study, developed the methodology, performed the statistical analyses, interpreted the data, and drafted the manuscript. TK contributed to the conception and design of the study, interpretation of the results, and critical revision of the manuscript. KY performed data collection, image segmentation, and data processing and analysis. SH, TY and OA supervised the study, interpreted the results, and critically reviewed and revised the manuscript. YY and KY directly accessed and verified the underlying data reported in the manuscript. All authors had full access to all the data in the study, reviewed and approved the final manuscript, and accept responsibility for the decision to submit for publication.

## References

1. Morin, S. N., Leslie, W. D. & Schousboe, J. T. Osteoporosis: a review. JAMA 334, 894–907 (2025).

2. Wu, A.-M. et al. Global, regional, and national burden of bone fractures in 204 countries and territories, 1990–2019: a systematic analysis from the Global Burden of Disease Study 2019. Lancet Healthy Longev. 2, e580–e592 (2021).

3. Williamson, S. et al. Costs of fragility hip fractures globally: a systematic review and meta-regression analysis. Osteoporos. Int. 28, 2791–2800 (2017).

4. Choi, Y. J. Dual-energy X-ray absorptiometry: beyond bone mineral density determination. Endocrinol. Metab. 31, 25–30 (2016).

5. Golding, P. H. Dual-energy X-ray absorptiometry (DXA) to measure bone mineral density (BMD) for diagnosis of osteoporosis - experimental data from artificial vertebrae confirms significant dependence on bone size. Bone Rep. 17, 101607 (2022).

6. Praveen, A. D., Sollmann, N., Baum, T., Ferguson, S. J. & Helgason, B. CT image-based biomarkers for opportunistic screening of osteoporotic fractures: a systematic review and meta-analysis. Osteoporos. Int. 35, 971–996 (2024).

7. Pickhardt, P. J. Value-added opportunistic CT screening: state of the art. Radiology 303, 241–254 (2022).

8. Pickhardt, P. J. et al. Opportunistic screening for osteoporosis using abdominal computed tomography scans obtained for other indications. Ann. Intern. Med. 158, 588–595 (2013).

9. Pickhardt, P. J. et al. Population-based opportunistic osteoporosis screening: validation of a fully automated CT tool for assessing longitudinal BMD changes. Br. J. Radiol. 92, 20180726 (2019).

10. Westerhoff, M. et al. Deep learning–based opportunistic CT osteoporosis screening and the establishment of normative values. Radiology 317, e250917 (2025).

11. Yaprak, G., Gemici, C., Seseogullari, O. O., Karabag, I. S. & Cini, N. CT derived Hounsfield unit: an easy way to determine osteoporosis and radiation related fracture risk in irradiated patients. Front. Oncol. 10, 742 (2020).

12. Wasserthal, J. et al. TotalSegmentator: robust segmentation of 104 anatomic structures in CT images. Radiol. Artif. Intell. 5, e230024 (2023).

13. Akashi, T. et al. Japan-Medical Image Database (J-MID): medical big data supporting data science. Juntendo Med. J. 71, 166–172 (2025).

14. Wachinger, C., Renger, B., Späth, C., Kirschke, J. & Makowski, M. Whole body CT attenuation and volume charts from routine clinical scans via LLM report filtering. npj Digit. Med. 9, 505 (2026).

15. Wachinger, C., Renger, B., Späth, C. & Makowski, M. R. Body charts from CT segmentations across the adult lifespan: large-scale cross-sectional and longitudinal analyses. Radiol. Artif. Intell. 8, e250506 (2026).

16. Riggs, B. L. et al. Population-based analysis of the relationship of whole bone strength indices and fall-related loads to age- and sex-specific patterns of hip and wrist fractures. J. Bone Miner. Res. 21, 315–323 (2006).

17. Yasui, T. et al. Factors associated with premature ovarian failure, early menopause and earlier onset of menopause in Japanese women. Maturitas 72, 249–255 (2012).

18. Bland, J. M. & Altman, D. G. Statistical methods for assessing agreement between two methods of clinical measurement. Lancet 1, 307–310 (1986).

19. Boutin, R. D. & Lenchik, L. Value-added opportunistic CT: insights into osteoporosis and sarcopenia. AJR Am. J. Roentgenol. 215, 582–594 (2020).

20. Kaesmacher, J., Liebl, H., Baum, T. & Kirschke, J. S. Bone mineral density estimations from routine multidetector CT: a comparative study of contrast and calibration effects. J. Comput. Assist. Tomogr. 41, 217–223 (2017).

21. Yoshimura, N. et al. Prevalence of knee osteoarthritis, lumbar spondylosis, and osteoporosis in Japanese men and women: the research on osteoarthritis/osteoporosis against disability study. J. Bone Miner. Metab. 27, 620–628 (2009).

22. Wood, S. N. Generalized Additive Models: An Introduction with R 2nd edn (CRC Press, 2017).

23. Koenker, R. & Bassett, G. Regression quantiles. Econometrica 46, 33–50 (1978).

24. Cole, T. J. & Green, P. J. Smoothing reference centile curves: the LMS method and penalized likelihood. Stat. Med. 11, 1305–1319 (1992).

25. Wei, Y., Pere, A., Koenker, R. & He, X. Quantile regression methods for reference growth charts. Stat. Med. 25, 1369–1382 (2006).

26. Linnet, K. Evaluation of regression procedures for methods comparison studies. Clin. Chem. 39, 424–432 (1993).

27. DeLong, E. R., DeLong, D. M. & Clarke-Pearson, D. L. Comparing the areas under two or more correlated receiver operating characteristic curves: a nonparametric approach. Biometrics 44, 837–845 (1988).

28. McGraw, K. O. & Wong, S. P. Forming inferences about some intraclass correlation coefficients. Psychol. Methods 1, 30–46 (1996).

29. Benjamini, Y. & Hochberg, Y. Controlling the false discovery rate: a practical and powerful approach to multiple testing. J. R. Stat. Soc. B 57, 289–300 (1995).

30. Kanis, J. A. et al. A reference standard for the description of osteoporosis. Bone 42, 467–475 (2008).

