## Supplementary information for "Sex-specific skeletal aging trajectories and reference centiles across vertebrae and ribs in over 78,000 patients"

**Contents**

Supplementary Figure 1. Study flow.

Supplementary Figure 2. Description of the cohort.

Supplementary Figure 3. Fitted mean CT attenuation against age for all 25 vertebral levels, with 95% confidence bands.

Supplementary Figure 4. Fitted mean CT attenuation against age for the left ribs, levels 1–12, with 95% confidence bands.

Supplementary Figure 5. Fitted mean CT attenuation against age for the right ribs, levels 1–12, with 95% confidence bands.

Supplementary Figure 6. Rate of change of CT attenuation against age for all 25 vertebral levels, with 95% confidence bands.

Supplementary Figure 7. Rate of change of CT attenuation against age for the left ribs, levels 1–12, with 95% confidence bands.

Supplementary Figure 8. Rate of change of CT attenuation against age for the right ribs, levels 1–12, with 95% confidence bands.

Supplementary Figure 9. Reference centiles (3rd, 10th, 25th, 50th, 75th, 90th and 97th) of CT attenuation for all 25 vertebral levels in men.

Supplementary Figure 10. Reference centiles of CT attenuation for all 25 vertebral levels in women.

Supplementary Figure 11. Reference centiles of CT attenuation for all 24 ribs in men.

Supplementary Figure 12. Reference centiles of CT attenuation for all 24 ribs in women.

Supplementary Figure 13. Age-adjusted correlation between every pair of the 49 skeletal sites, in men (A) and women (B).

Supplementary Figure 14. Conversion of attenuation at four representative sites to the L1 scale, by sex.

Supplementary Figure 15. Bland–Altman plots of left minus right rib attenuation for all 12 rib levels.

Supplementary Figure 16. Sensitivity of the fitted trajectories.

Supplementary Figure 17. Between-center variation in CT attenuation.

Supplementary Table 1. Quality control by skeletal site.

Supplementary Table 2. Trajectory statistics for all 49 skeletal sites.

Supplementary Table 2B. Difference between the sexes in the age trajectory, by site.

Supplementary Table 3. Reference centiles of CT attenuation by site, sex and age.

Supplementary Table 3B. Diagnostics of the centile fits.

Supplementary Table 4. Prevalence of L1 attenuation below published trabecular cut-offs.

Supplementary Table 5. Young-adult reference values.

Supplementary Table 6. Prevalence of a young-adult-referenced score below −2.5.

Supplementary Table 7. Sensitivity of the trajectories to the quality-control rule.

Supplementary Table 7B. Summary of the sensitivity analyses.

Supplementary Table 8. Attenuation by contributing center and skeletal site, adjusted for age and sex.

Supplementary Table 9. Leave-one-center-out refits.

Supplementary Table 10. Body-mass-index-adjusted models.

Supplementary Table 11. Dependence of mean attenuation on segmented volume.

Supplementary Table 12. Association with L1 and surrogacy, all sites.

Supplementary Table 13. Agreement between left and right ribs.

Supplementary Table 14. Between-center spread of attenuation by skeletal site.

**Supplementary figures**

**Supplementary Figure 1: Study flow. Examinations and patients remaining after each step (left column) with the exclusion applied to reach it (right). The first three steps were performed when the data were extracted from the Japan Medical Image Database; the segmentation export analyzed here begins at the 79 482 examinations of the third box, so those counts are reported as supplied by the extraction step. From that box onwards each examination comes from a different patient, so the two counts coincide.**

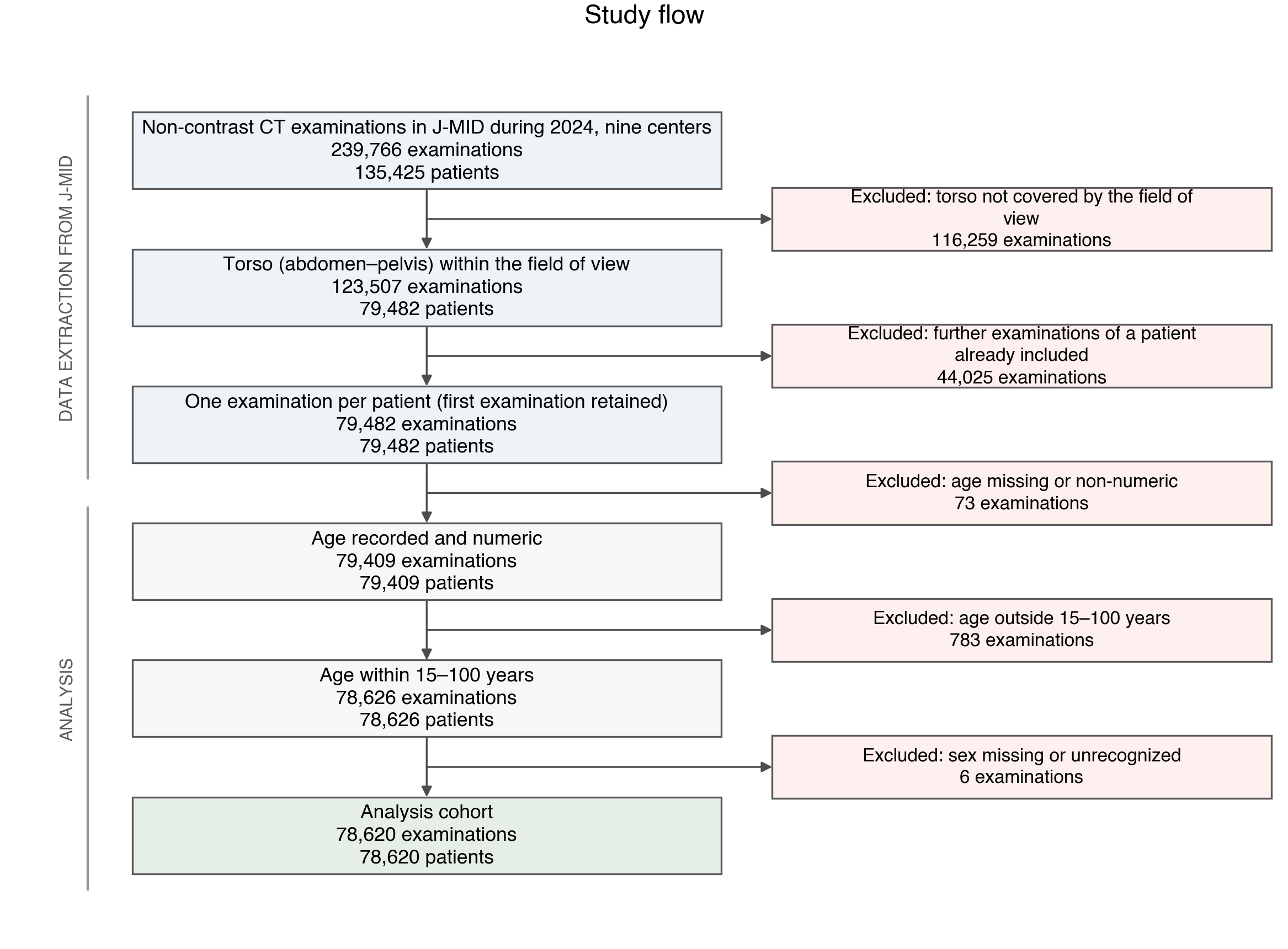

**Supplementary Figure 2: Description of the cohort.** (A) Age distribution by sex. (B) Number of patients contributed by each center, under pseudonyms ordered by decreasing size. (C) Distribution of body-mass index among the patients for whom height and weight were recorded.

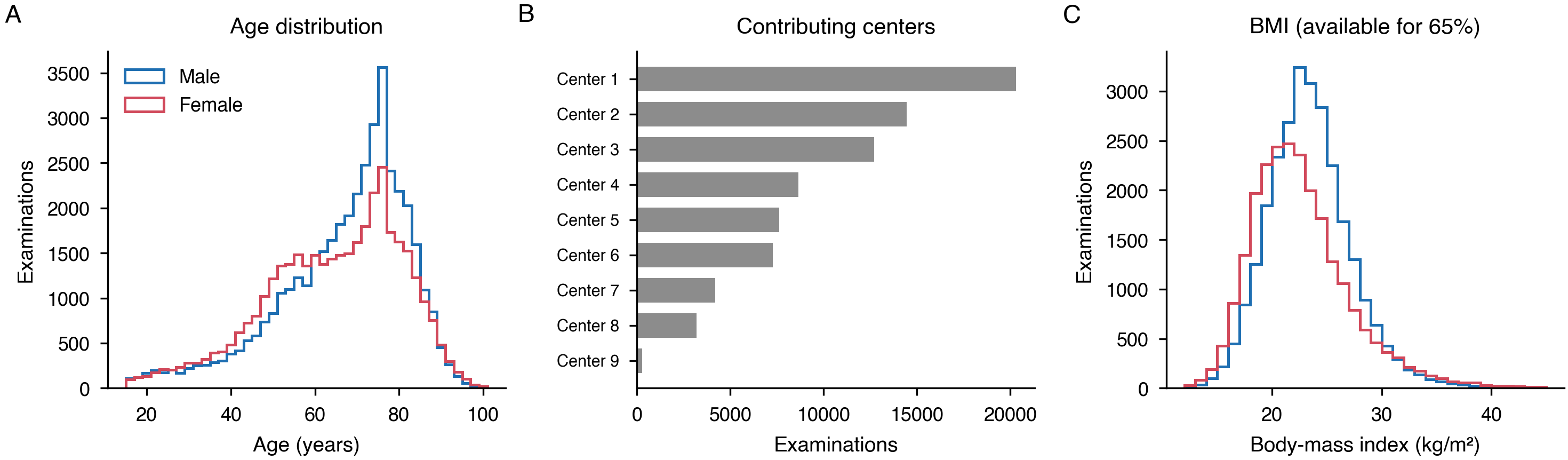

**Supplementary Figure 3: Fitted mean CT attenuation against age for all 25 vertebral levels, with 95% confidence bands.** Blue, men; red, women.

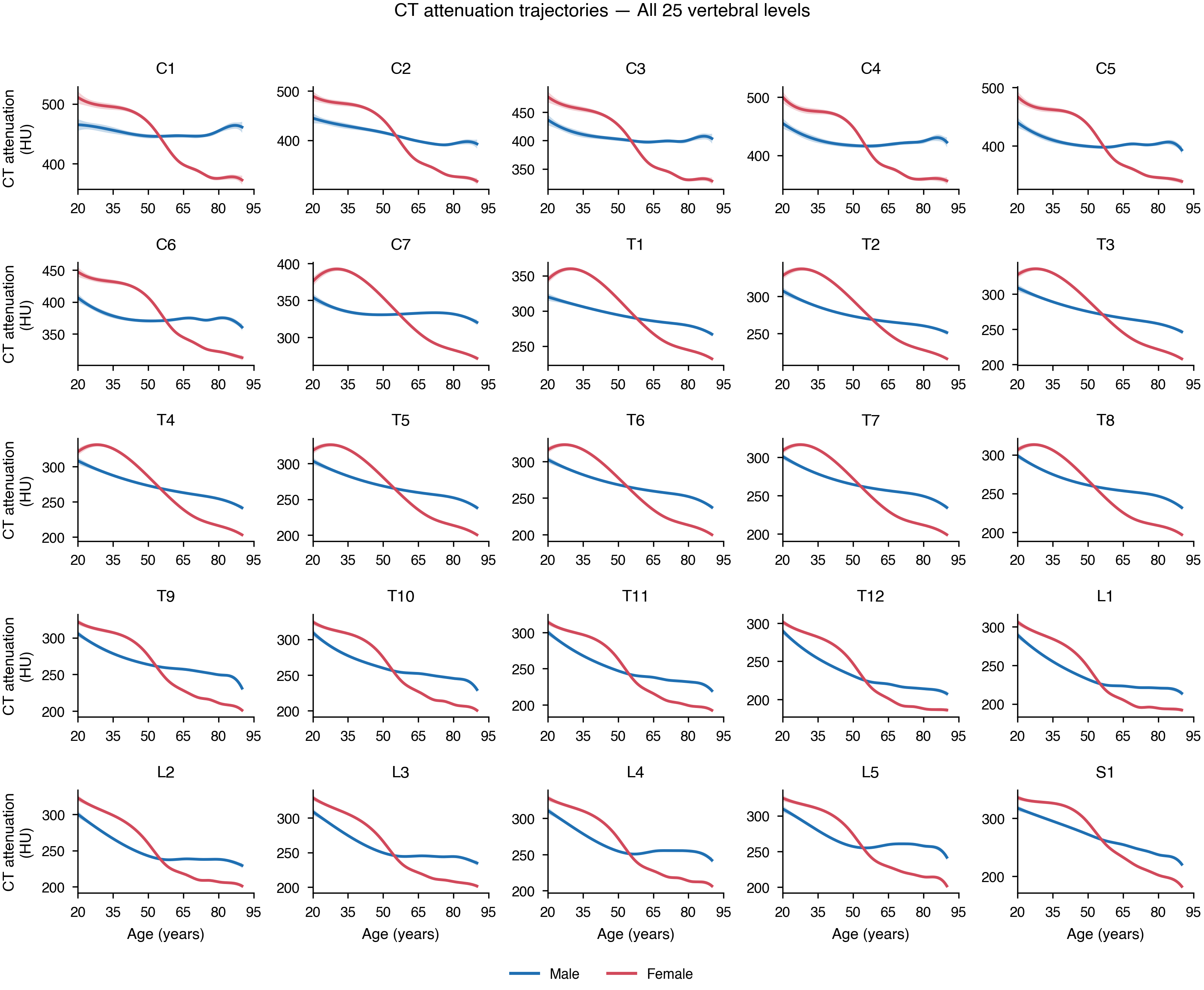

**Supplementary Figure 4: Fitted mean CT attenuation against age for the left ribs, levels 1–12, with 95% confidence bands.** Blue, men; red, women.

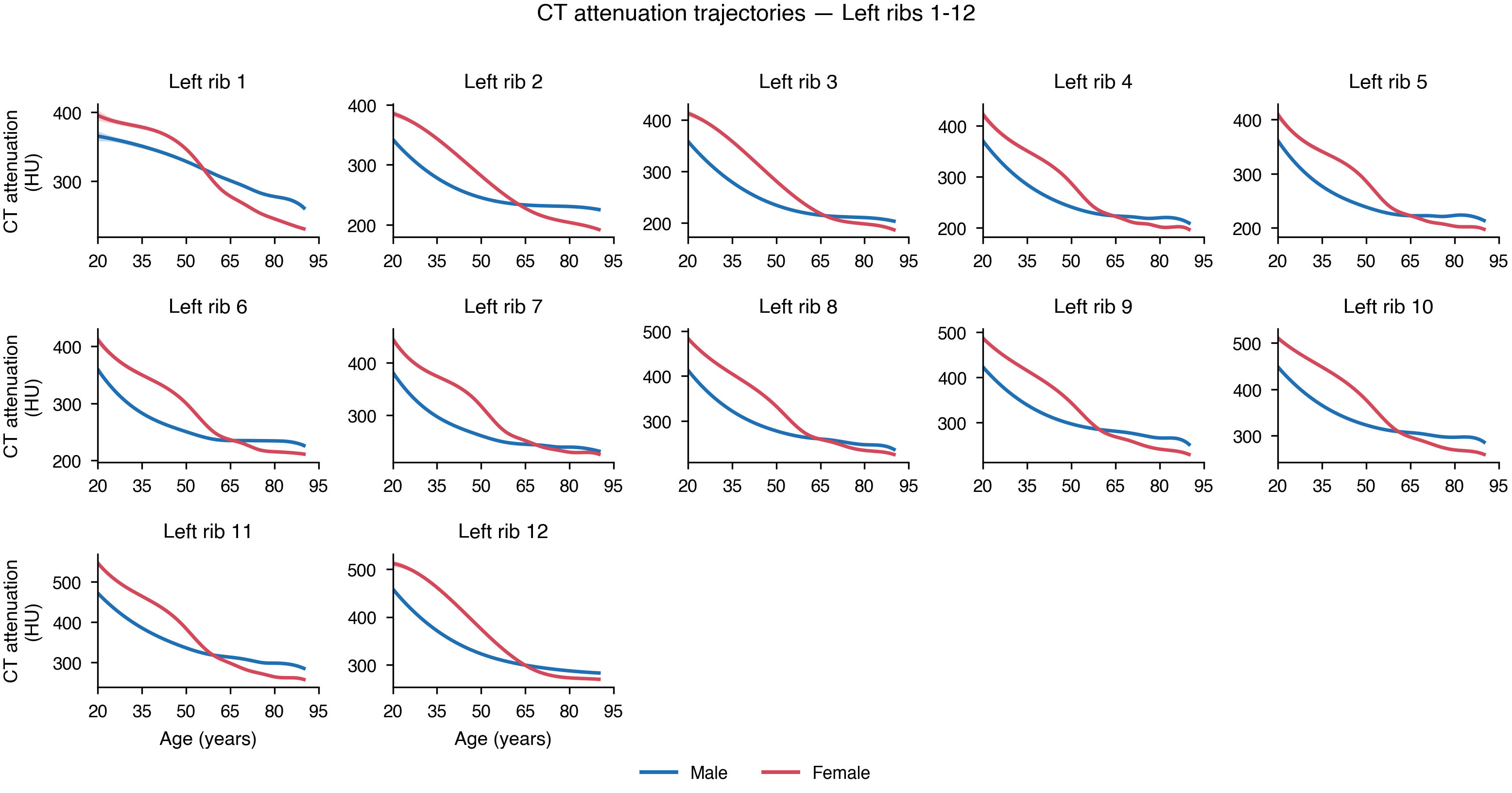

**Supplementary Figure 5: Fitted mean CT attenuation against age for the right ribs, levels 1–12, with 95% confidence bands.** Blue, men; red, women.

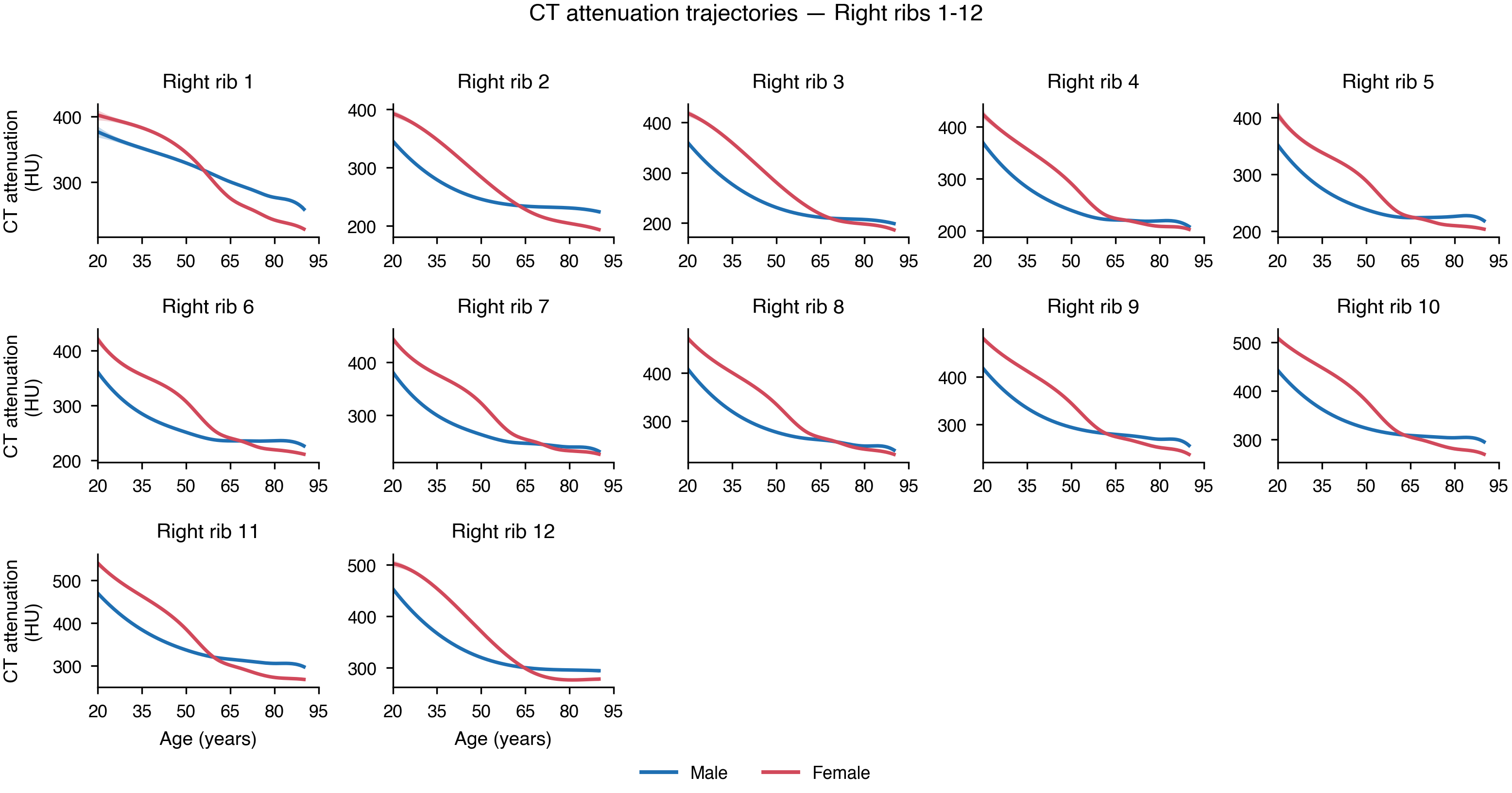

**Supplementary Figure 6: Rate of change of CT attenuation against age for all 25 vertebral levels, with 95% confidence bands.** The rate is the analytic derivative of the fitted spline. Blue, men; red, women.

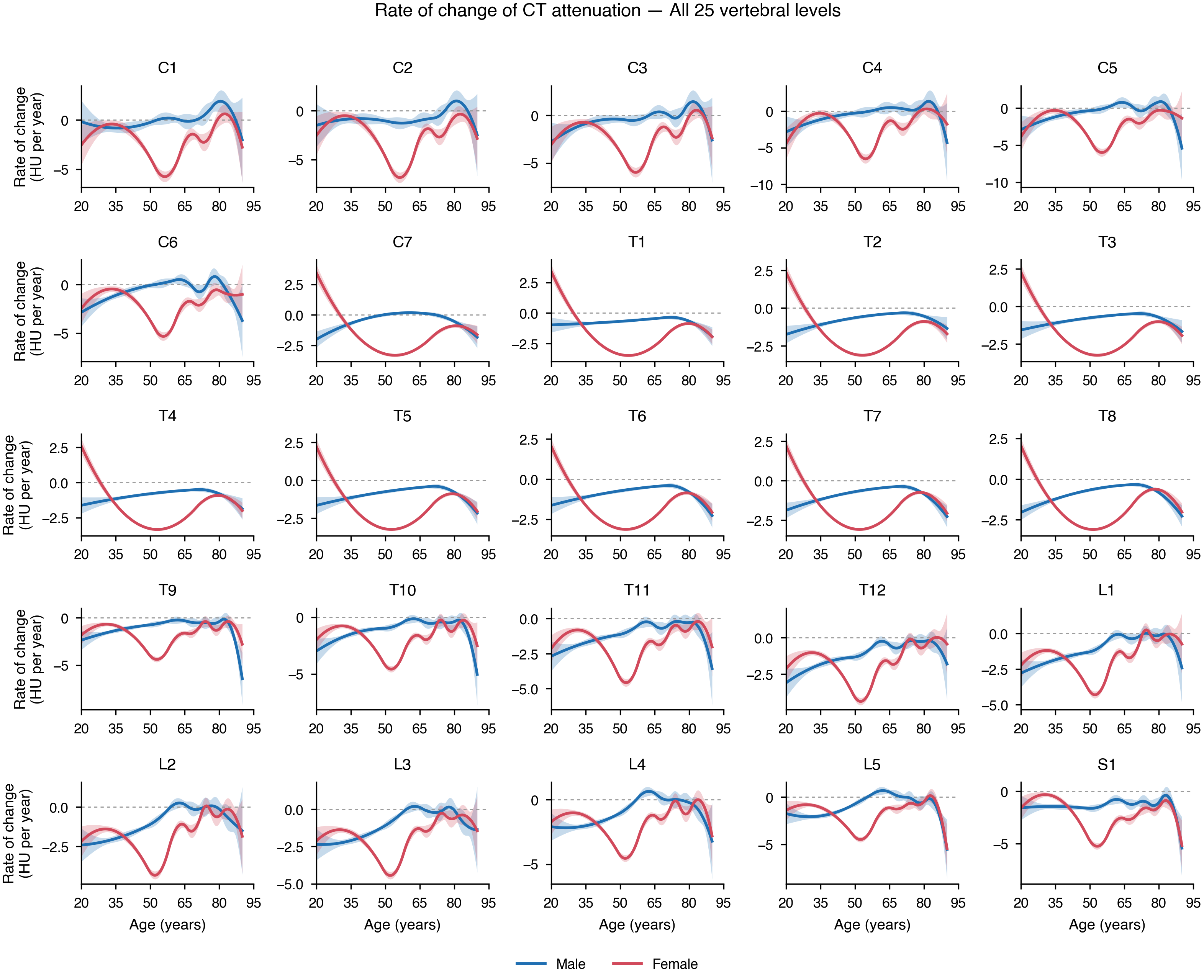

**Supplementary Figure 7: Rate of change of CT attenuation against age for the left ribs, levels 1–12, with 95% confidence bands.** Blue, men; red, women.

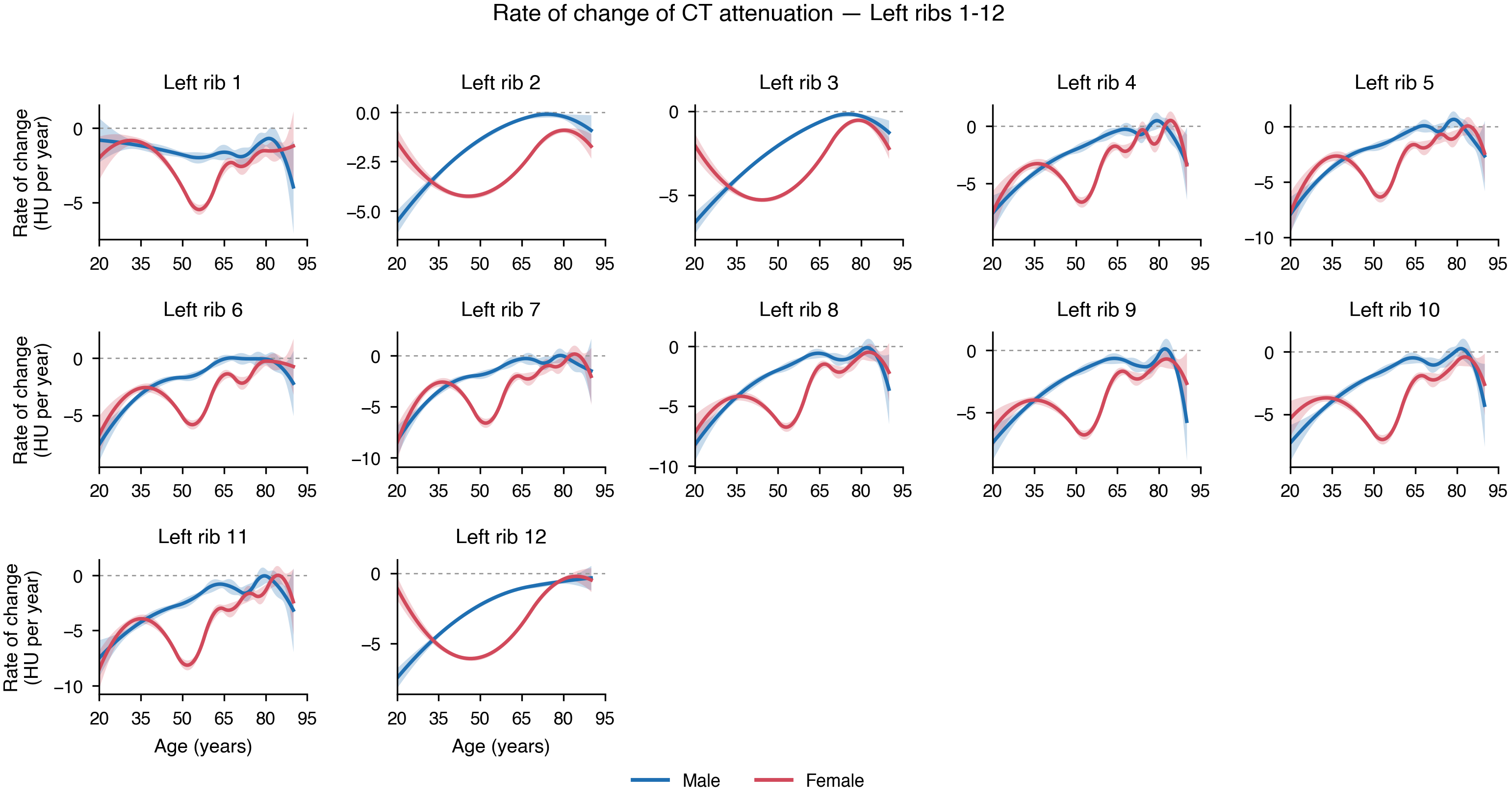

**Supplementary Figure 8: Rate of change of CT attenuation against age for the right ribs, levels 1–12, with 95% confidence bands.** Blue, men; red, women.

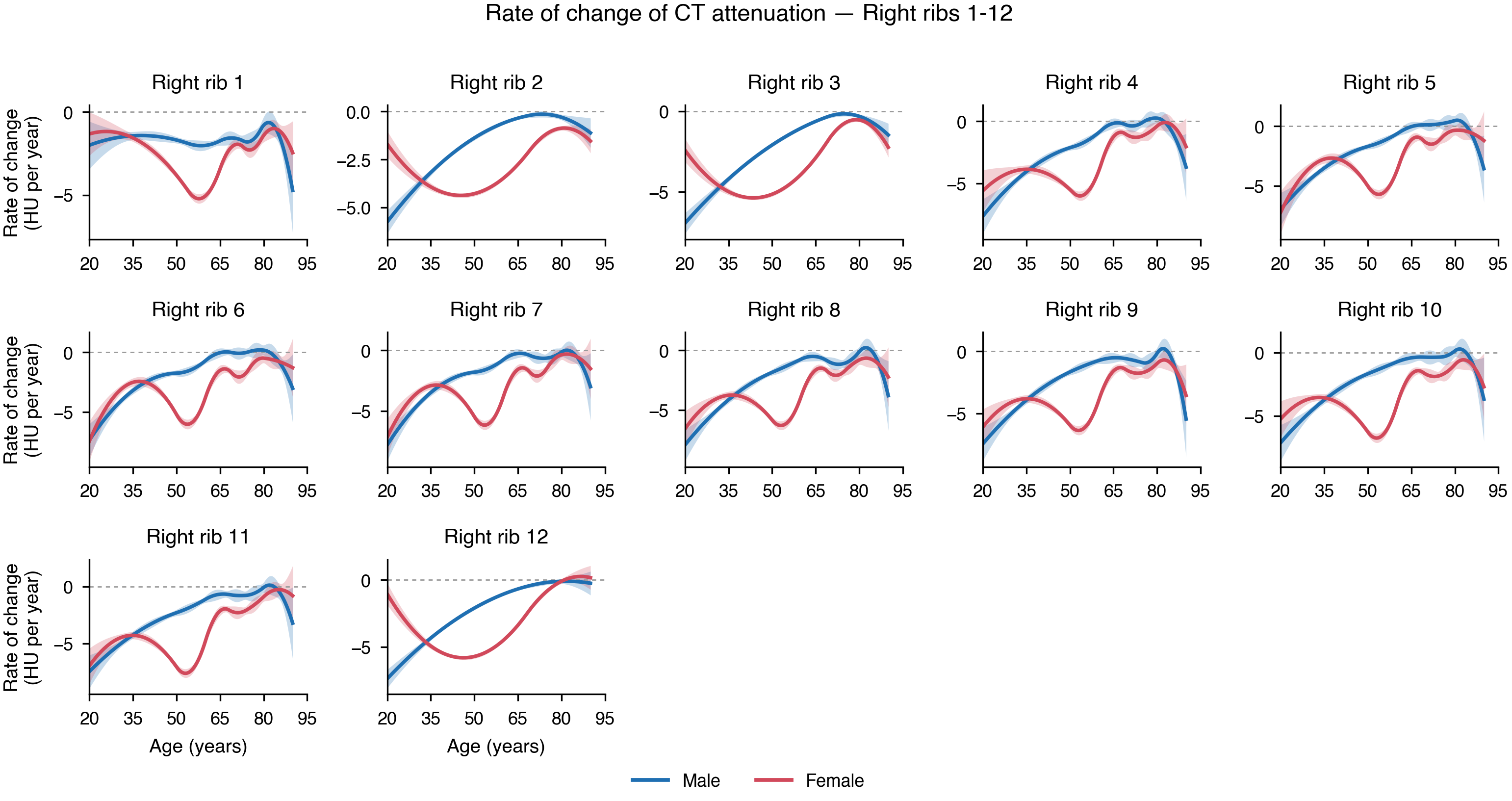

**Supplementary Figure 9: Reference centiles (3rd, 10th, 25th, 50th, 75th, 90th and 97th) of CT attenuation for all 25 vertebral levels in men.** The heavy line is the median.

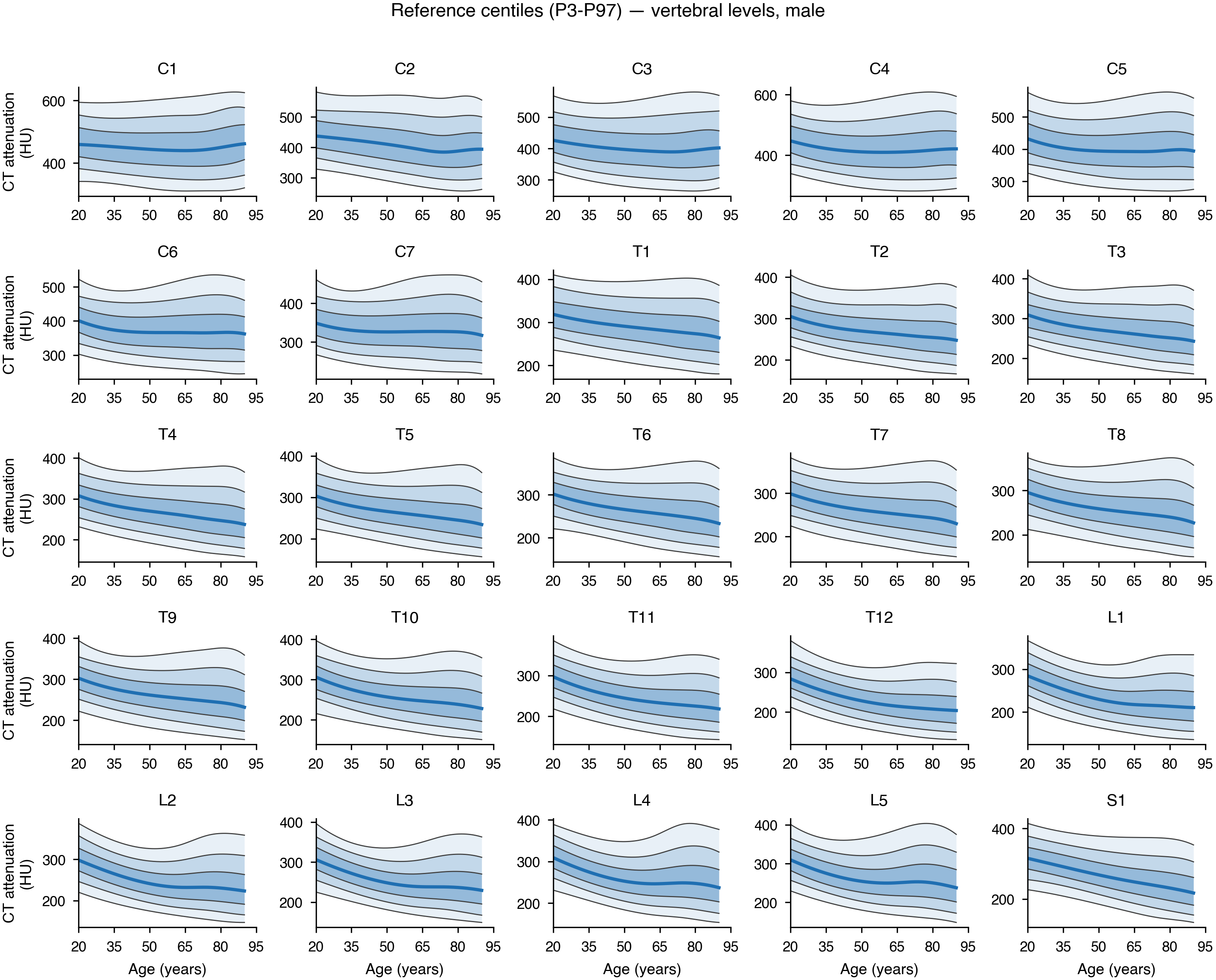

**Supplementary Figure 10: Reference centiles of CT attenuation for all 25 vertebral levels in women.** The heavy line is the median.

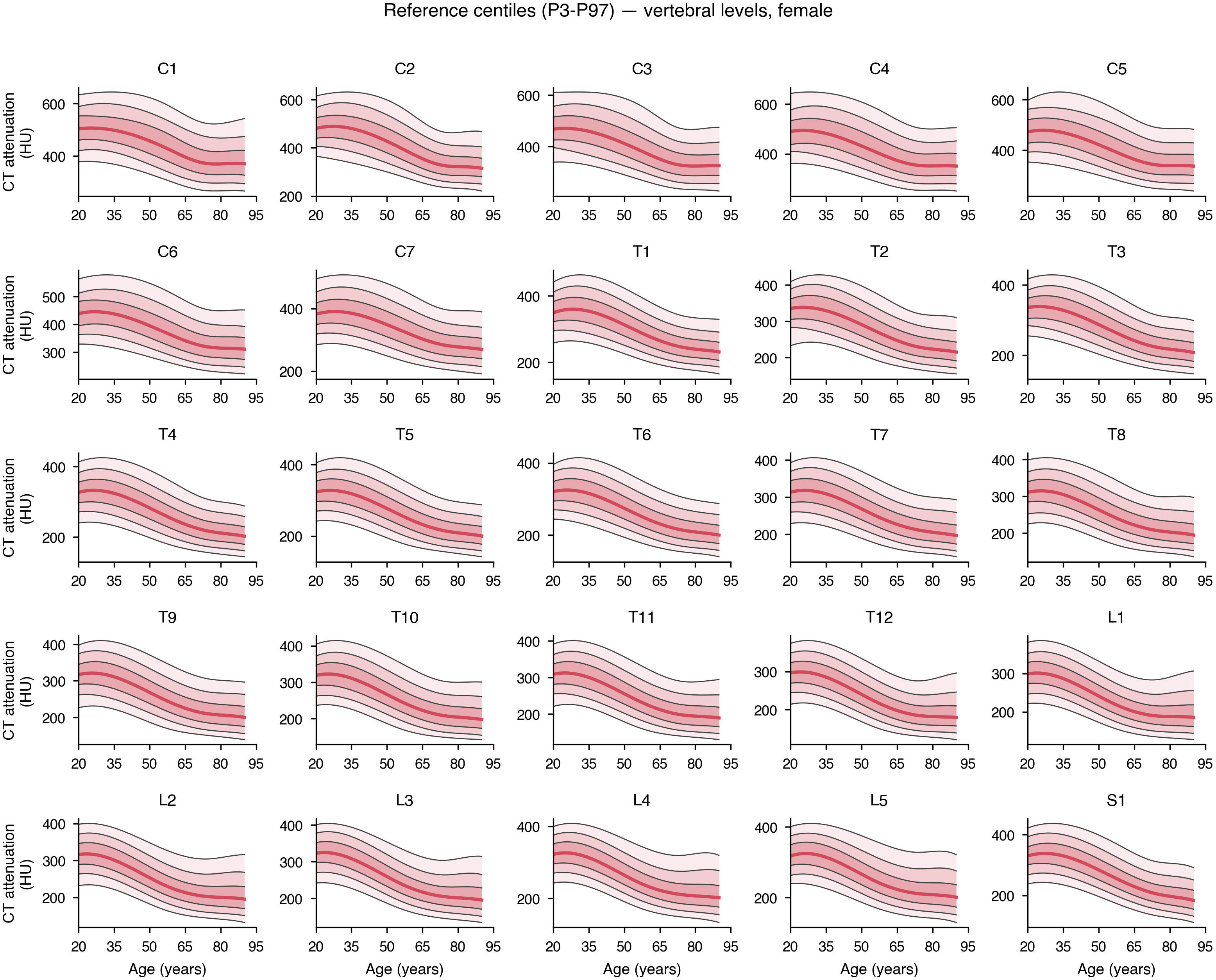

**Supplementary Figure 11: Reference centiles of CT attenuation for all 24 ribs in men.** The heavy line is the median.

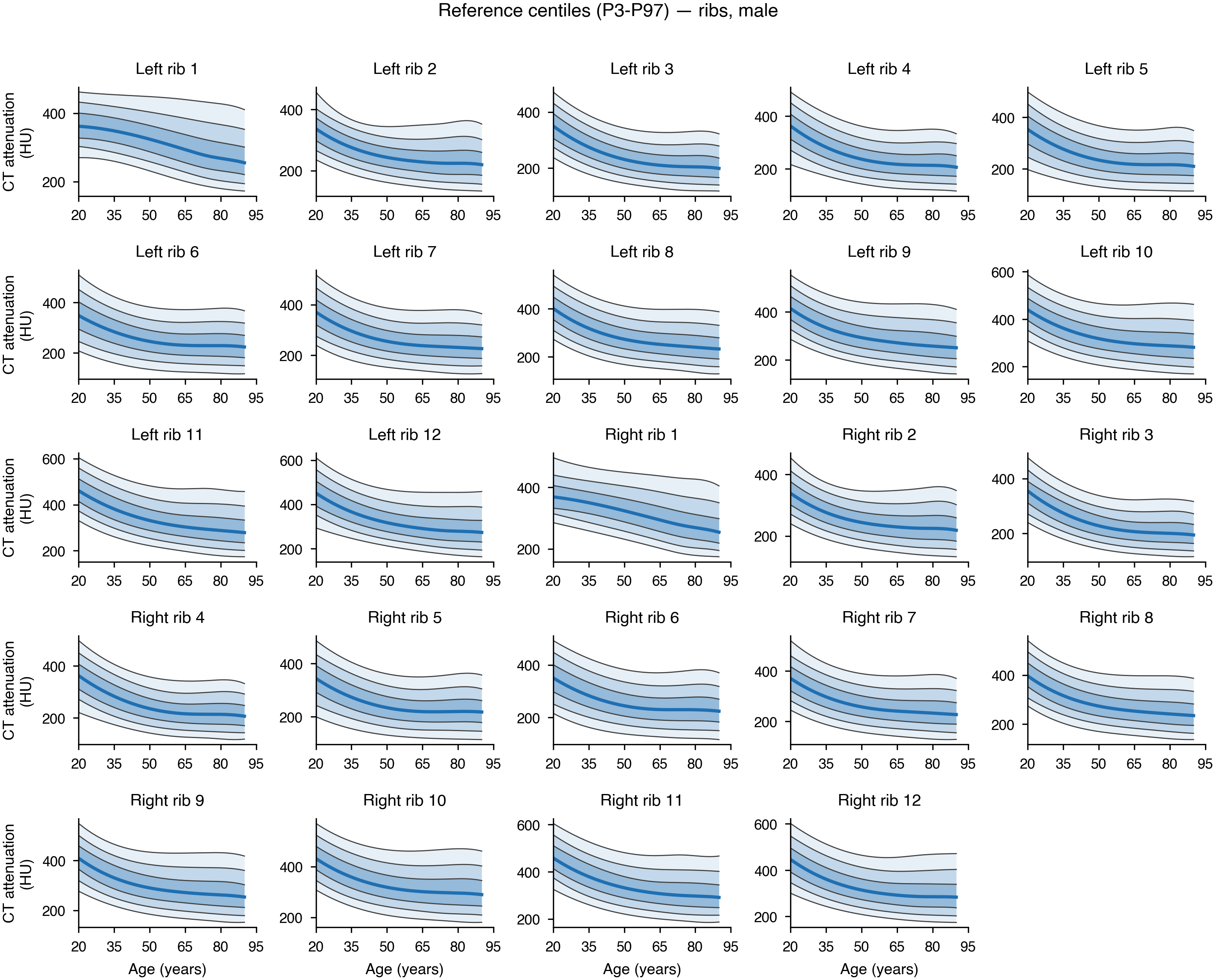

**Supplementary Figure 12: Reference centiles of CT attenuation for all 24 ribs in women.** The heavy line is the median.

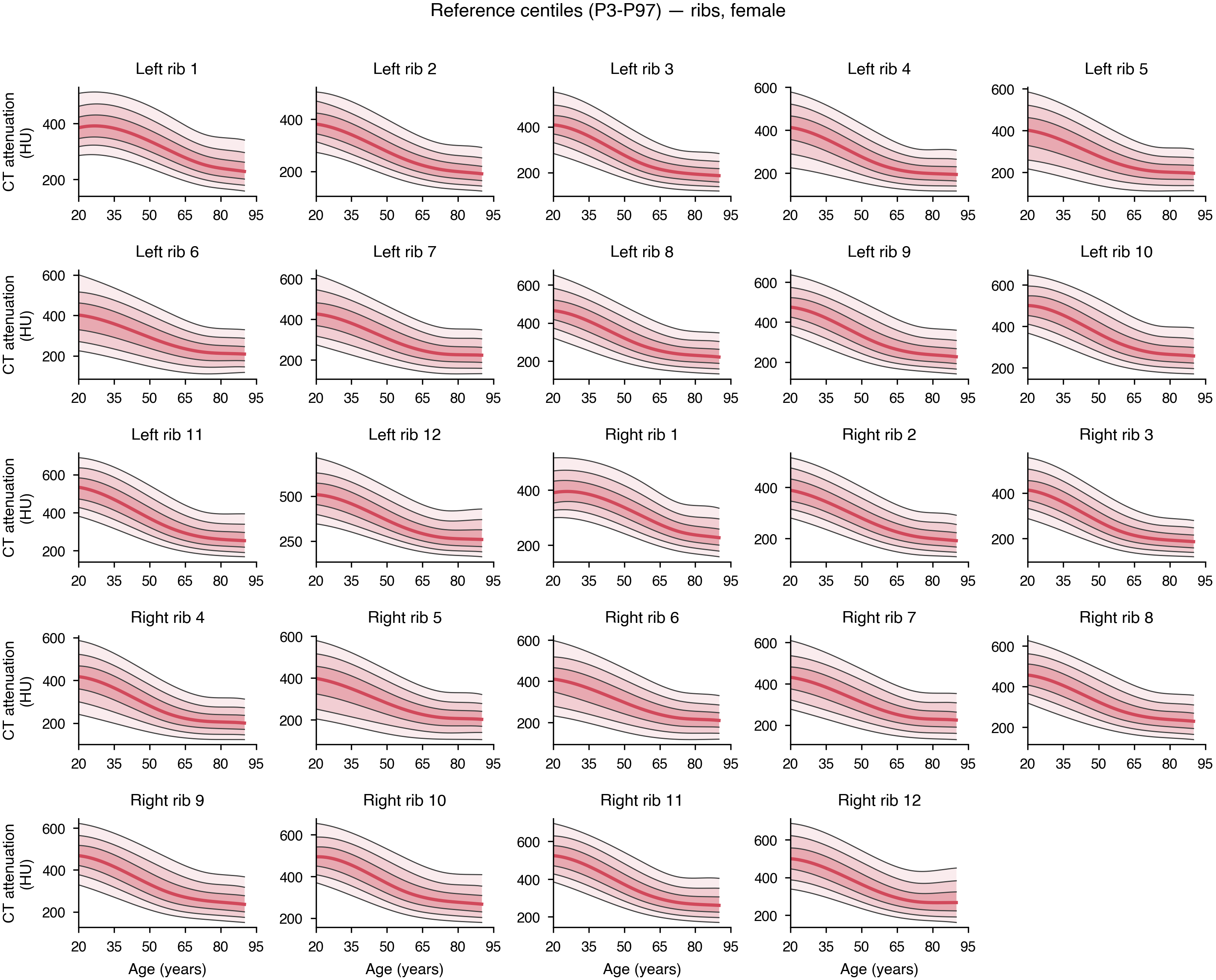

**Supplementary Figure 13: Age-adjusted correlation between every pair of the 49 skeletal sites, in men (A) and women (B).** Correlations are computed on residuals after removing a spline in age from both sites.

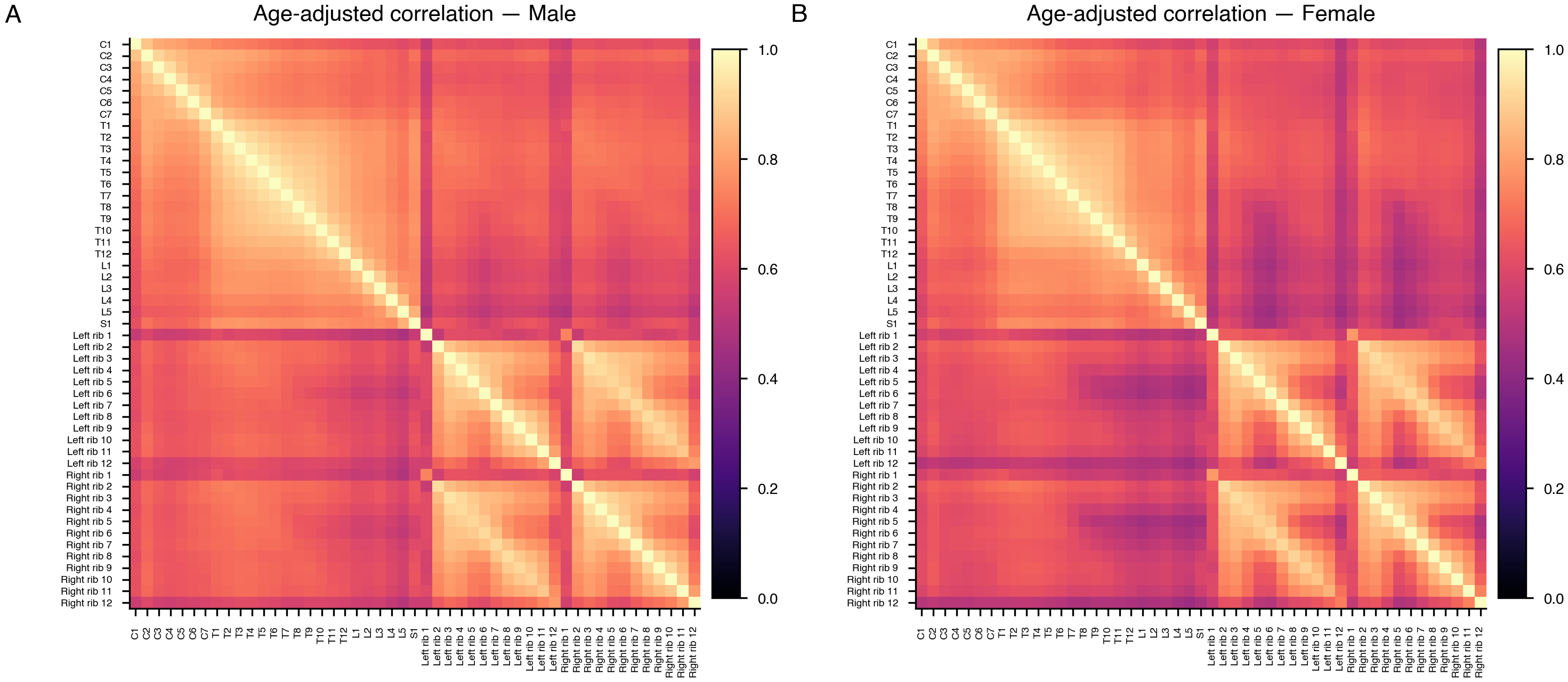

**Supplementary Figure 14: Conversion of attenuation at four representative sites to the L1 scale, by sex.** Points are individual patients (random subsample); the line is the Deming (errors-in-variables) fit, whose equation and prediction-error SD are given in each panel title.

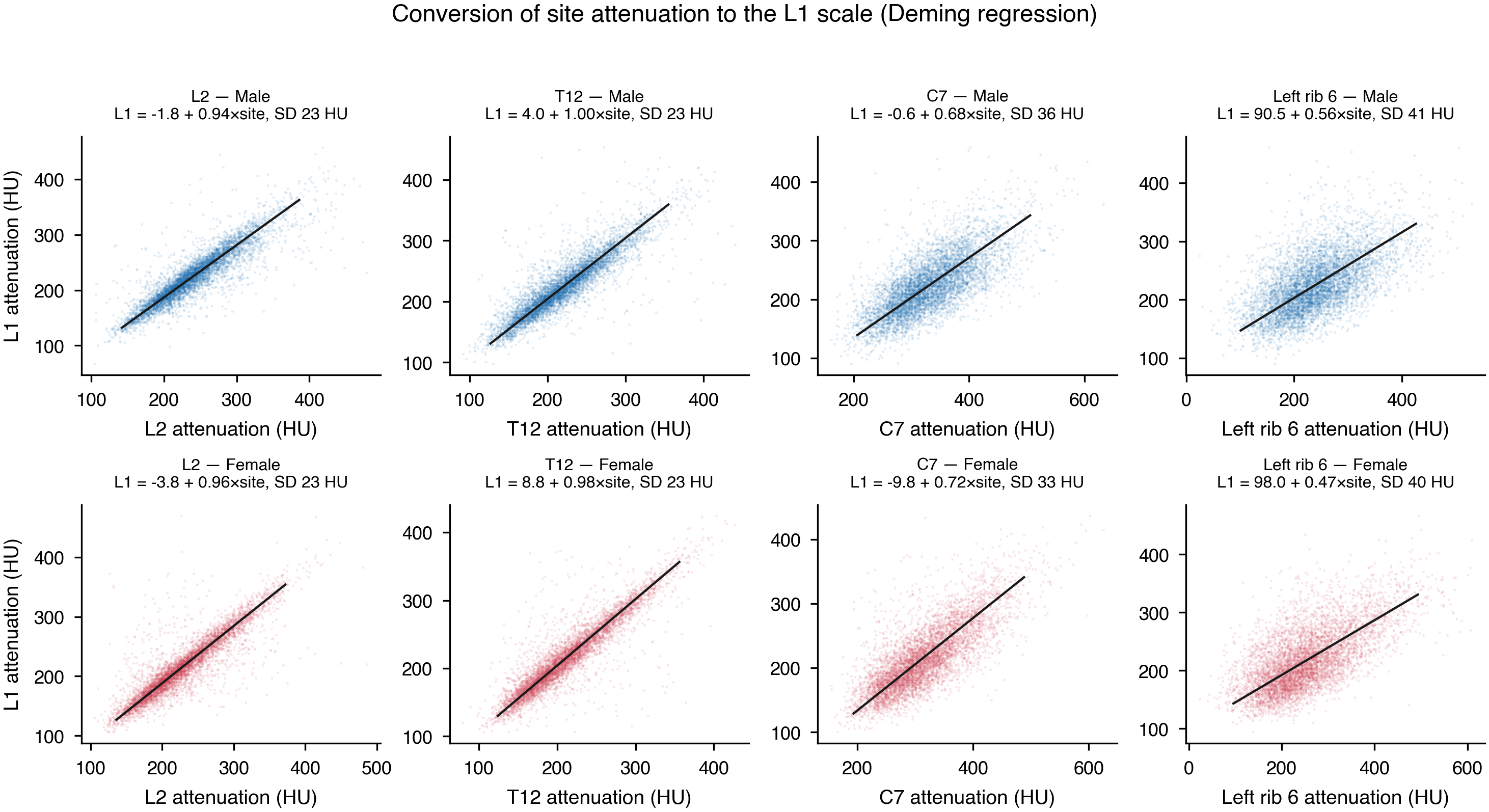

**Supplementary Figure 15: Bland–Altman plots of left minus right rib attenuation for all 12 rib levels.** Blue, men; red, women. Mean difference (solid) and 95% limits of agreement (dashed) are drawn separately within each sex, in the corresponding color, and each panel title gives the intraclass correlation and the limits of agreement for men (M) and women (F); the same statistics are tabulated in Supplementary Table 13.

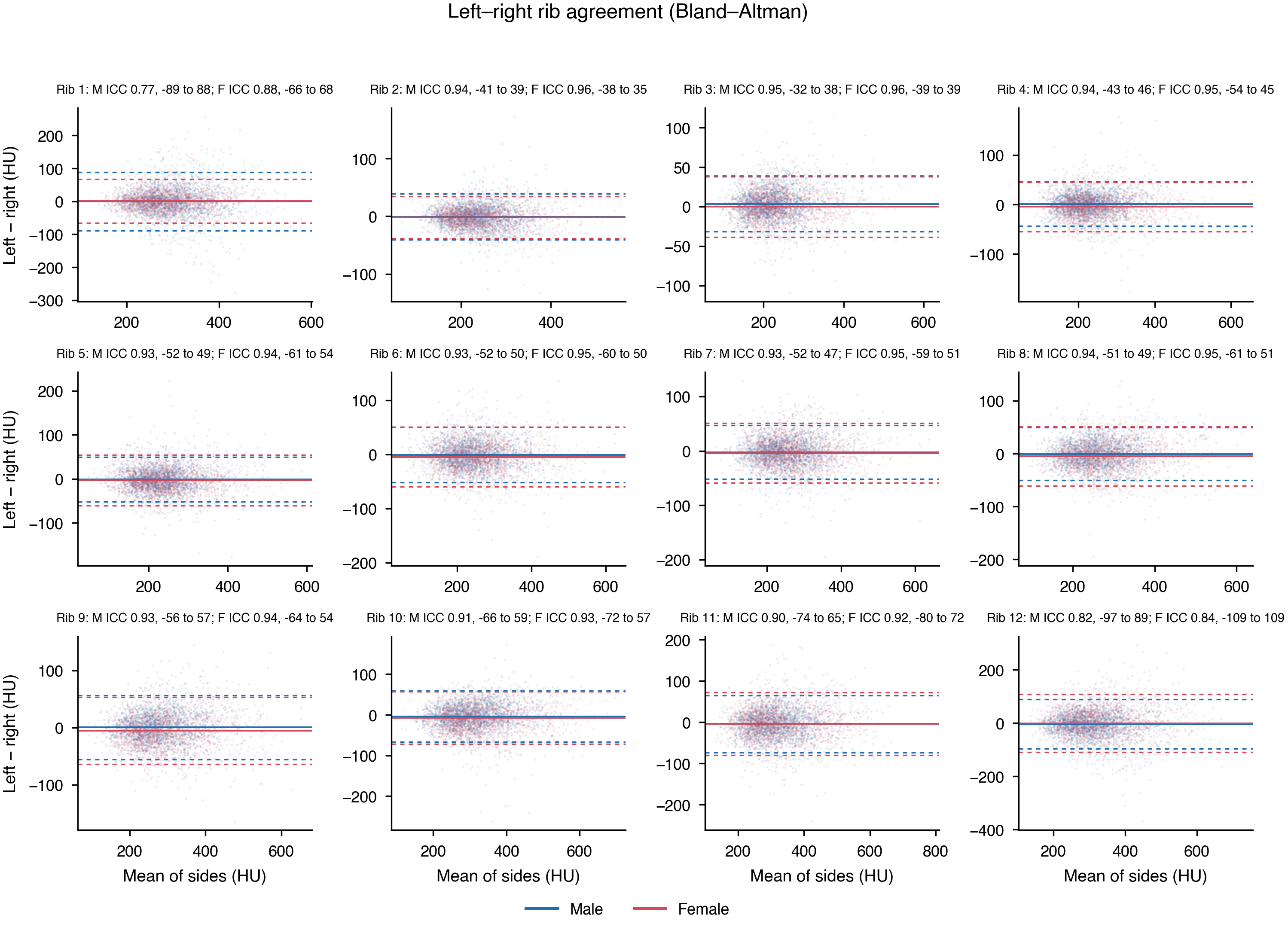

**Supplementary Figure 16: Sensitivity of the fitted trajectories.** Upper row: primary quality-control rule (solid) against the legacy 5th–95th percentile rule (dashed). Lower row: the trajectory refitted with each center omitted in turn, one line per omitted center. Blue, men; red, women.

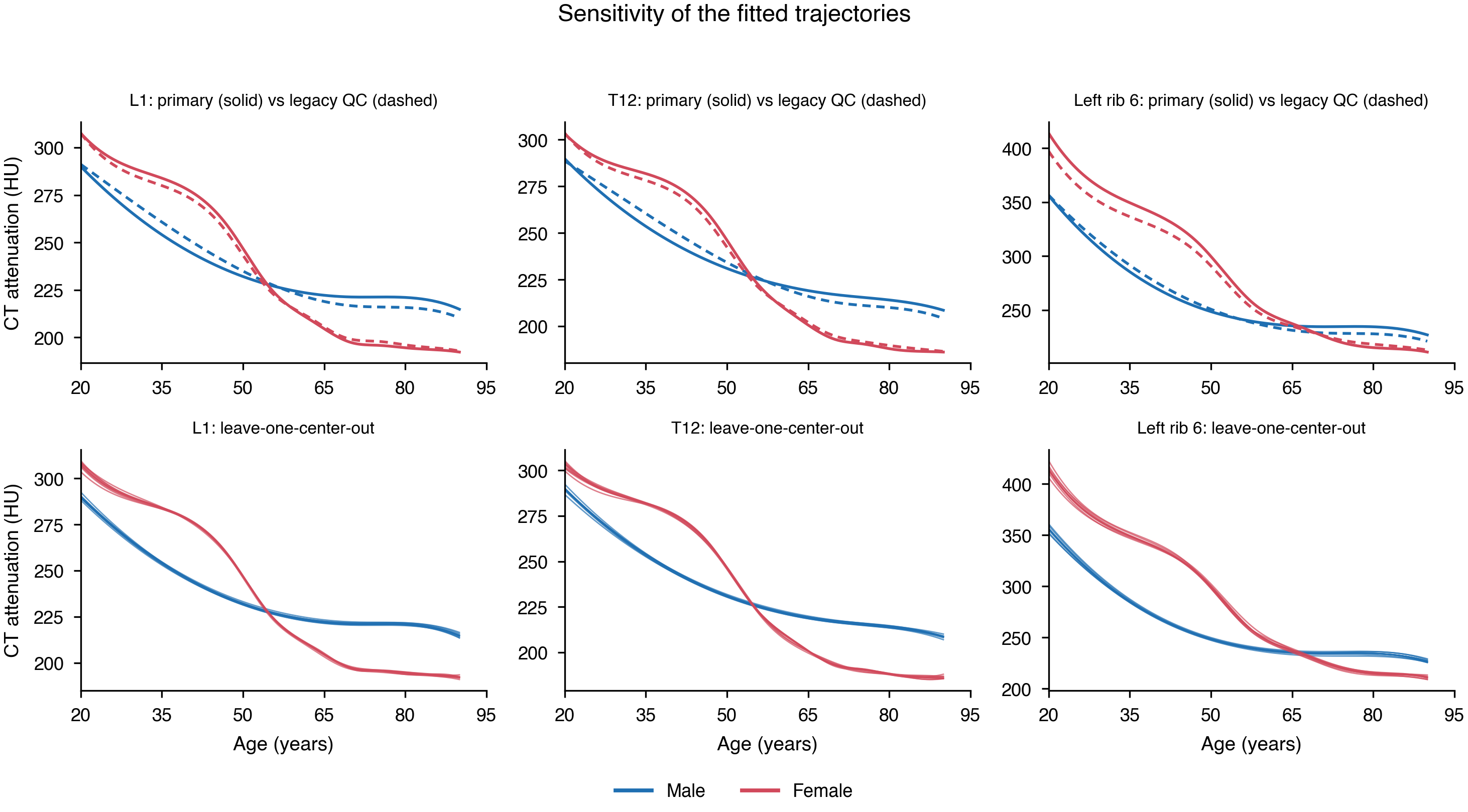

**Supplementary Figure 17: Between-center variation in CT attenuation.** (A) Age- and sex-adjusted deviation of each pseudonymized center from the cohort mean, for all 49 sites ordered anatomically. (B) Range of those deviations by anatomical region; the between-center range is smallest in the thoracolumbar spine and smallest of all at L1. (C) Between-center range against the error of converting a site to the L1 scale, showing that sites with a wide between-center range are also the poorer substitutes for L1. Between-center variation is an adjusted spread across centers, not a direct measure of scanner repeatability.

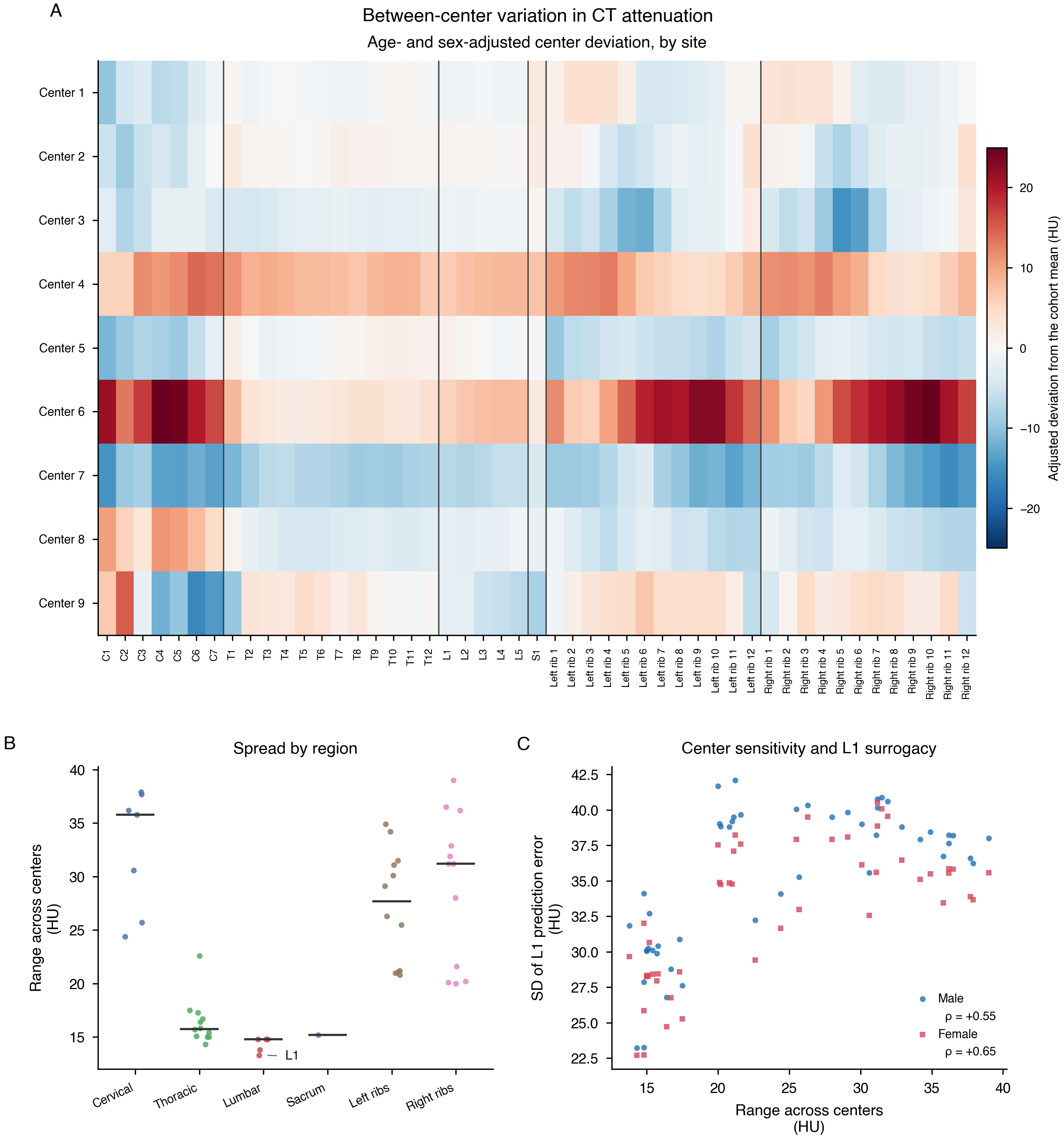

**Supplementary Table 1: Quality control by skeletal site**

| **Site** | **Region** | **Segmented, n** | **Excluded by adjacency rule, n** | **Excluded by truncation rule, n** | **Excluded by artifact rule, n** | **Analyzed, n** | **Men, n** | **Women, n** | **Analyzed, % of cohort** | **Median attenuation (HU)** | **Median segmented volume (cm³)** |
| --- | --- | --- | --- | --- | --- | --- | --- | --- | --- | --- | --- |
| C1 | Cervical vertebrae | 27704 | 1 | 1406 | 2 | 26295 | 12941 | 13354 | 33.4 | 427.5 | 13.9 |
| C2 | Cervical vertebrae | 29753 | 2050 | 1404 | 11 | 26288 | 12939 | 13349 | 33.4 | 381 | 18.7 |
| C3 | Cervical vertebrae | 32505 | 2752 | 1496 | 19 | 28238 | 13945 | 14293 | 35.9 | 379 | 12.5 |
| C4 | Cervical vertebrae | 38883 | 6379 | 1618 | 14 | 30872 | 15517 | 15355 | 39.3 | 400.5 | 12.7 |
| C5 | Cervical vertebrae | 50264 | 11384 | 1962 | 13 | 36905 | 19024 | 17881 | 46.9 | 384.5 | 14 |
| C6 | Cervical vertebrae | 56215 | 5953 | 2562 | 23 | 47677 | 25074 | 22603 | 60.6 | 358 | 15.1 |
| C7 | Cervical vertebrae | 56943 | 728 | 2858 | 49 | 53308 | 28177 | 25131 | 67.8 | 317 | 18.8 |
| T1 | Thoracic vertebrae | 56960 | 0 | 2862 | 58 | 54040 | 28549 | 25491 | 68.7 | 275 | 23.7 |
| T2 | Thoracic vertebrae | 56975 | 17 | 2878 | 79 | 54001 | 28532 | 25469 | 68.7 | 256 | 23.4 |
| T3 | Thoracic vertebrae | 57011 | 38 | 2848 | 74 | 54051 | 28568 | 25483 | 68.7 | 253 | 22 |
| T4 | Thoracic vertebrae | 57156 | 148 | 2876 | 78 | 54054 | 28569 | 25485 | 68.8 | 249 | 22.6 |
| T5 | Thoracic vertebrae | 58341 | 1190 | 2873 | 97 | 54181 | 28635 | 25546 | 68.9 | 246 | 24.6 |
| T6 | Thoracic vertebrae | 62459 | 4121 | 2963 | 94 | 55281 | 29225 | 26056 | 70.3 | 245 | 25.5 |
| T7 | Thoracic vertebrae | 68402 | 5944 | 3183 | 121 | 59154 | 31300 | 27854 | 75.2 | 242 | 26.8 |
| T8 | Thoracic vertebrae | 74177 | 5778 | 3486 | 132 | 64781 | 34324 | 30457 | 82.4 | 239.5 | 28.8 |
| T9 | Thoracic vertebrae | 77565 | 3391 | 3783 | 127 | 70264 | 37013 | 33251 | 89.4 | 243.5 | 32.2 |
| T10 | Thoracic vertebrae | 78620 | 1056 | 3928 | 162 | 73474 | 38463 | 35011 | 93.5 | 240 | 36.1 |
| T11 | Thoracic vertebrae | 78619 | 4 | 3986 | 163 | 74466 | 38907 | 35559 | 94.7 | 228 | 39.7 |
| T12 | Thoracic vertebrae | 78615 | 0 | 4009 | 235 | 74371 | 38876 | 35495 | 94.6 | 212 | 43.6 |
| L1 | Lumbar vertebrae | 78615 | 4 | 3989 | 238 | 74384 | 38886 | 35498 | 94.6 | 216 | 49.8 |
| L2 | Lumbar vertebrae | 78614 | 4 | 4004 | 198 | 74408 | 38881 | 35527 | 94.6 | 229 | 56.6 |
| L3 | Lumbar vertebrae | 78613 | 0 | 4000 | 224 | 74389 | 38881 | 35508 | 94.6 | 235 | 63.4 |
| L4 | Lumbar vertebrae | 78617 | 4 | 3995 | 231 | 74387 | 38879 | 35508 | 94.6 | 242 | 64.9 |
| L5 | Lumbar vertebrae | 78620 | 3 | 4005 | 202 | 74410 | 38902 | 35508 | 94.6 | 244 | 65.7 |
| S1 | Sacrum | 78612 | 0 | 3990 | 155 | 74467 | 38908 | 35559 | 94.7 | 242 | 46 |
| Left rib 1 | Left ribs | 56998 | 0 | 2864 | 34 | 54100 | 28579 | 25521 | 68.8 | 285 | 9.7 |
| Left rib 2 | Left ribs | 57111 | 0 | 2894 | 29 | 54188 | 28631 | 25557 | 68.9 | 231 | 11.6 |
| Left rib 3 | Left ribs | 58539 | 0 | 2971 | 35 | 55533 | 29200 | 26333 | 70.6 | 218 | 13.7 |
| Left rib 4 | Left ribs | 64961 | 0 | 3278 | 47 | 61636 | 32094 | 29542 | 78.4 | 224 | 16.4 |
| Left rib 5 | Left ribs | 74264 | 0 | 3765 | 38 | 70461 | 36632 | 33829 | 89.6 | 225.5 | 17.3 |
| Left rib 6 | Left ribs | 78266 | 0 | 3972 | 31 | 74263 | 38749 | 35514 | 94.5 | 238.5 | 19.2 |
| Left rib 7 | Left ribs | 78479 | 0 | 3981 | 31 | 74467 | 38883 | 35584 | 94.7 | 249 | 20.1 |
| Left rib 8 | Left ribs | 78517 | 0 | 3982 | 34 | 74501 | 38911 | 35590 | 94.8 | 260 | 18.2 |
| Left rib 9 | Left ribs | 78575 | 0 | 3989 | 30 | 74556 | 38937 | 35619 | 94.8 | 274 | 17.4 |
| Left rib 10 | Left ribs | 78620 | 0 | 3955 | 30 | 74635 | 38979 | 35656 | 94.9 | 303 | 14.4 |
| Left rib 11 | Left ribs | 78617 | 0 | 3938 | 34 | 74645 | 38973 | 35672 | 94.9 | 306 | 8.8 |
| Left rib 12 | Left ribs | 76997 | 0 | 3917 | 73 | 73007 | 38542 | 34465 | 92.9 | 300 | 3.7 |
| Right rib 1 | Right ribs | 56989 | 0 | 2882 | 49 | 54058 | 28562 | 25496 | 68.8 | 284.5 | 9.8 |
| Right rib 2 | Right ribs | 57083 | 0 | 2835 | 35 | 54213 | 28622 | 25591 | 69 | 232 | 11.7 |
| Right rib 3 | Right ribs | 58449 | 0 | 2956 | 32 | 55461 | 29164 | 26297 | 70.5 | 216 | 14.2 |
| Right rib 4 | Right ribs | 64632 | 0 | 3277 | 48 | 61307 | 31936 | 29371 | 78 | 226 | 16.9 |
| Right rib 5 | Right ribs | 73777 | 0 | 3749 | 42 | 69986 | 36386 | 33600 | 89 | 229 | 18.5 |
| Right rib 6 | Right ribs | 78219 | 0 | 3963 | 33 | 74223 | 38734 | 35489 | 94.4 | 241 | 19.8 |
| Right rib 7 | Right ribs | 78473 | 0 | 3971 | 46 | 74456 | 38875 | 35581 | 94.7 | 252 | 21.1 |
| Right rib 8 | Right ribs | 78508 | 0 | 3998 | 42 | 74468 | 38895 | 35573 | 94.7 | 264 | 18.9 |
| Right rib 9 | Right ribs | 78561 | 0 | 3995 | 32 | 74534 | 38925 | 35609 | 94.8 | 278 | 17.8 |
| Right rib 10 | Right ribs | 78611 | 0 | 3961 | 31 | 74619 | 38951 | 35668 | 94.9 | 309 | 14.3 |
| Right rib 11 | Right ribs | 78616 | 0 | 3894 | 41 | 74681 | 38989 | 35692 | 95 | 311 | 8.7 |
| Right rib 12 | Right ribs | 77455 | 0 | 3851 | 69 | 73535 | 38562 | 34973 | 93.5 | 302.5 | 3.6 |

Rules are applied in the order shown. Adjacency: a vertebra requires both within-region neighbors to be segmented. Truncation: segmented volume must lie between the site- and sex-specific 5th and 99.9th percentiles. Artifact: attenuation within median ± 5 scaled median absolute deviations inside site × sex × 5-year age cells.

**Supplementary Table 2: Trajectory statistics for all 49 skeletal sites (model fit and observed distribution)**

| **Site** | **Sex** | **Region** | **n** | **Spline df** | **R²** | **Residual SD (HU)** | **Observed mean (HU)** | **Observed SD (HU)** | **Observed median (HU)** |
| --- | --- | --- | --- | --- | --- | --- | --- | --- | --- |
| C1 | Male | Cervical vertebrae | 12941 | 8 | 0.005 | 80.1665 | 450.122 | 80.3447 | 445 |
| C1 | Female | Cervical vertebrae | 13354 | 8 | 0.2519 | 73.0199 | 417.085 | 84.4011 | 409 |
| C2 | Male | Cervical vertebrae | 12939 | 8 | 0.0234 | 78.2314 | 401.186 | 79.1415 | 395 |
| C2 | Female | Cervical vertebrae | 13349 | 8 | 0.3574 | 69.2004 | 377.89 | 86.3043 | 365 |
| C3 | Male | Cervical vertebrae | 13945 | 8 | 0.0051 | 80.2437 | 401.377 | 80.4308 | 394.5 |
| C3 | Female | Cervical vertebrae | 14293 | 8 | 0.2849 | 69.1145 | 373.494 | 81.7135 | 363 |
| C4 | Male | Cervical vertebrae | 15517 | 9 | 0.0049 | 82.1653 | 421.951 | 82.3457 | 414.5 |
| C4 | Female | Cervical vertebrae | 15355 | 9 | 0.2479 | 72.467 | 395.554 | 83.5386 | 385.5 |
| C5 | Male | Cervical vertebrae | 19024 | 9 | 0.0051 | 78.0275 | 402.822 | 78.2097 | 395.5 |
| C5 | Female | Cervical vertebrae | 17881 | 9 | 0.2578 | 70.0049 | 381.898 | 81.2383 | 371 |
| C6 | Male | Cervical vertebrae | 25074 | 9 | 0.0047 | 71.299 | 373.207 | 71.4569 | 367 |
| C6 | Female | Cervical vertebrae | 22603 | 9 | 0.2741 | 63.3629 | 356.077 | 74.359 | 347 |
| C7 | Male | Cervical vertebrae | 28177 | 5 | 0.0031 | 64.1518 | 331.839 | 64.2455 | 326 |
| C7 | Female | Cervical vertebrae | 25131 | 5 | 0.2751 | 55.3401 | 313.237 | 64.9949 | 306 |
| T1 | Male | Thoracic vertebrae | 28549 | 5 | 0.0235 | 54.8087 | 284.845 | 55.4599 | 281 |
| T1 | Female | Thoracic vertebrae | 25491 | 5 | 0.3655 | 46.7548 | 275.239 | 58.6893 | 268 |
| T2 | Male | Thoracic vertebrae | 28532 | 5 | 0.0231 | 52.3744 | 265.54 | 52.9872 | 261 |
| T2 | Female | Thoracic vertebrae | 25469 | 5 | 0.3492 | 44.9292 | 256.793 | 55.6893 | 250 |
| T3 | Male | Thoracic vertebrae | 28568 | 5 | 0.0319 | 54.11 | 264.689 | 54.9913 | 260 |
| T3 | Female | Thoracic vertebrae | 25483 | 5 | 0.3681 | 45.5454 | 251.875 | 57.2903 | 245 |
| T4 | Male | Thoracic vertebrae | 28569 | 5 | 0.0366 | 53.8862 | 261.704 | 54.8961 | 257 |
| T4 | Female | Thoracic vertebrae | 25485 | 5 | 0.3777 | 44.7988 | 246.26 | 56.7867 | 238 |
| T5 | Male | Thoracic vertebrae | 28635 | 5 | 0.0334 | 52.8703 | 258.551 | 53.7709 | 254 |
| T5 | Female | Thoracic vertebrae | 25546 | 5 | 0.3795 | 43.3977 | 242.477 | 55.0892 | 235 |
| T6 | Male | Thoracic vertebrae | 29225 | 5 | 0.034 | 52.7975 | 258.565 | 53.7143 | 255 |
| T6 | Female | Thoracic vertebrae | 26056 | 5 | 0.3647 | 43.7985 | 241.608 | 54.9453 | 234 |
| T7 | Male | Thoracic vertebrae | 31300 | 5 | 0.0362 | 52.6137 | 255.82 | 53.5892 | 252 |
| T7 | Female | Thoracic vertebrae | 27854 | 5 | 0.3459 | 43.9283 | 238.533 | 54.3118 | 231 |
| T8 | Male | Thoracic vertebrae | 34324 | 5 | 0.0362 | 52.5407 | 253.119 | 53.5144 | 249 |
| T8 | Female | Thoracic vertebrae | 30457 | 5 | 0.3408 | 44.244 | 234.993 | 54.4893 | 228 |
| T9 | Male | Thoracic vertebrae | 37013 | 10 | 0.0361 | 52.9938 | 256.483 | 53.9698 | 252.5 |
| T9 | Female | Thoracic vertebrae | 33251 | 10 | 0.3505 | 44.9226 | 239.577 | 55.7326 | 232 |
| T10 | Male | Thoracic vertebrae | 38463 | 10 | 0.04 | 52.1576 | 251.976 | 53.2277 | 248 |
| T10 | Female | Thoracic vertebrae | 35011 | 10 | 0.3647 | 44.4345 | 238.039 | 55.7407 | 230 |
| T11 | Male | Thoracic vertebrae | 38907 | 10 | 0.052 | 50.3559 | 238.556 | 51.7123 | 234.5 |
| T11 | Female | Thoracic vertebrae | 35559 | 10 | 0.3692 | 43.6656 | 227.747 | 54.9713 | 220 |
| T12 | Male | Thoracic vertebrae | 38876 | 10 | 0.0696 | 46.8452 | 221.397 | 48.5597 | 217 |
| T12 | Female | Thoracic vertebrae | 35495 | 10 | 0.3752 | 41.3503 | 214.132 | 52.3066 | 207 |
| L1 | Male | Lumbar vertebrae | 38886 | 10 | 0.0524 | 47.3729 | 225.505 | 48.6599 | 221 |
| L1 | Female | Lumbar vertebrae | 35498 | 10 | 0.3522 | 41.211 | 217.694 | 51.1948 | 210.5 |
| L2 | Male | Lumbar vertebrae | 38881 | 10 | 0.0353 | 50.1689 | 240.333 | 51.0735 | 235 |
| L2 | Female | Lumbar vertebrae | 35527 | 10 | 0.3348 | 43.2671 | 230.002 | 53.0414 | 222.5 |
| L3 | Male | Lumbar vertebrae | 38881 | 10 | 0.0385 | 51.0636 | 246.958 | 52.0695 | 242 |
| L3 | Female | Lumbar vertebrae | 35508 | 10 | 0.3664 | 43.0815 | 233.565 | 54.1143 | 226 |
| L4 | Male | Lumbar vertebrae | 38879 | 10 | 0.0254 | 54.2515 | 256.017 | 54.9479 | 250 |
| L4 | Female | Lumbar vertebrae | 35508 | 10 | 0.3282 | 45.7379 | 239.691 | 55.7966 | 232 |
| L5 | Male | Lumbar vertebrae | 38902 | 10 | 0.0182 | 59.3686 | 259.908 | 59.9086 | 253 |
| L5 | Female | Lumbar vertebrae | 35508 | 10 | 0.2959 | 48.9562 | 240.664 | 58.337 | 233 |
| S1 | Male | Sacrum | 38908 | 10 | 0.0907 | 56.9395 | 252.687 | 59.7035 | 248 |
| S1 | Female | Sacrum | 35559 | 10 | 0.4299 | 49.7842 | 242.804 | 65.9239 | 235 |
| Left rib 1 | Male | Left ribs | 28579 | 9 | 0.1134 | 63.1206 | 297.049 | 67.0277 | 290 |
| Left rib 1 | Female | Left ribs | 25521 | 9 | 0.4345 | 52.6704 | 288.231 | 70.0292 | 279 |
| Left rib 2 | Male | Left ribs | 28631 | 5 | 0.0878 | 55.6192 | 237.317 | 58.2305 | 232 |
| Left rib 2 | Female | Left ribs | 25557 | 5 | 0.4528 | 50.2898 | 241.059 | 67.9801 | 229 |
| Left rib 3 | Male | Left ribs | 29200 | 5 | 0.1649 | 54.5553 | 221.131 | 59.6947 | 214 |
| Left rib 3 | Female | Left ribs | 26333 | 5 | 0.493 | 53.0762 | 236.998 | 74.5356 | 222 |
| Left rib 4 | Male | Left ribs | 32094 | 10 | 0.1463 | 59.9936 | 229.389 | 64.9201 | 222.5 |
| Left rib 4 | Female | Left ribs | 29542 | 10 | 0.4135 | 61.1749 | 241.117 | 79.8689 | 226 |
| Left rib 5 | Male | Left ribs | 36632 | 10 | 0.1101 | 63.8348 | 230.22 | 67.6619 | 224 |
| Left rib 5 | Female | Left ribs | 33829 | 10 | 0.3417 | 67.9867 | 241.565 | 83.7858 | 227 |
| Left rib 6 | Male | Left ribs | 38749 | 9 | 0.0828 | 67.1406 | 241.694 | 70.0996 | 236 |
| Left rib 6 | Female | Left ribs | 35514 | 9 | 0.3129 | 71.3838 | 254.767 | 86.1078 | 242.5 |
| Left rib 7 | Male | Left ribs | 38883 | 10 | 0.1122 | 64.9583 | 249.51 | 68.9335 | 242.5 |
| Left rib 7 | Female | Left ribs | 35584 | 10 | 0.3664 | 68.0572 | 271.013 | 85.4887 | 257.5 |
| Left rib 8 | Male | Left ribs | 38911 | 9 | 0.1432 | 67.6336 | 263.356 | 73.0612 | 255.5 |
| Left rib 8 | Female | Left ribs | 35590 | 9 | 0.4407 | 67.4598 | 282.524 | 90.1945 | 266 |
| Left rib 9 | Male | Left ribs | 38937 | 9 | 0.1147 | 72.3087 | 282.986 | 76.8406 | 274.5 |
| Left rib 9 | Female | Left ribs | 35619 | 9 | 0.4419 | 67.9118 | 291.391 | 90.8958 | 274 |
| Left rib 10 | Male | Left ribs | 38979 | 9 | 0.0945 | 74.4538 | 310.1 | 78.2364 | 302 |
| Left rib 10 | Female | Left ribs | 35656 | 9 | 0.4505 | 67.8632 | 321.193 | 91.5371 | 304 |
| Left rib 11 | Male | Left ribs | 38973 | 10 | 0.1373 | 73.8922 | 316.088 | 79.5438 | 307 |
| Left rib 11 | Female | Left ribs | 35672 | 10 | 0.4997 | 69.046 | 323.219 | 97.5994 | 304 |
| Left rib 12 | Male | Left ribs | 38542 | 5 | 0.1447 | 73.0581 | 304.888 | 78.9927 | 295 |
| Left rib 12 | Female | Left ribs | 34465 | 5 | 0.4177 | 74.9902 | 324.061 | 98.2674 | 306 |
| Right rib 1 | Male | Right ribs | 28562 | 8 | 0.1194 | 63.0494 | 297.025 | 67.1801 | 291 |
| Right rib 1 | Female | Right ribs | 25496 | 8 | 0.4605 | 51.5433 | 287.099 | 70.1633 | 277 |
| Right rib 2 | Male | Right ribs | 28622 | 5 | 0.0886 | 55.8655 | 238.092 | 58.5141 | 233 |
| Right rib 2 | Female | Right ribs | 25591 | 5 | 0.4563 | 50.9461 | 242.545 | 69.0888 | 231 |
| Right rib 3 | Male | Right ribs | 29164 | 5 | 0.1752 | 53.6626 | 217.763 | 59.084 | 211 |
| Right rib 3 | Female | Right ribs | 26297 | 5 | 0.4976 | 53.1511 | 236.96 | 74.9838 | 222 |
| Right rib 4 | Male | Right ribs | 31936 | 9 | 0.1489 | 59.0679 | 228.211 | 64.0182 | 222 |
| Right rib 4 | Female | Right ribs | 29371 | 9 | 0.4019 | 61.9972 | 246.122 | 80.1516 | 231 |
| Right rib 5 | Male | Right ribs | 36386 | 9 | 0.0961 | 63.1457 | 231.548 | 66.4088 | 226 |
| Right rib 5 | Female | Right ribs | 33600 | 9 | 0.3059 | 70.1532 | 245.264 | 84.1941 | 233 |
| Right rib 6 | Male | Right ribs | 38734 | 9 | 0.0855 | 66.771 | 242.658 | 69.8149 | 236 |
| Right rib 6 | Female | Right ribs | 35489 | 9 | 0.3252 | 71.0098 | 259.696 | 86.4327 | 246.5 |
| Right rib 7 | Male | Right ribs | 38875 | 9 | 0.112 | 64.752 | 252.017 | 68.7078 | 245 |
| Right rib 7 | Female | Right ribs | 35581 | 9 | 0.3656 | 67.8563 | 275.052 | 85.1817 | 262 |
| Right rib 8 | Male | Right ribs | 38895 | 9 | 0.1359 | 66.4804 | 264.046 | 71.5116 | 256 |
| Right rib 8 | Female | Right ribs | 35573 | 9 | 0.4114 | 67.0038 | 287.123 | 87.3248 | 272 |
| Right rib 9 | Male | Right ribs | 38925 | 9 | 0.1023 | 70.971 | 282.847 | 74.8967 | 274.5 |
| Right rib 9 | Female | Right ribs | 35609 | 9 | 0.4054 | 67.5699 | 296.715 | 87.619 | 281 |
| Right rib 10 | Male | Right ribs | 38951 | 9 | 0.0785 | 73.229 | 313.777 | 76.2775 | 306 |
| Right rib 10 | Female | Right ribs | 35668 | 9 | 0.4166 | 67.9383 | 328.407 | 88.9388 | 313 |
| Right rib 11 | Male | Right ribs | 38989 | 9 | 0.1229 | 72.8642 | 320.586 | 77.7938 | 312 |
| Right rib 11 | Female | Right ribs | 35692 | 9 | 0.4782 | 68.8979 | 327.471 | 95.3688 | 310 |
| Right rib 12 | Male | Right ribs | 38562 | 5 | 0.1175 | 73.954 | 308.146 | 78.7194 | 298.5 |
| Right rib 12 | Female | Right ribs | 34973 | 5 | 0.3803 | 76.1142 | 323.995 | 96.6863 | 307.5 |

**Supplementary Table 2 (continued): Trajectory statistics for all 49 skeletal sites (fitted attenuation by age)**

| **Site** | **Sex** | **Attenuation at 20 years, HU (95% CI)** | **Attenuation at 30 years, HU (95% CI)** | **Attenuation at 40 years, HU (95% CI)** | **Attenuation at 50 years, HU (95% CI)** | **Attenuation at 60 years, HU (95% CI)** | **Attenuation at 70 years, HU (95% CI)** | **Attenuation at 80 years, HU (95% CI)** | **Attenuation at 90 years, HU (95% CI)** |
| --- | --- | --- | --- | --- | --- | --- | --- | --- | --- |
| C1 | Male | 465 (455 to 475) | 460 (454 to 466) | 452 (448 to 456) | 446 (443 to 450) | 447 (444 to 450) | 446 (443 to 449) | 455 (452 to 458) | 461 (452 to 471) |
| C1 | Female | 511 (501 to 521) | 497 (492 to 503) | 492 (488 to 495) | 469 (466 to 472) | 417 (414 to 420) | 390 (387 to 393) | 376 (373 to 379) | 373 (365 to 381) |
| C2 | Male | 445 (434 to 455) | 433 (427 to 439) | 424 (420 to 429) | 416 (412 to 419) | 404 (401 to 407) | 394 (391 to 397) | 393 (390 to 396) | 393 (383 to 402) |
| C2 | Female | 489 (479 to 499) | 476 (471 to 481) | 469 (465 to 473) | 442 (438 to 445) | 379 (377 to 382) | 347 (344 to 350) | 327 (324 to 329) | 317 (310 to 324) |
| C3 | Male | 437 (427 to 446) | 416 (411 to 422) | 407 (403 to 411) | 403 (399 to 406) | 398 (395 to 401) | 399 (396 to 402) | 401 (397 to 404) | 404 (395 to 412) |
| C3 | Female | 477 (467 to 487) | 459 (454 to 464) | 451 (447 to 454) | 427 (424 to 430) | 374 (371 to 376) | 347 (345 to 350) | 331 (328 to 333) | 329 (322 to 335) |
| C4 | Male | 455 (446 to 465) | 433 (428 to 439) | 422 (418 to 426) | 417 (414 to 420) | 417 (413 to 420) | 421 (418 to 424) | 424 (421 to 428) | 423 (414 to 431) |
| C4 | Female | 500 (490 to 510) | 478 (472 to 483) | 473 (470 to 477) | 448 (445 to 452) | 391 (388 to 394) | 371 (368 to 373) | 359 (356 to 363) | 356 (350 to 363) |
| C5 | Male | 440 (431 to 449) | 417 (412 to 422) | 405 (402 to 408) | 399 (396 to 402) | 398 (396 to 401) | 403 (401 to 406) | 404 (401 to 407) | 393 (386 to 400) |
| C5 | Female | 484 (474 to 493) | 464 (459 to 469) | 459 (456 to 463) | 436 (433 to 439) | 382 (379 to 384) | 359 (356 to 361) | 346 (343 to 349) | 340 (334 to 345) |
| C6 | Male | 407 (400 to 413) | 385 (381 to 389) | 374 (371 to 376) | 370 (368 to 373) | 372 (370 to 374) | 374 (372 to 376) | 375 (372 to 377) | 361 (355 to 366) |
| C6 | Female | 447 (439 to 455) | 434 (430 to 438) | 428 (426 to 431) | 407 (405 to 409) | 359 (357 to 361) | 335 (333 to 337) | 322 (320 to 325) | 313 (308 to 317) |
| C7 | Male | 354 (349 to 359) | 339 (337 to 341) | 332 (330 to 334) | 331 (329 to 332) | 332 (331 to 333) | 333 (332 to 334) | 331 (330 to 333) | 320 (316 to 324) |
| C7 | Female | 376 (370 to 382) | 392 (390 to 395) | 381 (379 to 383) | 354 (352 to 355) | 321 (320 to 322) | 295 (294 to 296) | 283 (282 to 284) | 272 (269 to 275) |
| T1 | Male | 320 (314 to 325) | 310 (308 to 313) | 302 (300 to 304) | 294 (293 to 296) | 288 (287 to 289) | 284 (283 to 284) | 279 (278 to 280) | 267 (264 to 270) |
| T1 | Female | 344 (339 to 350) | 360 (358 to 362) | 347 (345 to 348) | 317 (316 to 318) | 283 (282 to 284) | 256 (255 to 257) | 244 (243 to 245) | 232 (230 to 235) |
| T2 | Male | 307 (302 to 312) | 292 (290 to 294) | 281 (279 to 283) | 273 (272 to 274) | 268 (267 to 269) | 264 (263 to 265) | 260 (259 to 262) | 251 (248 to 255) |
| T2 | Female | 328 (323 to 332) | 336 (334 to 338) | 322 (320 to 323) | 294 (293 to 295) | 264 (263 to 265) | 240 (239 to 240) | 228 (227 to 229) | 216 (214 to 219) |
| T3 | Male | 309 (304 to 314) | 295 (293 to 297) | 284 (282 to 285) | 275 (274 to 276) | 268 (267 to 269) | 263 (262 to 264) | 258 (257 to 259) | 246 (243 to 250) |
| T3 | Female | 327 (323 to 332) | 335 (333 to 337) | 320 (318 to 321) | 291 (290 to 292) | 259 (258 to 260) | 234 (233 to 235) | 221 (220 to 222) | 208 (206 to 210) |
| T4 | Male | 308 (303 to 313) | 294 (291 to 296) | 282 (280 to 284) | 273 (272 to 274) | 266 (265 to 267) | 260 (259 to 261) | 254 (253 to 256) | 242 (238 to 245) |
| T4 | Female | 321 (316 to 326) | 331 (329 to 333) | 315 (314 to 317) | 286 (285 to 287) | 253 (252 to 254) | 228 (227 to 229) | 216 (215 to 217) | 203 (201 to 205) |
| T5 | Male | 303 (299 to 308) | 289 (287 to 291) | 277 (276 to 279) | 268 (267 to 270) | 262 (261 to 263) | 257 (256 to 258) | 252 (251 to 253) | 238 (235 to 241) |
| T5 | Female | 318 (314 to 323) | 326 (324 to 328) | 310 (308 to 311) | 281 (280 to 282) | 249 (248 to 250) | 225 (224 to 226) | 213 (212 to 214) | 200 (198 to 203) |
| T6 | Male | 302 (298 to 307) | 288 (286 to 290) | 277 (275 to 278) | 268 (267 to 269) | 262 (261 to 263) | 257 (256 to 258) | 252 (251 to 253) | 237 (234 to 241) |
| T6 | Female | 316 (312 to 320) | 322 (320 to 324) | 306 (304 to 308) | 278 (277 to 279) | 247 (246 to 248) | 224 (223 to 225) | 213 (212 to 214) | 200 (198 to 202) |
| T7 | Male | 301 (297 to 305) | 285 (283 to 286) | 273 (271 to 274) | 264 (263 to 265) | 259 (258 to 260) | 254 (254 to 255) | 249 (248 to 250) | 234 (231 to 238) |
| T7 | Female | 308 (304 to 312) | 316 (314 to 317) | 300 (299 to 302) | 272 (271 to 273) | 242 (241 to 243) | 221 (220 to 222) | 212 (211 to 213) | 199 (197 to 201) |
| T8 | Male | 299 (295 to 302) | 281 (280 to 283) | 269 (268 to 271) | 261 (260 to 262) | 255 (255 to 256) | 252 (251 to 252) | 247 (246 to 248) | 232 (229 to 235) |
| T8 | Female | 306 (302 to 310) | 312 (310 to 314) | 296 (294 to 297) | 267 (266 to 268) | 237 (237 to 238) | 217 (216 to 217) | 209 (208 to 210) | 197 (195 to 199) |
| T9 | Male | 306 (301 to 310) | 286 (283 to 288) | 272 (271 to 274) | 264 (262 to 265) | 258 (257 to 260) | 255 (254 to 256) | 250 (248 to 251) | 231 (228 to 235) |
| T9 | Female | 321 (317 to 326) | 311 (308 to 313) | 302 (300 to 303) | 276 (274 to 277) | 238 (236 to 239) | 220 (219 to 221) | 211 (210 to 213) | 201 (198 to 204) |
| T10 | Male | 309 (304 to 315) | 285 (283 to 288) | 270 (269 to 272) | 260 (259 to 261) | 253 (252 to 254) | 250 (249 to 251) | 245 (244 to 247) | 230 (226 to 233) |
| T10 | Female | 324 (320 to 329) | 312 (310 to 314) | 302 (301 to 304) | 275 (273 to 276) | 236 (235 to 237) | 218 (216 to 219) | 209 (208 to 210) | 201 (198 to 203) |
| T11 | Male | 301 (296 to 305) | 277 (275 to 280) | 260 (259 to 262) | 247 (246 to 248) | 240 (238 to 241) | 235 (233 to 236) | 232 (230 to 233) | 220 (216 to 223) |
| T11 | Female | 315 (310 to 319) | 301 (299 to 304) | 291 (290 to 292) | 263 (262 to 265) | 225 (224 to 226) | 207 (206 to 208) | 199 (198 to 200) | 193 (190 to 195) |
| T12 | Male | 290 (285 to 294) | 264 (262 to 266) | 245 (244 to 247) | 231 (230 to 232) | 222 (221 to 223) | 217 (216 to 218) | 214 (213 to 215) | 208 (204 to 211) |
| T12 | Female | 302 (298 to 306) | 287 (285 to 289) | 275 (274 to 276) | 247 (246 to 248) | 210 (209 to 211) | 193 (192 to 194) | 188 (187 to 189) | 186 (183 to 189) |
| L1 | Male | 289 (285 to 294) | 265 (263 to 267) | 246 (245 to 248) | 232 (231 to 233) | 224 (223 to 225) | 222 (220 to 223) | 221 (219 to 222) | 214 (210 to 217) |
| L1 | Female | 306 (302 to 310) | 290 (288 to 292) | 276 (275 to 278) | 248 (247 to 249) | 212 (211 to 213) | 197 (196 to 198) | 194 (193 to 196) | 192 (189 to 195) |
| L2 | Male | 300 (295 to 305) | 277 (275 to 279) | 257 (256 to 259) | 243 (241 to 244) | 237 (236 to 238) | 238 (237 to 239) | 238 (236 to 239) | 229 (225 to 233) |
| L2 | Female | 323 (318 to 327) | 306 (304 to 308) | 290 (289 to 291) | 260 (259 to 261) | 224 (223 to 225) | 211 (210 to 212) | 207 (205 to 208) | 201 (198 to 204) |
| L3 | Male | 309 (304 to 314) | 286 (283 to 288) | 265 (263 to 266) | 250 (248 to 251) | 244 (243 to 245) | 245 (243 to 246) | 244 (243 to 246) | 235 (231 to 239) |
| L3 | Female | 329 (325 to 334) | 313 (311 to 315) | 297 (295 to 298) | 266 (265 to 267) | 229 (228 to 230) | 214 (213 to 215) | 208 (206 to 209) | 201 (198 to 204) |
| L4 | Male | 311 (306 to 316) | 289 (287 to 292) | 269 (268 to 271) | 255 (253 to 256) | 251 (250 to 253) | 255 (254 to 257) | 255 (253 to 257) | 242 (238 to 246) |
| L4 | Female | 328 (324 to 332) | 316 (314 to 318) | 302 (301 to 304) | 272 (271 to 274) | 235 (234 to 236) | 220 (219 to 222) | 214 (212 to 215) | 207 (203 to 210) |
| L5 | Male | 309 (304 to 314) | 290 (287 to 292) | 270 (268 to 272) | 257 (256 to 258) | 256 (255 to 258) | 261 (259 to 262) | 258 (256 to 260) | 242 (238 to 246) |
| L5 | Female | 325 (320 to 329) | 315 (312 to 317) | 303 (301 to 304) | 274 (273 to 275) | 236 (235 to 237) | 223 (221 to 224) | 215 (213 to 216) | 202 (199 to 205) |
| S1 | Male | 317 (312 to 322) | 302 (300 to 305) | 288 (286 to 289) | 272 (271 to 274) | 259 (257 to 260) | 248 (247 to 250) | 237 (235 to 239) | 221 (216 to 225) |
| S1 | Female | 336 (331 to 340) | 328 (325 to 331) | 321 (320 to 323) | 292 (291 to 294) | 246 (245 to 247) | 219 (218 to 221) | 202 (200 to 203) | 182 (179 to 185) |
| Left rib 1 | Male | 365 (358 to 373) | 356 (353 to 360) | 344 (342 to 347) | 329 (327 to 331) | 309 (307 to 311) | 292 (290 to 294) | 278 (276 to 280) | 261 (256 to 266) |
| Left rib 1 | Female | 395 (388 to 403) | 383 (379 to 386) | 372 (370 to 375) | 346 (345 to 348) | 296 (294 to 298) | 266 (264 to 268) | 245 (244 to 247) | 231 (228 to 234) |
| Left rib 2 | Male | 342 (337 to 347) | 295 (293 to 298) | 264 (262 to 266) | 245 (244 to 246) | 235 (234 to 236) | 232 (231 to 233) | 230 (229 to 232) | 225 (222 to 229) |
| Left rib 2 | Female | 385 (380 to 391) | 361 (359 to 364) | 324 (322 to 326) | 281 (280 to 283) | 243 (242 to 244) | 216 (215 to 217) | 204 (203 to 205) | 192 (189 to 194) |
| Left rib 3 | Male | 359 (353 to 364) | 301 (299 to 304) | 261 (259 to 263) | 234 (233 to 236) | 219 (218 to 220) | 213 (212 to 213) | 210 (209 to 212) | 203 (200 to 207) |
| Left rib 3 | Female | 413 (407 to 419) | 381 (378 to 384) | 334 (331 to 336) | 281 (280 to 283) | 235 (234 to 236) | 207 (206 to 208) | 198 (197 to 199) | 187 (184 to 189) |
| Left rib 4 | Male | 371 (364 to 377) | 308 (304 to 312) | 266 (264 to 269) | 241 (239 to 243) | 226 (224 to 228) | 221 (220 to 223) | 219 (218 to 221) | 209 (205 to 213) |
| Left rib 4 | Female | 423 (414 to 431) | 369 (364 to 373) | 334 (332 to 337) | 287 (285 to 290) | 232 (231 to 234) | 212 (210 to 214) | 202 (200 to 203) | 196 (193 to 200) |
| Left rib 5 | Male | 361 (354 to 368) | 298 (294 to 302) | 261 (258 to 263) | 238 (236 to 240) | 225 (223 to 226) | 223 (221 to 224) | 223 (221 to 225) | 214 (209 to 218) |
| Left rib 5 | Female | 409 (401 to 418) | 356 (352 to 361) | 328 (325 to 330) | 286 (284 to 288) | 233 (231 to 234) | 213 (212 to 215) | 203 (201 to 205) | 197 (194 to 201) |
| Left rib 6 | Male | 359 (353 to 366) | 301 (298 to 305) | 269 (267 to 272) | 250 (248 to 252) | 237 (235 to 238) | 235 (233 to 236) | 234 (232 to 236) | 226 (222 to 231) |
| Left rib 6 | Female | 412 (403 to 420) | 364 (360 to 368) | 337 (334 to 340) | 300 (298 to 302) | 248 (246 to 249) | 228 (227 to 230) | 215 (213 to 217) | 211 (207 to 215) |
| Left rib 7 | Male | 382 (375 to 388) | 317 (314 to 321) | 282 (280 to 284) | 261 (259 to 262) | 247 (245 to 248) | 243 (241 to 244) | 239 (237 to 241) | 231 (227 to 236) |
| Left rib 7 | Female | 445 (438 to 453) | 389 (385 to 393) | 361 (359 to 364) | 317 (315 to 319) | 262 (261 to 264) | 240 (238 to 242) | 229 (227 to 231) | 225 (222 to 229) |
| Left rib 8 | Male | 413 (407 to 419) | 346 (342 to 350) | 303 (301 to 306) | 278 (276 to 280) | 263 (261 to 265) | 256 (255 to 258) | 247 (245 to 249) | 236 (232 to 241) |
| Left rib 8 | Female | 484 (477 to 492) | 427 (423 to 431) | 384 (382 to 386) | 333 (331 to 335) | 272 (270 to 274) | 251 (249 to 252) | 234 (232 to 236) | 225 (221 to 229) |
| Left rib 9 | Male | 423 (417 to 429) | 362 (358 to 365) | 321 (319 to 324) | 297 (295 to 299) | 284 (283 to 286) | 277 (275 to 279) | 266 (264 to 268) | 252 (247 to 257) |
| Left rib 9 | Female | 487 (480 to 494) | 436 (432 to 439) | 395 (393 to 397) | 344 (342 to 346) | 283 (281 to 284) | 260 (258 to 261) | 241 (239 to 243) | 229 (226 to 233) |
| Left rib 10 | Male | 448 (442 to 455) | 388 (384 to 391) | 348 (345 to 350) | 323 (321 to 325) | 310 (308 to 311) | 303 (302 to 305) | 297 (295 to 299) | 286 (281 to 292) |
| Left rib 10 | Female | 510 (504 to 517) | 467 (463 to 470) | 429 (426 to 431) | 377 (375 to 379) | 313 (311 to 315) | 287 (285 to 289) | 269 (268 to 271) | 260 (256 to 263) |
| Left rib 11 | Male | 471 (464 to 478) | 409 (405 to 413) | 366 (363 to 368) | 336 (334 to 338) | 317 (316 to 319) | 308 (306 to 310) | 298 (296 to 300) | 285 (280 to 291) |
| Left rib 11 | Female | 546 (538 to 553) | 484 (480 to 488) | 443 (441 to 446) | 384 (382 to 385) | 314 (313 to 316) | 283 (281 to 285) | 264 (263 to 266) | 258 (254 to 262) |
| Left rib 12 | Male | 458 (452 to 464) | 395 (393 to 398) | 351 (349 to 354) | 323 (321 to 324) | 305 (304 to 306) | 294 (293 to 295) | 287 (286 to 289) | 283 (279 to 287) |
| Left rib 12 | Female | 512 (505 to 519) | 485 (482 to 488) | 434 (432 to 437) | 375 (373 to 376) | 320 (318 to 321) | 284 (283 to 285) | 273 (271 to 274) | 269 (266 to 273) |
| Right rib 1 | Male | 377 (368 to 385) | 359 (356 to 362) | 345 (342 to 347) | 329 (327 to 331) | 310 (308 to 312) | 292 (291 to 294) | 277 (275 to 278) | 259 (254 to 263) |
| Right rib 1 | Female | 402 (394 to 410) | 390 (387 to 393) | 374 (371 to 376) | 345 (343 to 347) | 297 (296 to 299) | 263 (261 to 264) | 242 (241 to 244) | 228 (225 to 232) |
| Right rib 2 | Male | 345 (340 to 350) | 297 (295 to 299) | 265 (263 to 267) | 246 (245 to 247) | 236 (235 to 237) | 233 (232 to 234) | 231 (230 to 232) | 224 (221 to 228) |
| Right rib 2 | Female | 392 (386 to 398) | 366 (364 to 369) | 327 (325 to 329) | 284 (282 to 285) | 244 (243 to 245) | 217 (216 to 218) | 205 (203 to 206) | 194 (191 to 196) |
| Right rib 3 | Male | 360 (354 to 365) | 300 (298 to 303) | 258 (256 to 260) | 231 (230 to 232) | 216 (215 to 217) | 209 (208 to 210) | 207 (206 to 208) | 199 (196 to 202) |
| Right rib 3 | Female | 418 (412 to 425) | 383 (381 to 386) | 334 (332 to 336) | 281 (280 to 282) | 235 (234 to 236) | 207 (206 to 207) | 198 (197 to 199) | 186 (184 to 189) |
| Right rib 4 | Male | 369 (362 to 376) | 306 (302 to 310) | 265 (263 to 268) | 239 (237 to 241) | 223 (221 to 225) | 220 (218 to 221) | 219 (217 to 221) | 208 (204 to 212) |
| Right rib 4 | Female | 424 (415 to 432) | 377 (372 to 381) | 337 (335 to 340) | 291 (289 to 293) | 236 (234 to 238) | 219 (217 to 220) | 209 (207 to 210) | 203 (199 to 206) |
| Right rib 5 | Male | 351 (345 to 358) | 295 (291 to 298) | 259 (257 to 262) | 238 (236 to 240) | 225 (223 to 227) | 224 (222 to 225) | 226 (224 to 228) | 218 (214 to 222) |
| Right rib 5 | Female | 405 (396 to 413) | 354 (349 to 358) | 325 (322 to 328) | 288 (286 to 291) | 236 (234 to 238) | 220 (218 to 221) | 209 (207 to 211) | 203 (200 to 207) |
| Right rib 6 | Male | 360 (354 to 367) | 303 (300 to 307) | 271 (269 to 273) | 251 (249 to 253) | 237 (235 to 239) | 235 (234 to 237) | 236 (234 to 238) | 226 (222 to 231) |
| Right rib 6 | Female | 420 (412 to 428) | 369 (365 to 374) | 343 (340 to 346) | 307 (305 to 309) | 253 (251 to 254) | 233 (231 to 234) | 219 (217 to 221) | 211 (208 to 215) |
| Right rib 7 | Male | 381 (375 to 387) | 320 (316 to 323) | 285 (282 to 287) | 263 (262 to 265) | 249 (248 to 251) | 245 (244 to 247) | 240 (238 to 242) | 232 (228 to 236) |
| Right rib 7 | Female | 444 (437 to 451) | 393 (389 to 397) | 362 (360 to 365) | 323 (321 to 325) | 267 (265 to 269) | 247 (245 to 248) | 233 (231 to 235) | 226 (223 to 230) |
| Right rib 8 | Male | 409 (403 to 415) | 343 (340 to 347) | 302 (300 to 304) | 277 (275 to 279) | 264 (262 to 266) | 257 (256 to 259) | 248 (246 to 250) | 240 (235 to 244) |
| Right rib 8 | Female | 472 (465 to 479) | 421 (417 to 424) | 382 (380 to 385) | 335 (333 to 337) | 279 (277 to 280) | 258 (256 to 259) | 242 (240 to 243) | 231 (227 to 234) |
| Right rib 9 | Male | 418 (412 to 424) | 356 (353 to 360) | 317 (315 to 319) | 294 (292 to 296) | 282 (281 to 284) | 277 (275 to 278) | 269 (267 to 271) | 256 (251 to 261) |
| Right rib 9 | Female | 481 (474 to 487) | 431 (428 to 435) | 393 (390 to 395) | 344 (342 to 346) | 287 (285 to 289) | 268 (266 to 269) | 252 (250 to 254) | 237 (234 to 241) |
| Right rib 10 | Male | 442 (436 to 449) | 384 (380 to 387) | 346 (344 to 348) | 323 (321 to 325) | 311 (310 to 313) | 307 (305 to 308) | 304 (302 to 306) | 295 (290 to 301) |
| Right rib 10 | Female | 509 (502 to 515) | 466 (462 to 469) | 429 (427 to 432) | 380 (378 to 382) | 319 (317 to 321) | 298 (296 to 299) | 281 (279 to 283) | 270 (266 to 274) |
| Right rib 11 | Male | 470 (463 to 476) | 407 (404 to 411) | 365 (363 to 367) | 337 (335 to 339) | 319 (317 to 321) | 312 (310 to 313) | 305 (303 to 307) | 297 (292 to 303) |
| Right rib 11 | Female | 540 (533 to 548) | 485 (481 to 489) | 441 (439 to 444) | 385 (383 to 387) | 317 (315 to 319) | 291 (289 to 292) | 272 (271 to 274) | 268 (264 to 272) |
| Right rib 12 | Male | 454 (448 to 459) | 391 (388 to 394) | 347 (345 to 350) | 320 (318 to 321) | 304 (303 to 306) | 298 (297 to 299) | 296 (294 to 297) | 294 (290 to 298) |
| Right rib 12 | Female | 503 (496 to 510) | 476 (473 to 479) | 428 (425 to 430) | 371 (369 to 372) | 318 (317 to 320) | 285 (284 to 286) | 276 (275 to 278) | 278 (274 to 282) |

**Supplementary Table 2 (continued): Trajectory statistics for all 49 skeletal sites (rate of change)**

| **Site** | **Sex** | **Change 30–40 years, HU/year (95% CI)** | **Change 40–50 years, HU/year (95% CI)** | **Change 50–60 years, HU/year (95% CI)** | **Change 60–70 years, HU/year (95% CI)** | **Change 70–80 years, HU/year (95% CI)** | **Change 45–60 years, HU/year (95% CI)** | **Change 40 to 80 years, HU (95% CI)** | **Change 40 to 80 years (%)** |
| --- | --- | --- | --- | --- | --- | --- | --- | --- | --- |
| C1 | Male | -0.81 (-1.24 to -0.39) | -0.57 (-1.01 to -0.12) | 0.05 (-0.43 to 0.53) | -0.06 (-0.56 to 0.45) | 0.88 (0.37 to 1.39) | -0.11 (-0.46 to 0.24) | 3 (-2 to 8) | 0.6828 |
| C1 | Female | -0.57 (-0.96 to -0.19) | -2.24 (-2.63 to -1.85) | -5.24 (-5.67 to -4.81) | -2.65 (-3.11 to -2.19) | -1.46 (-1.93 to -0.99) | -4.48 (-4.79 to -4.17) | -116 (-121 to -111) | -23.5781 |
| C2 | Male | -0.84 (-1.25 to -0.43) | -0.88 (-1.33 to -0.44) | -1.20 (-1.68 to -0.72) | -0.99 (-1.48 to -0.50) | -0.07 (-0.57 to 0.42) | -1.11 (-1.46 to -0.76) | -31 (-37 to -26) | -7.4102 |
| C2 | Female | -0.70 (-1.09 to -0.32) | -2.75 (-3.15 to -2.36) | -6.22 (-6.64 to -5.80) | -3.24 (-3.66 to -2.82) | -2.02 (-2.44 to -1.59) | -5.35 (-5.65 to -5.05) | -142 (-147 to -138) | -30.3355 |
| C3 | Male | -0.91 (-1.29 to -0.53) | -0.45 (-0.86 to -0.03) | -0.49 (-0.94 to -0.04) | 0.14 (-0.32 to 0.60) | 0.15 (-0.36 to 0.66) | -0.46 (-0.79 to -0.14) | -6 (-12 to -1) | -1.589 |
| C3 | Female | -0.85 (-1.21 to -0.49) | -2.32 (-2.70 to -1.94) | -5.36 (-5.75 to -4.97) | -2.67 (-3.07 to -2.28) | -1.63 (-2.06 to -1.20) | -4.57 (-4.85 to -4.29) | -120 (-124 to -115) | -26.5801 |
| C4 | Male | -1.15 (-1.59 to -0.71) | -0.47 (-0.88 to -0.07) | -0.04 (-0.54 to 0.45) | 0.44 (-0.02 to 0.91) | 0.35 (-0.16 to 0.85) | -0.15 (-0.52 to 0.23) | 3 (-3 to 8) | 0.6413 |
| C4 | Female | -0.43 (-0.86 to -0.01) | -2.48 (-2.86 to -2.11) | -5.79 (-6.24 to -5.33) | -2.00 (-2.42 to -1.59) | -1.13 (-1.57 to -0.69) | -5.02 (-5.36 to -4.68) | -114 (-119 to -109) | -24.085 |
| C5 | Male | -1.21 (-1.59 to -0.83) | -0.56 (-0.92 to -0.20) | -0.09 (-0.52 to 0.34) | 0.50 (0.07 to 0.92) | 0.06 (-0.36 to 0.49) | -0.21 (-0.53 to 0.11) | -1 (-6 to 4) | -0.2256 |
| C5 | Female | -0.47 (-0.86 to -0.09) | -2.30 (-2.65 to -1.95) | -5.47 (-5.88 to -5.05) | -2.28 (-2.68 to -1.88) | -1.28 (-1.67 to -0.89) | -4.70 (-5.01 to -4.40) | -113 (-118 to -109) | -24.6517 |
| C6 | Male | -1.10 (-1.39 to -0.82) | -0.34 (-0.62 to -0.06) | 0.16 (-0.19 to 0.50) | 0.21 (-0.13 to 0.56) | 0.07 (-0.27 to 0.41) | 0.04 (-0.21 to 0.30) | 1 (-3 to 5) | 0.2799 |
| C6 | Female | -0.57 (-0.87 to -0.28) | -2.15 (-2.43 to -1.86) | -4.81 (-5.15 to -4.48) | -2.35 (-2.68 to -2.03) | -1.30 (-1.62 to -0.99) | -4.15 (-4.39 to -3.91) | -106 (-110 to -103) | -24.7781 |
| C7 | Male | -0.70 (-0.87 to -0.53) | -0.15 (-0.28 to -0.03) | 0.13 (-0.00 to 0.26) | 0.14 (0.06 to 0.23) | -0.17 (-0.34 to -0.01) | 0.07 (-0.06 to 0.20) | -1 (-3 to 2) | -0.1749 |
| C7 | Female | -1.12 (-1.31 to -0.93) | -2.76 (-2.88 to -2.64) | -3.25 (-3.37 to -3.12) | -2.58 (-2.66 to -2.50) | -1.24 (-1.38 to -1.09) | -3.17 (-3.30 to -3.05) | -98 (-100 to -96) | -25.7835 |
| T1 | Male | -0.86 (-1.02 to -0.69) | -0.75 (-0.86 to -0.64) | -0.62 (-0.73 to -0.50) | -0.46 (-0.53 to -0.38) | -0.44 (-0.58 to -0.30) | -0.65 (-0.76 to -0.54) | -23 (-25 to -21) | -7.4943 |
| T1 | Female | -1.32 (-1.49 to -1.16) | -2.98 (-3.08 to -2.88) | -3.42 (-3.53 to -3.31) | -2.64 (-2.71 to -2.57) | -1.18 (-1.30 to -1.06) | -3.36 (-3.47 to -3.26) | -102 (-104 to -100) | -29.4853 |
| T2 | Male | -1.10 (-1.26 to -0.94) | -0.78 (-0.88 to -0.67) | -0.53 (-0.64 to -0.43) | -0.37 (-0.45 to -0.30) | -0.38 (-0.51 to -0.25) | -0.59 (-0.69 to -0.49) | -21 (-23 to -19) | -7.344 |
| T2 | Female | -1.44 (-1.59 to -1.29) | -2.75 (-2.85 to -2.65) | -3.07 (-3.17 to -2.97) | -2.40 (-2.46 to -2.33) | -1.17 (-1.29 to -1.05) | -3.03 (-3.13 to -2.93) | -94 (-96 to -92) | -29.1705 |
| T3 | Male | -1.10 (-1.26 to -0.94) | -0.86 (-0.96 to -0.75) | -0.66 (-0.77 to -0.56) | -0.52 (-0.60 to -0.45) | -0.54 (-0.68 to -0.41) | -0.71 (-0.82 to -0.60) | -26 (-28 to -24) | -9.1206 |
| T3 | Female | -1.55 (-1.70 to -1.40) | -2.87 (-2.97 to -2.77) | -3.19 (-3.29 to -3.09) | -2.50 (-2.57 to -2.44) | -1.27 (-1.39 to -1.16) | -3.15 (-3.25 to -3.05) | -98 (-100 to -97) | -30.7842 |
| T4 | Male | -1.15 (-1.31 to -1.00) | -0.91 (-1.01 to -0.80) | -0.71 (-0.82 to -0.60) | -0.57 (-0.64 to -0.49) | -0.59 (-0.73 to -0.46) | -0.76 (-0.86 to -0.65) | -28 (-30 to -26) | -9.8366 |
| T4 | Female | -1.54 (-1.69 to -1.38) | -2.95 (-3.04 to -2.85) | -3.28 (-3.38 to -3.17) | -2.52 (-2.59 to -2.46) | -1.19 (-1.30 to -1.07) | -3.24 (-3.34 to -3.14) | -99 (-101 to -98) | -31.4916 |
| T5 | Male | -1.15 (-1.30 to -1.00) | -0.87 (-0.97 to -0.77) | -0.65 (-0.75 to -0.54) | -0.47 (-0.55 to -0.40) | -0.51 (-0.64 to -0.38) | -0.70 (-0.81 to -0.60) | -25 (-27 to -23) | -9.0398 |
| T5 | Female | -1.60 (-1.75 to -1.45) | -2.90 (-2.99 to -2.81) | -3.17 (-3.27 to -3.07) | -2.42 (-2.48 to -2.35) | -1.13 (-1.25 to -1.02) | -3.15 (-3.24 to -3.05) | -96 (-98 to -95) | -31.0815 |
| T6 | Male | -1.12 (-1.26 to -0.98) | -0.85 (-0.94 to -0.75) | -0.63 (-0.73 to -0.53) | -0.47 (-0.54 to -0.39) | -0.52 (-0.65 to -0.39) | -0.68 (-0.78 to -0.58) | -25 (-26 to -23) | -8.907 |
| T6 | Female | -1.60 (-1.73 to -1.46) | -2.82 (-2.91 to -2.73) | -3.06 (-3.16 to -2.97) | -2.32 (-2.38 to -2.26) | -1.08 (-1.20 to -0.97) | -3.04 (-3.14 to -2.95) | -93 (-95 to -91) | -30.3646 |
| T7 | Male | -1.19 (-1.31 to -1.06) | -0.84 (-0.93 to -0.74) | -0.58 (-0.68 to -0.48) | -0.41 (-0.48 to -0.34) | -0.52 (-0.65 to -0.40) | -0.64 (-0.74 to -0.54) | -24 (-25 to -22) | -8.6321 |
| T7 | Female | -1.56 (-1.68 to -1.43) | -2.79 (-2.88 to -2.70) | -2.99 (-3.08 to -2.89) | -2.15 (-2.21 to -2.08) | -0.92 (-1.03 to -0.82) | -2.98 (-3.07 to -2.89) | -88 (-90 to -87) | -29.4845 |
| T8 | Male | -1.23 (-1.34 to -1.11) | -0.82 (-0.91 to -0.74) | -0.54 (-0.63 to -0.45) | -0.37 (-0.44 to -0.31) | -0.51 (-0.62 to -0.39) | -0.61 (-0.70 to -0.52) | -22 (-24 to -21) | -8.3549 |
| T8 | Female | -1.66 (-1.78 to -1.53) | -2.84 (-2.92 to -2.75) | -2.98 (-3.06 to -2.89) | -2.07 (-2.13 to -2.01) | -0.80 (-0.91 to -0.70) | -2.99 (-3.07 to -2.90) | -87 (-89 to -85) | -29.4216 |
| T9 | Male | -1.34 (-1.55 to -1.13) | -0.90 (-1.05 to -0.74) | -0.51 (-0.69 to -0.33) | -0.35 (-0.54 to -0.15) | -0.53 (-0.75 to -0.32) | -0.61 (-0.76 to -0.46) | -23 (-25 to -21) | -8.3842 |
| T9 | Female | -0.90 (-1.10 to -0.69) | -2.60 (-2.75 to -2.44) | -3.82 (-3.99 to -3.65) | -1.73 (-1.91 to -1.56) | -0.89 (-1.08 to -0.71) | -3.63 (-3.78 to -3.49) | -90 (-93 to -88) | -29.9781 |
| T10 | Male | -1.51 (-1.72 to -1.30) | -1.06 (-1.22 to -0.90) | -0.67 (-0.84 to -0.49) | -0.31 (-0.50 to -0.12) | -0.47 (-0.68 to -0.26) | -0.78 (-0.93 to -0.63) | -25 (-27 to -23) | -9.2632 |
| T10 | Female | -0.99 (-1.19 to -0.79) | -2.75 (-2.90 to -2.60) | -3.89 (-4.06 to -3.73) | -1.81 (-1.98 to -1.64) | -0.85 (-1.04 to -0.67) | -3.74 (-3.88 to -3.60) | -93 (-95 to -91) | -30.7969 |
| T11 | Male | -1.72 (-1.93 to -1.52) | -1.29 (-1.45 to -1.14) | -0.75 (-0.92 to -0.58) | -0.47 (-0.66 to -0.28) | -0.30 (-0.50 to -0.10) | -0.91 (-1.05 to -0.76) | -28 (-30 to -26) | -10.8385 |
| T11 | Female | -1.03 (-1.22 to -0.83) | -2.76 (-2.90 to -2.62) | -3.88 (-4.04 to -3.72) | -1.76 (-1.94 to -1.59) | -0.78 (-0.96 to -0.60) | -3.74 (-3.87 to -3.61) | -92 (-94 to -90) | -31.5727 |
| T12 | Male | -1.85 (-2.04 to -1.66) | -1.44 (-1.58 to -1.29) | -0.91 (-1.07 to -0.76) | -0.49 (-0.66 to -0.31) | -0.30 (-0.48 to -0.11) | -1.07 (-1.20 to -0.94) | -31 (-33 to -29) | -12.7711 |
| T12 | Female | -1.23 (-1.41 to -1.05) | -2.80 (-2.93 to -2.66) | -3.72 (-3.87 to -3.58) | -1.70 (-1.87 to -1.54) | -0.49 (-0.67 to -0.31) | -3.62 (-3.74 to -3.50) | -87 (-89 to -85) | -31.6893 |
| L1 | Male | -1.85 (-2.04 to -1.67) | -1.44 (-1.59 to -1.30) | -0.81 (-0.96 to -0.66) | -0.21 (-0.38 to -0.04) | -0.11 (-0.30 to 0.08) | -0.99 (-1.12 to -0.87) | -26 (-28 to -24) | -10.4319 |
| L1 | Female | -1.38 (-1.56 to -1.20) | -2.86 (-2.99 to -2.72) | -3.55 (-3.70 to -3.41) | -1.50 (-1.65 to -1.34) | -0.28 (-0.46 to -0.10) | -3.51 (-3.63 to -3.39) | -82 (-84 to -80) | -29.6361 |
| L2 | Male | -1.99 (-2.18 to -1.79) | -1.45 (-1.60 to -1.30) | -0.53 (-0.69 to -0.37) | 0.08 (-0.10 to 0.25) | -0.02 (-0.23 to 0.18) | -0.78 (-0.91 to -0.65) | -19 (-21 to -17) | -7.4745 |
| L2 | Female | -1.61 (-1.79 to -1.43) | -3.01 (-3.15 to -2.87) | -3.55 (-3.70 to -3.40) | -1.34 (-1.50 to -1.17) | -0.43 (-0.61 to -0.24) | -3.55 (-3.67 to -3.42) | -83 (-85 to -81) | -28.7132 |
| L3 | Male | -2.09 (-2.29 to -1.90) | -1.50 (-1.65 to -1.36) | -0.53 (-0.69 to -0.37) | 0.05 (-0.13 to 0.23) | -0.06 (-0.27 to 0.14) | -0.79 (-0.93 to -0.66) | -20 (-23 to -18) | -7.7437 |
| L3 | Female | -1.62 (-1.80 to -1.43) | -3.07 (-3.21 to -2.93) | -3.75 (-3.90 to -3.60) | -1.47 (-1.64 to -1.31) | -0.59 (-0.78 to -0.41) | -3.71 (-3.83 to -3.58) | -89 (-91 to -87) | -29.9556 |
| L4 | Male | -2.01 (-2.21 to -1.81) | -1.45 (-1.61 to -1.30) | -0.32 (-0.48 to -0.15) | 0.40 (0.21 to 0.59) | -0.05 (-0.27 to 0.17) | -0.63 (-0.77 to -0.49) | -14 (-16 to -12) | -5.2626 |
| L4 | Female | -1.33 (-1.52 to -1.14) | -3.00 (-3.15 to -2.86) | -3.79 (-3.94 to -3.63) | -1.42 (-1.59 to -1.24) | -0.67 (-0.87 to -0.48) | -3.73 (-3.86 to -3.60) | -89 (-91 to -87) | -29.3663 |
| L5 | Male | -1.98 (-2.19 to -1.76) | -1.29 (-1.46 to -1.13) | -0.05 (-0.24 to 0.13) | 0.43 (0.22 to 0.64) | -0.29 (-0.53 to -0.05) | -0.38 (-0.54 to -0.22) | -12 (-15 to -10) | -4.4899 |
| L5 | Female | -1.17 (-1.38 to -0.97) | -2.91 (-3.06 to -2.76) | -3.79 (-3.96 to -3.62) | -1.34 (-1.53 to -1.15) | -0.81 (-1.02 to -0.60) | -3.70 (-3.84 to -3.56) | -88 (-91 to -86) | -29.1666 |
| S1 | Male | -1.46 (-1.69 to -1.22) | -1.55 (-1.73 to -1.38) | -1.34 (-1.54 to -1.15) | -1.02 (-1.24 to -0.81) | -1.14 (-1.37 to -0.92) | -1.43 (-1.59 to -1.26) | -51 (-53 to -48) | -17.5893 |
| S1 | Female | -0.66 (-0.88 to -0.44) | -2.89 (-3.05 to -2.72) | -4.64 (-4.83 to -4.46) | -2.66 (-2.86 to -2.46) | -1.76 (-1.97 to -1.55) | -4.34 (-4.49 to -4.19) | -120 (-122 to -117) | -37.1853 |
| Left rib 1 | Male | -1.19 (-1.45 to -0.94) | -1.57 (-1.82 to -1.31) | -1.93 (-2.22 to -1.64) | -1.72 (-2.01 to -1.43) | -1.45 (-1.73 to -1.17) | -1.84 (-2.05 to -1.64) | -67 (-70 to -64) | -19.3587 |
| Left rib 1 | Female | -1.03 (-1.27 to -0.79) | -2.60 (-2.84 to -2.35) | -5.03 (-5.29 to -4.77) | -3.01 (-3.27 to -2.75) | -2.07 (-2.31 to -1.83) | -4.43 (-4.61 to -4.24) | -127 (-130 to -124) | -34.1285 |
| Left rib 2 | Male | -3.13 (-3.29 to -2.97) | -1.90 (-2.01 to -1.79) | -0.98 (-1.09 to -0.86) | -0.36 (-0.44 to -0.28) | -0.13 (-0.27 to 0.00) | -1.19 (-1.31 to -1.08) | -34 (-36 to -32) | -12.7636 |
| Left rib 2 | Female | -3.74 (-3.93 to -3.56) | -4.22 (-4.34 to -4.11) | -3.87 (-3.99 to -3.75) | -2.68 (-2.76 to -2.61) | -1.23 (-1.35 to -1.10) | -3.99 (-4.11 to -3.88) | -120 (-122 to -118) | -37.087 |
| Left rib 3 | Male | -4.05 (-4.23 to -3.87) | -2.65 (-2.77 to -2.54) | -1.52 (-1.64 to -1.41) | -0.66 (-0.73 to -0.58) | -0.22 (-0.35 to -0.09) | -1.80 (-1.91 to -1.68) | -51 (-53 to -48) | -19.377 |
| Left rib 3 | Female | -4.76 (-4.95 to -4.56) | -5.23 (-5.35 to -5.11) | -4.59 (-4.72 to -4.47) | -2.86 (-2.93 to -2.78) | -0.87 (-1.00 to -0.74) | -4.80 (-4.92 to -4.67) | -135 (-138 to -133) | -40.6047 |
| Left rib 4 | Male | -4.12 (-4.45 to -3.79) | -2.58 (-2.83 to -2.33) | -1.46 (-1.71 to -1.21) | -0.46 (-0.69 to -0.22) | -0.21 (-0.46 to 0.04) | -1.73 (-1.94 to -1.52) | -47 (-50 to -44) | -17.6656 |
| Left rib 4 | Female | -3.42 (-3.80 to -3.05) | -4.69 (-4.97 to -4.41) | -5.51 (-5.78 to -5.23) | -2.05 (-2.29 to -1.80) | -1.03 (-1.27 to -0.79) | -5.48 (-5.71 to -5.25) | -133 (-136 to -130) | -39.7215 |
| Left rib 5 | Male | -3.73 (-4.06 to -3.40) | -2.24 (-2.49 to -1.99) | -1.36 (-1.62 to -1.11) | -0.21 (-0.45 to 0.02) | 0.05 (-0.20 to 0.29) | -1.58 (-1.78 to -1.37) | -38 (-41 to -35) | -14.4479 |
| Left rib 5 | Female | -2.86 (-3.24 to -2.48) | -4.16 (-4.44 to -3.88) | -5.34 (-5.63 to -5.06) | -1.91 (-2.16 to -1.65) | -1.04 (-1.29 to -0.79) | -5.21 (-5.45 to -4.98) | -124 (-128 to -121) | -38.0015 |
| Left rib 6 | Male | -3.20 (-3.49 to -2.92) | -1.91 (-2.15 to -1.66) | -1.37 (-1.64 to -1.10) | -0.18 (-0.42 to 0.07) | -0.05 (-0.30 to 0.19) | -1.50 (-1.70 to -1.29) | -35 (-38 to -32) | -13.0341 |
| Left rib 6 | Female | -2.74 (-3.06 to -2.42) | -3.68 (-3.96 to -3.40) | -5.26 (-5.56 to -4.95) | -1.92 (-2.20 to -1.65) | -1.31 (-1.57 to -1.06) | -4.93 (-5.16 to -4.70) | -122 (-125 to -119) | -36.138 |
| Left rib 7 | Male | -3.55 (-3.87 to -3.22) | -2.14 (-2.37 to -1.92) | -1.40 (-1.63 to -1.17) | -0.39 (-0.64 to -0.15) | -0.39 (-0.64 to -0.13) | -1.59 (-1.79 to -1.39) | -43 (-46 to -40) | -15.3342 |
| Left rib 7 | Female | -2.80 (-3.16 to -2.45) | -4.39 (-4.64 to -4.14) | -5.49 (-5.74 to -5.23) | -2.19 (-2.46 to -1.93) | -1.13 (-1.39 to -0.87) | -5.43 (-5.66 to -5.21) | -132 (-135 to -129) | -36.5632 |
| Left rib 8 | Male | -4.26 (-4.54 to -3.99) | -2.57 (-2.81 to -2.33) | -1.46 (-1.73 to -1.19) | -0.70 (-0.95 to -0.45) | -0.92 (-1.17 to -0.67) | -1.72 (-1.93 to -1.52) | -56 (-59 to -53) | -18.6182 |
| Left rib 8 | Female | -4.29 (-4.58 to -4.00) | -5.12 (-5.37 to -4.87) | -6.07 (-6.36 to -5.79) | -2.12 (-2.38 to -1.86) | -1.66 (-1.91 to -1.41) | -5.92 (-6.14 to -5.71) | -150 (-153 to -147) | -39.0079 |
| Left rib 9 | Male | -4.03 (-4.31 to -3.74) | -2.42 (-2.66 to -2.17) | -1.28 (-1.56 to -1.00) | -0.74 (-1.01 to -0.48) | -1.10 (-1.37 to -0.83) | -1.55 (-1.76 to -1.33) | -55 (-59 to -52) | -17.2367 |
| Left rib 9 | Female | -4.07 (-4.35 to -3.79) | -5.10 (-5.35 to -4.86) | -6.13 (-6.41 to -5.85) | -2.29 (-2.56 to -2.03) | -1.83 (-2.08 to -1.58) | -5.96 (-6.18 to -5.74) | -154 (-157 to -151) | -38.8903 |
| Left rib 10 | Male | -4.00 (-4.28 to -3.71) | -2.47 (-2.72 to -2.22) | -1.34 (-1.62 to -1.06) | -0.63 (-0.90 to -0.37) | -0.65 (-0.93 to -0.37) | -1.61 (-1.83 to -1.40) | -51 (-54 to -48) | -14.6538 |
| Left rib 10 | Female | -3.81 (-4.09 to -3.53) | -5.17 (-5.41 to -4.93) | -6.40 (-6.68 to -6.12) | -2.58 (-2.84 to -2.32) | -1.76 (-2.01 to -1.51) | -6.18 (-6.40 to -5.97) | -159 (-162 to -156) | -37.1252 |
| Left rib 11 | Male | -4.27 (-4.62 to -3.93) | -3.00 (-3.25 to -2.75) | -1.86 (-2.12 to -1.61) | -0.94 (-1.22 to -0.67) | -0.96 (-1.25 to -0.67) | -2.17 (-2.39 to -1.94) | -68 (-71 to -64) | -18.5031 |
| Left rib 11 | Female | -4.09 (-4.45 to -3.74) | -5.98 (-6.23 to -5.74) | -6.90 (-7.16 to -6.65) | -3.15 (-3.42 to -2.88) | -1.85 (-2.12 to -1.59) | -6.91 (-7.14 to -6.69) | -179 (-182 to -176) | -40.3589 |
| Left rib 12 | Male | -4.38 (-4.57 to -4.19) | -2.86 (-3.00 to -2.73) | -1.76 (-1.89 to -1.63) | -1.08 (-1.17 to -0.99) | -0.71 (-0.86 to -0.57) | -2.02 (-2.15 to -1.89) | -64 (-67 to -62) | -18.2716 |
| Left rib 12 | Female | -5.05 (-5.28 to -4.83) | -5.98 (-6.12 to -5.83) | -5.48 (-5.63 to -5.33) | -3.57 (-3.67 to -3.47) | -1.16 (-1.32 to -1.00) | -5.66 (-5.81 to -5.52) | -162 (-164 to -159) | -37.2673 |
| Right rib 1 | Male | -1.46 (-1.70 to -1.22) | -1.54 (-1.79 to -1.28) | -1.92 (-2.15 to -1.70) | -1.77 (-1.99 to -1.55) | -1.56 (-1.84 to -1.28) | -1.82 (-1.98 to -1.65) | -68 (-71 to -65) | -19.707 |
| Right rib 1 | Female | -1.61 (-1.83 to -1.39) | -2.88 (-3.12 to -2.65) | -4.77 (-4.97 to -4.57) | -3.46 (-3.64 to -3.27) | -2.04 (-2.28 to -1.81) | -4.28 (-4.43 to -4.13) | -132 (-134 to -129) | -35.1899 |
| Right rib 2 | Male | -3.20 (-3.36 to -3.03) | -1.91 (-2.02 to -1.80) | -0.96 (-1.07 to -0.85) | -0.36 (-0.44 to -0.28) | -0.18 (-0.31 to -0.04) | -1.18 (-1.30 to -1.07) | -34 (-36 to -32) | -12.8428 |
| Right rib 2 | Female | -3.90 (-4.09 to -3.71) | -4.35 (-4.46 to -4.23) | -3.96 (-4.08 to -3.84) | -2.73 (-2.81 to -2.66) | -1.22 (-1.35 to -1.09) | -4.09 (-4.21 to -3.97) | -123 (-125 to -121) | -37.4821 |
| Right rib 3 | Male | -4.20 (-4.38 to -4.02) | -2.73 (-2.84 to -2.62) | -1.54 (-1.66 to -1.43) | -0.64 (-0.72 to -0.57) | -0.21 (-0.34 to -0.08) | -1.83 (-1.94 to -1.71) | -51 (-53 to -49) | -19.8563 |
| Right rib 3 | Female | -4.93 (-5.14 to -4.73) | -5.30 (-5.43 to -5.18) | -4.61 (-4.73 to -4.48) | -2.84 (-2.92 to -2.76) | -0.85 (-0.99 to -0.72) | -4.83 (-4.95 to -4.70) | -136 (-138 to -134) | -40.7274 |
| Right rib 4 | Male | -4.08 (-4.35 to -3.82) | -2.61 (-2.87 to -2.36) | -1.62 (-1.89 to -1.35) | -0.32 (-0.57 to -0.07) | -0.07 (-0.31 to 0.17) | -1.86 (-2.06 to -1.66) | -46 (-49 to -43) | -17.4114 |
| Right rib 4 | Female | -3.93 (-4.23 to -3.62) | -4.64 (-4.94 to -4.35) | -5.50 (-5.80 to -5.20) | -1.70 (-1.97 to -1.43) | -1.04 (-1.28 to -0.79) | -5.34 (-5.56 to -5.12) | -129 (-132 to -126) | -38.1801 |
| Right rib 5 | Male | -3.54 (-3.81 to -3.27) | -2.16 (-2.41 to -1.91) | -1.26 (-1.52 to -0.99) | -0.14 (-0.39 to 0.10) | 0.20 (-0.04 to 0.45) | -1.47 (-1.67 to -1.28) | -34 (-37 to -31) | -12.9347 |
| Right rib 5 | Female | -2.89 (-3.22 to -2.56) | -3.64 (-3.94 to -3.34) | -5.21 (-5.52 to -4.90) | -1.66 (-1.94 to -1.38) | -1.02 (-1.28 to -0.77) | -4.87 (-5.10 to -4.64) | -115 (-119 to -112) | -35.5219 |
| Right rib 6 | Male | -3.24 (-3.52 to -2.97) | -1.98 (-2.23 to -1.74) | -1.39 (-1.66 to -1.13) | -0.17 (-0.41 to 0.08) | 0.04 (-0.21 to 0.28) | -1.54 (-1.74 to -1.33) | -35 (-38 to -32) | -12.9562 |
| Right rib 6 | Female | -2.65 (-2.97 to -2.34) | -3.60 (-3.88 to -3.33) | -5.44 (-5.74 to -5.14) | -1.98 (-2.25 to -1.70) | -1.34 (-1.59 to -1.09) | -5.05 (-5.27 to -4.82) | -124 (-127 to -120) | -36.047 |
| Right rib 7 | Male | -3.53 (-3.79 to -3.27) | -2.12 (-2.34 to -1.89) | -1.42 (-1.67 to -1.16) | -0.38 (-0.62 to -0.15) | -0.54 (-0.78 to -0.29) | -1.58 (-1.78 to -1.39) | -45 (-47 to -42) | -15.6463 |
| Right rib 7 | Female | -3.04 (-3.33 to -2.74) | -3.98 (-4.24 to -3.73) | -5.57 (-5.85 to -5.29) | -2.03 (-2.29 to -1.77) | -1.37 (-1.62 to -1.12) | -5.25 (-5.46 to -5.03) | -130 (-133 to -126) | -35.7435 |
| Right rib 8 | Male | -4.16 (-4.43 to -3.89) | -2.48 (-2.71 to -2.24) | -1.31 (-1.58 to -1.05) | -0.66 (-0.90 to -0.42) | -0.92 (-1.17 to -0.67) | -1.59 (-1.79 to -1.39) | -54 (-57 to -51) | -17.792 |
| Right rib 8 | Female | -3.85 (-4.14 to -3.57) | -4.69 (-4.94 to -4.44) | -5.67 (-5.95 to -5.39) | -2.08 (-2.34 to -1.83) | -1.62 (-1.87 to -1.37) | -5.51 (-5.72 to -5.29) | -141 (-144 to -138) | -36.8019 |
| Right rib 9 | Male | -3.94 (-4.21 to -3.67) | -2.30 (-2.54 to -2.06) | -1.17 (-1.44 to -0.90) | -0.58 (-0.84 to -0.32) | -0.74 (-1.01 to -0.48) | -1.43 (-1.64 to -1.22) | -48 (-51 to -45) | -15.1073 |
| Right rib 9 | Female | -3.88 (-4.16 to -3.61) | -4.81 (-5.05 to -4.57) | -5.72 (-6.00 to -5.44) | -1.97 (-2.23 to -1.71) | -1.57 (-1.82 to -1.32) | -5.58 (-5.79 to -5.36) | -141 (-144 to -138) | -35.8522 |
| Right rib 10 | Male | -3.76 (-4.05 to -3.48) | -2.24 (-2.49 to -2.00) | -1.21 (-1.49 to -0.93) | -0.47 (-0.73 to -0.20) | -0.28 (-0.56 to -0.01) | -1.45 (-1.67 to -1.24) | -42 (-45 to -39) | -12.1494 |
| Right rib 10 | Female | -3.65 (-3.93 to -3.37) | -4.95 (-5.19 to -4.72) | -6.09 (-6.37 to -5.81) | -2.13 (-2.39 to -1.87) | -1.65 (-1.90 to -1.39) | -5.90 (-6.12 to -5.69) | -148 (-151 to -145) | -34.5269 |
| Right rib 11 | Male | -4.25 (-4.54 to -3.96) | -2.82 (-3.07 to -2.57) | -1.75 (-2.04 to -1.47) | -0.77 (-1.03 to -0.51) | -0.63 (-0.90 to -0.35) | -2.01 (-2.23 to -1.79) | -60 (-63 to -57) | -16.3578 |
| Right rib 11 | Female | -4.37 (-4.66 to -4.07) | -5.60 (-5.85 to -5.35) | -6.86 (-7.14 to -6.58) | -2.62 (-2.87 to -2.36) | -1.81 (-2.07 to -1.56) | -6.64 (-6.86 to -6.43) | -169 (-172 to -166) | -38.2795 |
| Right rib 12 | Male | -4.34 (-4.52 to -4.15) | -2.76 (-2.89 to -2.63) | -1.54 (-1.67 to -1.41) | -0.68 (-0.78 to -0.59) | -0.20 (-0.35 to -0.05) | -1.83 (-1.97 to -1.70) | -52 (-54 to -50) | -14.9304 |
| Right rib 12 | Female | -4.86 (-5.08 to -4.65) | -5.72 (-5.87 to -5.58) | -5.21 (-5.35 to -5.06) | -3.32 (-3.42 to -3.22) | -0.90 (-1.06 to -0.74) | -5.39 (-5.54 to -5.25) | -151 (-154 to -149) | -35.4104 |

**Supplementary Table 2 (continued): Trajectory statistics for all 49 skeletal sites (age of steepest decline)**

| **Site** | **Sex** | **Age of steepest decline, years (95% CI)** | **Interior maximum present** | **Steepest rate of change, HU/year (95% CI)** |
| --- | --- | --- | --- | --- |
| C1 | Male | not identified | False | -0.84 (-1.37 to -0.45) |
| C1 | Female | 55.5 (52.8 to 60.0) | True | -5.76 (-6.33 to -5.23) |
| C2 | Male | 48.5 (35.0 to 72.8) | True | -1.26 (-1.94 to -1.10) |
| C2 | Female | 55.5 (53.0 to 59.8) | True | -6.81 (-7.37 to -6.28) |
| C3 | Male | not identified | False | -0.88 (-1.40 to -0.68) |
| C3 | Female | 55.2 (52.8 to 60.0) | True | -5.99 (-6.54 to -5.48) |
| C4 | Male | not identified | False | -1.13 (-1.60 to -0.71) |
| C4 | Female | 55.0 (51.8 to 58.5) | True | -6.54 (-7.18 to -5.96) |
| C5 | Male | not identified | False | -1.20 (-1.60 to -0.84) |
| C5 | Female | 54.2 (52.5 to 59.0) | True | -6.01 (-6.54 to -5.48) |
| C6 | Male | not identified | False | -1.09 (-1.50 to -0.83) |
| C6 | Female | 54.2 (52.5 to 59.5) | True | -5.31 (-5.76 to -4.91) |
| C7 | Male | not identified | False | -0.69 (-0.86 to -0.54) |
| C7 | Female | 54.2 (52.8 to 58.5) | True | -3.30 (-3.43 to -3.17) |
| T1 | Male | not identified | False | -0.86 (-1.03 to -0.73) |
| T1 | Female | 54.0 (52.5 to 57.5) | True | -3.48 (-3.60 to -3.37) |
| T2 | Male | not identified | False | -1.10 (-1.26 to -0.94) |
| T2 | Female | 52.8 (52.2 to 57.8) | True | -3.13 (-3.23 to -3.02) |
| T3 | Male | not identified | False | -1.10 (-1.27 to -0.94) |
| T3 | Female | 52.5 (52.0 to 58.0) | True | -3.25 (-3.36 to -3.14) |
| T4 | Male | not identified | False | -1.15 (-1.32 to -1.00) |
| T4 | Female | 52.5 (52.0 to 57.8) | True | -3.34 (-3.45 to -3.23) |
| T5 | Male | not identified | False | -1.15 (-1.30 to -1.00) |
| T5 | Female | 52.5 (51.5 to 57.5) | True | -3.24 (-3.34 to -3.14) |
| T6 | Male | not identified | False | -1.12 (-1.27 to -0.98) |
| T6 | Female | 52.2 (51.0 to 57.5) | True | -3.14 (-3.24 to -3.03) |
| T7 | Male | not identified | False | -1.18 (-1.31 to -1.06) |
| T7 | Female | 52.0 (50.8 to 56.8) | True | -3.08 (-3.18 to -2.98) |
| T8 | Male | not identified | False | -1.22 (-1.34 to -1.11) |
| T8 | Female | 51.5 (50.2 to 56.2) | True | -3.09 (-3.19 to -2.99) |
| T9 | Male | not identified | False | -1.33 (-1.56 to -1.11) |
| T9 | Female | 51.2 (50.2 to 56.0) | True | -4.38 (-4.68 to -4.09) |
| T10 | Male | not identified | False | -1.49 (-1.71 to -1.27) |
| T10 | Female | 51.0 (50.0 to 55.8) | True | -4.56 (-4.86 to -4.28) |
| T11 | Male | not identified | False | -1.72 (-1.93 to -1.49) |
| T11 | Female | 51.0 (49.8 to 55.8) | True | -4.57 (-4.86 to -4.30) |
| T12 | Male | not identified | False | -1.84 (-2.04 to -1.63) |
| T12 | Female | 51.0 (49.0 to 55.2) | True | -4.39 (-4.65 to -4.14) |
| L1 | Male | not identified | False | -1.85 (-2.05 to -1.65) |
| L1 | Female | 50.5 (48.0 to 54.5) | True | -4.31 (-4.56 to -4.07) |
| L2 | Male | not identified | False | -2.00 (-2.20 to -1.79) |
| L2 | Female | 50.5 (47.0 to 53.5) | True | -4.32 (-4.58 to -4.07) |
| L3 | Male | not identified | False | -2.11 (-2.31 to -1.90) |
| L3 | Female | 50.8 (47.5 to 54.0) | True | -4.45 (-4.71 to -4.20) |
| L4 | Male | not identified | False | -2.02 (-2.24 to -1.81) |
| L4 | Female | 50.8 (48.0 to 54.0) | True | -4.55 (-4.80 to -4.28) |
| L5 | Male | not identified | False | -2.00 (-2.24 to -1.77) |
| L5 | Female | 52.5 (48.5 to 54.2) | True | -4.49 (-4.76 to -4.20) |
| S1 | Male | not identified | False | -1.64 (-2.06 to -1.51) |
| S1 | Female | 53.2 (51.5 to 57.0) | True | -5.23 (-5.55 to -4.93) |
| Left rib 1 | Male | 56.0 (52.8 to 74.5) | True | -1.98 (-2.54 to -1.85) |
| Left rib 1 | Female | 54.5 (52.8 to 59.5) | True | -5.45 (-5.81 to -5.13) |
| Left rib 2 | Male | not identified | False | -3.12 (-3.28 to -2.97) |
| Left rib 2 | Female | 51.0 (44.5 to 55.2) | True | -4.26 (-4.38 to -4.15) |
| Left rib 3 | Male | not identified | False | -4.04 (-4.23 to -3.87) |
| Left rib 3 | Female | 51.8 (43.5 to 53.0) | True | -5.27 (-5.40 to -5.16) |
| Left rib 4 | Male | not identified | False | -4.09 (-4.45 to -3.72) |
| Left rib 4 | Female | 50.8 (43.0 to 54.5) | True | -6.63 (-7.10 to -6.16) |
| Left rib 5 | Male | not identified | False | -3.68 (-4.04 to -3.31) |
| Left rib 5 | Female | 52.0 (43.8 to 55.2) | True | -6.37 (-6.85 to -5.89) |
| Left rib 6 | Male | not identified | False | -3.15 (-3.45 to -2.85) |
| Left rib 6 | Female | 52.2 (45.2 to 56.0) | True | -5.82 (-6.23 to -5.40) |
| Left rib 7 | Male | not identified | False | -3.48 (-3.83 to -3.12) |
| Left rib 7 | Female | 52.0 (44.2 to 55.5) | True | -6.60 (-7.07 to -6.17) |
| Left rib 8 | Male | not identified | False | -4.23 (-4.51 to -3.93) |
| Left rib 8 | Female | 51.5 (42.5 to 54.2) | True | -6.76 (-7.13 to -6.37) |
| Left rib 9 | Male | not identified | False | -4.00 (-4.29 to -3.70) |
| Left rib 9 | Female | 51.8 (44.2 to 54.2) | True | -6.78 (-7.16 to -6.39) |
| Left rib 10 | Male | not identified | False | -3.97 (-4.27 to -3.66) |
| Left rib 10 | Female | 51.8 (46.0 to 54.8) | True | -7.02 (-7.39 to -6.62) |
| Left rib 11 | Male | not identified | False | -4.25 (-4.62 to -3.85) |
| Left rib 11 | Female | 51.8 (45.2 to 54.5) | True | -8.14 (-8.62 to -7.71) |
| Left rib 12 | Male | not identified | False | -4.37 (-4.56 to -4.18) |
| Left rib 12 | Female | 51.8 (45.5 to 54.8) | True | -6.05 (-6.20 to -5.90) |
| Right rib 1 | Male | 54.2 (51.2 to 75.0) | True | -2.04 (-2.43 to -1.85) |
| Right rib 1 | Female | 54.8 (52.5 to 60.0) | True | -5.22 (-5.51 to -4.94) |
| Right rib 2 | Male | not identified | False | -3.18 (-3.35 to -3.03) |
| Right rib 2 | Female | 52.5 (44.0 to 55.0) | True | -4.38 (-4.51 to -4.27) |
| Right rib 3 | Male | not identified | False | -4.19 (-4.37 to -4.02) |
| Right rib 3 | Female | 51.8 (42.5 to 53.0) | True | -5.36 (-5.49 to -5.25) |
| Right rib 4 | Male | not identified | False | -4.05 (-4.34 to -3.77) |
| Right rib 4 | Female | 52.2 (43.0 to 54.2) | True | -6.02 (-6.40 to -5.63) |
| Right rib 5 | Male | not identified | False | -3.51 (-3.80 to -3.22) |
| Right rib 5 | Female | 52.2 (43.5 to 56.0) | True | -5.70 (-6.09 to -5.29) |
| Right rib 6 | Male | not identified | False | -3.19 (-3.48 to -2.90) |
| Right rib 6 | Female | 52.5 (45.5 to 56.2) | True | -6.01 (-6.41 to -5.59) |
| Right rib 7 | Male | not identified | False | -3.48 (-3.75 to -3.20) |
| Right rib 7 | Female | 52.2 (45.0 to 55.8) | True | -6.16 (-6.53 to -5.76) |
| Right rib 8 | Male | not identified | False | -4.14 (-4.41 to -3.85) |
| Right rib 8 | Female | 51.8 (43.2 to 54.5) | True | -6.29 (-6.65 to -5.90) |
| Right rib 9 | Male | not identified | False | -3.92 (-4.20 to -3.62) |
| Right rib 9 | Female | 51.8 (43.5 to 54.2) | True | -6.35 (-6.71 to -5.96) |
| Right rib 10 | Male | not identified | False | -3.74 (-4.03 to -3.43) |
| Right rib 10 | Female | 52.0 (45.5 to 54.5) | True | -6.74 (-7.11 to -6.37) |
| Right rib 11 | Male | not identified | False | -4.22 (-4.52 to -3.91) |
| Right rib 11 | Female | 51.8 (44.8 to 54.5) | True | -7.58 (-7.96 to -7.17) |
| Right rib 12 | Male | not identified | False | -4.32 (-4.51 to -4.15) |
| Right rib 12 | Female | 51.8 (45.2 to 54.8) | True | -5.79 (-5.94 to -5.64) |

One row per site and sex. Values in parentheses are 95% confidence intervals, derived analytically from the spline coefficient covariance (HC3). The age of steepest decline is reported only where the derivative has an interior minimum; its interval pools coefficient draws across all candidate spline dimensions, 2000 draws per dimension with equal weight. The comparison between the sexes is a property of the site rather than of one sex and is reported once per site in Supplementary Table 2B.

**Supplementary Table 2B: Difference between the sexes in the age trajectory, by site**

| **Site** | **Region** | **Excess female change 45–60 years, HU/year (95% CI)** | **Excess female change 40 to 80 years, HU (95% CI)** | **Excess female change, p (FDR)** | **Sex × age interaction, χ²** | **Sex × age interaction, df** | **Sex × age interaction, p** | **Sex × age interaction, p (FDR)** |
| --- | --- | --- | --- | --- | --- | --- | --- | --- |
| C1 | Cervical vertebrae | -4.37 (-4.84 to -3.91) | -119 (-126 to -112) | <0.0001 | 2008.91 | 7 | <0.0001 | <0.0001 |
| C2 | Cervical vertebrae | -4.24 (-4.70 to -3.78) | -111 (-118 to -104) | <0.0001 | 1757.81 | 7 | <0.0001 | <0.0001 |
| C3 | Cervical vertebrae | -4.10 (-4.53 to -3.68) | -113 (-120 to -106) | <0.0001 | 2022.28 | 7 | <0.0001 | <0.0001 |
| C4 | Cervical vertebrae | -4.87 (-5.38 to -4.36) | -117 (-124 to -109) | <0.0001 | 2174.44 | 8 | <0.0001 | <0.0001 |
| C5 | Cervical vertebrae | -4.49 (-4.93 to -4.05) | -112 (-119 to -106) | <0.0001 | 2573.94 | 8 | <0.0001 | <0.0001 |
| C6 | Cervical vertebrae | -4.19 (-4.54 to -3.84) | -107 (-112 to -102) | <0.0001 | 3599.14 | 8 | <0.0001 | <0.0001 |
| C7 | Cervical vertebrae | -3.24 (-3.42 to -3.06) | -98 (-101 to -94) | <0.0001 | 3881.95 | 4 | <0.0001 | <0.0001 |
| T1 | Thoracic vertebrae | -2.71 (-2.87 to -2.56) | -80 (-82 to -77) | <0.0001 | 3414.26 | 4 | <0.0001 | <0.0001 |
| T2 | Thoracic vertebrae | -2.44 (-2.59 to -2.30) | -73 (-76 to -71) | <0.0001 | 3282.52 | 4 | <0.0001 | <0.0001 |
| T3 | Thoracic vertebrae | -2.44 (-2.59 to -2.29) | -73 (-75 to -70) | <0.0001 | 3118.36 | 4 | <0.0001 | <0.0001 |
| T4 | Thoracic vertebrae | -2.48 (-2.63 to -2.34) | -72 (-74 to -69) | <0.0001 | 3042 | 4 | <0.0001 | <0.0001 |
| T5 | Thoracic vertebrae | -2.44 (-2.59 to -2.30) | -71 (-74 to -69) | <0.0001 | 3198.41 | 4 | <0.0001 | <0.0001 |
| T6 | Thoracic vertebrae | -2.36 (-2.50 to -2.23) | -68 (-71 to -66) | <0.0001 | 3091.59 | 4 | <0.0001 | <0.0001 |
| T7 | Thoracic vertebrae | -2.34 (-2.47 to -2.21) | -65 (-67 to -63) | <0.0001 | 3027.86 | 4 | <0.0001 | <0.0001 |
| T8 | Thoracic vertebrae | -2.38 (-2.51 to -2.25) | -64 (-67 to -62) | <0.0001 | 3358.6 | 4 | <0.0001 | <0.0001 |
| T9 | Thoracic vertebrae | -3.02 (-3.23 to -2.81) | -68 (-71 to -65) | <0.0001 | 3893.09 | 9 | <0.0001 | <0.0001 |
| T10 | Thoracic vertebrae | -2.96 (-3.16 to -2.76) | -68 (-71 to -65) | <0.0001 | 4046.42 | 9 | <0.0001 | <0.0001 |
| T11 | Thoracic vertebrae | -2.83 (-3.03 to -2.64) | -64 (-67 to -61) | <0.0001 | 3767.66 | 9 | <0.0001 | <0.0001 |
| T12 | Thoracic vertebrae | -2.55 (-2.73 to -2.37) | -56 (-59 to -53) | <0.0001 | 3351.04 | 9 | <0.0001 | <0.0001 |
| L1 | Lumbar vertebrae | -2.52 (-2.70 to -2.34) | -56 (-59 to -53) | <0.0001 | 3529.37 | 9 | <0.0001 | <0.0001 |
| L2 | Lumbar vertebrae | -2.76 (-2.95 to -2.58) | -64 (-67 to -61) | <0.0001 | 4153.12 | 9 | <0.0001 | <0.0001 |
| L3 | Lumbar vertebrae | -2.92 (-3.10 to -2.73) | -68 (-71 to -65) | <0.0001 | 4703.61 | 9 | <0.0001 | <0.0001 |
| L4 | Lumbar vertebrae | -3.10 (-3.29 to -2.91) | -75 (-78 to -72) | <0.0001 | 5069.43 | 9 | <0.0001 | <0.0001 |
| L5 | Lumbar vertebrae | -3.32 (-3.53 to -3.11) | -76 (-80 to -73) | <0.0001 | 4744.14 | 9 | <0.0001 | <0.0001 |
| S1 | Sacrum | -2.91 (-3.14 to -2.69) | -69 (-72 to -66) | <0.0001 | 3682.99 | 9 | <0.0001 | <0.0001 |
| Left rib 1 | Left ribs | -2.58 (-2.86 to -2.30) | -60 (-65 to -56) | <0.0001 | 1759.45 | 8 | <0.0001 | <0.0001 |
| Left rib 2 | Left ribs | -2.80 (-2.96 to -2.64) | -86 (-89 to -84) | <0.0001 | 3878.38 | 4 | <0.0001 | <0.0001 |
| Left rib 3 | Left ribs | -3.00 (-3.17 to -2.83) | -85 (-88 to -82) | <0.0001 | 3305.48 | 4 | <0.0001 | <0.0001 |
| Left rib 4 | Left ribs | -3.75 (-4.06 to -3.44) | -86 (-90 to -81) | <0.0001 | 2621.56 | 9 | <0.0001 | <0.0001 |
| Left rib 5 | Left ribs | -3.64 (-3.95 to -3.32) | -87 (-91 to -82) | <0.0001 | 2726.1 | 9 | <0.0001 | <0.0001 |
| Left rib 6 | Left ribs | -3.43 (-3.74 to -3.13) | -87 (-91 to -82) | <0.0001 | 2935.32 | 8 | <0.0001 | <0.0001 |
| Left rib 7 | Left ribs | -3.84 (-4.14 to -3.54) | -89 (-93 to -84) | <0.0001 | 3142.29 | 9 | <0.0001 | <0.0001 |
| Left rib 8 | Left ribs | -4.20 (-4.50 to -3.90) | -93 (-98 to -89) | <0.0001 | 3579.64 | 8 | <0.0001 | <0.0001 |
| Left rib 9 | Left ribs | -4.41 (-4.72 to -4.11) | -98 (-103 to -94) | <0.0001 | 3933.42 | 8 | <0.0001 | <0.0001 |
| Left rib 10 | Left ribs | -4.57 (-4.88 to -4.26) | -108 (-112 to -104) | <0.0001 | 4565.49 | 8 | <0.0001 | <0.0001 |
| Left rib 11 | Left ribs | -4.75 (-5.06 to -4.43) | -111 (-116 to -107) | <0.0001 | 4533.59 | 9 | <0.0001 | <0.0001 |
| Left rib 12 | Left ribs | -3.64 (-3.84 to -3.45) | -98 (-101 to -94) | <0.0001 | 3251.13 | 4 | <0.0001 | <0.0001 |
| Right rib 1 | Right ribs | -2.47 (-2.69 to -2.24) | -64 (-68 to -60) | <0.0001 | 1885.85 | 7 | <0.0001 | <0.0001 |
| Right rib 2 | Right ribs | -2.91 (-3.07 to -2.74) | -89 (-91 to -86) | <0.0001 | 3965.85 | 4 | <0.0001 | <0.0001 |
| Right rib 3 | Right ribs | -3.00 (-3.17 to -2.83) | -85 (-88 to -82) | <0.0001 | 3307.01 | 4 | <0.0001 | <0.0001 |
| Right rib 4 | Right ribs | -3.48 (-3.77 to -3.18) | -83 (-87 to -78) | <0.0001 | 2520.88 | 8 | <0.0001 | <0.0001 |
| Right rib 5 | Right ribs | -3.40 (-3.70 to -3.09) | -82 (-86 to -77) | <0.0001 | 2652.76 | 8 | <0.0001 | <0.0001 |
| Right rib 6 | Right ribs | -3.51 (-3.82 to -3.20) | -89 (-93 to -84) | <0.0001 | 3202.51 | 8 | <0.0001 | <0.0001 |
| Right rib 7 | Right ribs | -3.66 (-3.96 to -3.37) | -85 (-89 to -81) | <0.0001 | 3199.21 | 8 | <0.0001 | <0.0001 |
| Right rib 8 | Right ribs | -3.92 (-4.21 to -3.62) | -87 (-91 to -83) | <0.0001 | 3257.77 | 8 | <0.0001 | <0.0001 |
| Right rib 9 | Right ribs | -4.15 (-4.45 to -3.85) | -93 (-97 to -89) | <0.0001 | 3556.37 | 8 | <0.0001 | <0.0001 |
| Right rib 10 | Right ribs | -4.45 (-4.75 to -4.15) | -106 (-111 to -102) | <0.0001 | 4416.51 | 8 | <0.0001 | <0.0001 |
| Right rib 11 | Right ribs | -4.63 (-4.94 to -4.32) | -109 (-114 to -105) | <0.0001 | 4639.05 | 8 | <0.0001 | <0.0001 |
| Right rib 12 | Right ribs | -3.56 (-3.76 to -3.37) | -100 (-103 to -96) | <0.0001 | 3362.63 | 4 | <0.0001 | <0.0001 |

One row per site: each statistic compares women with men at that site, so it is not defined separately within a sex. Excess female change is the difference between two independently estimated contrasts, women minus men, so a negative value means a steeper decline in women. The sex × age interaction is a joint Wald test of the interaction block in a model pooling both sexes. p values are controlled across the 49 sites by the Benjamini–Hochberg procedure at a 5% false discovery rate.

**Supplementary Table 3: Reference centiles of CT attenuation by site, sex and age**

| **Site** | **Sex** | **Age (y)** | **3rd centile (HU)** | **10th centile (HU)** | **25th centile (HU)** | **50th centile (HU)** | **75th centile (HU)** | **90th centile (HU)** | **97th centile (HU)** |
| --- | --- | --- | --- | --- | --- | --- | --- | --- | --- |
| C1 | Female | 20 | 377 | 419 | 460 | 503 | 552 | 588 | 633 |
| C1 | Female | 30 | 375 | 422 | 460 | 504 | 550 | 600 | 643 |
| C1 | Female | 40 | 357 | 401 | 440 | 487 | 539 | 591 | 641 |
| C1 | Female | 50 | 330 | 367 | 408 | 457 | 514 | 564 | 622 |
| C1 | Female | 60 | 298 | 330 | 369 | 416 | 474 | 524 | 582 |
| C1 | Female | 70 | 272 | 302 | 336 | 379 | 429 | 483 | 534 |
| C1 | Female | 80 | 266 | 296 | 328 | 369 | 417 | 468 | 523 |
| C1 | Female | 90 | 265 | 293 | 326 | 370 | 421 | 474 | 542 |
| C1 | Male | 20 | 340 | 382 | 420 | 459 | 513 | 553 | 594 |
| C1 | Male | 30 | 337 | 372 | 409 | 455 | 502 | 544 | 592 |
| C1 | Male | 40 | 328 | 363 | 401 | 449 | 497 | 543 | 595 |
| C1 | Male | 50 | 318 | 355 | 395 | 443 | 496 | 547 | 600 |
| C1 | Male | 60 | 311 | 349 | 391 | 440 | 497 | 551 | 607 |
| C1 | Male | 70 | 310 | 346 | 389 | 440 | 498 | 554 | 614 |
| C1 | Male | 80 | 311 | 349 | 393 | 449 | 510 | 569 | 624 |
| C1 | Male | 90 | 320 | 361 | 410 | 461 | 522 | 577 | 625 |
| C2 | Female | 20 | 365 | 406 | 440 | 480 | 525 | 566 | 615 |
| C2 | Female | 30 | 348 | 395 | 439 | 488 | 535 | 587 | 631 |
| C2 | Female | 40 | 326 | 370 | 416 | 467 | 518 | 575 | 627 |
| C2 | Female | 50 | 300 | 337 | 379 | 427 | 482 | 537 | 599 |
| C2 | Female | 60 | 270 | 300 | 335 | 378 | 430 | 482 | 542 |
| C2 | Female | 70 | 242 | 268 | 297 | 336 | 381 | 429 | 479 |
| C2 | Female | 80 | 232 | 255 | 285 | 322 | 364 | 409 | 464 |
| C2 | Female | 90 | 220 | 249 | 280 | 315 | 356 | 404 | 468 |
| C2 | Male | 20 | 328 | 365 | 398 | 437 | 488 | 522 | 582 |
| C2 | Male | 30 | 317 | 350 | 386 | 429 | 477 | 519 | 571 |
| C2 | Male | 40 | 304 | 337 | 373 | 420 | 469 | 517 | 569 |
| C2 | Male | 50 | 289 | 324 | 360 | 410 | 462 | 514 | 571 |
| C2 | Male | 60 | 276 | 311 | 348 | 398 | 452 | 508 | 570 |
| C2 | Male | 70 | 264 | 298 | 337 | 386 | 440 | 500 | 562 |
| C2 | Male | 80 | 256 | 292 | 334 | 387 | 443 | 503 | 567 |
| C2 | Male | 90 | 261 | 297 | 343 | 394 | 446 | 500 | 555 |
| C3 | Female | 20 | 340 | 397 | 426 | 468 | 514 | 555 | 610 |
| C3 | Female | 30 | 336 | 378 | 421 | 468 | 518 | 565 | 611 |
| C3 | Female | 40 | 318 | 354 | 399 | 447 | 500 | 553 | 607 |
| C3 | Female | 50 | 292 | 326 | 366 | 413 | 467 | 522 | 587 |
| C3 | Female | 60 | 264 | 295 | 329 | 372 | 424 | 476 | 542 |
| C3 | Female | 70 | 242 | 268 | 297 | 336 | 384 | 429 | 484 |
| C3 | Female | 80 | 234 | 258 | 288 | 325 | 368 | 412 | 467 |
| C3 | Female | 90 | 229 | 257 | 289 | 327 | 371 | 418 | 475 |
| C3 | Male | 20 | 325 | 357 | 388 | 426 | 476 | 517 | 568 |
| C3 | Male | 30 | 305 | 336 | 369 | 413 | 463 | 503 | 549 |
| C3 | Male | 40 | 290 | 322 | 357 | 404 | 454 | 497 | 544 |
| C3 | Male | 50 | 279 | 312 | 350 | 397 | 448 | 497 | 549 |
| C3 | Male | 60 | 272 | 305 | 345 | 392 | 446 | 502 | 560 |
| C3 | Male | 70 | 266 | 300 | 339 | 389 | 447 | 509 | 574 |
| C3 | Male | 80 | 264 | 297 | 339 | 394 | 456 | 515 | 580 |
| C3 | Male | 90 | 274 | 306 | 346 | 402 | 456 | 519 | 569 |
| C4 | Female | 20 | 361 | 410 | 437 | 489 | 532 | 576 | 642 |
| C4 | Female | 30 | 355 | 399 | 440 | 491 | 541 | 592 | 648 |
| C4 | Female | 40 | 335 | 375 | 418 | 468 | 524 | 579 | 636 |
| C4 | Female | 50 | 307 | 343 | 383 | 431 | 489 | 545 | 608 |
| C4 | Female | 60 | 278 | 310 | 346 | 391 | 446 | 501 | 563 |
| C4 | Female | 70 | 257 | 286 | 318 | 360 | 408 | 459 | 515 |
| C4 | Female | 80 | 254 | 279 | 312 | 352 | 400 | 445 | 500 |
| C4 | Female | 90 | 251 | 279 | 312 | 351 | 401 | 451 | 505 |
| C4 | Male | 20 | 340 | 371 | 409 | 447 | 497 | 536 | 579 |
| C4 | Male | 30 | 320 | 354 | 385 | 429 | 478 | 517 | 566 |
| C4 | Male | 40 | 304 | 340 | 371 | 417 | 467 | 511 | 566 |
| C4 | Male | 50 | 293 | 328 | 363 | 411 | 463 | 514 | 575 |
| C4 | Male | 60 | 285 | 321 | 360 | 410 | 465 | 523 | 589 |
| C4 | Male | 70 | 282 | 319 | 360 | 411 | 471 | 534 | 603 |
| C4 | Male | 80 | 284 | 320 | 365 | 417 | 479 | 545 | 608 |
| C4 | Male | 90 | 290 | 325 | 367 | 421 | 478 | 537 | 594 |
| C5 | Female | 20 | 352 | 391 | 427 | 472 | 520 | 562 | 598 |
| C5 | Female | 30 | 342 | 388 | 428 | 477 | 527 | 569 | 629 |
| C5 | Female | 40 | 324 | 366 | 407 | 456 | 509 | 557 | 625 |
| C5 | Female | 50 | 299 | 334 | 373 | 421 | 475 | 529 | 594 |
| C5 | Female | 60 | 274 | 303 | 337 | 381 | 434 | 488 | 548 |
| C5 | Female | 70 | 252 | 279 | 309 | 348 | 397 | 446 | 501 |
| C5 | Female | 80 | 243 | 269 | 301 | 338 | 383 | 429 | 486 |
| C5 | Female | 90 | 236 | 266 | 299 | 336 | 382 | 428 | 482 |
| C5 | Male | 20 | 326 | 353 | 389 | 432 | 480 | 521 | 576 |
| C5 | Male | 30 | 306 | 341 | 369 | 411 | 457 | 501 | 550 |
| C5 | Male | 40 | 292 | 327 | 356 | 399 | 445 | 492 | 542 |
| C5 | Male | 50 | 282 | 316 | 350 | 394 | 443 | 492 | 547 |
| C5 | Male | 60 | 275 | 308 | 346 | 393 | 446 | 499 | 559 |
| C5 | Male | 70 | 272 | 306 | 345 | 392 | 451 | 509 | 572 |
| C5 | Male | 80 | 270 | 306 | 346 | 397 | 455 | 516 | 577 |
| C5 | Male | 90 | 275 | 305 | 343 | 394 | 445 | 504 | 559 |
| C6 | Female | 20 | 329 | 363 | 395 | 439 | 483 | 512 | 563 |
| C6 | Female | 30 | 323 | 361 | 401 | 444 | 487 | 527 | 578 |
| C6 | Female | 40 | 306 | 343 | 383 | 427 | 472 | 518 | 572 |
| C6 | Female | 50 | 282 | 315 | 352 | 395 | 442 | 492 | 548 |
| C6 | Female | 60 | 258 | 285 | 318 | 358 | 405 | 453 | 508 |
| C6 | Female | 70 | 239 | 261 | 291 | 327 | 368 | 414 | 465 |
| C6 | Female | 80 | 228 | 251 | 279 | 315 | 355 | 397 | 449 |
| C6 | Female | 90 | 222 | 246 | 274 | 311 | 352 | 392 | 452 |
| C6 | Male | 20 | 303 | 334 | 367 | 401 | 440 | 472 | 522 |
| C6 | Male | 30 | 286 | 317 | 343 | 379 | 420 | 458 | 494 |
| C6 | Male | 40 | 274 | 304 | 331 | 369 | 412 | 453 | 488 |
| C6 | Male | 50 | 266 | 296 | 326 | 366 | 411 | 455 | 497 |
| C6 | Male | 60 | 261 | 290 | 323 | 366 | 415 | 462 | 514 |
| C6 | Male | 70 | 256 | 285 | 320 | 365 | 418 | 472 | 530 |
| C6 | Male | 80 | 249 | 282 | 319 | 366 | 419 | 476 | 534 |
| C6 | Male | 90 | 245 | 281 | 315 | 362 | 412 | 460 | 519 |
| C7 | Female | 20 | 284 | 318 | 349 | 382 | 418 | 452 | 495 |
| C7 | Female | 30 | 287 | 320 | 352 | 390 | 430 | 469 | 508 |
| C7 | Female | 40 | 272 | 304 | 336 | 376 | 418 | 459 | 502 |
| C7 | Female | 50 | 249 | 279 | 310 | 349 | 391 | 432 | 480 |
| C7 | Female | 60 | 226 | 252 | 281 | 317 | 357 | 397 | 446 |
| C7 | Female | 70 | 210 | 232 | 258 | 291 | 326 | 364 | 407 |
| C7 | Female | 80 | 200 | 222 | 247 | 277 | 312 | 347 | 393 |
| C7 | Female | 90 | 191 | 214 | 239 | 269 | 305 | 340 | 389 |
| C7 | Male | 20 | 267 | 291 | 317 | 349 | 385 | 418 | 461 |
| C7 | Male | 30 | 251 | 274 | 301 | 334 | 372 | 407 | 435 |
| C7 | Male | 40 | 241 | 266 | 294 | 328 | 367 | 404 | 434 |
| C7 | Male | 50 | 233 | 261 | 291 | 326 | 368 | 406 | 447 |
| C7 | Male | 60 | 228 | 258 | 289 | 326 | 371 | 413 | 464 |
| C7 | Male | 70 | 224 | 253 | 286 | 327 | 373 | 421 | 472 |
| C7 | Male | 80 | 222 | 250 | 283 | 326 | 373 | 422 | 473 |
| C7 | Male | 90 | 217 | 246 | 278 | 317 | 362 | 404 | 456 |
| T1 | Female | 20 | 258 | 295 | 322 | 349 | 382 | 411 | 441 |
| T1 | Female | 30 | 264 | 293 | 325 | 359 | 395 | 430 | 463 |
| T1 | Female | 40 | 250 | 276 | 308 | 344 | 380 | 416 | 453 |
| T1 | Female | 50 | 226 | 251 | 279 | 313 | 349 | 383 | 423 |
| T1 | Female | 60 | 202 | 225 | 249 | 280 | 313 | 345 | 385 |
| T1 | Female | 70 | 185 | 204 | 226 | 253 | 283 | 314 | 351 |
| T1 | Female | 80 | 175 | 194 | 214 | 240 | 269 | 298 | 334 |
| T1 | Female | 90 | 165 | 186 | 206 | 231 | 260 | 291 | 329 |
| T1 | Male | 20 | 236 | 265 | 288 | 318 | 348 | 383 | 410 |
| T1 | Male | 30 | 226 | 252 | 277 | 306 | 339 | 371 | 400 |
| T1 | Male | 40 | 217 | 242 | 268 | 298 | 331 | 363 | 395 |
| T1 | Male | 50 | 208 | 233 | 260 | 291 | 325 | 359 | 395 |
| T1 | Male | 60 | 199 | 225 | 253 | 285 | 320 | 357 | 397 |
| T1 | Male | 70 | 192 | 217 | 244 | 279 | 317 | 356 | 401 |
| T1 | Male | 80 | 185 | 209 | 237 | 274 | 314 | 356 | 402 |
| T1 | Male | 90 | 180 | 202 | 230 | 264 | 302 | 345 | 386 |
| T2 | Female | 20 | 232 | 283 | 305 | 335 | 361 | 388 | 409 |
| T2 | Female | 30 | 242 | 275 | 303 | 336 | 370 | 401 | 427 |
| T2 | Female | 40 | 230 | 256 | 286 | 319 | 355 | 388 | 421 |
| T2 | Female | 50 | 207 | 233 | 259 | 291 | 325 | 358 | 397 |
| T2 | Female | 60 | 185 | 209 | 232 | 261 | 292 | 324 | 363 |
| T2 | Female | 70 | 171 | 189 | 210 | 236 | 266 | 296 | 329 |
| T2 | Female | 80 | 161 | 178 | 199 | 223 | 251 | 281 | 316 |
| T2 | Female | 90 | 155 | 171 | 191 | 215 | 242 | 274 | 310 |
| T2 | Male | 20 | 234 | 256 | 278 | 305 | 331 | 364 | 405 |
| T2 | Male | 30 | 217 | 241 | 261 | 288 | 316 | 344 | 381 |
| T2 | Male | 40 | 204 | 228 | 250 | 277 | 307 | 335 | 370 |
| T2 | Male | 50 | 195 | 217 | 242 | 270 | 302 | 333 | 368 |
| T2 | Male | 60 | 186 | 208 | 235 | 265 | 298 | 333 | 371 |
| T2 | Male | 70 | 177 | 200 | 227 | 259 | 295 | 332 | 376 |
| T2 | Male | 80 | 170 | 194 | 220 | 254 | 294 | 334 | 383 |
| T2 | Male | 90 | 167 | 187 | 213 | 248 | 286 | 325 | 376 |
| T3 | Female | 20 | 255 | 284 | 305 | 336 | 362 | 385 | 418 |
| T3 | Female | 30 | 245 | 273 | 301 | 336 | 369 | 398 | 429 |
| T3 | Female | 40 | 226 | 253 | 283 | 317 | 352 | 385 | 420 |
| T3 | Female | 50 | 202 | 227 | 256 | 287 | 323 | 356 | 395 |
| T3 | Female | 60 | 180 | 202 | 227 | 255 | 289 | 321 | 360 |
| T3 | Female | 70 | 164 | 182 | 203 | 230 | 260 | 290 | 325 |
| T3 | Female | 80 | 154 | 172 | 191 | 217 | 245 | 276 | 308 |
| T3 | Female | 90 | 146 | 164 | 182 | 207 | 235 | 266 | 299 |
| T3 | Male | 20 | 234 | 254 | 278 | 309 | 335 | 366 | 409 |
| T3 | Male | 30 | 217 | 240 | 264 | 291 | 321 | 348 | 382 |
| T3 | Male | 40 | 204 | 227 | 253 | 280 | 311 | 340 | 372 |
| T3 | Male | 50 | 193 | 217 | 243 | 271 | 305 | 337 | 373 |
| T3 | Male | 60 | 183 | 207 | 234 | 265 | 300 | 336 | 377 |
| T3 | Male | 70 | 174 | 197 | 224 | 258 | 296 | 334 | 380 |
| T3 | Male | 80 | 167 | 190 | 217 | 251 | 292 | 335 | 384 |
| T3 | Male | 90 | 161 | 183 | 209 | 243 | 282 | 322 | 371 |
| T4 | Female | 20 | 240 | 271 | 297 | 327 | 356 | 383 | 414 |
| T4 | Female | 30 | 239 | 269 | 297 | 331 | 364 | 394 | 426 |
| T4 | Female | 40 | 222 | 250 | 279 | 313 | 347 | 380 | 416 |
| T4 | Female | 50 | 197 | 223 | 251 | 282 | 317 | 350 | 390 |
| T4 | Female | 60 | 175 | 197 | 222 | 250 | 282 | 315 | 354 |
| T4 | Female | 70 | 160 | 178 | 198 | 224 | 253 | 284 | 318 |
| T4 | Female | 80 | 151 | 168 | 187 | 211 | 239 | 269 | 300 |
| T4 | Female | 90 | 143 | 160 | 179 | 202 | 229 | 258 | 288 |
| T4 | Male | 20 | 232 | 254 | 281 | 308 | 334 | 363 | 401 |
| T4 | Male | 30 | 216 | 239 | 264 | 290 | 318 | 347 | 377 |
| T4 | Male | 40 | 203 | 226 | 251 | 278 | 309 | 339 | 368 |
| T4 | Male | 50 | 192 | 215 | 240 | 270 | 302 | 334 | 368 |
| T4 | Male | 60 | 182 | 205 | 231 | 263 | 298 | 332 | 373 |
| T4 | Male | 70 | 172 | 195 | 221 | 255 | 292 | 331 | 377 |
| T4 | Male | 80 | 164 | 186 | 213 | 247 | 287 | 330 | 380 |
| T4 | Male | 90 | 158 | 179 | 205 | 237 | 275 | 317 | 366 |
| T5 | Female | 20 | 241 | 269 | 296 | 323 | 353 | 377 | 404 |
| T5 | Female | 30 | 239 | 267 | 294 | 326 | 358 | 387 | 419 |
| T5 | Female | 40 | 220 | 248 | 275 | 307 | 340 | 372 | 408 |
| T5 | Female | 50 | 195 | 221 | 247 | 277 | 310 | 342 | 380 |
| T5 | Female | 60 | 172 | 194 | 218 | 245 | 277 | 308 | 345 |
| T5 | Female | 70 | 160 | 176 | 196 | 221 | 250 | 279 | 313 |
| T5 | Female | 80 | 151 | 166 | 185 | 209 | 235 | 264 | 295 |
| T5 | Female | 90 | 142 | 158 | 177 | 200 | 227 | 254 | 287 |
| T5 | Male | 20 | 223 | 250 | 278 | 303 | 330 | 358 | 395 |
| T5 | Male | 30 | 213 | 235 | 260 | 286 | 314 | 343 | 369 |
| T5 | Male | 40 | 201 | 223 | 247 | 274 | 304 | 332 | 359 |
| T5 | Male | 50 | 190 | 213 | 237 | 266 | 298 | 327 | 360 |
| T5 | Male | 60 | 179 | 203 | 229 | 259 | 293 | 326 | 366 |
| T5 | Male | 70 | 170 | 193 | 220 | 252 | 288 | 328 | 373 |
| T5 | Male | 80 | 163 | 185 | 211 | 245 | 284 | 328 | 379 |
| T5 | Male | 90 | 156 | 177 | 202 | 234 | 273 | 312 | 360 |
| T6 | Female | 20 | 244 | 270 | 294 | 320 | 348 | 376 | 396 |
| T6 | Female | 30 | 235 | 262 | 290 | 323 | 355 | 386 | 414 |
| T6 | Female | 40 | 216 | 243 | 271 | 304 | 338 | 370 | 404 |
| T6 | Female | 50 | 193 | 219 | 244 | 274 | 307 | 339 | 376 |
| T6 | Female | 60 | 172 | 194 | 216 | 243 | 274 | 306 | 342 |
| T6 | Female | 70 | 157 | 174 | 195 | 220 | 250 | 281 | 314 |
| T6 | Female | 80 | 149 | 164 | 184 | 208 | 236 | 265 | 297 |
| T6 | Female | 90 | 139 | 157 | 176 | 200 | 227 | 256 | 287 |
| T6 | Male | 20 | 221 | 250 | 278 | 302 | 329 | 353 | 387 |
| T6 | Male | 30 | 213 | 236 | 260 | 286 | 315 | 340 | 367 |
| T6 | Male | 40 | 202 | 224 | 247 | 274 | 304 | 332 | 359 |
| T6 | Male | 50 | 190 | 213 | 237 | 266 | 298 | 328 | 360 |
| T6 | Male | 60 | 179 | 202 | 229 | 259 | 293 | 327 | 366 |
| T6 | Male | 70 | 169 | 192 | 219 | 252 | 289 | 327 | 374 |
| T6 | Male | 80 | 162 | 184 | 211 | 245 | 286 | 328 | 379 |
| T6 | Male | 90 | 155 | 177 | 202 | 233 | 272 | 312 | 362 |
| T7 | Female | 20 | 229 | 260 | 285 | 313 | 344 | 369 | 393 |
| T7 | Female | 30 | 228 | 256 | 284 | 316 | 348 | 379 | 406 |
| T7 | Female | 40 | 212 | 238 | 266 | 298 | 330 | 363 | 395 |
| T7 | Female | 50 | 189 | 214 | 239 | 268 | 301 | 333 | 369 |
| T7 | Female | 60 | 168 | 190 | 212 | 239 | 270 | 301 | 338 |
| T7 | Female | 70 | 155 | 172 | 192 | 217 | 246 | 276 | 312 |
| T7 | Female | 80 | 148 | 163 | 182 | 206 | 233 | 265 | 300 |
| T7 | Female | 90 | 140 | 155 | 174 | 197 | 225 | 259 | 293 |
| T7 | Male | 20 | 225 | 249 | 273 | 299 | 327 | 353 | 382 |
| T7 | Male | 30 | 208 | 234 | 256 | 282 | 310 | 337 | 363 |
| T7 | Male | 40 | 195 | 221 | 243 | 270 | 299 | 328 | 356 |
| T7 | Male | 50 | 185 | 210 | 233 | 261 | 293 | 323 | 357 |
| T7 | Male | 60 | 176 | 200 | 224 | 255 | 289 | 323 | 363 |
| T7 | Male | 70 | 167 | 190 | 217 | 249 | 285 | 325 | 371 |
| T7 | Male | 80 | 159 | 181 | 208 | 243 | 283 | 325 | 375 |
| T7 | Male | 90 | 153 | 174 | 199 | 230 | 268 | 306 | 354 |
| T8 | Female | 20 | 224 | 252 | 284 | 310 | 339 | 366 | 399 |
| T8 | Female | 30 | 226 | 252 | 282 | 312 | 345 | 374 | 404 |
| T8 | Female | 40 | 209 | 235 | 262 | 293 | 326 | 357 | 392 |
| T8 | Female | 50 | 184 | 209 | 233 | 263 | 295 | 328 | 367 |
| T8 | Female | 60 | 163 | 184 | 206 | 233 | 264 | 297 | 335 |
| T8 | Female | 70 | 151 | 167 | 188 | 212 | 242 | 273 | 306 |
| T8 | Female | 80 | 145 | 159 | 178 | 202 | 231 | 263 | 299 |
| T8 | Female | 90 | 135 | 152 | 171 | 194 | 224 | 257 | 297 |
| T8 | Male | 20 | 212 | 245 | 271 | 295 | 325 | 350 | 373 |
| T8 | Male | 30 | 202 | 231 | 253 | 279 | 307 | 334 | 358 |
| T8 | Male | 40 | 192 | 218 | 240 | 267 | 296 | 324 | 353 |
| T8 | Male | 50 | 182 | 206 | 230 | 258 | 289 | 319 | 355 |
| T8 | Male | 60 | 173 | 196 | 221 | 252 | 285 | 318 | 360 |
| T8 | Male | 70 | 165 | 187 | 214 | 246 | 283 | 322 | 367 |
| T8 | Male | 80 | 157 | 179 | 206 | 240 | 280 | 324 | 373 |
| T8 | Male | 90 | 151 | 170 | 196 | 227 | 267 | 305 | 357 |
| T9 | Female | 20 | 226 | 262 | 290 | 317 | 345 | 374 | 398 |
| T9 | Female | 30 | 229 | 258 | 287 | 319 | 351 | 382 | 410 |
| T9 | Female | 40 | 212 | 239 | 266 | 299 | 332 | 364 | 398 |
| T9 | Female | 50 | 186 | 212 | 238 | 268 | 301 | 334 | 371 |
| T9 | Female | 60 | 165 | 187 | 210 | 237 | 268 | 301 | 339 |
| T9 | Female | 70 | 154 | 170 | 190 | 216 | 246 | 276 | 313 |
| T9 | Female | 80 | 147 | 162 | 181 | 206 | 235 | 267 | 301 |
| T9 | Female | 90 | 139 | 155 | 175 | 200 | 230 | 263 | 297 |
| T9 | Male | 20 | 222 | 251 | 276 | 303 | 331 | 355 | 394 |
| T9 | Male | 30 | 206 | 234 | 257 | 284 | 312 | 338 | 367 |
| T9 | Male | 40 | 193 | 220 | 243 | 270 | 300 | 327 | 356 |
| T9 | Male | 50 | 182 | 208 | 233 | 262 | 292 | 322 | 358 |
| T9 | Male | 60 | 174 | 199 | 225 | 255 | 288 | 322 | 364 |
| T9 | Male | 70 | 166 | 190 | 216 | 249 | 286 | 326 | 370 |
| T9 | Male | 80 | 159 | 182 | 209 | 243 | 284 | 328 | 376 |
| T9 | Male | 90 | 153 | 173 | 199 | 232 | 271 | 311 | 359 |
| T10 | Female | 20 | 236 | 262 | 291 | 319 | 347 | 372 | 404 |
| T10 | Female | 30 | 235 | 260 | 287 | 319 | 352 | 382 | 414 |
| T10 | Female | 40 | 214 | 240 | 266 | 298 | 332 | 365 | 400 |
| T10 | Female | 50 | 187 | 212 | 237 | 266 | 299 | 333 | 372 |
| T10 | Female | 60 | 164 | 185 | 208 | 235 | 266 | 299 | 338 |
| T10 | Female | 70 | 152 | 167 | 188 | 214 | 243 | 273 | 309 |
| T10 | Female | 80 | 146 | 160 | 179 | 204 | 233 | 264 | 300 |
| T10 | Female | 90 | 143 | 155 | 174 | 198 | 227 | 261 | 301 |
| T10 | Male | 20 | 216 | 254 | 276 | 307 | 335 | 360 | 397 |
| T10 | Male | 30 | 202 | 235 | 256 | 284 | 311 | 341 | 371 |
| T10 | Male | 40 | 191 | 218 | 241 | 268 | 296 | 327 | 356 |
| T10 | Male | 50 | 181 | 205 | 230 | 257 | 288 | 318 | 352 |
| T10 | Male | 60 | 172 | 194 | 220 | 250 | 283 | 316 | 356 |
| T10 | Male | 70 | 163 | 186 | 212 | 245 | 281 | 321 | 366 |
| T10 | Male | 80 | 156 | 178 | 205 | 239 | 279 | 322 | 370 |
| T10 | Male | 90 | 151 | 170 | 196 | 228 | 267 | 309 | 354 |
| T11 | Female | 20 | 221 | 255 | 281 | 309 | 337 | 363 | 392 |
| T11 | Female | 30 | 225 | 251 | 278 | 309 | 342 | 371 | 401 |
| T11 | Female | 40 | 206 | 230 | 257 | 288 | 321 | 352 | 386 |
| T11 | Female | 50 | 179 | 202 | 227 | 256 | 288 | 320 | 357 |
| T11 | Female | 60 | 155 | 176 | 198 | 224 | 254 | 286 | 323 |
| T11 | Female | 70 | 142 | 159 | 177 | 203 | 231 | 261 | 296 |
| T11 | Female | 80 | 136 | 151 | 169 | 194 | 223 | 252 | 288 |
| T11 | Female | 90 | 130 | 146 | 165 | 189 | 219 | 253 | 294 |
| T11 | Male | 20 | 218 | 247 | 270 | 297 | 326 | 351 | 386 |
| T11 | Male | 30 | 200 | 226 | 249 | 274 | 303 | 330 | 359 |
| T11 | Male | 40 | 184 | 209 | 232 | 257 | 286 | 314 | 343 |
| T11 | Male | 50 | 172 | 194 | 218 | 245 | 274 | 304 | 336 |
| T11 | Male | 60 | 161 | 183 | 207 | 236 | 268 | 300 | 337 |
| T11 | Male | 70 | 152 | 174 | 199 | 230 | 265 | 302 | 346 |
| T11 | Male | 80 | 145 | 167 | 193 | 225 | 264 | 305 | 351 |
| T11 | Male | 90 | 143 | 161 | 185 | 218 | 254 | 294 | 340 |
| T12 | Female | 20 | 214 | 244 | 269 | 297 | 323 | 348 | 374 |
| T12 | Female | 30 | 215 | 240 | 266 | 295 | 324 | 352 | 380 |
| T12 | Female | 40 | 196 | 219 | 244 | 272 | 302 | 332 | 364 |
| T12 | Female | 50 | 168 | 190 | 214 | 240 | 270 | 299 | 334 |
| T12 | Female | 60 | 145 | 165 | 185 | 209 | 237 | 266 | 301 |
| T12 | Female | 70 | 133 | 148 | 166 | 189 | 215 | 243 | 278 |
| T12 | Female | 80 | 128 | 142 | 159 | 182 | 209 | 239 | 280 |
| T12 | Female | 90 | 121 | 139 | 157 | 179 | 210 | 246 | 296 |
| T12 | Male | 20 | 213 | 239 | 261 | 284 | 314 | 339 | 380 |
| T12 | Male | 30 | 190 | 215 | 238 | 261 | 290 | 314 | 346 |
| T12 | Male | 40 | 173 | 196 | 219 | 243 | 271 | 295 | 324 |
| T12 | Male | 50 | 160 | 181 | 204 | 228 | 256 | 283 | 313 |
| T12 | Male | 60 | 150 | 170 | 192 | 218 | 248 | 278 | 313 |
| T12 | Male | 70 | 141 | 161 | 184 | 212 | 245 | 280 | 321 |
| T12 | Male | 80 | 134 | 154 | 177 | 208 | 243 | 283 | 325 |
| T12 | Male | 90 | 131 | 150 | 172 | 204 | 239 | 276 | 322 |
| L1 | Female | 20 | 222 | 246 | 273 | 299 | 329 | 352 | 381 |
| L1 | Female | 30 | 217 | 244 | 269 | 296 | 326 | 354 | 383 |
| L1 | Female | 40 | 198 | 222 | 246 | 273 | 302 | 332 | 363 |
| L1 | Female | 50 | 172 | 194 | 216 | 241 | 269 | 298 | 332 |
| L1 | Female | 60 | 150 | 168 | 188 | 211 | 238 | 266 | 301 |
| L1 | Female | 70 | 138 | 153 | 171 | 193 | 219 | 248 | 284 |
| L1 | Female | 80 | 132 | 147 | 165 | 188 | 216 | 249 | 291 |
| L1 | Female | 90 | 128 | 144 | 162 | 186 | 218 | 255 | 307 |
| L1 | Male | 20 | 211 | 240 | 262 | 284 | 313 | 339 | 367 |
| L1 | Male | 30 | 191 | 217 | 238 | 263 | 289 | 313 | 341 |
| L1 | Male | 40 | 175 | 198 | 219 | 244 | 270 | 294 | 321 |
| L1 | Male | 50 | 163 | 184 | 205 | 230 | 256 | 283 | 311 |
| L1 | Male | 60 | 154 | 173 | 195 | 220 | 249 | 280 | 312 |
| L1 | Male | 70 | 145 | 166 | 188 | 216 | 250 | 285 | 325 |
| L1 | Male | 80 | 139 | 159 | 183 | 213 | 251 | 291 | 334 |
| L1 | Male | 90 | 135 | 155 | 179 | 210 | 247 | 285 | 334 |
| L2 | Female | 20 | 232 | 263 | 289 | 316 | 346 | 371 | 398 |
| L2 | Female | 30 | 230 | 259 | 283 | 312 | 343 | 370 | 397 |
| L2 | Female | 40 | 210 | 235 | 258 | 287 | 318 | 346 | 376 |
| L2 | Female | 50 | 182 | 204 | 227 | 253 | 283 | 313 | 348 |
| L2 | Female | 60 | 159 | 178 | 199 | 223 | 252 | 282 | 321 |
| L2 | Female | 70 | 147 | 163 | 183 | 206 | 233 | 265 | 305 |
| L2 | Female | 80 | 141 | 157 | 176 | 200 | 230 | 266 | 308 |
| L2 | Female | 90 | 132 | 151 | 169 | 195 | 229 | 268 | 316 |
| L2 | Male | 20 | 220 | 246 | 272 | 298 | 327 | 357 | 386 |
| L2 | Male | 30 | 201 | 226 | 249 | 275 | 301 | 327 | 357 |
| L2 | Male | 40 | 186 | 209 | 230 | 255 | 281 | 307 | 335 |
| L2 | Male | 50 | 175 | 195 | 216 | 241 | 268 | 295 | 326 |
| L2 | Male | 60 | 165 | 184 | 206 | 233 | 263 | 295 | 330 |
| L2 | Male | 70 | 157 | 179 | 202 | 232 | 267 | 305 | 349 |
| L2 | Male | 80 | 150 | 172 | 197 | 230 | 269 | 312 | 362 |
| L2 | Male | 90 | 147 | 165 | 191 | 223 | 263 | 308 | 358 |
| L3 | Female | 20 | 242 | 270 | 297 | 323 | 352 | 378 | 401 |
| L3 | Female | 30 | 237 | 265 | 291 | 319 | 350 | 378 | 401 |
| L3 | Female | 40 | 215 | 241 | 265 | 294 | 325 | 354 | 382 |
| L3 | Female | 50 | 187 | 210 | 232 | 259 | 289 | 319 | 353 |
| L3 | Female | 60 | 163 | 182 | 203 | 228 | 256 | 286 | 324 |
| L3 | Female | 70 | 149 | 166 | 186 | 209 | 235 | 266 | 305 |
| L3 | Female | 80 | 141 | 158 | 178 | 201 | 231 | 266 | 308 |
| L3 | Female | 90 | 134 | 151 | 170 | 195 | 228 | 265 | 315 |
| L3 | Male | 20 | 225 | 255 | 282 | 306 | 337 | 363 | 396 |
| L3 | Male | 30 | 209 | 234 | 257 | 282 | 310 | 335 | 360 |
| L3 | Male | 40 | 194 | 216 | 238 | 263 | 289 | 314 | 340 |
| L3 | Male | 50 | 181 | 200 | 223 | 248 | 275 | 303 | 335 |
| L3 | Male | 60 | 170 | 190 | 213 | 240 | 270 | 302 | 342 |
| L3 | Male | 70 | 162 | 184 | 208 | 238 | 274 | 312 | 359 |
| L3 | Male | 80 | 155 | 177 | 203 | 236 | 277 | 319 | 370 |
| L3 | Male | 90 | 149 | 167 | 195 | 229 | 270 | 312 | 363 |
| L4 | Female | 20 | 242 | 270 | 296 | 322 | 351 | 375 | 399 |
| L4 | Female | 30 | 241 | 268 | 294 | 323 | 354 | 381 | 407 |
| L4 | Female | 40 | 220 | 245 | 270 | 299 | 331 | 360 | 392 |
| L4 | Female | 50 | 191 | 214 | 238 | 264 | 296 | 327 | 365 |
| L4 | Female | 60 | 166 | 187 | 208 | 233 | 264 | 295 | 337 |
| L4 | Female | 70 | 152 | 170 | 190 | 215 | 244 | 277 | 320 |
| L4 | Female | 80 | 144 | 162 | 182 | 207 | 239 | 277 | 324 |
| L4 | Female | 90 | 133 | 154 | 176 | 202 | 235 | 277 | 319 |
| L4 | Male | 20 | 231 | 260 | 284 | 309 | 338 | 364 | 389 |
| L4 | Male | 30 | 214 | 239 | 260 | 285 | 313 | 339 | 369 |
| L4 | Male | 40 | 198 | 220 | 241 | 266 | 293 | 321 | 353 |
| L4 | Male | 50 | 184 | 205 | 227 | 253 | 282 | 311 | 348 |
| L4 | Male | 60 | 173 | 195 | 219 | 247 | 279 | 313 | 356 |
| L4 | Male | 70 | 167 | 190 | 216 | 248 | 286 | 328 | 381 |
| L4 | Male | 80 | 161 | 185 | 212 | 247 | 290 | 337 | 390 |
| L4 | Male | 90 | 153 | 175 | 203 | 237 | 280 | 322 | 377 |
| L5 | Female | 20 | 239 | 262 | 291 | 317 | 349 | 374 | 400 |
| L5 | Female | 30 | 234 | 263 | 290 | 322 | 355 | 381 | 409 |
| L5 | Female | 40 | 214 | 242 | 268 | 300 | 334 | 363 | 398 |
| L5 | Female | 50 | 186 | 211 | 236 | 266 | 300 | 333 | 375 |
| L5 | Female | 60 | 162 | 184 | 207 | 234 | 267 | 303 | 349 |
| L5 | Female | 70 | 148 | 167 | 189 | 215 | 247 | 284 | 330 |
| L5 | Female | 80 | 141 | 159 | 181 | 208 | 242 | 283 | 330 |
| L5 | Female | 90 | 129 | 151 | 173 | 201 | 236 | 275 | 321 |
| L5 | Male | 20 | 230 | 257 | 282 | 310 | 337 | 365 | 401 |
| L5 | Male | 30 | 209 | 235 | 257 | 285 | 314 | 342 | 371 |
| L5 | Male | 40 | 193 | 217 | 238 | 266 | 296 | 326 | 360 |
| L5 | Male | 50 | 180 | 203 | 226 | 254 | 286 | 320 | 364 |
| L5 | Male | 60 | 171 | 193 | 219 | 250 | 286 | 326 | 377 |
| L5 | Male | 70 | 164 | 189 | 216 | 252 | 295 | 342 | 396 |
| L5 | Male | 80 | 158 | 183 | 212 | 250 | 298 | 347 | 402 |
| L5 | Male | 90 | 148 | 172 | 201 | 238 | 284 | 329 | 375 |
| S1 | Female | 20 | 238 | 268 | 301 | 330 | 360 | 393 | 422 |
| S1 | Female | 30 | 241 | 272 | 303 | 337 | 371 | 405 | 437 |
| S1 | Female | 40 | 224 | 254 | 283 | 318 | 354 | 389 | 426 |
| S1 | Female | 50 | 196 | 224 | 251 | 284 | 320 | 356 | 398 |
| S1 | Female | 60 | 166 | 191 | 216 | 246 | 280 | 316 | 359 |
| S1 | Female | 70 | 141 | 164 | 187 | 215 | 246 | 281 | 321 |
| S1 | Female | 80 | 127 | 147 | 169 | 197 | 229 | 264 | 306 |
| S1 | Female | 90 | 111 | 132 | 155 | 184 | 217 | 251 | 291 |
| S1 | Male | 20 | 226 | 255 | 285 | 315 | 346 | 378 | 414 |
| S1 | Male | 30 | 215 | 244 | 270 | 299 | 332 | 362 | 397 |
| S1 | Male | 40 | 201 | 230 | 254 | 283 | 317 | 349 | 385 |
| S1 | Male | 50 | 185 | 213 | 238 | 268 | 303 | 338 | 378 |
| S1 | Male | 60 | 169 | 195 | 222 | 254 | 291 | 329 | 375 |
| S1 | Male | 70 | 153 | 180 | 208 | 242 | 282 | 323 | 374 |
| S1 | Male | 80 | 143 | 166 | 195 | 231 | 274 | 319 | 371 |
| S1 | Male | 90 | 133 | 154 | 182 | 217 | 261 | 305 | 353 |
| Left rib 1 | Female | 20 | 285 | 317 | 346 | 385 | 423 | 462 | 508 |
| Left rib 1 | Female | 30 | 286 | 321 | 350 | 390 | 430 | 469 | 512 |
| Left rib 1 | Female | 40 | 269 | 302 | 332 | 371 | 411 | 452 | 495 |
| Left rib 1 | Female | 50 | 240 | 270 | 300 | 337 | 377 | 416 | 463 |
| Left rib 1 | Female | 60 | 209 | 235 | 263 | 297 | 334 | 372 | 418 |
| Left rib 1 | Female | 70 | 182 | 206 | 230 | 261 | 294 | 329 | 371 |
| Left rib 1 | Female | 80 | 167 | 188 | 211 | 240 | 272 | 307 | 350 |
| Left rib 1 | Female | 90 | 158 | 179 | 201 | 228 | 261 | 296 | 341 |
| Left rib 1 | Male | 20 | 270 | 303 | 327 | 362 | 400 | 432 | 462 |
| Left rib 1 | Male | 30 | 265 | 292 | 320 | 354 | 392 | 424 | 456 |
| Left rib 1 | Male | 40 | 251 | 277 | 306 | 341 | 379 | 414 | 452 |
| Left rib 1 | Male | 50 | 231 | 258 | 287 | 324 | 364 | 403 | 448 |
| Left rib 1 | Male | 60 | 210 | 238 | 266 | 304 | 346 | 391 | 443 |
| Left rib 1 | Male | 70 | 192 | 217 | 245 | 283 | 328 | 378 | 435 |
| Left rib 1 | Male | 80 | 179 | 202 | 231 | 268 | 314 | 366 | 427 |
| Left rib 1 | Male | 90 | 172 | 194 | 220 | 255 | 301 | 353 | 410 |
| Left rib 2 | Female | 20 | 273 | 314 | 344 | 382 | 424 | 470 | 505 |
| Left rib 2 | Female | 30 | 251 | 284 | 319 | 359 | 403 | 442 | 486 |
| Left rib 2 | Female | 40 | 219 | 248 | 282 | 320 | 363 | 401 | 448 |
| Left rib 2 | Female | 50 | 185 | 213 | 242 | 277 | 316 | 355 | 400 |
| Left rib 2 | Female | 60 | 157 | 182 | 207 | 238 | 274 | 311 | 351 |
| Left rib 2 | Female | 70 | 139 | 161 | 184 | 212 | 245 | 276 | 314 |
| Left rib 2 | Female | 80 | 131 | 150 | 172 | 199 | 229 | 261 | 299 |
| Left rib 2 | Female | 90 | 125 | 144 | 164 | 191 | 219 | 252 | 292 |
| Left rib 2 | Male | 20 | 236 | 266 | 298 | 337 | 373 | 403 | 457 |
| Left rib 2 | Male | 30 | 201 | 230 | 257 | 292 | 325 | 357 | 387 |
| Left rib 2 | Male | 40 | 177 | 204 | 230 | 262 | 294 | 326 | 353 |
| Left rib 2 | Male | 50 | 161 | 185 | 212 | 243 | 276 | 309 | 344 |
| Left rib 2 | Male | 60 | 150 | 173 | 200 | 232 | 267 | 303 | 347 |
| Left rib 2 | Male | 70 | 142 | 165 | 191 | 225 | 264 | 304 | 351 |
| Left rib 2 | Male | 80 | 136 | 160 | 188 | 224 | 266 | 311 | 361 |
| Left rib 2 | Male | 90 | 133 | 156 | 185 | 219 | 260 | 301 | 352 |
| Left rib 3 | Female | 20 | 283 | 331 | 369 | 409 | 449 | 495 | 553 |
| Left rib 3 | Female | 30 | 250 | 295 | 338 | 383 | 428 | 472 | 523 |
| Left rib 3 | Female | 40 | 212 | 250 | 289 | 333 | 378 | 422 | 471 |
| Left rib 3 | Female | 50 | 175 | 205 | 238 | 276 | 318 | 361 | 410 |
| Left rib 3 | Female | 60 | 146 | 170 | 197 | 229 | 267 | 306 | 350 |
| Left rib 3 | Female | 70 | 129 | 151 | 176 | 204 | 237 | 271 | 306 |
| Left rib 3 | Female | 80 | 122 | 143 | 166 | 193 | 223 | 255 | 291 |
| Left rib 3 | Female | 90 | 119 | 138 | 158 | 186 | 217 | 249 | 283 |
| Left rib 3 | Male | 20 | 238 | 275 | 306 | 350 | 395 | 432 | 472 |
| Left rib 3 | Male | 30 | 194 | 227 | 258 | 296 | 337 | 374 | 411 |
| Left rib 3 | Male | 40 | 165 | 194 | 223 | 257 | 296 | 332 | 369 |
| Left rib 3 | Male | 50 | 146 | 171 | 198 | 231 | 268 | 303 | 343 |
| Left rib 3 | Male | 60 | 134 | 157 | 183 | 215 | 252 | 287 | 330 |
| Left rib 3 | Male | 70 | 124 | 148 | 175 | 207 | 245 | 283 | 327 |
| Left rib 3 | Male | 80 | 119 | 143 | 171 | 204 | 245 | 286 | 332 |
| Left rib 3 | Male | 90 | 118 | 140 | 166 | 199 | 236 | 279 | 322 |
| Left rib 4 | Female | 20 | 225 | 290 | 356 | 412 | 466 | 521 | 577 |
| Left rib 4 | Female | 30 | 206 | 265 | 326 | 382 | 438 | 488 | 542 |
| Left rib 4 | Female | 40 | 182 | 230 | 280 | 332 | 386 | 435 | 489 |
| Left rib 4 | Female | 50 | 156 | 193 | 232 | 277 | 326 | 374 | 429 |
| Left rib 4 | Female | 60 | 136 | 164 | 195 | 232 | 275 | 318 | 368 |
| Left rib 4 | Female | 70 | 123 | 148 | 175 | 207 | 244 | 280 | 321 |
| Left rib 4 | Female | 80 | 118 | 142 | 168 | 199 | 233 | 269 | 309 |
| Left rib 4 | Female | 90 | 117 | 141 | 164 | 194 | 230 | 265 | 307 |
| Left rib 4 | Male | 20 | 216 | 271 | 313 | 363 | 408 | 451 | 492 |
| Left rib 4 | Male | 30 | 186 | 224 | 261 | 305 | 348 | 391 | 434 |
| Left rib 4 | Male | 40 | 162 | 192 | 225 | 264 | 305 | 346 | 391 |
| Left rib 4 | Male | 50 | 143 | 171 | 201 | 237 | 277 | 317 | 362 |
| Left rib 4 | Male | 60 | 130 | 158 | 186 | 221 | 261 | 301 | 347 |
| Left rib 4 | Male | 70 | 123 | 151 | 179 | 215 | 254 | 299 | 347 |
| Left rib 4 | Male | 80 | 121 | 146 | 176 | 213 | 256 | 302 | 351 |
| Left rib 4 | Male | 90 | 116 | 142 | 172 | 206 | 250 | 296 | 333 |
| Left rib 5 | Female | 20 | 216 | 259 | 328 | 402 | 462 | 521 | 585 |
| Left rib 5 | Female | 30 | 191 | 238 | 301 | 373 | 435 | 492 | 550 |
| Left rib 5 | Female | 40 | 163 | 208 | 263 | 327 | 387 | 442 | 498 |
| Left rib 5 | Female | 50 | 137 | 178 | 224 | 277 | 331 | 383 | 438 |
| Left rib 5 | Female | 60 | 119 | 154 | 192 | 235 | 281 | 328 | 379 |
| Left rib 5 | Female | 70 | 111 | 141 | 172 | 209 | 248 | 288 | 331 |
| Left rib 5 | Female | 80 | 111 | 137 | 166 | 200 | 237 | 274 | 317 |
| Left rib 5 | Female | 90 | 111 | 136 | 164 | 195 | 233 | 269 | 310 |
| Left rib 5 | Male | 20 | 197 | 245 | 297 | 353 | 405 | 452 | 499 |
| Left rib 5 | Male | 30 | 168 | 207 | 249 | 298 | 348 | 389 | 439 |
| Left rib 5 | Male | 40 | 146 | 179 | 216 | 259 | 305 | 345 | 396 |
| Left rib 5 | Male | 50 | 131 | 162 | 195 | 234 | 277 | 318 | 367 |
| Left rib 5 | Male | 60 | 121 | 152 | 183 | 221 | 262 | 305 | 353 |
| Left rib 5 | Male | 70 | 117 | 147 | 178 | 216 | 259 | 303 | 351 |
| Left rib 5 | Male | 80 | 115 | 145 | 177 | 216 | 262 | 309 | 360 |
| Left rib 5 | Male | 90 | 114 | 143 | 174 | 210 | 258 | 303 | 349 |
| Left rib 6 | Female | 20 | 225 | 271 | 329 | 402 | 461 | 516 | 600 |
| Left rib 6 | Female | 30 | 203 | 248 | 307 | 377 | 439 | 494 | 559 |
| Left rib 6 | Female | 40 | 175 | 219 | 274 | 336 | 397 | 453 | 511 |
| Left rib 6 | Female | 50 | 147 | 189 | 237 | 291 | 347 | 402 | 457 |
| Left rib 6 | Female | 60 | 125 | 163 | 204 | 250 | 300 | 350 | 402 |
| Left rib 6 | Female | 70 | 112 | 146 | 182 | 223 | 265 | 308 | 355 |
| Left rib 6 | Female | 80 | 112 | 144 | 176 | 212 | 251 | 291 | 334 |
| Left rib 6 | Female | 90 | 118 | 145 | 176 | 209 | 245 | 287 | 329 |
| Left rib 6 | Male | 20 | 208 | 245 | 294 | 348 | 403 | 452 | 510 |
| Left rib 6 | Male | 30 | 173 | 210 | 254 | 302 | 352 | 396 | 446 |
| Left rib 6 | Male | 40 | 149 | 185 | 224 | 268 | 315 | 356 | 405 |
| Left rib 6 | Male | 50 | 134 | 169 | 205 | 245 | 290 | 332 | 382 |
| Left rib 6 | Male | 60 | 126 | 160 | 193 | 233 | 277 | 322 | 373 |
| Left rib 6 | Male | 70 | 123 | 155 | 189 | 229 | 275 | 322 | 373 |
| Left rib 6 | Male | 80 | 119 | 152 | 186 | 229 | 276 | 325 | 377 |
| Left rib 6 | Male | 90 | 117 | 149 | 181 | 223 | 269 | 318 | 367 |
| Left rib 7 | Female | 20 | 274 | 317 | 369 | 426 | 481 | 545 | 619 |
| Left rib 7 | Female | 30 | 241 | 288 | 343 | 401 | 459 | 515 | 576 |
| Left rib 7 | Female | 40 | 208 | 252 | 302 | 356 | 414 | 468 | 523 |
| Left rib 7 | Female | 50 | 177 | 215 | 257 | 306 | 361 | 414 | 466 |
| Left rib 7 | Female | 60 | 152 | 184 | 219 | 262 | 311 | 360 | 410 |
| Left rib 7 | Female | 70 | 136 | 165 | 197 | 235 | 277 | 319 | 367 |
| Left rib 7 | Female | 80 | 131 | 159 | 190 | 225 | 265 | 306 | 352 |
| Left rib 7 | Female | 90 | 132 | 158 | 187 | 223 | 261 | 304 | 348 |
| Left rib 7 | Male | 20 | 240 | 276 | 321 | 371 | 420 | 467 | 518 |
| Left rib 7 | Male | 30 | 196 | 233 | 272 | 317 | 364 | 407 | 453 |
| Left rib 7 | Male | 40 | 169 | 204 | 238 | 279 | 324 | 366 | 411 |
| Left rib 7 | Male | 50 | 153 | 185 | 217 | 255 | 298 | 340 | 387 |
| Left rib 7 | Male | 60 | 144 | 173 | 204 | 242 | 285 | 328 | 378 |
| Left rib 7 | Male | 70 | 135 | 164 | 197 | 236 | 280 | 328 | 378 |
| Left rib 7 | Male | 80 | 128 | 159 | 192 | 231 | 279 | 328 | 380 |
| Left rib 7 | Male | 90 | 128 | 159 | 188 | 227 | 273 | 320 | 364 |
| Left rib 8 | Female | 20 | 322 | 374 | 419 | 465 | 521 | 582 | 652 |
| Left rib 8 | Female | 30 | 277 | 329 | 379 | 435 | 491 | 546 | 607 |
| Left rib 8 | Female | 40 | 233 | 279 | 326 | 381 | 437 | 492 | 550 |
| Left rib 8 | Female | 50 | 194 | 232 | 272 | 321 | 375 | 430 | 488 |
| Left rib 8 | Female | 60 | 166 | 197 | 230 | 272 | 320 | 370 | 426 |
| Left rib 8 | Female | 70 | 149 | 176 | 205 | 243 | 284 | 325 | 377 |
| Left rib 8 | Female | 80 | 140 | 166 | 195 | 231 | 269 | 309 | 355 |
| Left rib 8 | Female | 90 | 134 | 159 | 188 | 222 | 262 | 303 | 348 |
| Left rib 8 | Male | 20 | 274 | 311 | 355 | 402 | 450 | 495 | 541 |
| Left rib 8 | Male | 30 | 223 | 256 | 297 | 342 | 390 | 436 | 480 |
| Left rib 8 | Male | 40 | 189 | 221 | 258 | 300 | 347 | 392 | 438 |
| Left rib 8 | Male | 50 | 168 | 199 | 234 | 274 | 319 | 364 | 412 |
| Left rib 8 | Male | 60 | 156 | 185 | 218 | 257 | 303 | 349 | 400 |
| Left rib 8 | Male | 70 | 147 | 176 | 208 | 248 | 296 | 344 | 398 |
| Left rib 8 | Male | 80 | 136 | 166 | 198 | 240 | 289 | 341 | 398 |
| Left rib 8 | Male | 90 | 130 | 159 | 191 | 233 | 280 | 331 | 388 |
| Left rib 9 | Female | 20 | 339 | 381 | 425 | 474 | 522 | 573 | 636 |
| Left rib 9 | Female | 30 | 295 | 342 | 389 | 445 | 499 | 546 | 604 |
| Left rib 9 | Female | 40 | 248 | 292 | 337 | 392 | 449 | 500 | 557 |
| Left rib 9 | Female | 50 | 207 | 243 | 282 | 331 | 387 | 442 | 501 |
| Left rib 9 | Female | 60 | 176 | 205 | 238 | 281 | 331 | 385 | 442 |
| Left rib 9 | Female | 70 | 160 | 185 | 213 | 250 | 294 | 338 | 391 |
| Left rib 9 | Female | 80 | 150 | 174 | 201 | 236 | 276 | 317 | 367 |
| Left rib 9 | Female | 90 | 141 | 165 | 192 | 227 | 267 | 308 | 359 |
| Left rib 9 | Male | 20 | 286 | 327 | 368 | 414 | 464 | 508 | 553 |
| Left rib 9 | Male | 30 | 240 | 274 | 311 | 356 | 406 | 448 | 495 |
| Left rib 9 | Male | 40 | 208 | 238 | 274 | 317 | 366 | 410 | 459 |
| Left rib 9 | Male | 50 | 186 | 215 | 250 | 293 | 340 | 389 | 440 |
| Left rib 9 | Male | 60 | 172 | 201 | 235 | 277 | 326 | 379 | 433 |
| Left rib 9 | Male | 70 | 160 | 190 | 223 | 266 | 320 | 373 | 433 |
| Left rib 9 | Male | 80 | 149 | 180 | 213 | 258 | 312 | 367 | 430 |
| Left rib 9 | Male | 90 | 143 | 171 | 205 | 251 | 298 | 354 | 410 |
| Left rib 10 | Female | 20 | 367 | 410 | 452 | 500 | 546 | 594 | 648 |
| Left rib 10 | Female | 30 | 327 | 375 | 423 | 476 | 530 | 577 | 625 |
| Left rib 10 | Female | 40 | 282 | 326 | 372 | 425 | 482 | 533 | 584 |
| Left rib 10 | Female | 50 | 238 | 276 | 315 | 365 | 420 | 474 | 531 |
| Left rib 10 | Female | 60 | 204 | 236 | 269 | 312 | 362 | 414 | 470 |
| Left rib 10 | Female | 70 | 185 | 212 | 241 | 278 | 321 | 365 | 417 |
| Left rib 10 | Female | 80 | 175 | 201 | 229 | 264 | 304 | 347 | 397 |
| Left rib 10 | Female | 90 | 170 | 196 | 222 | 257 | 298 | 339 | 392 |
| Left rib 10 | Male | 20 | 308 | 353 | 395 | 440 | 488 | 534 | 587 |
| Left rib 10 | Male | 30 | 261 | 299 | 337 | 384 | 431 | 477 | 526 |
| Left rib 10 | Male | 40 | 228 | 263 | 299 | 344 | 392 | 438 | 487 |
| Left rib 10 | Male | 50 | 206 | 240 | 275 | 318 | 367 | 414 | 466 |
| Left rib 10 | Male | 60 | 192 | 225 | 260 | 302 | 353 | 404 | 460 |
| Left rib 10 | Male | 70 | 182 | 214 | 249 | 293 | 347 | 403 | 464 |
| Left rib 10 | Male | 80 | 174 | 205 | 241 | 288 | 344 | 402 | 468 |
| Left rib 10 | Male | 90 | 169 | 198 | 235 | 281 | 338 | 395 | 462 |
| Left rib 11 | Female | 20 | 381 | 427 | 472 | 533 | 583 | 636 | 689 |
| Left rib 11 | Female | 30 | 340 | 389 | 438 | 497 | 554 | 611 | 663 |
| Left rib 11 | Female | 40 | 291 | 336 | 383 | 437 | 496 | 555 | 611 |
| Left rib 11 | Female | 50 | 244 | 282 | 322 | 370 | 427 | 483 | 543 |
| Left rib 11 | Female | 60 | 207 | 237 | 270 | 312 | 362 | 414 | 471 |
| Left rib 11 | Female | 70 | 184 | 210 | 238 | 276 | 317 | 363 | 414 |
| Left rib 11 | Female | 80 | 172 | 197 | 224 | 259 | 301 | 342 | 393 |
| Left rib 11 | Female | 90 | 167 | 189 | 218 | 252 | 295 | 339 | 393 |
| Left rib 11 | Male | 20 | 331 | 373 | 414 | 461 | 512 | 559 | 604 |
| Left rib 11 | Male | 30 | 279 | 319 | 357 | 404 | 456 | 503 | 549 |
| Left rib 11 | Male | 40 | 245 | 280 | 316 | 361 | 412 | 459 | 508 |
| Left rib 11 | Male | 50 | 222 | 253 | 288 | 331 | 379 | 428 | 482 |
| Left rib 11 | Male | 60 | 205 | 234 | 269 | 310 | 359 | 410 | 469 |
| Left rib 11 | Male | 70 | 190 | 219 | 254 | 297 | 350 | 405 | 470 |
| Left rib 11 | Male | 80 | 178 | 208 | 243 | 288 | 343 | 403 | 465 |
| Left rib 11 | Male | 90 | 174 | 201 | 234 | 279 | 333 | 394 | 457 |
| Left rib 12 | Female | 20 | 346 | 398 | 453 | 508 | 572 | 629 | 714 |
| Left rib 12 | Female | 30 | 319 | 364 | 421 | 481 | 547 | 602 | 675 |
| Left rib 12 | Female | 40 | 280 | 320 | 372 | 428 | 491 | 549 | 617 |
| Left rib 12 | Female | 50 | 238 | 274 | 316 | 366 | 424 | 483 | 550 |
| Left rib 12 | Female | 60 | 203 | 234 | 269 | 311 | 363 | 419 | 482 |
| Left rib 12 | Female | 70 | 180 | 206 | 237 | 276 | 322 | 372 | 430 |
| Left rib 12 | Female | 80 | 170 | 195 | 224 | 263 | 311 | 362 | 419 |
| Left rib 12 | Female | 90 | 163 | 190 | 219 | 259 | 313 | 369 | 428 |
| Left rib 12 | Male | 20 | 293 | 352 | 392 | 451 | 505 | 558 | 611 |
| Left rib 12 | Male | 30 | 260 | 301 | 340 | 390 | 442 | 490 | 539 |
| Left rib 12 | Male | 40 | 233 | 266 | 303 | 346 | 396 | 443 | 493 |
| Left rib 12 | Male | 50 | 212 | 242 | 276 | 317 | 365 | 413 | 468 |
| Left rib 12 | Male | 60 | 194 | 224 | 257 | 298 | 346 | 397 | 456 |
| Left rib 12 | Male | 70 | 180 | 209 | 242 | 284 | 337 | 390 | 454 |
| Left rib 12 | Male | 80 | 169 | 199 | 234 | 278 | 331 | 390 | 454 |
| Left rib 12 | Male | 90 | 163 | 193 | 229 | 273 | 327 | 387 | 458 |
| Right rib 1 | Female | 20 | 300 | 326 | 353 | 390 | 432 | 470 | 517 |
| Right rib 1 | Female | 30 | 294 | 325 | 357 | 393 | 432 | 470 | 514 |
| Right rib 1 | Female | 40 | 272 | 303 | 336 | 372 | 411 | 451 | 496 |
| Right rib 1 | Female | 50 | 242 | 269 | 302 | 337 | 376 | 417 | 463 |
| Right rib 1 | Female | 60 | 210 | 234 | 263 | 296 | 333 | 371 | 416 |
| Right rib 1 | Female | 70 | 184 | 206 | 230 | 258 | 291 | 324 | 364 |
| Right rib 1 | Female | 80 | 168 | 189 | 210 | 237 | 269 | 302 | 344 |
| Right rib 1 | Female | 90 | 158 | 178 | 200 | 227 | 258 | 295 | 334 |
| Right rib 1 | Male | 20 | 285 | 314 | 332 | 369 | 406 | 440 | 496 |
| Right rib 1 | Male | 30 | 267 | 295 | 322 | 357 | 392 | 426 | 473 |
| Right rib 1 | Male | 40 | 250 | 276 | 306 | 342 | 378 | 415 | 459 |
| Right rib 1 | Male | 50 | 231 | 257 | 288 | 324 | 363 | 404 | 449 |
| Right rib 1 | Male | 60 | 212 | 237 | 267 | 304 | 346 | 392 | 441 |
| Right rib 1 | Male | 70 | 192 | 216 | 246 | 284 | 328 | 377 | 433 |
| Right rib 1 | Male | 80 | 179 | 203 | 231 | 269 | 312 | 365 | 427 |
| Right rib 1 | Male | 90 | 174 | 195 | 219 | 254 | 298 | 350 | 405 |
| Right rib 2 | Female | 20 | 280 | 315 | 346 | 389 | 433 | 477 | 517 |
| Right rib 2 | Female | 30 | 252 | 289 | 322 | 363 | 406 | 448 | 490 |
| Right rib 2 | Female | 40 | 218 | 252 | 285 | 323 | 365 | 406 | 449 |
| Right rib 2 | Female | 50 | 185 | 214 | 243 | 280 | 318 | 358 | 402 |
| Right rib 2 | Female | 60 | 157 | 182 | 208 | 241 | 275 | 312 | 353 |
| Right rib 2 | Female | 70 | 139 | 162 | 185 | 213 | 245 | 277 | 314 |
| Right rib 2 | Female | 80 | 132 | 151 | 173 | 199 | 230 | 263 | 300 |
| Right rib 2 | Female | 90 | 128 | 145 | 166 | 191 | 222 | 255 | 291 |
| Right rib 2 | Male | 20 | 240 | 271 | 301 | 339 | 377 | 412 | 454 |
| Right rib 2 | Male | 30 | 203 | 232 | 259 | 293 | 328 | 361 | 393 |
| Right rib 2 | Male | 40 | 179 | 204 | 230 | 263 | 296 | 329 | 360 |
| Right rib 2 | Male | 50 | 162 | 186 | 212 | 244 | 277 | 312 | 347 |
| Right rib 2 | Male | 60 | 151 | 174 | 200 | 233 | 268 | 305 | 346 |
| Right rib 2 | Male | 70 | 142 | 165 | 192 | 227 | 265 | 306 | 351 |
| Right rib 2 | Male | 80 | 136 | 160 | 189 | 225 | 267 | 312 | 359 |
| Right rib 2 | Male | 90 | 132 | 156 | 183 | 218 | 259 | 302 | 348 |
| Right rib 3 | Female | 20 | 288 | 333 | 370 | 414 | 456 | 506 | 556 |
| Right rib 3 | Female | 30 | 253 | 298 | 336 | 384 | 431 | 477 | 526 |
| Right rib 3 | Female | 40 | 213 | 251 | 288 | 333 | 379 | 424 | 473 |
| Right rib 3 | Female | 50 | 175 | 204 | 237 | 276 | 318 | 362 | 410 |
| Right rib 3 | Female | 60 | 146 | 169 | 197 | 229 | 266 | 306 | 351 |
| Right rib 3 | Female | 70 | 131 | 152 | 176 | 204 | 236 | 269 | 308 |
| Right rib 3 | Female | 80 | 124 | 144 | 166 | 193 | 223 | 255 | 289 |
| Right rib 3 | Female | 90 | 120 | 139 | 159 | 187 | 216 | 248 | 280 |
| Right rib 3 | Male | 20 | 240 | 271 | 311 | 355 | 389 | 431 | 476 |
| Right rib 3 | Male | 30 | 197 | 226 | 257 | 297 | 337 | 370 | 412 |
| Right rib 3 | Male | 40 | 166 | 192 | 220 | 256 | 295 | 326 | 367 |
| Right rib 3 | Male | 50 | 144 | 169 | 195 | 228 | 265 | 298 | 338 |
| Right rib 3 | Male | 60 | 131 | 154 | 180 | 212 | 246 | 283 | 324 |
| Right rib 3 | Male | 70 | 123 | 146 | 172 | 204 | 240 | 279 | 323 |
| Right rib 3 | Male | 80 | 118 | 141 | 168 | 201 | 240 | 281 | 325 |
| Right rib 3 | Male | 90 | 116 | 137 | 164 | 195 | 233 | 272 | 316 |
| Right rib 4 | Female | 20 | 241 | 299 | 360 | 417 | 467 | 521 | 586 |
| Right rib 4 | Female | 30 | 213 | 269 | 330 | 388 | 444 | 493 | 552 |
| Right rib 4 | Female | 40 | 184 | 232 | 284 | 337 | 392 | 441 | 497 |
| Right rib 4 | Female | 50 | 157 | 196 | 235 | 281 | 331 | 379 | 433 |
| Right rib 4 | Female | 60 | 137 | 167 | 198 | 236 | 279 | 323 | 373 |
| Right rib 4 | Female | 70 | 126 | 151 | 180 | 212 | 249 | 286 | 331 |
| Right rib 4 | Female | 80 | 123 | 147 | 174 | 205 | 240 | 275 | 318 |
| Right rib 4 | Female | 90 | 123 | 145 | 170 | 200 | 236 | 272 | 312 |
| Right rib 4 | Male | 20 | 221 | 271 | 311 | 362 | 406 | 449 | 497 |
| Right rib 4 | Male | 30 | 188 | 224 | 260 | 305 | 350 | 391 | 430 |
| Right rib 4 | Male | 40 | 162 | 191 | 223 | 264 | 306 | 345 | 385 |
| Right rib 4 | Male | 50 | 142 | 169 | 199 | 236 | 274 | 314 | 357 |
| Right rib 4 | Male | 60 | 130 | 156 | 185 | 220 | 256 | 297 | 344 |
| Right rib 4 | Male | 70 | 125 | 150 | 178 | 214 | 253 | 296 | 341 |
| Right rib 4 | Male | 80 | 119 | 146 | 176 | 214 | 256 | 301 | 346 |
| Right rib 4 | Male | 90 | 119 | 143 | 170 | 207 | 248 | 291 | 332 |
| Right rib 5 | Female | 20 | 204 | 252 | 324 | 398 | 456 | 516 | 580 |
| Right rib 5 | Female | 30 | 183 | 227 | 298 | 369 | 431 | 489 | 545 |
| Right rib 5 | Female | 40 | 157 | 201 | 263 | 326 | 386 | 442 | 495 |
| Right rib 5 | Female | 50 | 133 | 176 | 226 | 280 | 333 | 386 | 439 |
| Right rib 5 | Female | 60 | 116 | 155 | 195 | 240 | 286 | 334 | 384 |
| Right rib 5 | Female | 70 | 108 | 140 | 175 | 215 | 255 | 296 | 342 |
| Right rib 5 | Female | 80 | 106 | 137 | 171 | 206 | 245 | 284 | 330 |
| Right rib 5 | Female | 90 | 105 | 139 | 169 | 202 | 241 | 278 | 321 |
| Right rib 5 | Male | 20 | 200 | 243 | 292 | 344 | 392 | 434 | 486 |
| Right rib 5 | Male | 30 | 167 | 208 | 247 | 293 | 341 | 383 | 428 |
| Right rib 5 | Male | 40 | 145 | 181 | 215 | 257 | 302 | 344 | 386 |
| Right rib 5 | Male | 50 | 131 | 163 | 195 | 234 | 276 | 317 | 361 |
| Right rib 5 | Male | 60 | 123 | 153 | 184 | 222 | 262 | 303 | 350 |
| Right rib 5 | Male | 70 | 118 | 150 | 180 | 219 | 260 | 304 | 352 |
| Right rib 5 | Male | 80 | 116 | 148 | 181 | 220 | 265 | 312 | 362 |
| Right rib 5 | Male | 90 | 114 | 146 | 178 | 218 | 261 | 305 | 357 |
| Right rib 6 | Female | 20 | 232 | 279 | 349 | 411 | 466 | 518 | 598 |
| Right rib 6 | Female | 30 | 210 | 255 | 316 | 385 | 444 | 499 | 563 |
| Right rib 6 | Female | 40 | 182 | 225 | 279 | 344 | 403 | 459 | 517 |
| Right rib 6 | Female | 50 | 154 | 195 | 242 | 297 | 353 | 407 | 463 |
| Right rib 6 | Female | 60 | 131 | 169 | 209 | 255 | 305 | 355 | 407 |
| Right rib 6 | Female | 70 | 119 | 152 | 187 | 227 | 270 | 313 | 359 |
| Right rib 6 | Female | 80 | 117 | 148 | 179 | 216 | 254 | 294 | 340 |
| Right rib 6 | Female | 90 | 119 | 147 | 177 | 210 | 247 | 285 | 330 |
| Right rib 6 | Male | 20 | 216 | 252 | 298 | 350 | 401 | 448 | 492 |
| Right rib 6 | Male | 30 | 177 | 215 | 256 | 302 | 353 | 398 | 444 |
| Right rib 6 | Male | 40 | 152 | 189 | 226 | 267 | 316 | 361 | 407 |
| Right rib 6 | Male | 50 | 138 | 172 | 206 | 245 | 291 | 335 | 383 |
| Right rib 6 | Male | 60 | 130 | 161 | 194 | 233 | 277 | 323 | 371 |
| Right rib 6 | Male | 70 | 124 | 155 | 189 | 230 | 274 | 321 | 372 |
| Right rib 6 | Male | 80 | 121 | 153 | 188 | 230 | 278 | 326 | 381 |
| Right rib 6 | Male | 90 | 116 | 150 | 183 | 224 | 270 | 318 | 370 |
| Right rib 7 | Female | 20 | 277 | 322 | 375 | 432 | 482 | 536 | 610 |
| Right rib 7 | Female | 30 | 246 | 290 | 345 | 404 | 460 | 511 | 574 |
| Right rib 7 | Female | 40 | 212 | 254 | 305 | 360 | 417 | 470 | 526 |
| Right rib 7 | Female | 50 | 181 | 220 | 262 | 311 | 365 | 419 | 471 |
| Right rib 7 | Female | 60 | 157 | 190 | 226 | 268 | 316 | 366 | 416 |
| Right rib 7 | Female | 70 | 141 | 170 | 202 | 239 | 281 | 323 | 371 |
| Right rib 7 | Female | 80 | 134 | 162 | 193 | 229 | 268 | 309 | 355 |
| Right rib 7 | Female | 90 | 130 | 160 | 190 | 224 | 263 | 308 | 353 |
| Right rib 7 | Male | 20 | 245 | 283 | 322 | 370 | 417 | 460 | 520 |
| Right rib 7 | Male | 30 | 204 | 237 | 274 | 317 | 364 | 405 | 453 |
| Right rib 7 | Male | 40 | 176 | 207 | 241 | 281 | 326 | 367 | 412 |
| Right rib 7 | Male | 50 | 158 | 187 | 220 | 258 | 302 | 343 | 389 |
| Right rib 7 | Male | 60 | 146 | 176 | 208 | 245 | 288 | 331 | 381 |
| Right rib 7 | Male | 70 | 138 | 168 | 199 | 239 | 283 | 329 | 381 |
| Right rib 7 | Male | 80 | 130 | 161 | 193 | 233 | 282 | 330 | 386 |
| Right rib 7 | Male | 90 | 129 | 156 | 190 | 227 | 274 | 322 | 370 |
| Right rib 8 | Female | 20 | 319 | 364 | 407 | 457 | 512 | 562 | 627 |
| Right rib 8 | Female | 30 | 274 | 324 | 374 | 428 | 486 | 536 | 591 |
| Right rib 8 | Female | 40 | 234 | 279 | 326 | 379 | 436 | 488 | 542 |
| Right rib 8 | Female | 50 | 199 | 236 | 276 | 324 | 377 | 431 | 487 |
| Right rib 8 | Female | 60 | 172 | 202 | 236 | 278 | 325 | 376 | 431 |
| Right rib 8 | Female | 70 | 155 | 183 | 213 | 250 | 290 | 334 | 384 |
| Right rib 8 | Female | 80 | 146 | 172 | 201 | 237 | 277 | 317 | 364 |
| Right rib 8 | Female | 90 | 138 | 164 | 193 | 229 | 269 | 309 | 357 |
| Right rib 8 | Male | 20 | 274 | 307 | 353 | 397 | 449 | 495 | 541 |
| Right rib 8 | Male | 30 | 222 | 257 | 294 | 338 | 387 | 430 | 473 |
| Right rib 8 | Male | 40 | 189 | 223 | 256 | 298 | 345 | 387 | 431 |
| Right rib 8 | Male | 50 | 170 | 201 | 233 | 273 | 318 | 362 | 408 |
| Right rib 8 | Male | 60 | 158 | 188 | 219 | 258 | 303 | 349 | 399 |
| Right rib 8 | Male | 70 | 149 | 178 | 209 | 249 | 295 | 343 | 397 |
| Right rib 8 | Male | 80 | 140 | 169 | 200 | 241 | 290 | 341 | 397 |
| Right rib 8 | Male | 90 | 137 | 162 | 195 | 235 | 282 | 333 | 387 |
| Right rib 9 | Female | 20 | 329 | 377 | 422 | 468 | 517 | 565 | 622 |
| Right rib 9 | Female | 30 | 291 | 341 | 388 | 439 | 495 | 541 | 596 |
| Right rib 9 | Female | 40 | 249 | 293 | 337 | 388 | 446 | 496 | 552 |
| Right rib 9 | Female | 50 | 210 | 245 | 283 | 332 | 387 | 441 | 500 |
| Right rib 9 | Female | 60 | 182 | 210 | 243 | 285 | 335 | 387 | 445 |
| Right rib 9 | Female | 70 | 167 | 192 | 222 | 259 | 302 | 346 | 399 |
| Right rib 9 | Female | 80 | 158 | 183 | 211 | 246 | 287 | 330 | 378 |
| Right rib 9 | Female | 90 | 150 | 174 | 200 | 236 | 277 | 318 | 367 |
| Right rib 9 | Male | 20 | 277 | 319 | 365 | 409 | 458 | 500 | 548 |
| Right rib 9 | Male | 30 | 234 | 268 | 307 | 351 | 402 | 440 | 485 |
| Right rib 9 | Male | 40 | 204 | 235 | 269 | 313 | 362 | 403 | 449 |
| Right rib 9 | Male | 50 | 184 | 214 | 247 | 289 | 337 | 383 | 433 |
| Right rib 9 | Male | 60 | 171 | 201 | 234 | 276 | 323 | 374 | 429 |
| Right rib 9 | Male | 70 | 163 | 192 | 225 | 267 | 317 | 372 | 430 |
| Right rib 9 | Male | 80 | 154 | 184 | 217 | 262 | 315 | 370 | 431 |
| Right rib 9 | Male | 90 | 152 | 178 | 211 | 253 | 302 | 360 | 417 |
| Right rib 10 | Female | 20 | 370 | 408 | 450 | 494 | 541 | 589 | 654 |
| Right rib 10 | Female | 30 | 331 | 377 | 422 | 476 | 527 | 575 | 629 |
| Right rib 10 | Female | 40 | 286 | 330 | 373 | 427 | 481 | 532 | 586 |
| Right rib 10 | Female | 50 | 243 | 279 | 319 | 367 | 421 | 476 | 532 |
| Right rib 10 | Female | 60 | 211 | 241 | 276 | 317 | 367 | 419 | 476 |
| Right rib 10 | Female | 70 | 193 | 221 | 252 | 289 | 333 | 376 | 430 |
| Right rib 10 | Female | 80 | 184 | 210 | 240 | 276 | 317 | 360 | 411 |
| Right rib 10 | Female | 90 | 179 | 203 | 230 | 267 | 308 | 354 | 408 |
| Right rib 10 | Male | 20 | 308 | 346 | 388 | 431 | 480 | 524 | 569 |
| Right rib 10 | Male | 30 | 261 | 295 | 335 | 379 | 428 | 472 | 517 |
| Right rib 10 | Male | 40 | 229 | 263 | 299 | 342 | 390 | 436 | 484 |
| Right rib 10 | Male | 50 | 208 | 242 | 276 | 318 | 366 | 414 | 466 |
| Right rib 10 | Male | 60 | 196 | 229 | 262 | 305 | 353 | 404 | 460 |
| Right rib 10 | Male | 70 | 189 | 219 | 254 | 299 | 350 | 404 | 465 |
| Right rib 10 | Male | 80 | 183 | 213 | 249 | 296 | 352 | 408 | 470 |
| Right rib 10 | Male | 90 | 180 | 209 | 244 | 289 | 345 | 402 | 461 |
| Right rib 11 | Female | 20 | 386 | 428 | 471 | 525 | 578 | 630 | 696 |
| Right rib 11 | Female | 30 | 342 | 389 | 437 | 495 | 552 | 605 | 658 |
| Right rib 11 | Female | 40 | 293 | 337 | 383 | 438 | 495 | 551 | 605 |
| Right rib 11 | Female | 50 | 247 | 284 | 324 | 372 | 427 | 482 | 542 |
| Right rib 11 | Female | 60 | 211 | 241 | 274 | 316 | 365 | 416 | 475 |
| Right rib 11 | Female | 70 | 189 | 214 | 245 | 282 | 324 | 368 | 420 |
| Right rib 11 | Female | 80 | 177 | 202 | 231 | 267 | 309 | 352 | 405 |
| Right rib 11 | Female | 90 | 170 | 196 | 225 | 261 | 306 | 352 | 405 |
| Right rib 11 | Male | 20 | 326 | 374 | 413 | 458 | 510 | 556 | 604 |
| Right rib 11 | Male | 30 | 279 | 317 | 356 | 401 | 455 | 497 | 549 |
| Right rib 11 | Male | 40 | 244 | 278 | 316 | 360 | 411 | 455 | 508 |
| Right rib 11 | Male | 50 | 221 | 253 | 289 | 332 | 379 | 428 | 481 |
| Right rib 11 | Male | 60 | 205 | 237 | 271 | 313 | 360 | 413 | 469 |
| Right rib 11 | Male | 70 | 195 | 224 | 259 | 302 | 354 | 408 | 470 |
| Right rib 11 | Male | 80 | 187 | 217 | 251 | 297 | 352 | 406 | 467 |
| Right rib 11 | Male | 90 | 187 | 216 | 248 | 291 | 344 | 401 | 466 |
| Right rib 12 | Female | 20 | 339 | 388 | 446 | 500 | 557 | 622 | 687 |
| Right rib 12 | Female | 30 | 313 | 361 | 415 | 473 | 535 | 595 | 660 |
| Right rib 12 | Female | 40 | 275 | 318 | 366 | 422 | 483 | 543 | 607 |
| Right rib 12 | Female | 50 | 235 | 270 | 312 | 364 | 420 | 481 | 543 |
| Right rib 12 | Female | 60 | 201 | 231 | 266 | 311 | 361 | 420 | 481 |
| Right rib 12 | Female | 70 | 179 | 206 | 238 | 277 | 322 | 376 | 439 |
| Right rib 12 | Female | 80 | 169 | 194 | 225 | 266 | 316 | 370 | 435 |
| Right rib 12 | Female | 90 | 161 | 190 | 222 | 267 | 323 | 382 | 450 |
| Right rib 12 | Male | 20 | 301 | 346 | 392 | 446 | 496 | 547 | 602 |
| Right rib 12 | Male | 30 | 260 | 297 | 336 | 384 | 435 | 487 | 538 |
| Right rib 12 | Male | 40 | 230 | 262 | 298 | 342 | 391 | 442 | 492 |
| Right rib 12 | Male | 50 | 209 | 238 | 273 | 314 | 362 | 412 | 464 |
| Right rib 12 | Male | 60 | 194 | 222 | 256 | 297 | 346 | 397 | 454 |
| Right rib 12 | Male | 70 | 183 | 211 | 245 | 288 | 341 | 395 | 460 |
| Right rib 12 | Male | 80 | 177 | 206 | 241 | 285 | 340 | 399 | 468 |
| Right rib 12 | Male | 90 | 174 | 202 | 237 | 283 | 339 | 403 | 471 |

Estimated by quantile regression on a cubic spline in age, separately for each site and sex. Values may be interpolated linearly between the tabulated ages.

**Supplementary Table 3B: Diagnostics of the centile fits**

| **Site** | **Sex** | **n** | **Grid points repaired for crossing, n** | **Quantile fits reaching the iteration cap, n** | **Quantile fits attempted, n** |
| --- | --- | --- | --- | --- | --- |
| C1 | Male | 12941 | 0 | 0 | 7 |
| C1 | Female | 13354 | 0 | 0 | 7 |
| C2 | Male | 12939 | 0 | 0 | 7 |
| C2 | Female | 13349 | 0 | 0 | 7 |
| C3 | Male | 13945 | 0 | 0 | 7 |
| C3 | Female | 14293 | 0 | 0 | 7 |
| C4 | Male | 15517 | 0 | 0 | 7 |
| C4 | Female | 15355 | 0 | 0 | 7 |
| C5 | Male | 19024 | 0 | 0 | 7 |
| C5 | Female | 17881 | 0 | 0 | 7 |
| C6 | Male | 25074 | 0 | 0 | 7 |
| C6 | Female | 22603 | 0 | 0 | 7 |
| C7 | Male | 28177 | 0 | 0 | 7 |
| C7 | Female | 25131 | 0 | 0 | 7 |
| T1 | Male | 28549 | 0 | 0 | 7 |
| T1 | Female | 25491 | 0 | 0 | 7 |
| T2 | Male | 28532 | 0 | 0 | 7 |
| T2 | Female | 25469 | 0 | 0 | 7 |
| T3 | Male | 28568 | 0 | 0 | 7 |
| T3 | Female | 25483 | 0 | 0 | 7 |
| T4 | Male | 28569 | 0 | 0 | 7 |
| T4 | Female | 25485 | 0 | 0 | 7 |
| T5 | Male | 28635 | 0 | 0 | 7 |
| T5 | Female | 25546 | 0 | 0 | 7 |
| T6 | Male | 29225 | 0 | 0 | 7 |
| T6 | Female | 26056 | 0 | 0 | 7 |
| T7 | Male | 31300 | 0 | 0 | 7 |
| T7 | Female | 27854 | 0 | 0 | 7 |
| T8 | Male | 34324 | 0 | 0 | 7 |
| T8 | Female | 30457 | 0 | 0 | 7 |
| T9 | Male | 37013 | 0 | 0 | 7 |
| T9 | Female | 33251 | 0 | 0 | 7 |
| T10 | Male | 38463 | 0 | 0 | 7 |
| T10 | Female | 35011 | 0 | 0 | 7 |
| T11 | Male | 38907 | 0 | 0 | 7 |
| T11 | Female | 35559 | 0 | 0 | 7 |
| T12 | Male | 38876 | 0 | 0 | 7 |
| T12 | Female | 35495 | 0 | 0 | 7 |
| L1 | Male | 38886 | 0 | 0 | 7 |
| L1 | Female | 35498 | 0 | 0 | 7 |
| L2 | Male | 38881 | 0 | 0 | 7 |
| L2 | Female | 35527 | 0 | 0 | 7 |
| L3 | Male | 38881 | 0 | 0 | 7 |
| L3 | Female | 35508 | 0 | 0 | 7 |
| L4 | Male | 38879 | 0 | 0 | 7 |
| L4 | Female | 35508 | 0 | 0 | 7 |
| L5 | Male | 38902 | 0 | 0 | 7 |
| L5 | Female | 35508 | 0 | 0 | 7 |
| S1 | Male | 38908 | 0 | 0 | 7 |
| S1 | Female | 35559 | 0 | 0 | 7 |
| Left rib 1 | Male | 28579 | 0 | 0 | 7 |
| Left rib 1 | Female | 25521 | 0 | 0 | 7 |
| Left rib 2 | Male | 28631 | 0 | 0 | 7 |
| Left rib 2 | Female | 25557 | 0 | 0 | 7 |
| Left rib 3 | Male | 29200 | 0 | 0 | 7 |
| Left rib 3 | Female | 26333 | 0 | 0 | 7 |
| Left rib 4 | Male | 32094 | 0 | 0 | 7 |
| Left rib 4 | Female | 29542 | 0 | 0 | 7 |
| Left rib 5 | Male | 36632 | 0 | 0 | 7 |
| Left rib 5 | Female | 33829 | 0 | 0 | 7 |
| Left rib 6 | Male | 38749 | 0 | 0 | 7 |
| Left rib 6 | Female | 35514 | 0 | 0 | 7 |
| Left rib 7 | Male | 38883 | 0 | 0 | 7 |
| Left rib 7 | Female | 35584 | 0 | 0 | 7 |
| Left rib 8 | Male | 38911 | 0 | 0 | 7 |
| Left rib 8 | Female | 35590 | 0 | 0 | 7 |
| Left rib 9 | Male | 38937 | 0 | 0 | 7 |
| Left rib 9 | Female | 35619 | 0 | 0 | 7 |
| Left rib 10 | Male | 38979 | 0 | 0 | 7 |
| Left rib 10 | Female | 35656 | 0 | 0 | 7 |
| Left rib 11 | Male | 38973 | 0 | 0 | 7 |
| Left rib 11 | Female | 35672 | 0 | 0 | 7 |
| Left rib 12 | Male | 38542 | 0 | 0 | 7 |
| Left rib 12 | Female | 34465 | 0 | 0 | 7 |
| Right rib 1 | Male | 28562 | 0 | 0 | 7 |
| Right rib 1 | Female | 25496 | 0 | 0 | 7 |
| Right rib 2 | Male | 28622 | 0 | 0 | 7 |
| Right rib 2 | Female | 25591 | 0 | 0 | 7 |
| Right rib 3 | Male | 29164 | 0 | 0 | 7 |
| Right rib 3 | Female | 26297 | 0 | 0 | 7 |
| Right rib 4 | Male | 31936 | 0 | 0 | 7 |
| Right rib 4 | Female | 29371 | 0 | 0 | 7 |
| Right rib 5 | Male | 36386 | 0 | 0 | 7 |
| Right rib 5 | Female | 33600 | 0 | 0 | 7 |
| Right rib 6 | Male | 38734 | 0 | 0 | 7 |
| Right rib 6 | Female | 35489 | 0 | 0 | 7 |
| Right rib 7 | Male | 38875 | 0 | 0 | 7 |
| Right rib 7 | Female | 35581 | 0 | 0 | 7 |
| Right rib 8 | Male | 38895 | 0 | 0 | 7 |
| Right rib 8 | Female | 35573 | 0 | 0 | 7 |
| Right rib 9 | Male | 38925 | 0 | 0 | 7 |
| Right rib 9 | Female | 35609 | 0 | 0 | 7 |
| Right rib 10 | Male | 38951 | 0 | 0 | 7 |
| Right rib 10 | Female | 35668 | 0 | 0 | 7 |
| Right rib 11 | Male | 38989 | 0 | 0 | 7 |
| Right rib 11 | Female | 35692 | 0 | 0 | 7 |
| Right rib 12 | Male | 38562 | 0 | 0 | 7 |
| Right rib 12 | Female | 34973 | 0 | 0 | 7 |

Separate quantile fits can cross where data are sparse; any crossing is repaired by a running maximum across increasing centile and counted here. Reaching the iteration cap is not an error: the returned coefficients are the best iterate, and every set of centiles was verified to be monotone.

**Supplementary Table 4: Prevalence of L1 attenuation below published trabecular cut-offs**

| **Site** | **Sex** | **Age group (y)** | **n** | **<100 HU, n** | **<100 HU, %** | **<100 HU, lower 95% limit, %** | **<100 HU, upper 95% limit, %** | **<150 HU, n** | **<150 HU, %** | **<150 HU, lower 95% limit, %** | **<150 HU, upper 95% limit, %** |
| --- | --- | --- | --- | --- | --- | --- | --- | --- | --- | --- | --- |
| L1 | Male | All ages | 38886 | 14 | 0 | 0 | 0.1 | 1499 | 3.9 | 3.7 | 4.1 |
| L1 | Male | 20-29 | 703 | 0 | 0 | 0 | 0.5 | 0 | 0 | 0 | 0.5 |
| L1 | Male | 30-39 | 1201 | 0 | 0 | 0 | 0.3 | 7 | 0.6 | 0.3 | 1.2 |
| L1 | Male | 40-49 | 2647 | 0 | 0 | 0 | 0.1 | 19 | 0.7 | 0.5 | 1.1 |
| L1 | Male | 50-59 | 5463 | 0 | 0 | 0 | 0.1 | 90 | 1.6 | 1.3 | 2 |
| L1 | Male | 60-69 | 8346 | 2 | 0 | 0 | 0.1 | 238 | 2.9 | 2.5 | 3.2 |
| L1 | Male | 70-79 | 13048 | 9 | 0.1 | 0 | 0.1 | 658 | 5 | 4.7 | 5.4 |
| L1 | Male | 80-89 | 6678 | 2 | 0 | 0 | 0.1 | 442 | 6.6 | 6 | 7.2 |
| L1 | Male | 90-99 | 631 | 1 | 0.2 | 0 | 0.9 | 45 | 7.1 | 5.4 | 9.4 |
| L1 | Female | All ages | 35498 | 8 | 0 | 0 | 0 | 2182 | 6.1 | 5.9 | 6.4 |
| L1 | Female | 20-29 | 837 | 0 | 0 | 0 | 0.5 | 2 | 0.2 | 0.1 | 0.9 |
| L1 | Female | 30-39 | 1578 | 0 | 0 | 0 | 0.2 | 2 | 0.1 | 0 | 0.5 |
| L1 | Female | 40-49 | 3781 | 0 | 0 | 0 | 0.1 | 10 | 0.3 | 0.1 | 0.5 |
| L1 | Female | 50-59 | 6675 | 0 | 0 | 0 | 0.1 | 106 | 1.6 | 1.3 | 1.9 |
| L1 | Female | 60-69 | 7008 | 1 | 0 | 0 | 0.1 | 361 | 5.2 | 4.7 | 5.7 |
| L1 | Female | 70-79 | 9309 | 4 | 0 | 0 | 0.1 | 915 | 9.8 | 9.2 | 10.5 |
| L1 | Female | 80-89 | 5326 | 2 | 0 | 0 | 0.1 | 659 | 12.4 | 11.5 | 13.3 |
| L1 | Female | 90-99 | 781 | 1 | 0.1 | 0 | 0.7 | 124 | 15.9 | 13.5 | 18.6 |

Intervals are Wilson 95% intervals. The 100 HU and 150 HU thresholds refer to a trabecular region of interest and are not transferable to the whole-mask attenuation measured here; this table documents that fact rather than estimating disease prevalence.

**Supplementary Table 5: Young-adult reference values**

| **Site** | **Sex** | **Young adults, n** | **Mean attenuation (HU)** | **SD (HU)** |
| --- | --- | --- | --- | --- |
| C1 | Male | 459 | 459.763 | 66.1025 |
| C1 | Female | 510 | 498.731 | 69.3443 |
| C2 | Male | 449 | 432.845 | 62.98 |
| C2 | Female | 499 | 476.086 | 68.8033 |
| C3 | Male | 482 | 419.5 | 62.1257 |
| C3 | Female | 526 | 459.781 | 69.0595 |
| C4 | Male | 518 | 437.343 | 63.6196 |
| C4 | Female | 578 | 482.939 | 72.418 |
| C5 | Male | 604 | 421.564 | 62.5727 |
| C5 | Female | 626 | 468.081 | 69.6911 |
| C6 | Male | 783 | 388.921 | 55.5006 |
| C6 | Female | 810 | 436.087 | 61.8713 |
| C7 | Male | 789 | 340.435 | 50.0886 |
| C7 | Female | 809 | 384.507 | 55.9849 |
| T1 | Male | 748 | 309.779 | 45.4572 |
| T1 | Female | 824 | 353.949 | 50.6718 |
| T2 | Male | 755 | 293.348 | 42.8753 |
| T2 | Female | 839 | 330.055 | 47.4964 |
| T3 | Male | 771 | 294.551 | 43.3798 |
| T3 | Female | 862 | 328.695 | 47.4301 |
| T4 | Male | 760 | 294.201 | 43.2533 |
| T4 | Female | 852 | 324.126 | 48.7162 |
| T5 | Male | 768 | 289.476 | 42.7878 |
| T5 | Female | 844 | 319.972 | 46.6077 |
| T6 | Male | 831 | 289.37 | 41.9524 |
| T6 | Female | 917 | 317.068 | 46.5176 |
| T7 | Male | 1012 | 286.362 | 41.7015 |
| T7 | Female | 1080 | 310.667 | 47.3789 |
| T8 | Male | 1170 | 284.359 | 41.7595 |
| T8 | Female | 1256 | 307.46 | 46.6553 |
| T9 | Male | 1242 | 289.159 | 42.5686 |
| T9 | Female | 1466 | 312.929 | 46.2404 |
| T10 | Male | 1195 | 289.346 | 43.7465 |
| T10 | Female | 1464 | 314.554 | 46.0171 |
| T11 | Male | 1234 | 280.996 | 43.2388 |
| T11 | Female | 1501 | 303.663 | 44.7676 |
| T12 | Male | 1245 | 267.611 | 41.5055 |
| T12 | Female | 1517 | 289.955 | 42.1943 |
| L1 | Male | 1220 | 268.335 | 40.4282 |
| L1 | Female | 1484 | 292.924 | 42.093 |
| L2 | Male | 1168 | 279.794 | 41.7052 |
| L2 | Female | 1443 | 308.975 | 43.6866 |
| L3 | Male | 1185 | 288.408 | 41.4111 |
| L3 | Female | 1461 | 315.867 | 43.3663 |
| L4 | Male | 1187 | 292.194 | 41.1686 |
| L4 | Female | 1436 | 318.275 | 43.3152 |
| L5 | Male | 1189 | 293.028 | 44.1721 |
| L5 | Female | 1431 | 316.823 | 45.2603 |
| S1 | Male | 1286 | 303.755 | 48.0263 |
| S1 | Female | 1493 | 329.441 | 49.9023 |
| Left rib 1 | Male | 742 | 355.99 | 52.3421 |
| Left rib 1 | Female | 847 | 383.662 | 56.3522 |
| Left rib 2 | Male | 913 | 303.623 | 54.5668 |
| Left rib 2 | Female | 994 | 363.019 | 62.2196 |
| Left rib 3 | Male | 959 | 312.895 | 62.888 |
| Left rib 3 | Female | 1018 | 384.73 | 69.3935 |
| Left rib 4 | Male | 1100 | 321.75 | 70.3381 |
| Left rib 4 | Female | 1248 | 378.865 | 88.9502 |
| Left rib 5 | Male | 1308 | 312.214 | 77.9364 |
| Left rib 5 | Female | 1541 | 365.223 | 97.825 |
| Left rib 6 | Male | 1408 | 314.456 | 77.3506 |
| Left rib 6 | Female | 1643 | 372.328 | 93.9718 |
| Left rib 7 | Male | 1409 | 332.305 | 73.289 |
| Left rib 7 | Female | 1659 | 400.935 | 87.8602 |
| Left rib 8 | Male | 1414 | 360.836 | 74.7886 |
| Left rib 8 | Female | 1659 | 437.98 | 85.7028 |
| Left rib 9 | Male | 1402 | 375.361 | 73.6161 |
| Left rib 9 | Female | 1650 | 445.673 | 81.0152 |
| Left rib 10 | Male | 1326 | 399.569 | 75.0595 |
| Left rib 10 | Female | 1638 | 475.287 | 78.5772 |
| Left rib 11 | Male | 1342 | 421.181 | 77.3437 |
| Left rib 11 | Female | 1652 | 497.37 | 86.3995 |
| Left rib 12 | Male | 1434 | 407.633 | 83.4446 |
| Left rib 12 | Female | 1617 | 483.951 | 93.4161 |
| Right rib 1 | Male | 717 | 359.747 | 53.4626 |
| Right rib 1 | Female | 843 | 389.695 | 56.0557 |
| Right rib 2 | Male | 898 | 305.53 | 55.1589 |
| Right rib 2 | Female | 986 | 367.705 | 62.2385 |
| Right rib 3 | Male | 953 | 311.863 | 63.4507 |
| Right rib 3 | Female | 1010 | 387.282 | 70.6203 |
| Right rib 4 | Male | 1102 | 319.973 | 69.833 |
| Right rib 4 | Female | 1241 | 385.118 | 87.9344 |
| Right rib 5 | Male | 1292 | 308.002 | 73.3777 |
| Right rib 5 | Female | 1549 | 361.997 | 99.151 |
| Right rib 6 | Male | 1417 | 316.532 | 75.3662 |
| Right rib 6 | Female | 1664 | 379.166 | 93.76 |
| Right rib 7 | Male | 1417 | 333.734 | 71.0987 |
| Right rib 7 | Female | 1661 | 403.251 | 86.604 |
| Right rib 8 | Male | 1397 | 357.991 | 72.6953 |
| Right rib 8 | Female | 1671 | 430.642 | 83.5929 |
| Right rib 9 | Male | 1385 | 369.592 | 71.1257 |
| Right rib 9 | Female | 1653 | 441.984 | 79.0167 |
| Right rib 10 | Male | 1317 | 395.135 | 73.2478 |
| Right rib 10 | Female | 1632 | 474.937 | 77.8219 |
| Right rib 11 | Male | 1362 | 419.42 | 76.5762 |
| Right rib 11 | Female | 1660 | 496.104 | 85.3295 |
| Right rib 12 | Male | 1454 | 401.28 | 80.8895 |
| Right rib 12 | Female | 1636 | 475.601 | 90.3928 |

Participants aged 20–34 years of the same sex. A standardized score is (measured − mean) / SD, mirroring the construction of a DXA T-score.

**Supplementary Table 6: Prevalence of a young-adult-referenced score below −2.5**

| **Site** | **Sex** | **Age group (y)** | **n** | **Score < −2.5, n** | **Score < −2.5, %** | **Lower 95% limit, %** | **Upper 95% limit, %** |
| --- | --- | --- | --- | --- | --- | --- | --- |
| C1 | Male | 20-29 | 264 | 2 | 0.8 | 0.2 | 2.7 |
| C1 | Male | 30-39 | 413 | 3 | 0.7 | 0.2 | 2.1 |
| C1 | Male | 40-49 | 973 | 6 | 0.6 | 0.3 | 1.3 |
| C1 | Male | 50-59 | 1929 | 31 | 1.6 | 1.1 | 2.3 |
| C1 | Male | 60-69 | 3036 | 51 | 1.7 | 1.3 | 2.2 |
| C1 | Male | 70-79 | 4154 | 66 | 1.6 | 1.3 | 2 |
| C1 | Male | 80-89 | 1887 | 30 | 1.6 | 1.1 | 2.3 |
| C1 | Male | 90-99 | 185 | 0 | 0 | 0 | 2 |
| C1 | Female | 20-29 | 307 | 3 | 1 | 0.3 | 2.8 |
| C1 | Female | 30-39 | 530 | 7 | 1.3 | 0.6 | 2.7 |
| C1 | Female | 40-49 | 1458 | 16 | 1.1 | 0.7 | 1.8 |
| C1 | Female | 50-59 | 2651 | 136 | 5.1 | 4.4 | 6 |
| C1 | Female | 60-69 | 2872 | 388 | 13.5 | 12.3 | 14.8 |
| C1 | Female | 70-79 | 3371 | 743 | 22 | 20.7 | 23.5 |
| C1 | Female | 80-89 | 1792 | 419 | 23.4 | 21.5 | 25.4 |
| C1 | Female | 90-99 | 276 | 85 | 30.8 | 25.6 | 36.5 |
| C2 | Male | 20-29 | 264 | 1 | 0.4 | 0.1 | 2.1 |
| C2 | Male | 30-39 | 407 | 2 | 0.5 | 0.1 | 1.8 |
| C2 | Male | 40-49 | 976 | 11 | 1.1 | 0.6 | 2 |
| C2 | Male | 50-59 | 1933 | 41 | 2.1 | 1.6 | 2.9 |
| C2 | Male | 60-69 | 3051 | 118 | 3.9 | 3.2 | 4.6 |
| C2 | Male | 70-79 | 4181 | 211 | 5 | 4.4 | 5.8 |
| C2 | Male | 80-89 | 1869 | 118 | 6.3 | 5.3 | 7.5 |
| C2 | Male | 90-99 | 170 | 7 | 4.1 | 2 | 8.3 |
| C2 | Female | 20-29 | 303 | 2 | 0.7 | 0.2 | 2.4 |
| C2 | Female | 30-39 | 516 | 3 | 0.6 | 0.2 | 1.7 |
| C2 | Female | 40-49 | 1439 | 24 | 1.7 | 1.1 | 2.5 |
| C2 | Female | 50-59 | 2674 | 176 | 6.6 | 5.7 | 7.6 |
| C2 | Female | 60-69 | 2897 | 557 | 19.2 | 17.8 | 20.7 |
| C2 | Female | 70-79 | 3378 | 1155 | 34.2 | 32.6 | 35.8 |
| C2 | Female | 80-89 | 1782 | 721 | 40.5 | 38.2 | 42.8 |
| C2 | Female | 90-99 | 269 | 131 | 48.7 | 42.8 | 54.6 |
| C3 | Male | 20-29 | 279 | 1 | 0.4 | 0.1 | 2 |
| C3 | Male | 30-39 | 437 | 3 | 0.7 | 0.2 | 2 |
| C3 | Male | 40-49 | 1008 | 14 | 1.4 | 0.8 | 2.3 |
| C3 | Male | 50-59 | 2056 | 39 | 1.9 | 1.4 | 2.6 |
| C3 | Male | 60-69 | 3214 | 69 | 2.1 | 1.7 | 2.7 |
| C3 | Male | 70-79 | 4538 | 129 | 2.8 | 2.4 | 3.4 |
| C3 | Male | 80-89 | 2113 | 62 | 2.9 | 2.3 | 3.7 |
| C3 | Male | 90-99 | 209 | 2 | 1 | 0.3 | 3.4 |
| C3 | Female | 20-29 | 316 | 1 | 0.3 | 0.1 | 1.8 |
| C3 | Female | 30-39 | 555 | 2 | 0.4 | 0.1 | 1.3 |
| C3 | Female | 40-49 | 1519 | 18 | 1.2 | 0.8 | 1.9 |
| C3 | Female | 50-59 | 2809 | 133 | 4.7 | 4 | 5.6 |
| C3 | Female | 60-69 | 3022 | 375 | 12.4 | 11.3 | 13.6 |
| C3 | Female | 70-79 | 3636 | 791 | 21.8 | 20.4 | 23.1 |
| C3 | Female | 80-89 | 2011 | 493 | 24.5 | 22.7 | 26.4 |
| C3 | Female | 90-99 | 326 | 89 | 27.3 | 22.7 | 32.4 |
| C4 | Male | 20-29 | 305 | 1 | 0.3 | 0.1 | 1.8 |
| C4 | Male | 30-39 | 462 | 1 | 0.2 | 0 | 1.2 |
| C4 | Male | 40-49 | 1092 | 12 | 1.1 | 0.6 | 1.9 |
| C4 | Male | 50-59 | 2211 | 44 | 2 | 1.5 | 2.7 |
| C4 | Male | 60-69 | 3525 | 90 | 2.6 | 2.1 | 3.1 |
| C4 | Male | 70-79 | 5143 | 115 | 2.2 | 1.9 | 2.7 |
| C4 | Male | 80-89 | 2429 | 60 | 2.5 | 1.9 | 3.2 |
| C4 | Male | 90-99 | 248 | 2 | 0.8 | 0.2 | 2.9 |
| C4 | Female | 20-29 | 347 | 2 | 0.6 | 0.2 | 2.1 |
| C4 | Female | 30-39 | 599 | 3 | 0.5 | 0.2 | 1.5 |
| C4 | Female | 40-49 | 1597 | 20 | 1.3 | 0.8 | 1.9 |
| C4 | Female | 50-59 | 2967 | 128 | 4.3 | 3.6 | 5.1 |
| C4 | Female | 60-69 | 3195 | 371 | 11.6 | 10.5 | 12.8 |
| C4 | Female | 70-79 | 3951 | 711 | 18 | 16.8 | 19.2 |
| C4 | Female | 80-89 | 2223 | 412 | 18.5 | 17 | 20.2 |
| C4 | Female | 90-99 | 378 | 92 | 24.3 | 20.3 | 28.9 |
| C5 | Male | 20-29 | 347 | 2 | 0.6 | 0.2 | 2.1 |
| C5 | Male | 30-39 | 549 | 2 | 0.4 | 0.1 | 1.3 |
| C5 | Male | 40-49 | 1278 | 12 | 0.9 | 0.5 | 1.6 |
| C5 | Male | 50-59 | 2615 | 49 | 1.9 | 1.4 | 2.5 |
| C5 | Male | 60-69 | 4169 | 80 | 1.9 | 1.5 | 2.4 |
| C5 | Male | 70-79 | 6416 | 152 | 2.4 | 2 | 2.8 |
| C5 | Male | 80-89 | 3210 | 81 | 2.5 | 2 | 3.1 |
| C5 | Male | 90-99 | 320 | 2 | 0.6 | 0.2 | 2.2 |
| C5 | Female | 20-29 | 371 | 3 | 0.8 | 0.3 | 2.4 |
| C5 | Female | 30-39 | 665 | 3 | 0.5 | 0.2 | 1.3 |
| C5 | Female | 40-49 | 1794 | 24 | 1.3 | 0.9 | 2 |
| C5 | Female | 50-59 | 3330 | 150 | 4.5 | 3.9 | 5.3 |
| C5 | Female | 60-69 | 3656 | 439 | 12 | 11 | 13.1 |
| C5 | Female | 70-79 | 4696 | 879 | 18.7 | 17.6 | 19.9 |
| C5 | Female | 80-89 | 2773 | 594 | 21.4 | 19.9 | 23 |
| C5 | Female | 90-99 | 490 | 122 | 24.9 | 21.3 | 28.9 |
| C6 | Male | 20-29 | 457 | 2 | 0.4 | 0.1 | 1.6 |
| C6 | Male | 30-39 | 718 | 4 | 0.6 | 0.2 | 1.4 |
| C6 | Male | 40-49 | 1633 | 21 | 1.3 | 0.8 | 2 |
| C6 | Male | 50-59 | 3372 | 61 | 1.8 | 1.4 | 2.3 |
| C6 | Male | 60-69 | 5334 | 102 | 1.9 | 1.6 | 2.3 |
| C6 | Male | 70-79 | 8531 | 225 | 2.6 | 2.3 | 3 |
| C6 | Male | 80-89 | 4438 | 148 | 3.3 | 2.8 | 3.9 |
| C6 | Male | 90-99 | 443 | 10 | 2.3 | 1.2 | 4.1 |
| C6 | Female | 20-29 | 471 | 2 | 0.4 | 0.1 | 1.5 |
| C6 | Female | 30-39 | 848 | 4 | 0.5 | 0.2 | 1.2 |
| C6 | Female | 40-49 | 2248 | 35 | 1.6 | 1.1 | 2.2 |
| C6 | Female | 50-59 | 4131 | 221 | 5.3 | 4.7 | 6.1 |
| C6 | Female | 60-69 | 4539 | 629 | 13.9 | 12.9 | 14.9 |
| C6 | Female | 70-79 | 6021 | 1373 | 22.8 | 21.8 | 23.9 |
| C6 | Female | 80-89 | 3609 | 995 | 27.6 | 26.1 | 29.1 |
| C6 | Female | 90-99 | 603 | 207 | 34.3 | 30.6 | 38.2 |
| C7 | Male | 20-29 | 455 | 1 | 0.2 | 0 | 1.2 |
| C7 | Male | 30-39 | 749 | 2 | 0.3 | 0.1 | 1 |
| C7 | Male | 40-49 | 1753 | 18 | 1 | 0.7 | 1.6 |
| C7 | Male | 50-59 | 3738 | 48 | 1.3 | 1 | 1.7 |
| C7 | Male | 60-69 | 5967 | 94 | 1.6 | 1.3 | 1.9 |
| C7 | Male | 70-79 | 9724 | 182 | 1.9 | 1.6 | 2.2 |
| C7 | Male | 80-89 | 5163 | 126 | 2.4 | 2.1 | 2.9 |
| C7 | Male | 90-99 | 505 | 15 | 3 | 1.8 | 4.8 |
| C7 | Female | 20-29 | 450 | 3 | 0.7 | 0.2 | 1.9 |
| C7 | Female | 30-39 | 914 | 4 | 0.4 | 0.2 | 1.1 |
| C7 | Female | 40-49 | 2474 | 31 | 1.3 | 0.9 | 1.8 |
| C7 | Female | 50-59 | 4597 | 206 | 4.5 | 3.9 | 5.1 |
| C7 | Female | 60-69 | 5022 | 585 | 11.6 | 10.8 | 12.6 |
| C7 | Female | 70-79 | 6808 | 1320 | 19.4 | 18.5 | 20.3 |
| C7 | Female | 80-89 | 4087 | 1032 | 25.3 | 23.9 | 26.6 |
| C7 | Female | 90-99 | 659 | 210 | 31.9 | 28.4 | 35.5 |
| T1 | Male | 20-29 | 427 | 0 | 0 | 0 | 0.9 |
| T1 | Male | 30-39 | 745 | 5 | 0.7 | 0.3 | 1.6 |
| T1 | Male | 40-49 | 1783 | 20 | 1.1 | 0.7 | 1.7 |
| T1 | Male | 50-59 | 3808 | 77 | 2 | 1.6 | 2.5 |
| T1 | Male | 60-69 | 6069 | 170 | 2.8 | 2.4 | 3.2 |
| T1 | Male | 70-79 | 9892 | 452 | 4.6 | 4.2 | 5 |
| T1 | Male | 80-89 | 5224 | 331 | 6.3 | 5.7 | 7 |
| T1 | Male | 90-99 | 497 | 36 | 7.2 | 5.3 | 9.9 |
| T1 | Female | 20-29 | 459 | 4 | 0.9 | 0.3 | 2.2 |
| T1 | Female | 30-39 | 917 | 4 | 0.4 | 0.2 | 1.1 |
| T1 | Female | 40-49 | 2482 | 39 | 1.6 | 1.2 | 2.1 |
| T1 | Female | 50-59 | 4680 | 299 | 6.4 | 5.7 | 7.1 |
| T1 | Female | 60-69 | 5099 | 979 | 19.2 | 18.1 | 20.3 |
| T1 | Female | 70-79 | 6907 | 2161 | 31.3 | 30.2 | 32.4 |
| T1 | Female | 80-89 | 4164 | 1682 | 40.4 | 38.9 | 41.9 |
| T1 | Female | 90-99 | 668 | 326 | 48.8 | 45 | 52.6 |
| T2 | Male | 20-29 | 427 | 2 | 0.5 | 0.1 | 1.7 |
| T2 | Male | 30-39 | 746 | 4 | 0.5 | 0.2 | 1.4 |
| T2 | Male | 40-49 | 1772 | 25 | 1.4 | 1 | 2.1 |
| T2 | Male | 50-59 | 3794 | 93 | 2.5 | 2 | 3 |
| T2 | Male | 60-69 | 6088 | 220 | 3.6 | 3.2 | 4.1 |
| T2 | Male | 70-79 | 9904 | 611 | 6.2 | 5.7 | 6.7 |
| T2 | Male | 80-89 | 5207 | 418 | 8 | 7.3 | 8.8 |
| T2 | Male | 90-99 | 498 | 51 | 10.2 | 7.9 | 13.2 |
| T2 | Female | 20-29 | 470 | 3 | 0.6 | 0.2 | 1.9 |
| T2 | Female | 30-39 | 921 | 8 | 0.9 | 0.4 | 1.7 |
| T2 | Female | 40-49 | 2501 | 48 | 1.9 | 1.5 | 2.5 |
| T2 | Female | 50-59 | 4664 | 323 | 6.9 | 6.2 | 7.7 |
| T2 | Female | 60-69 | 5099 | 960 | 18.8 | 17.8 | 19.9 |
| T2 | Female | 70-79 | 6881 | 2157 | 31.3 | 30.3 | 32.5 |
| T2 | Female | 80-89 | 4147 | 1657 | 40 | 38.5 | 41.5 |
| T2 | Female | 90-99 | 667 | 332 | 49.8 | 46 | 53.6 |
| T3 | Male | 20-29 | 444 | 2 | 0.5 | 0.1 | 1.6 |
| T3 | Male | 30-39 | 752 | 2 | 0.3 | 0.1 | 1 |
| T3 | Male | 40-49 | 1788 | 25 | 1.4 | 0.9 | 2.1 |
| T3 | Male | 50-59 | 3796 | 111 | 2.9 | 2.4 | 3.5 |
| T3 | Male | 60-69 | 6095 | 302 | 5 | 4.4 | 5.5 |
| T3 | Male | 70-79 | 9880 | 756 | 7.7 | 7.1 | 8.2 |
| T3 | Male | 80-89 | 5204 | 496 | 9.5 | 8.8 | 10.4 |
| T3 | Male | 90-99 | 504 | 60 | 11.9 | 9.4 | 15 |
| T3 | Female | 20-29 | 484 | 5 | 1 | 0.4 | 2.4 |
| T3 | Female | 30-39 | 931 | 10 | 1.1 | 0.6 | 2 |
| T3 | Female | 40-49 | 2519 | 67 | 2.7 | 2.1 | 3.4 |
| T3 | Female | 50-59 | 4686 | 410 | 8.7 | 8 | 9.6 |
| T3 | Female | 60-69 | 5079 | 1122 | 22.1 | 21 | 23.3 |
| T3 | Female | 70-79 | 6857 | 2527 | 36.9 | 35.7 | 38 |
| T3 | Female | 80-89 | 4128 | 1940 | 47 | 45.5 | 48.5 |
| T3 | Female | 90-99 | 674 | 387 | 57.4 | 53.7 | 61.1 |
| T4 | Male | 20-29 | 437 | 3 | 0.7 | 0.2 | 2 |
| T4 | Male | 30-39 | 747 | 7 | 0.9 | 0.5 | 1.9 |
| T4 | Male | 40-49 | 1793 | 27 | 1.5 | 1 | 2.2 |
| T4 | Male | 50-59 | 3818 | 124 | 3.2 | 2.7 | 3.9 |
| T4 | Male | 60-69 | 6107 | 324 | 5.3 | 4.8 | 5.9 |
| T4 | Male | 70-79 | 9869 | 812 | 8.2 | 7.7 | 8.8 |
| T4 | Male | 80-89 | 5193 | 567 | 10.9 | 10.1 | 11.8 |
| T4 | Male | 90-99 | 509 | 73 | 14.3 | 11.6 | 17.7 |
| T4 | Female | 20-29 | 482 | 3 | 0.6 | 0.2 | 1.8 |
| T4 | Female | 30-39 | 914 | 6 | 0.7 | 0.3 | 1.4 |
| T4 | Female | 40-49 | 2503 | 46 | 1.8 | 1.4 | 2.4 |
| T4 | Female | 50-59 | 4680 | 365 | 7.8 | 7.1 | 8.6 |
| T4 | Female | 60-69 | 5073 | 1050 | 20.7 | 19.6 | 21.8 |
| T4 | Female | 70-79 | 6859 | 2329 | 34 | 32.8 | 35.1 |
| T4 | Female | 80-89 | 4173 | 1854 | 44.4 | 42.9 | 45.9 |
| T4 | Female | 90-99 | 674 | 361 | 53.6 | 49.8 | 57.3 |
| T5 | Male | 20-29 | 444 | 0 | 0 | 0 | 0.9 |
| T5 | Male | 30-39 | 743 | 8 | 1.1 | 0.5 | 2.1 |
| T5 | Male | 40-49 | 1797 | 21 | 1.2 | 0.8 | 1.8 |
| T5 | Male | 50-59 | 3826 | 109 | 2.8 | 2.4 | 3.4 |
| T5 | Male | 60-69 | 6110 | 288 | 4.7 | 4.2 | 5.3 |
| T5 | Male | 70-79 | 9897 | 739 | 7.5 | 7 | 8 |
| T5 | Male | 80-89 | 5204 | 507 | 9.7 | 9 | 10.6 |
| T5 | Male | 90-99 | 512 | 67 | 13.1 | 10.4 | 16.3 |
| T5 | Female | 20-29 | 469 | 2 | 0.4 | 0.1 | 1.5 |
| T5 | Female | 30-39 | 931 | 7 | 0.8 | 0.4 | 1.5 |
| T5 | Female | 40-49 | 2511 | 60 | 2.4 | 1.9 | 3.1 |
| T5 | Female | 50-59 | 4684 | 439 | 9.4 | 8.6 | 10.2 |
| T5 | Female | 60-69 | 5086 | 1197 | 23.5 | 22.4 | 24.7 |
| T5 | Female | 70-79 | 6874 | 2529 | 36.8 | 35.7 | 37.9 |
| T5 | Female | 80-89 | 4191 | 2037 | 48.6 | 47.1 | 50.1 |
| T5 | Female | 90-99 | 675 | 390 | 57.8 | 54 | 61.4 |
| T6 | Male | 20-29 | 480 | 2 | 0.4 | 0.1 | 1.5 |
| T6 | Male | 30-39 | 806 | 9 | 1.1 | 0.6 | 2.1 |
| T6 | Male | 40-49 | 1862 | 22 | 1.2 | 0.8 | 1.8 |
| T6 | Male | 50-59 | 3942 | 129 | 3.3 | 2.8 | 3.9 |
| T6 | Male | 60-69 | 6204 | 341 | 5.5 | 5 | 6.1 |
| T6 | Male | 70-79 | 10030 | 866 | 8.6 | 8.1 | 9.2 |
| T6 | Male | 80-89 | 5276 | 576 | 10.9 | 10.1 | 11.8 |
| T6 | Male | 90-99 | 511 | 76 | 14.9 | 12.1 | 18.2 |
| T6 | Female | 20-29 | 516 | 3 | 0.6 | 0.2 | 1.7 |
| T6 | Female | 30-39 | 985 | 8 | 0.8 | 0.4 | 1.6 |
| T6 | Female | 40-49 | 2614 | 55 | 2.1 | 1.6 | 2.7 |
| T6 | Female | 50-59 | 4772 | 402 | 8.4 | 7.7 | 9.2 |
| T6 | Female | 60-69 | 5167 | 1146 | 22.2 | 21.1 | 23.3 |
| T6 | Female | 70-79 | 6930 | 2434 | 35.1 | 34 | 36.3 |
| T6 | Female | 80-89 | 4249 | 1933 | 45.5 | 44 | 47 |
| T6 | Female | 90-99 | 678 | 374 | 55.2 | 51.4 | 58.9 |
| T7 | Male | 20-29 | 597 | 4 | 0.7 | 0.3 | 1.7 |
| T7 | Male | 30-39 | 938 | 10 | 1.1 | 0.6 | 2 |
| T7 | Male | 40-49 | 2074 | 38 | 1.8 | 1.3 | 2.5 |
| T7 | Male | 50-59 | 4297 | 148 | 3.4 | 2.9 | 4 |
| T7 | Male | 60-69 | 6600 | 346 | 5.2 | 4.7 | 5.8 |
| T7 | Male | 70-79 | 10524 | 910 | 8.6 | 8.1 | 9.2 |
| T7 | Male | 80-89 | 5551 | 634 | 11.4 | 10.6 | 12.3 |
| T7 | Male | 90-99 | 538 | 74 | 13.8 | 11.1 | 16.9 |
| T7 | Female | 20-29 | 613 | 3 | 0.5 | 0.2 | 1.4 |
| T7 | Female | 30-39 | 1111 | 8 | 0.7 | 0.4 | 1.4 |
| T7 | Female | 40-49 | 2813 | 46 | 1.6 | 1.2 | 2.2 |
| T7 | Female | 50-59 | 5093 | 359 | 7 | 6.4 | 7.8 |
| T7 | Female | 60-69 | 5489 | 1020 | 18.6 | 17.6 | 19.6 |
| T7 | Female | 70-79 | 7363 | 2201 | 29.9 | 28.9 | 30.9 |
| T7 | Female | 80-89 | 4469 | 1760 | 39.4 | 38 | 40.8 |
| T7 | Female | 90-99 | 714 | 349 | 48.9 | 45.2 | 52.5 |
| T8 | Male | 20-29 | 690 | 4 | 0.6 | 0.2 | 1.5 |
| T8 | Male | 30-39 | 1084 | 14 | 1.3 | 0.8 | 2.2 |
| T8 | Male | 40-49 | 2329 | 43 | 1.8 | 1.4 | 2.5 |
| T8 | Male | 50-59 | 4778 | 181 | 3.8 | 3.3 | 4.4 |
| T8 | Male | 60-69 | 7233 | 407 | 5.6 | 5.1 | 6.2 |
| T8 | Male | 70-79 | 11447 | 981 | 8.6 | 8.1 | 9.1 |
| T8 | Male | 80-89 | 5953 | 677 | 11.4 | 10.6 | 12.2 |
| T8 | Male | 90-99 | 583 | 83 | 14.2 | 11.6 | 17.3 |
| T8 | Female | 20-29 | 717 | 4 | 0.6 | 0.2 | 1.4 |
| T8 | Female | 30-39 | 1282 | 6 | 0.5 | 0.2 | 1 |
| T8 | Female | 40-49 | 3092 | 60 | 1.9 | 1.5 | 2.5 |
| T8 | Female | 50-59 | 5538 | 486 | 8.8 | 8.1 | 9.5 |
| T8 | Female | 60-69 | 5996 | 1230 | 20.5 | 19.5 | 21.6 |
| T8 | Female | 70-79 | 8043 | 2582 | 32.1 | 31.1 | 33.1 |
| T8 | Female | 80-89 | 4813 | 1961 | 40.7 | 39.4 | 42.1 |
| T8 | Female | 90-99 | 754 | 374 | 49.6 | 46 | 53.2 |
| T9 | Male | 20-29 | 735 | 7 | 1 | 0.5 | 2 |
| T9 | Male | 30-39 | 1174 | 15 | 1.3 | 0.8 | 2.1 |
| T9 | Male | 40-49 | 2544 | 55 | 2.2 | 1.7 | 2.8 |
| T9 | Male | 50-59 | 5168 | 217 | 4.2 | 3.7 | 4.8 |
| T9 | Male | 60-69 | 7887 | 469 | 5.9 | 5.4 | 6.5 |
| T9 | Male | 70-79 | 12279 | 1067 | 8.7 | 8.2 | 9.2 |
| T9 | Male | 80-89 | 6390 | 730 | 11.4 | 10.7 | 12.2 |
| T9 | Male | 90-99 | 619 | 104 | 16.8 | 14.1 | 19.9 |
| T9 | Female | 20-29 | 839 | 9 | 1.1 | 0.6 | 2 |
| T9 | Female | 30-39 | 1465 | 10 | 0.7 | 0.4 | 1.3 |
| T9 | Female | 40-49 | 3436 | 81 | 2.4 | 1.9 | 2.9 |
| T9 | Female | 50-59 | 6067 | 618 | 10.2 | 9.5 | 11 |
| T9 | Female | 60-69 | 6552 | 1577 | 24.1 | 23 | 25.1 |
| T9 | Female | 70-79 | 8749 | 3120 | 35.7 | 34.7 | 36.7 |
| T9 | Female | 80-89 | 5125 | 2221 | 43.3 | 42 | 44.7 |
| T9 | Female | 90-99 | 776 | 405 | 52.2 | 48.7 | 55.7 |
| T10 | Male | 20-29 | 690 | 2 | 0.3 | 0.1 | 1.1 |
| T10 | Male | 30-39 | 1181 | 14 | 1.2 | 0.7 | 2 |
| T10 | Male | 40-49 | 2597 | 55 | 2.1 | 1.6 | 2.7 |
| T10 | Male | 50-59 | 5424 | 217 | 4 | 3.5 | 4.6 |
| T10 | Male | 60-69 | 8270 | 484 | 5.9 | 5.4 | 6.4 |
| T10 | Male | 70-79 | 12862 | 1169 | 9.1 | 8.6 | 9.6 |
| T10 | Male | 80-89 | 6656 | 785 | 11.8 | 11 | 12.6 |
| T10 | Male | 90-99 | 635 | 106 | 16.7 | 14 | 19.8 |
| T10 | Female | 20-29 | 827 | 6 | 0.7 | 0.3 | 1.6 |
| T10 | Female | 30-39 | 1543 | 11 | 0.7 | 0.4 | 1.3 |
| T10 | Female | 40-49 | 3679 | 95 | 2.6 | 2.1 | 3.1 |
| T10 | Female | 50-59 | 6506 | 759 | 11.7 | 10.9 | 12.5 |
| T10 | Female | 60-69 | 6946 | 1872 | 27 | 25.9 | 28 |
| T10 | Female | 70-79 | 9200 | 3747 | 40.7 | 39.7 | 41.7 |
| T10 | Female | 80-89 | 5313 | 2528 | 47.6 | 46.2 | 48.9 |
| T10 | Female | 90-99 | 790 | 439 | 55.6 | 52.1 | 59 |
| T11 | Male | 20-29 | 712 | 3 | 0.4 | 0.1 | 1.2 |
| T11 | Male | 30-39 | 1213 | 11 | 0.9 | 0.5 | 1.6 |
| T11 | Male | 40-49 | 2664 | 62 | 2.3 | 1.8 | 3 |
| T11 | Male | 50-59 | 5502 | 263 | 4.8 | 4.2 | 5.4 |
| T11 | Male | 60-69 | 8346 | 602 | 7.2 | 6.7 | 7.8 |
| T11 | Male | 70-79 | 13015 | 1420 | 10.9 | 10.4 | 11.5 |
| T11 | Male | 80-89 | 6654 | 897 | 13.5 | 12.7 | 14.3 |
| T11 | Male | 90-99 | 638 | 108 | 16.9 | 14.2 | 20 |
| T11 | Female | 20-29 | 844 | 5 | 0.6 | 0.3 | 1.4 |
| T11 | Female | 30-39 | 1591 | 14 | 0.9 | 0.5 | 1.5 |
| T11 | Female | 40-49 | 3803 | 97 | 2.6 | 2.1 | 3.1 |
| T11 | Female | 50-59 | 6686 | 824 | 12.3 | 11.6 | 13.1 |
| T11 | Female | 60-69 | 7058 | 2049 | 29 | 28 | 30.1 |
| T11 | Female | 70-79 | 9287 | 3982 | 42.9 | 41.9 | 43.9 |
| T11 | Female | 80-89 | 5298 | 2591 | 48.9 | 47.6 | 50.3 |
| T11 | Female | 90-99 | 780 | 441 | 56.5 | 53 | 60 |
| T12 | Male | 20-29 | 716 | 4 | 0.6 | 0.2 | 1.4 |
| T12 | Male | 30-39 | 1218 | 12 | 1 | 0.6 | 1.7 |
| T12 | Male | 40-49 | 2656 | 67 | 2.5 | 2 | 3.2 |
| T12 | Male | 50-59 | 5461 | 324 | 5.9 | 5.3 | 6.6 |
| T12 | Male | 60-69 | 8335 | 762 | 9.1 | 8.5 | 9.8 |
| T12 | Male | 70-79 | 13020 | 1735 | 13.3 | 12.8 | 13.9 |
| T12 | Male | 80-89 | 6674 | 1065 | 16 | 15.1 | 16.9 |
| T12 | Male | 90-99 | 625 | 123 | 19.7 | 16.8 | 23 |
| T12 | Female | 20-29 | 855 | 2 | 0.2 | 0.1 | 0.8 |
| T12 | Female | 30-39 | 1596 | 19 | 1.2 | 0.8 | 1.9 |
| T12 | Female | 40-49 | 3805 | 123 | 3.2 | 2.7 | 3.8 |
| T12 | Female | 50-59 | 6682 | 1033 | 15.5 | 14.6 | 16.3 |
| T12 | Female | 60-69 | 7037 | 2473 | 35.1 | 34 | 36.3 |
| T12 | Female | 70-79 | 9307 | 4521 | 48.6 | 47.6 | 49.6 |
| T12 | Female | 80-89 | 5240 | 2794 | 53.3 | 52 | 54.7 |
| T12 | Female | 90-99 | 762 | 438 | 57.5 | 53.9 | 60.9 |
| L1 | Male | 20-29 | 703 | 4 | 0.6 | 0.2 | 1.5 |
| L1 | Male | 30-39 | 1201 | 16 | 1.3 | 0.8 | 2.2 |
| L1 | Male | 40-49 | 2647 | 67 | 2.5 | 2 | 3.2 |
| L1 | Male | 50-59 | 5463 | 330 | 6 | 5.4 | 6.7 |
| L1 | Male | 60-69 | 8346 | 686 | 8.2 | 7.6 | 8.8 |
| L1 | Male | 70-79 | 13048 | 1676 | 12.8 | 12.3 | 13.4 |
| L1 | Male | 80-89 | 6678 | 1008 | 15.1 | 14.3 | 16 |
| L1 | Male | 90-99 | 631 | 117 | 18.5 | 15.7 | 21.8 |
| L1 | Female | 20-29 | 837 | 3 | 0.4 | 0.1 | 1 |
| L1 | Female | 30-39 | 1578 | 20 | 1.3 | 0.8 | 1.9 |
| L1 | Female | 40-49 | 3781 | 116 | 3.1 | 2.6 | 3.7 |
| L1 | Female | 50-59 | 6675 | 1007 | 15.1 | 14.2 | 16 |
| L1 | Female | 60-69 | 7008 | 2403 | 34.3 | 33.2 | 35.4 |
| L1 | Female | 70-79 | 9309 | 4351 | 46.7 | 45.7 | 47.8 |
| L1 | Female | 80-89 | 5326 | 2705 | 50.8 | 49.4 | 52.1 |
| L1 | Female | 90-99 | 781 | 436 | 55.8 | 52.3 | 59.3 |
| L2 | Male | 20-29 | 669 | 3 | 0.4 | 0.2 | 1.3 |
| L2 | Male | 30-39 | 1167 | 14 | 1.2 | 0.7 | 2 |
| L2 | Male | 40-49 | 2612 | 60 | 2.3 | 1.8 | 2.9 |
| L2 | Male | 50-59 | 5473 | 266 | 4.9 | 4.3 | 5.5 |
| L2 | Male | 60-69 | 8375 | 572 | 6.8 | 6.3 | 7.4 |
| L2 | Male | 70-79 | 13067 | 1295 | 9.9 | 9.4 | 10.4 |
| L2 | Male | 80-89 | 6734 | 870 | 12.9 | 12.1 | 13.7 |
| L2 | Male | 90-99 | 638 | 90 | 14.1 | 11.6 | 17 |
| L2 | Female | 20-29 | 803 | 3 | 0.4 | 0.1 | 1.1 |
| L2 | Female | 30-39 | 1555 | 20 | 1.3 | 0.8 | 2 |
| L2 | Female | 40-49 | 3755 | 136 | 3.6 | 3.1 | 4.3 |
| L2 | Female | 50-59 | 6656 | 1079 | 16.2 | 15.3 | 17.1 |
| L2 | Female | 60-69 | 7015 | 2407 | 34.3 | 33.2 | 35.4 |
| L2 | Female | 70-79 | 9377 | 4332 | 46.2 | 45.2 | 47.2 |
| L2 | Female | 80-89 | 5386 | 2731 | 50.7 | 49.4 | 52 |
| L2 | Female | 90-99 | 795 | 460 | 57.9 | 54.4 | 61.2 |
| L3 | Male | 20-29 | 676 | 3 | 0.4 | 0.2 | 1.3 |
| L3 | Male | 30-39 | 1181 | 17 | 1.4 | 0.9 | 2.3 |
| L3 | Male | 40-49 | 2627 | 65 | 2.5 | 1.9 | 3.1 |
| L3 | Male | 50-59 | 5469 | 328 | 6 | 5.4 | 6.7 |
| L3 | Male | 60-69 | 8379 | 695 | 8.3 | 7.7 | 8.9 |
| L3 | Male | 70-79 | 13071 | 1525 | 11.7 | 11.1 | 12.2 |
| L3 | Male | 80-89 | 6692 | 988 | 14.8 | 13.9 | 15.6 |
| L3 | Male | 90-99 | 629 | 118 | 18.8 | 15.9 | 22 |
| L3 | Female | 20-29 | 813 | 4 | 0.5 | 0.2 | 1.3 |
| L3 | Female | 30-39 | 1572 | 22 | 1.4 | 0.9 | 2.1 |
| L3 | Female | 40-49 | 3754 | 150 | 4 | 3.4 | 4.7 |
| L3 | Female | 50-59 | 6649 | 1209 | 18.2 | 17.3 | 19.1 |
| L3 | Female | 60-69 | 7003 | 2732 | 39 | 37.9 | 40.2 |
| L3 | Female | 70-79 | 9380 | 4844 | 51.6 | 50.6 | 52.7 |
| L3 | Female | 80-89 | 5364 | 3087 | 57.6 | 56.2 | 58.9 |
| L3 | Female | 90-99 | 788 | 479 | 60.8 | 57.3 | 64.1 |
| L4 | Male | 20-29 | 682 | 4 | 0.6 | 0.2 | 1.5 |
| L4 | Male | 30-39 | 1173 | 14 | 1.2 | 0.7 | 2 |
| L4 | Male | 40-49 | 2620 | 64 | 2.4 | 1.9 | 3.1 |
| L4 | Male | 50-59 | 5444 | 369 | 6.8 | 6.1 | 7.5 |
| L4 | Male | 60-69 | 8398 | 703 | 8.4 | 7.8 | 9 |
| L4 | Male | 70-79 | 13084 | 1418 | 10.8 | 10.3 | 11.4 |
| L4 | Male | 80-89 | 6699 | 888 | 13.3 | 12.5 | 14.1 |
| L4 | Male | 90-99 | 629 | 118 | 18.8 | 15.9 | 22 |
| L4 | Female | 20-29 | 795 | 5 | 0.6 | 0.3 | 1.5 |
| L4 | Female | 30-39 | 1562 | 17 | 1.1 | 0.7 | 1.7 |
| L4 | Female | 40-49 | 3745 | 137 | 3.7 | 3.1 | 4.3 |
| L4 | Female | 50-59 | 6664 | 1069 | 16 | 15.2 | 16.9 |
| L4 | Female | 60-69 | 7030 | 2531 | 36 | 34.9 | 37.1 |
| L4 | Female | 70-79 | 9373 | 4495 | 48 | 46.9 | 49 |
| L4 | Female | 80-89 | 5363 | 2939 | 54.8 | 53.5 | 56.1 |
| L4 | Female | 90-99 | 787 | 461 | 58.6 | 55.1 | 62 |
| L5 | Male | 20-29 | 685 | 3 | 0.4 | 0.1 | 1.3 |
| L5 | Male | 30-39 | 1173 | 14 | 1.2 | 0.7 | 2 |
| L5 | Male | 40-49 | 2634 | 65 | 2.5 | 1.9 | 3.1 |
| L5 | Male | 50-59 | 5463 | 287 | 5.3 | 4.7 | 5.9 |
| L5 | Male | 60-69 | 8395 | 525 | 6.3 | 5.8 | 6.8 |
| L5 | Male | 70-79 | 13096 | 1109 | 8.5 | 8 | 9 |
| L5 | Male | 80-89 | 6684 | 729 | 10.9 | 10.2 | 11.7 |
| L5 | Male | 90-99 | 622 | 106 | 17 | 14.3 | 20.2 |
| L5 | Female | 20-29 | 804 | 4 | 0.5 | 0.2 | 1.3 |
| L5 | Female | 30-39 | 1548 | 14 | 0.9 | 0.5 | 1.5 |
| L5 | Female | 40-49 | 3751 | 128 | 3.4 | 2.9 | 4 |
| L5 | Female | 50-59 | 6640 | 925 | 13.9 | 13.1 | 14.8 |
| L5 | Female | 60-69 | 7039 | 2177 | 30.9 | 29.9 | 32 |
| L5 | Female | 70-79 | 9373 | 3912 | 41.7 | 40.7 | 42.7 |
| L5 | Female | 80-89 | 5384 | 2573 | 47.8 | 46.5 | 49.1 |
| L5 | Female | 90-99 | 786 | 437 | 55.6 | 52.1 | 59 |
| S1 | Male | 20-29 | 751 | 7 | 0.9 | 0.5 | 1.9 |
| S1 | Male | 30-39 | 1222 | 9 | 0.7 | 0.4 | 1.4 |
| S1 | Male | 40-49 | 2645 | 53 | 2 | 1.5 | 2.6 |
| S1 | Male | 50-59 | 5476 | 250 | 4.6 | 4 | 5.2 |
| S1 | Male | 60-69 | 8370 | 682 | 8.1 | 7.6 | 8.8 |
| S1 | Male | 70-79 | 13010 | 1883 | 14.5 | 13.9 | 15.1 |
| S1 | Male | 80-89 | 6619 | 1317 | 19.9 | 19 | 20.9 |
| S1 | Male | 90-99 | 621 | 171 | 27.5 | 24.2 | 31.2 |
| S1 | Female | 20-29 | 835 | 5 | 0.6 | 0.3 | 1.4 |
| S1 | Female | 30-39 | 1576 | 22 | 1.4 | 0.9 | 2.1 |
| S1 | Female | 40-49 | 3756 | 94 | 2.5 | 2 | 3.1 |
| S1 | Female | 50-59 | 6657 | 639 | 9.6 | 8.9 | 10.3 |
| S1 | Female | 60-69 | 7049 | 2051 | 29.1 | 28 | 30.2 |
| S1 | Female | 70-79 | 9340 | 4478 | 47.9 | 46.9 | 49 |
| S1 | Female | 80-89 | 5349 | 3216 | 60.1 | 58.8 | 61.4 |
| S1 | Female | 90-99 | 783 | 555 | 70.9 | 67.6 | 74 |
| Left rib 1 | Male | 20-29 | 392 | 0 | 0 | 0 | 1 |
| Left rib 1 | Male | 30-39 | 803 | 7 | 0.9 | 0.4 | 1.8 |
| Left rib 1 | Male | 40-49 | 1879 | 20 | 1.1 | 0.7 | 1.6 |
| Left rib 1 | Male | 50-59 | 3895 | 150 | 3.9 | 3.3 | 4.5 |
| Left rib 1 | Male | 60-69 | 6110 | 598 | 9.8 | 9.1 | 10.6 |
| Left rib 1 | Male | 70-79 | 9832 | 1763 | 17.9 | 17.2 | 18.7 |
| Left rib 1 | Male | 80-89 | 5113 | 1196 | 23.4 | 22.3 | 24.6 |
| Left rib 1 | Male | 90-99 | 481 | 152 | 31.6 | 27.6 | 35.9 |
| Left rib 1 | Female | 20-29 | 472 | 1 | 0.2 | 0 | 1.2 |
| Left rib 1 | Female | 30-39 | 947 | 4 | 0.4 | 0.2 | 1.1 |
| Left rib 1 | Female | 40-49 | 2583 | 38 | 1.5 | 1.1 | 2 |
| Left rib 1 | Female | 50-59 | 4744 | 347 | 7.3 | 6.6 | 8.1 |
| Left rib 1 | Female | 60-69 | 5105 | 1197 | 23.4 | 22.3 | 24.6 |
| Left rib 1 | Female | 70-79 | 6797 | 2903 | 42.7 | 41.5 | 43.9 |
| Left rib 1 | Female | 80-89 | 4109 | 2331 | 56.7 | 55.2 | 58.2 |
| Left rib 1 | Female | 90-99 | 648 | 425 | 65.6 | 61.8 | 69.1 |
| Left rib 2 | Male | 20-29 | 539 | 0 | 0 | 0 | 0.7 |
| Left rib 2 | Male | 30-39 | 834 | 9 | 1.1 | 0.6 | 2 |
| Left rib 2 | Male | 40-49 | 1842 | 48 | 2.6 | 2 | 3.4 |
| Left rib 2 | Male | 50-59 | 3834 | 231 | 6 | 5.3 | 6.8 |
| Left rib 2 | Male | 60-69 | 6047 | 565 | 9.3 | 8.6 | 10.1 |
| Left rib 2 | Male | 70-79 | 9767 | 1216 | 12.5 | 11.8 | 13.1 |
| Left rib 2 | Male | 80-89 | 5132 | 690 | 13.4 | 12.5 | 14.4 |
| Left rib 2 | Male | 90-99 | 474 | 75 | 15.8 | 12.8 | 19.4 |
| Left rib 2 | Female | 20-29 | 572 | 2 | 0.3 | 0.1 | 1.3 |
| Left rib 2 | Female | 30-39 | 1030 | 16 | 1.6 | 1 | 2.5 |
| Left rib 2 | Female | 40-49 | 2623 | 101 | 3.9 | 3.2 | 4.7 |
| Left rib 2 | Female | 50-59 | 4760 | 753 | 15.8 | 14.8 | 16.9 |
| Left rib 2 | Female | 60-69 | 5057 | 1772 | 35 | 33.7 | 36.4 |
| Left rib 2 | Female | 70-79 | 6687 | 3399 | 50.8 | 49.6 | 52 |
| Left rib 2 | Female | 80-89 | 4022 | 2477 | 61.6 | 60.1 | 63.1 |
| Left rib 2 | Female | 90-99 | 630 | 425 | 67.5 | 63.7 | 71 |
| Left rib 3 | Male | 20-29 | 570 | 0 | 0 | 0 | 0.7 |
| Left rib 3 | Male | 30-39 | 872 | 8 | 0.9 | 0.5 | 1.8 |
| Left rib 3 | Male | 40-49 | 1898 | 59 | 3.1 | 2.4 | 4 |
| Left rib 3 | Male | 50-59 | 3896 | 288 | 7.4 | 6.6 | 8.3 |
| Left rib 3 | Male | 60-69 | 6161 | 695 | 11.3 | 10.5 | 12.1 |
| Left rib 3 | Male | 70-79 | 9926 | 1501 | 15.1 | 14.4 | 15.8 |
| Left rib 3 | Male | 80-89 | 5205 | 853 | 16.4 | 15.4 | 17.4 |
| Left rib 3 | Male | 90-99 | 496 | 96 | 19.4 | 16.1 | 23.1 |
| Left rib 3 | Female | 20-29 | 581 | 2 | 0.3 | 0.1 | 1.2 |
| Left rib 3 | Female | 30-39 | 1048 | 18 | 1.7 | 1.1 | 2.7 |
| Left rib 3 | Female | 40-49 | 2677 | 168 | 6.3 | 5.4 | 7.3 |
| Left rib 3 | Female | 50-59 | 4818 | 1152 | 23.9 | 22.7 | 25.1 |
| Left rib 3 | Female | 60-69 | 5202 | 2402 | 46.2 | 44.8 | 47.5 |
| Left rib 3 | Female | 70-79 | 6934 | 4189 | 60.4 | 59.3 | 61.6 |
| Left rib 3 | Female | 80-89 | 4218 | 2907 | 68.9 | 67.5 | 70.3 |
| Left rib 3 | Female | 90-99 | 677 | 484 | 71.5 | 68 | 74.8 |
| Left rib 4 | Male | 20-29 | 663 | 3 | 0.5 | 0.2 | 1.3 |
| Left rib 4 | Male | 30-39 | 975 | 11 | 1.1 | 0.6 | 2 |
| Left rib 4 | Male | 40-49 | 2119 | 55 | 2.6 | 2 | 3.4 |
| Left rib 4 | Male | 50-59 | 4324 | 205 | 4.7 | 4.1 | 5.4 |
| Left rib 4 | Male | 60-69 | 6705 | 485 | 7.2 | 6.6 | 7.9 |
| Left rib 4 | Male | 70-79 | 10810 | 979 | 9.1 | 8.5 | 9.6 |
| Left rib 4 | Male | 80-89 | 5707 | 591 | 10.4 | 9.6 | 11.2 |
| Left rib 4 | Male | 90-99 | 564 | 64 | 11.3 | 9 | 14.2 |
| Left rib 4 | Female | 20-29 | 730 | 3 | 0.4 | 0.1 | 1.2 |
| Left rib 4 | Female | 30-39 | 1237 | 7 | 0.6 | 0.3 | 1.2 |
| Left rib 4 | Female | 40-49 | 2974 | 52 | 1.7 | 1.3 | 2.3 |
| Left rib 4 | Female | 50-59 | 5350 | 285 | 5.3 | 4.8 | 6 |
| Left rib 4 | Female | 60-69 | 5752 | 617 | 10.7 | 10 | 11.6 |
| Left rib 4 | Female | 70-79 | 7806 | 1186 | 15.2 | 14.4 | 16 |
| Left rib 4 | Female | 80-89 | 4709 | 809 | 17.2 | 16.1 | 18.3 |
| Left rib 4 | Female | 90-99 | 759 | 175 | 23.1 | 20.2 | 26.2 |
| Left rib 5 | Male | 20-29 | 791 | 2 | 0.3 | 0.1 | 0.9 |
| Left rib 5 | Male | 30-39 | 1166 | 11 | 0.9 | 0.5 | 1.7 |
| Left rib 5 | Male | 40-49 | 2492 | 22 | 0.9 | 0.6 | 1.3 |
| Left rib 5 | Male | 50-59 | 5035 | 106 | 2.1 | 1.7 | 2.5 |
| Left rib 5 | Male | 60-69 | 7667 | 212 | 2.8 | 2.4 | 3.2 |
| Left rib 5 | Male | 70-79 | 12189 | 392 | 3.2 | 2.9 | 3.5 |
| Left rib 5 | Male | 80-89 | 6387 | 220 | 3.4 | 3 | 3.9 |
| Left rib 5 | Male | 90-99 | 631 | 28 | 4.4 | 3.1 | 6.3 |
| Left rib 5 | Female | 20-29 | 890 | 2 | 0.2 | 0.1 | 0.8 |
| Left rib 5 | Female | 30-39 | 1526 | 8 | 0.5 | 0.3 | 1 |
| Left rib 5 | Female | 40-49 | 3507 | 29 | 0.8 | 0.6 | 1.2 |
| Left rib 5 | Female | 50-59 | 6149 | 151 | 2.5 | 2.1 | 2.9 |
| Left rib 5 | Female | 60-69 | 6582 | 264 | 4 | 3.6 | 4.5 |
| Left rib 5 | Female | 70-79 | 8853 | 411 | 4.6 | 4.2 | 5.1 |
| Left rib 5 | Female | 80-89 | 5255 | 264 | 5 | 4.5 | 5.6 |
| Left rib 5 | Female | 90-99 | 802 | 40 | 5 | 3.7 | 6.7 |
| Left rib 6 | Male | 20-29 | 846 | 3 | 0.4 | 0.1 | 1 |
| Left rib 6 | Male | 30-39 | 1270 | 7 | 0.6 | 0.3 | 1.1 |
| Left rib 6 | Male | 40-49 | 2666 | 28 | 1.1 | 0.7 | 1.5 |
| Left rib 6 | Male | 50-59 | 5369 | 92 | 1.7 | 1.4 | 2.1 |
| Left rib 6 | Male | 60-69 | 8169 | 240 | 2.9 | 2.6 | 3.3 |
| Left rib 6 | Male | 70-79 | 12811 | 399 | 3.1 | 2.8 | 3.4 |
| Left rib 6 | Male | 80-89 | 6693 | 236 | 3.5 | 3.1 | 4 |
| Left rib 6 | Male | 90-99 | 647 | 23 | 3.6 | 2.4 | 5.3 |
| Left rib 6 | Female | 20-29 | 957 | 1 | 0.1 | 0 | 0.6 |
| Left rib 6 | Female | 30-39 | 1634 | 5 | 0.3 | 0.1 | 0.7 |
| Left rib 6 | Female | 40-49 | 3752 | 40 | 1.1 | 0.8 | 1.4 |
| Left rib 6 | Female | 50-59 | 6494 | 235 | 3.6 | 3.2 | 4.1 |
| Left rib 6 | Female | 60-69 | 6912 | 418 | 6 | 5.5 | 6.6 |
| Left rib 6 | Female | 70-79 | 9261 | 735 | 7.9 | 7.4 | 8.5 |
| Left rib 6 | Female | 80-89 | 5413 | 407 | 7.5 | 6.8 | 8.3 |
| Left rib 6 | Female | 90-99 | 818 | 73 | 8.9 | 7.2 | 11.1 |
| Left rib 7 | Male | 20-29 | 850 | 2 | 0.2 | 0.1 | 0.9 |
| Left rib 7 | Male | 30-39 | 1275 | 7 | 0.5 | 0.3 | 1.1 |
| Left rib 7 | Male | 40-49 | 2679 | 49 | 1.8 | 1.4 | 2.4 |
| Left rib 7 | Male | 50-59 | 5416 | 170 | 3.1 | 2.7 | 3.6 |
| Left rib 7 | Male | 60-69 | 8240 | 389 | 4.7 | 4.3 | 5.2 |
| Left rib 7 | Male | 70-79 | 12836 | 864 | 6.7 | 6.3 | 7.2 |
| Left rib 7 | Male | 80-89 | 6671 | 475 | 7.1 | 6.5 | 7.8 |
| Left rib 7 | Male | 90-99 | 644 | 51 | 7.9 | 6.1 | 10.3 |
| Left rib 7 | Female | 20-29 | 962 | 3 | 0.3 | 0.1 | 0.9 |
| Left rib 7 | Female | 30-39 | 1645 | 9 | 0.5 | 0.3 | 1 |
| Left rib 7 | Female | 40-49 | 3780 | 67 | 1.8 | 1.4 | 2.2 |
| Left rib 7 | Female | 50-59 | 6538 | 424 | 6.5 | 5.9 | 7.1 |
| Left rib 7 | Female | 60-69 | 6934 | 859 | 12.4 | 11.6 | 13.2 |
| Left rib 7 | Female | 70-79 | 9243 | 1668 | 18 | 17.3 | 18.8 |
| Left rib 7 | Female | 80-89 | 5397 | 1091 | 20.2 | 19.2 | 21.3 |
| Left rib 7 | Female | 90-99 | 813 | 186 | 22.9 | 20.1 | 25.9 |
| Left rib 8 | Male | 20-29 | 853 | 2 | 0.2 | 0.1 | 0.9 |
| Left rib 8 | Male | 30-39 | 1268 | 13 | 1 | 0.6 | 1.7 |
| Left rib 8 | Male | 40-49 | 2679 | 73 | 2.7 | 2.2 | 3.4 |
| Left rib 8 | Male | 50-59 | 5431 | 281 | 5.2 | 4.6 | 5.8 |
| Left rib 8 | Male | 60-69 | 8232 | 627 | 7.6 | 7.1 | 8.2 |
| Left rib 8 | Male | 70-79 | 12844 | 1417 | 11 | 10.5 | 11.6 |
| Left rib 8 | Male | 80-89 | 6684 | 938 | 14 | 13.2 | 14.9 |
| Left rib 8 | Male | 90-99 | 647 | 114 | 17.6 | 14.9 | 20.7 |
| Left rib 8 | Female | 20-29 | 965 | 4 | 0.4 | 0.2 | 1.1 |
| Left rib 8 | Female | 30-39 | 1649 | 14 | 0.8 | 0.5 | 1.4 |
| Left rib 8 | Female | 40-49 | 3793 | 141 | 3.7 | 3.2 | 4.4 |
| Left rib 8 | Female | 50-59 | 6566 | 970 | 14.8 | 13.9 | 15.7 |
| Left rib 8 | Female | 60-69 | 6937 | 2036 | 29.3 | 28.3 | 30.4 |
| Left rib 8 | Female | 70-79 | 9223 | 3752 | 40.7 | 39.7 | 41.7 |
| Left rib 8 | Female | 80-89 | 5381 | 2529 | 47 | 45.7 | 48.3 |
| Left rib 8 | Female | 90-99 | 803 | 435 | 54.2 | 50.7 | 57.6 |
| Left rib 9 | Male | 20-29 | 839 | 1 | 0.1 | 0 | 0.7 |
| Left rib 9 | Male | 30-39 | 1273 | 16 | 1.3 | 0.8 | 2 |
| Left rib 9 | Male | 40-49 | 2670 | 72 | 2.7 | 2.1 | 3.4 |
| Left rib 9 | Male | 50-59 | 5438 | 305 | 5.6 | 5 | 6.3 |
| Left rib 9 | Male | 60-69 | 8280 | 694 | 8.4 | 7.8 | 9 |
| Left rib 9 | Male | 70-79 | 12892 | 1567 | 12.2 | 11.6 | 12.7 |
| Left rib 9 | Male | 80-89 | 6665 | 1004 | 15.1 | 14.2 | 15.9 |
| Left rib 9 | Male | 90-99 | 637 | 141 | 22.1 | 19.1 | 25.5 |
| Left rib 9 | Female | 20-29 | 959 | 5 | 0.5 | 0.2 | 1.2 |
| Left rib 9 | Female | 30-39 | 1641 | 25 | 1.5 | 1 | 2.2 |
| Left rib 9 | Female | 40-49 | 3789 | 186 | 4.9 | 4.3 | 5.6 |
| Left rib 9 | Female | 50-59 | 6583 | 1282 | 19.5 | 18.5 | 20.4 |
| Left rib 9 | Female | 60-69 | 6959 | 2559 | 36.8 | 35.6 | 37.9 |
| Left rib 9 | Female | 70-79 | 9243 | 4584 | 49.6 | 48.6 | 50.6 |
| Left rib 9 | Female | 80-89 | 5379 | 3117 | 57.9 | 56.6 | 59.3 |
| Left rib 9 | Female | 90-99 | 800 | 516 | 64.5 | 61.1 | 67.7 |
| Left rib 10 | Male | 20-29 | 782 | 0 | 0 | 0 | 0.5 |
| Left rib 10 | Male | 30-39 | 1254 | 11 | 0.9 | 0.5 | 1.6 |
| Left rib 10 | Male | 40-49 | 2695 | 63 | 2.3 | 1.8 | 3 |
| Left rib 10 | Male | 50-59 | 5473 | 276 | 5 | 4.5 | 5.7 |
| Left rib 10 | Male | 60-69 | 8361 | 629 | 7.5 | 7 | 8.1 |
| Left rib 10 | Male | 70-79 | 12962 | 1402 | 10.8 | 10.3 | 11.4 |
| Left rib 10 | Male | 80-89 | 6629 | 839 | 12.7 | 11.9 | 13.5 |
| Left rib 10 | Male | 90-99 | 621 | 110 | 17.7 | 14.9 | 20.9 |
| Left rib 10 | Female | 20-29 | 942 | 6 | 0.6 | 0.3 | 1.4 |
| Left rib 10 | Female | 30-39 | 1665 | 32 | 1.9 | 1.4 | 2.7 |
| Left rib 10 | Female | 40-49 | 3834 | 210 | 5.5 | 4.8 | 6.2 |
| Left rib 10 | Female | 50-59 | 6648 | 1367 | 20.6 | 19.6 | 21.6 |
| Left rib 10 | Female | 60-69 | 6995 | 2841 | 40.6 | 39.5 | 41.8 |
| Left rib 10 | Female | 70-79 | 9239 | 5044 | 54.6 | 53.6 | 55.6 |
| Left rib 10 | Female | 80-89 | 5311 | 3293 | 62 | 60.7 | 63.3 |
| Left rib 10 | Female | 90-99 | 777 | 520 | 66.9 | 63.5 | 70.1 |
| Left rib 11 | Male | 20-29 | 790 | 1 | 0.1 | 0 | 0.7 |
| Left rib 11 | Male | 30-39 | 1260 | 16 | 1.3 | 0.8 | 2.1 |
| Left rib 11 | Male | 40-49 | 2692 | 62 | 2.3 | 1.8 | 2.9 |
| Left rib 11 | Male | 50-59 | 5512 | 328 | 6 | 5.4 | 6.6 |
| Left rib 11 | Male | 60-69 | 8366 | 855 | 10.2 | 9.6 | 10.9 |
| Left rib 11 | Male | 70-79 | 12959 | 1955 | 15.1 | 14.5 | 15.7 |
| Left rib 11 | Male | 80-89 | 6571 | 1201 | 18.3 | 17.4 | 19.2 |
| Left rib 11 | Male | 90-99 | 613 | 151 | 24.6 | 21.4 | 28.2 |
| Left rib 11 | Female | 20-29 | 947 | 4 | 0.4 | 0.2 | 1.1 |
| Left rib 11 | Female | 30-39 | 1672 | 27 | 1.6 | 1.1 | 2.3 |
| Left rib 11 | Female | 40-49 | 3853 | 170 | 4.4 | 3.8 | 5.1 |
| Left rib 11 | Female | 50-59 | 6692 | 1326 | 19.8 | 18.9 | 20.8 |
| Left rib 11 | Female | 60-69 | 6985 | 3002 | 43 | 41.8 | 44.1 |
| Left rib 11 | Female | 70-79 | 9214 | 5415 | 58.8 | 57.8 | 59.8 |
| Left rib 11 | Female | 80-89 | 5295 | 3545 | 66.9 | 65.7 | 68.2 |
| Left rib 11 | Female | 90-99 | 764 | 533 | 69.8 | 66.4 | 72.9 |
| Left rib 12 | Male | 20-29 | 857 | 2 | 0.2 | 0.1 | 0.8 |
| Left rib 12 | Male | 30-39 | 1294 | 7 | 0.5 | 0.3 | 1.1 |
| Left rib 12 | Male | 40-49 | 2677 | 28 | 1 | 0.7 | 1.5 |
| Left rib 12 | Male | 50-59 | 5403 | 142 | 2.6 | 2.2 | 3.1 |
| Left rib 12 | Male | 60-69 | 8199 | 429 | 5.2 | 4.8 | 5.7 |
| Left rib 12 | Male | 70-79 | 12699 | 1094 | 8.6 | 8.1 | 9.1 |
| Left rib 12 | Male | 80-89 | 6518 | 693 | 10.6 | 9.9 | 11.4 |
| Left rib 12 | Male | 90-99 | 616 | 87 | 14.1 | 11.6 | 17.1 |
| Left rib 12 | Female | 20-29 | 930 | 7 | 0.8 | 0.4 | 1.5 |
| Left rib 12 | Female | 30-39 | 1607 | 9 | 0.6 | 0.3 | 1.1 |
| Left rib 12 | Female | 40-49 | 3692 | 78 | 2.1 | 1.7 | 2.6 |
| Left rib 12 | Female | 50-59 | 6411 | 644 | 10 | 9.3 | 10.8 |
| Left rib 12 | Female | 60-69 | 6749 | 1649 | 24.4 | 23.4 | 25.5 |
| Left rib 12 | Female | 70-79 | 8928 | 3283 | 36.8 | 35.8 | 37.8 |
| Left rib 12 | Female | 80-89 | 5117 | 2245 | 43.9 | 42.5 | 45.2 |
| Left rib 12 | Female | 90-99 | 766 | 348 | 45.4 | 41.9 | 49 |
| Right rib 1 | Male | 20-29 | 374 | 0 | 0 | 0 | 1 |
| Right rib 1 | Male | 30-39 | 788 | 6 | 0.8 | 0.3 | 1.7 |
| Right rib 1 | Male | 40-49 | 1856 | 22 | 1.2 | 0.8 | 1.8 |
| Right rib 1 | Male | 50-59 | 3890 | 162 | 4.2 | 3.6 | 4.8 |
| Right rib 1 | Male | 60-69 | 6139 | 607 | 9.9 | 9.2 | 10.7 |
| Right rib 1 | Male | 70-79 | 9832 | 1780 | 18.1 | 17.4 | 18.9 |
| Right rib 1 | Male | 80-89 | 5139 | 1260 | 24.5 | 23.4 | 25.7 |
| Right rib 1 | Male | 90-99 | 474 | 166 | 35 | 30.9 | 39.4 |
| Right rib 1 | Female | 20-29 | 462 | 2 | 0.4 | 0.1 | 1.6 |
| Right rib 1 | Female | 30-39 | 955 | 12 | 1.3 | 0.7 | 2.2 |
| Right rib 1 | Female | 40-49 | 2571 | 53 | 2.1 | 1.6 | 2.7 |
| Right rib 1 | Female | 50-59 | 4735 | 447 | 9.4 | 8.6 | 10.3 |
| Right rib 1 | Female | 60-69 | 5107 | 1522 | 29.8 | 28.6 | 31.1 |
| Right rib 1 | Female | 70-79 | 6800 | 3448 | 50.7 | 49.5 | 51.9 |
| Right rib 1 | Female | 80-89 | 4103 | 2653 | 64.7 | 63.2 | 66.1 |
| Right rib 1 | Female | 90-99 | 650 | 463 | 71.2 | 67.6 | 74.6 |
| Right rib 2 | Male | 20-29 | 528 | 0 | 0 | 0 | 0.7 |
| Right rib 2 | Male | 30-39 | 830 | 9 | 1.1 | 0.6 | 2 |
| Right rib 2 | Male | 40-49 | 1847 | 44 | 2.4 | 1.8 | 3.2 |
| Right rib 2 | Male | 50-59 | 3840 | 236 | 6.1 | 5.4 | 7 |
| Right rib 2 | Male | 60-69 | 6058 | 555 | 9.2 | 8.5 | 9.9 |
| Right rib 2 | Male | 70-79 | 9756 | 1211 | 12.4 | 11.8 | 13.1 |
| Right rib 2 | Male | 80-89 | 5127 | 708 | 13.8 | 12.9 | 14.8 |
| Right rib 2 | Male | 90-99 | 479 | 78 | 16.3 | 13.2 | 19.9 |
| Right rib 2 | Female | 20-29 | 562 | 3 | 0.5 | 0.2 | 1.6 |
| Right rib 2 | Female | 30-39 | 1031 | 16 | 1.6 | 1 | 2.5 |
| Right rib 2 | Female | 40-49 | 2627 | 121 | 4.6 | 3.9 | 5.5 |
| Right rib 2 | Female | 50-59 | 4755 | 857 | 18 | 17 | 19.1 |
| Right rib 2 | Female | 60-69 | 5070 | 1970 | 38.9 | 37.5 | 40.2 |
| Right rib 2 | Female | 70-79 | 6692 | 3730 | 55.7 | 54.5 | 56.9 |
| Right rib 2 | Female | 80-89 | 4050 | 2612 | 64.5 | 63 | 66 |
| Right rib 2 | Female | 90-99 | 634 | 433 | 68.3 | 64.6 | 71.8 |
| Right rib 3 | Male | 20-29 | 559 | 0 | 0 | 0 | 0.7 |
| Right rib 3 | Male | 30-39 | 872 | 8 | 0.9 | 0.5 | 1.8 |
| Right rib 3 | Male | 40-49 | 1886 | 58 | 3.1 | 2.4 | 4 |
| Right rib 3 | Male | 50-59 | 3893 | 292 | 7.5 | 6.7 | 8.4 |
| Right rib 3 | Male | 60-69 | 6168 | 692 | 11.2 | 10.5 | 12 |
| Right rib 3 | Male | 70-79 | 9916 | 1486 | 15 | 14.3 | 15.7 |
| Right rib 3 | Male | 80-89 | 5205 | 865 | 16.6 | 15.6 | 17.7 |
| Right rib 3 | Male | 90-99 | 494 | 106 | 21.5 | 18.1 | 25.3 |
| Right rib 3 | Female | 20-29 | 578 | 2 | 0.3 | 0.1 | 1.3 |
| Right rib 3 | Female | 30-39 | 1043 | 13 | 1.2 | 0.7 | 2.1 |
| Right rib 3 | Female | 40-49 | 2667 | 163 | 6.1 | 5.3 | 7.1 |
| Right rib 3 | Female | 50-59 | 4812 | 1147 | 23.8 | 22.7 | 25.1 |
| Right rib 3 | Female | 60-69 | 5183 | 2390 | 46.1 | 44.8 | 47.5 |
| Right rib 3 | Female | 70-79 | 6951 | 4194 | 60.3 | 59.2 | 61.5 |
| Right rib 3 | Female | 80-89 | 4214 | 2874 | 68.2 | 66.8 | 69.6 |
| Right rib 3 | Female | 90-99 | 666 | 489 | 73.4 | 69.9 | 76.6 |
| Right rib 4 | Male | 20-29 | 662 | 1 | 0.2 | 0 | 0.9 |
| Right rib 4 | Male | 30-39 | 977 | 13 | 1.3 | 0.8 | 2.3 |
| Right rib 4 | Male | 40-49 | 2106 | 46 | 2.2 | 1.6 | 2.9 |
| Right rib 4 | Male | 50-59 | 4305 | 222 | 5.2 | 4.5 | 5.9 |
| Right rib 4 | Male | 60-69 | 6679 | 486 | 7.3 | 6.7 | 7.9 |
| Right rib 4 | Male | 70-79 | 10741 | 965 | 9 | 8.5 | 9.5 |
| Right rib 4 | Male | 80-89 | 5682 | 562 | 9.9 | 9.1 | 10.7 |
| Right rib 4 | Male | 90-99 | 559 | 62 | 11.1 | 8.7 | 14 |
| Right rib 4 | Female | 20-29 | 726 | 3 | 0.4 | 0.1 | 1.2 |
| Right rib 4 | Female | 30-39 | 1228 | 16 | 1.3 | 0.8 | 2.1 |
| Right rib 4 | Female | 40-49 | 2964 | 61 | 2.1 | 1.6 | 2.6 |
| Right rib 4 | Female | 50-59 | 5316 | 336 | 6.3 | 5.7 | 7 |
| Right rib 4 | Female | 60-69 | 5727 | 744 | 13 | 12.1 | 13.9 |
| Right rib 4 | Female | 70-79 | 7734 | 1341 | 17.3 | 16.5 | 18.2 |
| Right rib 4 | Female | 80-89 | 4696 | 935 | 19.9 | 18.8 | 21.1 |
| Right rib 4 | Female | 90-99 | 754 | 176 | 23.3 | 20.5 | 26.5 |
| Right rib 5 | Male | 20-29 | 785 | 1 | 0.1 | 0 | 0.7 |
| Right rib 5 | Male | 30-39 | 1150 | 14 | 1.2 | 0.7 | 2 |
| Right rib 5 | Male | 40-49 | 2460 | 38 | 1.5 | 1.1 | 2.1 |
| Right rib 5 | Male | 50-59 | 4978 | 136 | 2.7 | 2.3 | 3.2 |
| Right rib 5 | Male | 60-69 | 7620 | 281 | 3.7 | 3.3 | 4.1 |
| Right rib 5 | Male | 70-79 | 12127 | 493 | 4.1 | 3.7 | 4.4 |
| Right rib 5 | Male | 80-89 | 6356 | 269 | 4.2 | 3.8 | 4.8 |
| Right rib 5 | Male | 90-99 | 632 | 33 | 5.2 | 3.7 | 7.2 |
| Right rib 5 | Female | 20-29 | 895 | 0 | 0 | 0 | 0.4 |
| Right rib 5 | Female | 30-39 | 1521 | 1 | 0.1 | 0 | 0.4 |
| Right rib 5 | Female | 40-49 | 3486 | 16 | 0.5 | 0.3 | 0.7 |
| Right rib 5 | Female | 50-59 | 6086 | 129 | 2.1 | 1.8 | 2.5 |
| Right rib 5 | Female | 60-69 | 6530 | 232 | 3.6 | 3.1 | 4 |
| Right rib 5 | Female | 70-79 | 8797 | 358 | 4.1 | 3.7 | 4.5 |
| Right rib 5 | Female | 80-89 | 5217 | 227 | 4.4 | 3.8 | 4.9 |
| Right rib 5 | Female | 90-99 | 803 | 32 | 4 | 2.8 | 5.6 |
| Right rib 6 | Male | 20-29 | 854 | 2 | 0.2 | 0.1 | 0.8 |
| Right rib 6 | Male | 30-39 | 1277 | 6 | 0.5 | 0.2 | 1 |
| Right rib 6 | Male | 40-49 | 2678 | 31 | 1.2 | 0.8 | 1.6 |
| Right rib 6 | Male | 50-59 | 5355 | 129 | 2.4 | 2 | 2.9 |
| Right rib 6 | Male | 60-69 | 8159 | 263 | 3.2 | 2.9 | 3.6 |
| Right rib 6 | Male | 70-79 | 12806 | 497 | 3.9 | 3.6 | 4.2 |
| Right rib 6 | Male | 80-89 | 6669 | 273 | 4.1 | 3.6 | 4.6 |
| Right rib 6 | Male | 90-99 | 652 | 35 | 5.4 | 3.9 | 7.4 |
| Right rib 6 | Female | 20-29 | 965 | 3 | 0.3 | 0.1 | 0.9 |
| Right rib 6 | Female | 30-39 | 1644 | 9 | 0.5 | 0.3 | 1 |
| Right rib 6 | Female | 40-49 | 3749 | 36 | 1 | 0.7 | 1.3 |
| Right rib 6 | Female | 50-59 | 6507 | 242 | 3.7 | 3.3 | 4.2 |
| Right rib 6 | Female | 60-69 | 6904 | 440 | 6.4 | 5.8 | 7 |
| Right rib 6 | Female | 70-79 | 9234 | 802 | 8.7 | 8.1 | 9.3 |
| Right rib 6 | Female | 80-89 | 5396 | 483 | 9 | 8.2 | 9.7 |
| Right rib 6 | Female | 90-99 | 815 | 77 | 9.4 | 7.6 | 11.7 |
| Right rib 7 | Male | 20-29 | 859 | 3 | 0.3 | 0.1 | 1 |
| Right rib 7 | Male | 30-39 | 1270 | 10 | 0.8 | 0.4 | 1.4 |
| Right rib 7 | Male | 40-49 | 2689 | 55 | 2 | 1.6 | 2.7 |
| Right rib 7 | Male | 50-59 | 5397 | 190 | 3.5 | 3.1 | 4 |
| Right rib 7 | Male | 60-69 | 8215 | 451 | 5.5 | 5 | 6 |
| Right rib 7 | Male | 70-79 | 12839 | 940 | 7.3 | 6.9 | 7.8 |
| Right rib 7 | Male | 80-89 | 6677 | 563 | 8.4 | 7.8 | 9.1 |
| Right rib 7 | Male | 90-99 | 649 | 74 | 11.4 | 9.2 | 14.1 |
| Right rib 7 | Female | 20-29 | 965 | 2 | 0.2 | 0.1 | 0.8 |
| Right rib 7 | Female | 30-39 | 1647 | 9 | 0.5 | 0.3 | 1 |
| Right rib 7 | Female | 40-49 | 3780 | 69 | 1.8 | 1.4 | 2.3 |
| Right rib 7 | Female | 50-59 | 6554 | 410 | 6.3 | 5.7 | 6.9 |
| Right rib 7 | Female | 60-69 | 6918 | 870 | 12.6 | 11.8 | 13.4 |
| Right rib 7 | Female | 70-79 | 9256 | 1760 | 19 | 18.2 | 19.8 |
| Right rib 7 | Female | 80-89 | 5380 | 1183 | 22 | 20.9 | 23.1 |
| Right rib 7 | Female | 90-99 | 807 | 206 | 25.5 | 22.6 | 28.6 |
| Right rib 8 | Male | 20-29 | 846 | 3 | 0.4 | 0.1 | 1 |
| Right rib 8 | Male | 30-39 | 1261 | 12 | 1 | 0.5 | 1.7 |
| Right rib 8 | Male | 40-49 | 2672 | 82 | 3.1 | 2.5 | 3.8 |
| Right rib 8 | Male | 50-59 | 5415 | 302 | 5.6 | 5 | 6.2 |
| Right rib 8 | Male | 60-69 | 8236 | 639 | 7.8 | 7.2 | 8.4 |
| Right rib 8 | Male | 70-79 | 12854 | 1436 | 11.2 | 10.6 | 11.7 |
| Right rib 8 | Male | 80-89 | 6688 | 899 | 13.4 | 12.6 | 14.3 |
| Right rib 8 | Male | 90-99 | 649 | 113 | 17.4 | 14.7 | 20.5 |
| Right rib 8 | Female | 20-29 | 970 | 5 | 0.5 | 0.2 | 1.2 |
| Right rib 8 | Female | 30-39 | 1642 | 16 | 1 | 0.6 | 1.6 |
| Right rib 8 | Female | 40-49 | 3775 | 123 | 3.3 | 2.7 | 3.9 |
| Right rib 8 | Female | 50-59 | 6543 | 788 | 12 | 11.3 | 12.9 |
| Right rib 8 | Female | 60-69 | 6923 | 1604 | 23.2 | 22.2 | 24.2 |
| Right rib 8 | Female | 70-79 | 9249 | 3148 | 34 | 33.1 | 35 |
| Right rib 8 | Female | 80-89 | 5392 | 2219 | 41.2 | 39.8 | 42.5 |
| Right rib 8 | Female | 90-99 | 808 | 379 | 46.9 | 43.5 | 50.4 |
| Right rib 9 | Male | 20-29 | 828 | 1 | 0.1 | 0 | 0.7 |
| Right rib 9 | Male | 30-39 | 1263 | 13 | 1 | 0.6 | 1.8 |
| Right rib 9 | Male | 40-49 | 2678 | 79 | 2.9 | 2.4 | 3.7 |
| Right rib 9 | Male | 50-59 | 5419 | 321 | 5.9 | 5.3 | 6.6 |
| Right rib 9 | Male | 60-69 | 8289 | 701 | 8.5 | 7.9 | 9.1 |
| Right rib 9 | Male | 70-79 | 12876 | 1485 | 11.5 | 11 | 12.1 |
| Right rib 9 | Male | 80-89 | 6683 | 896 | 13.4 | 12.6 | 14.2 |
| Right rib 9 | Male | 90-99 | 641 | 113 | 17.6 | 14.9 | 20.8 |
| Right rib 9 | Female | 20-29 | 955 | 3 | 0.3 | 0.1 | 0.9 |
| Right rib 9 | Female | 30-39 | 1641 | 29 | 1.8 | 1.2 | 2.5 |
| Right rib 9 | Female | 40-49 | 3770 | 189 | 5 | 4.4 | 5.8 |
| Right rib 9 | Female | 50-59 | 6568 | 1173 | 17.9 | 17 | 18.8 |
| Right rib 9 | Female | 60-69 | 6969 | 2340 | 33.6 | 32.5 | 34.7 |
| Right rib 9 | Female | 70-79 | 9247 | 4044 | 43.7 | 42.7 | 44.7 |
| Right rib 9 | Female | 80-89 | 5384 | 2742 | 50.9 | 49.6 | 52.3 |
| Right rib 9 | Female | 90-99 | 808 | 486 | 60.1 | 56.7 | 63.5 |
| Right rib 10 | Male | 20-29 | 785 | 1 | 0.1 | 0 | 0.7 |
| Right rib 10 | Male | 30-39 | 1240 | 16 | 1.3 | 0.8 | 2.1 |
| Right rib 10 | Male | 40-49 | 2669 | 64 | 2.4 | 1.9 | 3.1 |
| Right rib 10 | Male | 50-59 | 5466 | 254 | 4.6 | 4.1 | 5.2 |
| Right rib 10 | Male | 60-69 | 8359 | 590 | 7.1 | 6.5 | 7.6 |
| Right rib 10 | Male | 70-79 | 12952 | 1170 | 9 | 8.6 | 9.5 |
| Right rib 10 | Male | 80-89 | 6651 | 639 | 9.6 | 8.9 | 10.3 |
| Right rib 10 | Male | 90-99 | 626 | 80 | 12.8 | 10.4 | 15.6 |
| Right rib 10 | Female | 20-29 | 942 | 7 | 0.7 | 0.4 | 1.5 |
| Right rib 10 | Female | 30-39 | 1652 | 25 | 1.5 | 1 | 2.2 |
| Right rib 10 | Female | 40-49 | 3810 | 191 | 5 | 4.4 | 5.8 |
| Right rib 10 | Female | 50-59 | 6616 | 1231 | 18.6 | 17.7 | 19.6 |
| Right rib 10 | Female | 60-69 | 6991 | 2604 | 37.2 | 36.1 | 38.4 |
| Right rib 10 | Female | 70-79 | 9257 | 4430 | 47.9 | 46.8 | 48.9 |
| Right rib 10 | Female | 80-89 | 5355 | 2958 | 55.2 | 53.9 | 56.6 |
| Right rib 10 | Female | 90-99 | 795 | 491 | 61.8 | 58.3 | 65.1 |
| Right rib 11 | Male | 20-29 | 805 | 1 | 0.1 | 0 | 0.7 |
| Right rib 11 | Male | 30-39 | 1272 | 15 | 1.2 | 0.7 | 1.9 |
| Right rib 11 | Male | 40-49 | 2690 | 68 | 2.5 | 2 | 3.2 |
| Right rib 11 | Male | 50-59 | 5503 | 329 | 6 | 5.4 | 6.6 |
| Right rib 11 | Male | 60-69 | 8382 | 798 | 9.5 | 8.9 | 10.2 |
| Right rib 11 | Male | 70-79 | 12942 | 1614 | 12.5 | 11.9 | 13.1 |
| Right rib 11 | Male | 80-89 | 6565 | 905 | 13.8 | 13 | 14.6 |
| Right rib 11 | Male | 90-99 | 610 | 99 | 16.2 | 13.5 | 19.4 |
| Right rib 11 | Female | 20-29 | 950 | 5 | 0.5 | 0.2 | 1.2 |
| Right rib 11 | Female | 30-39 | 1673 | 21 | 1.3 | 0.8 | 1.9 |
| Right rib 11 | Female | 40-49 | 3832 | 165 | 4.3 | 3.7 | 5 |
| Right rib 11 | Female | 50-59 | 6684 | 1266 | 18.9 | 18 | 19.9 |
| Right rib 11 | Female | 60-69 | 6992 | 2853 | 40.8 | 39.7 | 42 |
| Right rib 11 | Female | 70-79 | 9247 | 5060 | 54.7 | 53.7 | 55.7 |
| Right rib 11 | Female | 80-89 | 5291 | 3328 | 62.9 | 61.6 | 64.2 |
| Right rib 11 | Female | 90-99 | 776 | 491 | 63.3 | 59.8 | 66.6 |
| Right rib 12 | Male | 20-29 | 873 | 3 | 0.3 | 0.1 | 1 |
| Right rib 12 | Male | 30-39 | 1315 | 8 | 0.6 | 0.3 | 1.2 |
| Right rib 12 | Male | 40-49 | 2676 | 27 | 1 | 0.7 | 1.5 |
| Right rib 12 | Male | 50-59 | 5409 | 167 | 3.1 | 2.7 | 3.6 |
| Right rib 12 | Male | 60-69 | 8182 | 413 | 5 | 4.6 | 5.5 |
| Right rib 12 | Male | 70-79 | 12685 | 924 | 7.3 | 6.8 | 7.7 |
| Right rib 12 | Male | 80-89 | 6509 | 544 | 8.4 | 7.7 | 9.1 |
| Right rib 12 | Male | 90-99 | 627 | 59 | 9.4 | 7.4 | 11.9 |
| Right rib 12 | Female | 20-29 | 944 | 2 | 0.2 | 0.1 | 0.8 |
| Right rib 12 | Female | 30-39 | 1635 | 17 | 1 | 0.7 | 1.7 |
| Right rib 12 | Female | 40-49 | 3773 | 95 | 2.5 | 2.1 | 3.1 |
| Right rib 12 | Female | 50-59 | 6549 | 698 | 10.7 | 9.9 | 11.4 |
| Right rib 12 | Female | 60-69 | 6842 | 1676 | 24.5 | 23.5 | 25.5 |
| Right rib 12 | Female | 70-79 | 9066 | 3226 | 35.6 | 34.6 | 36.6 |
| Right rib 12 | Female | 80-89 | 5142 | 2118 | 41.2 | 39.9 | 42.5 |
| Right rib 12 | Female | 90-99 | 755 | 293 | 38.8 | 35.4 | 42.3 |

Score = (measured attenuation − young-adult mean) / young-adult SD for the same site and sex (Supplementary Table 5). Intervals are Wilson 95% intervals.

**Supplementary Table 7: Sensitivity of the trajectories to the quality-control rule (Sample size)**

| **Site** | **Sex** | **Analyzed under primary rule, n** | **Analyzed under legacy rule, n** |
| --- | --- | --- | --- |
| Left rib 1 | Female | 25521 | 22399 |
| Left rib 1 | Male | 28579 | 24141 |
| Left rib 10 | Female | 35656 | 31133 |
| Left rib 10 | Male | 38979 | 32699 |
| Left rib 11 | Female | 35672 | 30790 |
| Left rib 11 | Male | 38973 | 33015 |
| Left rib 12 | Female | 34465 | 30516 |
| Left rib 12 | Male | 38542 | 32219 |
| Left rib 2 | Female | 25557 | 22192 |
| Left rib 2 | Male | 28631 | 24239 |
| Left rib 3 | Female | 26333 | 23432 |
| Left rib 3 | Male | 29200 | 24326 |
| Left rib 4 | Female | 29542 | 26798 |
| Left rib 4 | Male | 32094 | 26416 |
| Left rib 5 | Female | 33829 | 30571 |
| Left rib 5 | Male | 36632 | 30221 |
| Left rib 6 | Female | 35514 | 31749 |
| Left rib 6 | Male | 38749 | 32242 |
| Left rib 7 | Female | 35584 | 31287 |
| Left rib 7 | Male | 38883 | 32646 |
| Left rib 8 | Female | 35590 | 31290 |
| Left rib 8 | Male | 38911 | 32579 |
| Left rib 9 | Female | 35619 | 31370 |
| Left rib 9 | Male | 38937 | 32432 |
| Right rib 1 | Female | 25496 | 22326 |
| Right rib 1 | Male | 28562 | 24221 |
| Right rib 10 | Female | 35668 | 31061 |
| Right rib 10 | Male | 38951 | 32751 |
| Right rib 11 | Female | 35692 | 30771 |
| Right rib 11 | Male | 38989 | 33092 |
| Right rib 12 | Female | 34973 | 30781 |
| Right rib 12 | Male | 38562 | 32311 |
| Right rib 2 | Female | 25591 | 22171 |
| Right rib 2 | Male | 28622 | 24217 |
| Right rib 3 | Female | 26297 | 23354 |
| Right rib 3 | Male | 29164 | 24293 |
| Right rib 4 | Female | 29371 | 26578 |
| Right rib 4 | Male | 31936 | 26328 |
| Right rib 5 | Female | 33600 | 30135 |
| Right rib 5 | Male | 36386 | 30213 |
| Right rib 6 | Female | 35489 | 31752 |
| Right rib 6 | Male | 38734 | 32187 |
| Right rib 7 | Female | 35581 | 31325 |
| Right rib 7 | Male | 38875 | 32637 |
| Right rib 8 | Female | 35573 | 31317 |
| Right rib 8 | Male | 38895 | 32538 |
| Right rib 9 | Female | 35609 | 31409 |
| Right rib 9 | Male | 38925 | 32455 |
| C1 | Female | 13354 | 11627 |
| C1 | Male | 12941 | 10734 |
| C2 | Female | 13349 | 11491 |
| C2 | Male | 12939 | 10904 |
| C3 | Female | 14293 | 12355 |
| C3 | Male | 13945 | 11738 |
| C4 | Female | 15355 | 13201 |
| C4 | Male | 15517 | 13168 |
| C5 | Female | 17881 | 15446 |
| C5 | Male | 19024 | 16059 |
| C6 | Female | 22603 | 19771 |
| C6 | Male | 25074 | 21014 |
| C7 | Female | 25131 | 21690 |
| C7 | Male | 28177 | 23888 |
| L1 | Female | 35498 | 30893 |
| L1 | Male | 38886 | 33147 |
| L2 | Female | 35527 | 30650 |
| L2 | Male | 38881 | 33336 |
| L3 | Female | 35508 | 30399 |
| L3 | Male | 38881 | 33687 |
| L4 | Female | 35508 | 30702 |
| L4 | Male | 38879 | 33381 |
| L5 | Female | 35508 | 30574 |
| L5 | Male | 38902 | 33335 |
| S1 | Female | 35559 | 30766 |
| S1 | Male | 38908 | 33083 |
| T1 | Female | 25491 | 21972 |
| T1 | Male | 28549 | 24208 |
| T10 | Female | 35011 | 30344 |
| T10 | Male | 38463 | 32669 |
| T11 | Female | 35559 | 30827 |
| T11 | Male | 38907 | 33125 |
| T12 | Female | 35495 | 30809 |
| T12 | Male | 38876 | 33182 |
| T2 | Female | 25469 | 21881 |
| T2 | Male | 28532 | 24293 |
| T3 | Female | 25483 | 21866 |
| T3 | Male | 28568 | 24348 |
| T4 | Female | 25485 | 21888 |
| T4 | Male | 28569 | 24373 |
| T5 | Female | 25546 | 21906 |
| T5 | Male | 28635 | 24459 |
| T6 | Female | 26056 | 22635 |
| T6 | Male | 29225 | 24711 |
| T7 | Female | 27854 | 24881 |
| T7 | Male | 31300 | 26042 |
| T8 | Female | 30457 | 27422 |
| T8 | Male | 34324 | 28513 |
| T9 | Female | 33251 | 29575 |
| T9 | Male | 37013 | 30859 |

**Supplementary Table 7 (continued): Sensitivity of the trajectories to the quality-control rule (Fitted attenuation)**

| **Site** | **Sex** | **Attenuation at 50 y, primary rule (HU)** | **Attenuation at 50 y, legacy rule (HU)** | **Difference at 50 y (HU)** | **Attenuation at 70 y, primary rule (HU)** | **Attenuation at 70 y, legacy rule (HU)** | **Difference at 70 y (HU)** |
| --- | --- | --- | --- | --- | --- | --- | --- |
| Left rib 1 | Female | 346.907 | 343.175 | 3.732 | 265.479 | 269.151 | -3.671 |
| Left rib 1 | Male | 328.141 | 326.011 | 2.13 | 291.795 | 281.103 | 10.692 |
| Left rib 10 | Female | 376.335 | 366.613 | 9.722 | 286.269 | 286.518 | -0.249 |
| Left rib 10 | Male | 323.161 | 330.336 | -7.175 | 302.466 | 296.911 | 5.556 |
| Left rib 11 | Female | 383.483 | 374.151 | 9.332 | 283.899 | 285.184 | -1.285 |
| Left rib 11 | Male | 336.132 | 342.665 | -6.534 | 306.518 | 300.147 | 6.372 |
| Left rib 12 | Female | 379.268 | 367.283 | 11.985 | 285.816 | 283.007 | 2.809 |
| Left rib 12 | Male | 322.743 | 326.765 | -4.022 | 294.319 | 288.291 | 6.028 |
| Left rib 2 | Female | 284 | 276.812 | 7.188 | 215.157 | 216.885 | -1.728 |
| Left rib 2 | Male | 245.419 | 249.137 | -3.719 | 230.608 | 224.417 | 6.191 |
| Left rib 3 | Female | 284.049 | 274.624 | 9.424 | 209.794 | 207.722 | 2.072 |
| Left rib 3 | Male | 234.389 | 238.613 | -4.224 | 212.439 | 208.761 | 3.678 |
| Left rib 4 | Female | 286.135 | 278.01 | 8.125 | 211.61 | 212.381 | -0.772 |
| Left rib 4 | Male | 240.389 | 243.299 | -2.909 | 220.633 | 216.885 | 3.748 |
| Left rib 5 | Female | 285.626 | 277.82 | 7.806 | 214.309 | 215.203 | -0.894 |
| Left rib 5 | Male | 237.26 | 239.979 | -2.72 | 221.916 | 217.535 | 4.381 |
| Left rib 6 | Female | 299.663 | 290.821 | 8.843 | 227.263 | 227.325 | -0.062 |
| Left rib 6 | Male | 248.436 | 250.636 | -2.2 | 234.441 | 228.833 | 5.609 |
| Left rib 7 | Female | 317.232 | 308.071 | 9.161 | 241.256 | 240.168 | 1.089 |
| Left rib 7 | Male | 259.263 | 263.19 | -3.927 | 241.957 | 237.582 | 4.375 |
| Left rib 8 | Female | 332.208 | 322.543 | 9.665 | 249.83 | 249.087 | 0.743 |
| Left rib 8 | Male | 277.578 | 282.413 | -4.835 | 255.255 | 250.902 | 4.353 |
| Left rib 9 | Female | 343.479 | 334.132 | 9.348 | 258.861 | 259.369 | -0.508 |
| Left rib 9 | Male | 297.575 | 302.038 | -4.463 | 276.101 | 269.595 | 6.506 |
| Right rib 1 | Female | 346.646 | 343.318 | 3.328 | 263.202 | 267.041 | -3.839 |
| Right rib 1 | Male | 328.277 | 326.282 | 1.994 | 292.17 | 281.241 | 10.929 |
| Right rib 10 | Female | 379.254 | 369.206 | 10.047 | 296.852 | 295.537 | 1.315 |
| Right rib 10 | Male | 323.219 | 330.778 | -7.56 | 306.455 | 302.203 | 4.253 |
| Right rib 11 | Female | 384.703 | 375.461 | 9.242 | 289.839 | 291.146 | -1.307 |
| Right rib 11 | Male | 336.548 | 342.864 | -6.316 | 310.825 | 304.482 | 6.343 |
| Right rib 12 | Female | 375.181 | 364.988 | 10.193 | 287.043 | 285.778 | 1.264 |
| Right rib 12 | Male | 319.909 | 324.78 | -4.871 | 297.593 | 291.574 | 6.02 |
| Right rib 2 | Female | 287.049 | 279.398 | 7.652 | 217.934 | 217.945 | -0.011 |
| Right rib 2 | Male | 246.267 | 249.983 | -3.716 | 231.65 | 225.655 | 5.994 |
| Right rib 3 | Female | 283.858 | 274.077 | 9.781 | 209.441 | 206.168 | 3.273 |
| Right rib 3 | Male | 231.031 | 235.883 | -4.852 | 209.028 | 206.257 | 2.771 |
| Right rib 4 | Female | 290.982 | 281.786 | 9.196 | 218.326 | 217.186 | 1.14 |
| Right rib 4 | Male | 238.422 | 241.571 | -3.149 | 219.001 | 216.021 | 2.98 |
| Right rib 5 | Female | 288.009 | 280.104 | 7.905 | 218.818 | 220.08 | -1.262 |
| Right rib 5 | Male | 236.815 | 239.265 | -2.45 | 223.364 | 219.894 | 3.471 |
| Right rib 6 | Female | 306.34 | 296.468 | 9.872 | 231.698 | 230.975 | 0.723 |
| Right rib 6 | Male | 249.313 | 251.564 | -2.251 | 234.949 | 229.607 | 5.342 |
| Right rib 7 | Female | 322.037 | 312.536 | 9.5 | 245.585 | 244.134 | 1.451 |
| Right rib 7 | Male | 262.172 | 266.414 | -4.242 | 244.597 | 240.419 | 4.178 |
| Right rib 8 | Female | 334.794 | 324.958 | 9.836 | 256.882 | 255.148 | 1.734 |
| Right rib 8 | Male | 277.205 | 282.628 | -5.423 | 256.38 | 252.494 | 3.886 |
| Right rib 9 | Female | 344.011 | 334.209 | 9.802 | 266.917 | 265.45 | 1.467 |
| Right rib 9 | Male | 294.148 | 299.278 | -5.13 | 276.033 | 270.877 | 5.156 |
| C1 | Female | 469.968 | 464.869 | 5.099 | 390.373 | 395.169 | -4.796 |
| C1 | Male | 447.364 | 450.356 | -2.992 | 446.901 | 437.47 | 9.431 |
| C2 | Female | 442.333 | 436.291 | 6.042 | 346.992 | 351.825 | -4.833 |
| C2 | Male | 414.466 | 419.181 | -4.715 | 394.491 | 383.945 | 10.546 |
| C3 | Female | 428.673 | 422.094 | 6.579 | 347.655 | 354.423 | -6.768 |
| C3 | Male | 401.454 | 403.354 | -1.9 | 398.373 | 385.997 | 12.375 |
| C4 | Female | 447.865 | 440.874 | 6.991 | 370.829 | 376.288 | -5.459 |
| C4 | Male | 416.739 | 417.277 | -0.538 | 420.401 | 407.235 | 13.167 |
| C5 | Female | 436.483 | 428.368 | 8.115 | 358.017 | 362.797 | -4.78 |
| C5 | Male | 399.362 | 402.073 | -2.711 | 402.04 | 391.721 | 10.319 |
| C6 | Female | 407.358 | 399.319 | 8.039 | 334.633 | 338.498 | -3.865 |
| C6 | Male | 370.72 | 373.39 | -2.669 | 373.569 | 363.802 | 9.768 |
| C7 | Female | 358.41 | 351.909 | 6.501 | 296.11 | 298.61 | -2.5 |
| C7 | Male | 330.531 | 333.724 | -3.193 | 333.179 | 325.017 | 8.161 |
| L1 | Female | 246.786 | 243.243 | 3.543 | 197.235 | 198.943 | -1.708 |
| L1 | Male | 232.031 | 234.884 | -2.853 | 221.25 | 216.632 | 4.618 |
| L2 | Female | 258.799 | 255.292 | 3.507 | 210.804 | 212.525 | -1.721 |
| L2 | Male | 243.541 | 245.947 | -2.406 | 237.293 | 232.118 | 5.175 |
| L3 | Female | 265.145 | 261.926 | 3.219 | 213.646 | 215.976 | -2.331 |
| L3 | Male | 250.706 | 252.132 | -1.426 | 243.803 | 238.988 | 4.815 |
| L4 | Female | 271.407 | 267.925 | 3.482 | 220.228 | 223.405 | -3.177 |
| L4 | Male | 254.787 | 257.497 | -2.71 | 255.924 | 249.005 | 6.919 |
| L5 | Female | 273.915 | 269.545 | 4.37 | 223.254 | 225.15 | -1.897 |
| L5 | Male | 257.47 | 258.992 | -1.522 | 261.413 | 253.536 | 7.878 |
| S1 | Female | 292.51 | 288.736 | 3.774 | 220.27 | 222.691 | -2.421 |
| S1 | Male | 272.298 | 275.379 | -3.081 | 248.169 | 242.43 | 5.739 |
| T1 | Female | 320.541 | 315.608 | 4.933 | 257.424 | 259.565 | -2.14 |
| T1 | Male | 294.318 | 298.816 | -4.499 | 283.522 | 278.708 | 4.814 |
| T10 | Female | 273.539 | 270.296 | 3.244 | 217.344 | 220.158 | -2.815 |
| T10 | Male | 259.725 | 262.602 | -2.878 | 249.734 | 243.724 | 6.01 |
| T11 | Female | 262.436 | 258.767 | 3.669 | 206.923 | 208.826 | -1.903 |
| T11 | Male | 247.399 | 250.591 | -3.192 | 235.039 | 230.06 | 4.978 |
| T12 | Female | 246.147 | 242.381 | 3.766 | 192.891 | 194.68 | -1.789 |
| T12 | Male | 230.988 | 234.19 | -3.202 | 216.957 | 212.81 | 4.147 |
| T2 | Female | 298.13 | 293.409 | 4.722 | 239.501 | 242.022 | -2.521 |
| T2 | Male | 273.275 | 276.865 | -3.59 | 264.163 | 259.333 | 4.83 |
| T3 | Female | 294.597 | 290.952 | 3.645 | 233.374 | 236.221 | -2.846 |
| T3 | Male | 275.12 | 277.531 | -2.411 | 263.176 | 257.738 | 5.438 |
| T4 | Female | 289.588 | 286.402 | 3.186 | 227.059 | 230.522 | -3.463 |
| T4 | Male | 272.996 | 275.235 | -2.239 | 260.184 | 254.017 | 6.167 |
| T5 | Female | 284.397 | 281.271 | 3.126 | 224.271 | 227.81 | -3.539 |
| T5 | Male | 268.492 | 270.525 | -2.033 | 257.203 | 251.116 | 6.088 |
| T6 | Female | 281.195 | 278.547 | 2.649 | 224.158 | 227.08 | -2.922 |
| T6 | Male | 268.252 | 270.036 | -1.785 | 257.246 | 250.892 | 6.354 |
| T7 | Female | 275.417 | 273.65 | 1.767 | 220.879 | 224.636 | -3.757 |
| T7 | Male | 264.407 | 266.487 | -2.08 | 254.342 | 248.782 | 5.56 |
| T8 | Female | 269.736 | 268.492 | 1.243 | 216.497 | 220.866 | -4.369 |
| T8 | Male | 260.842 | 263.094 | -2.252 | 251.619 | 246.327 | 5.292 |
| T9 | Female | 274.726 | 272.894 | 1.832 | 219.725 | 223.735 | -4.01 |
| T9 | Male | 263.728 | 266.284 | -2.556 | 254.757 | 248.64 | 6.117 |

**Supplementary Table 7 (continued): Sensitivity of the trajectories to the quality-control rule (Rate of change)**

| **Site** | **Sex** | **Change 45–60 y, primary rule (HU/y)** | **Change 45–60 y, legacy rule (HU/y)** | **Difference in change 45–60 y (HU/y)** |
| --- | --- | --- | --- | --- |
| Left rib 1 | Female | -4.528 | -4.136 | -0.392 |
| Left rib 1 | Male | -1.847 | -2.155 | 0.308 |
| Left rib 10 | Female | -6.241 | -5.784 | -0.457 |
| Left rib 10 | Male | -1.596 | -2.317 | 0.721 |
| Left rib 11 | Female | -6.913 | -6.323 | -0.59 |
| Left rib 11 | Male | -2.075 | -2.809 | 0.734 |
| Left rib 12 | Female | -6.38 | -5.809 | -0.571 |
| Left rib 12 | Male | -2.033 | -2.563 | 0.531 |
| Left rib 2 | Female | -4.378 | -4.016 | -0.362 |
| Left rib 2 | Male | -1.072 | -1.627 | 0.554 |
| Left rib 3 | Female | -5.362 | -4.96 | -0.403 |
| Left rib 3 | Male | -1.797 | -2.214 | 0.417 |
| Left rib 4 | Female | -5.461 | -5.008 | -0.453 |
| Left rib 4 | Male | -1.708 | -2.073 | 0.366 |
| Left rib 5 | Female | -5.224 | -4.735 | -0.489 |
| Left rib 5 | Male | -1.495 | -1.884 | 0.389 |
| Left rib 6 | Female | -5.001 | -4.575 | -0.425 |
| Left rib 6 | Male | -1.332 | -1.753 | 0.421 |
| Left rib 7 | Female | -5.436 | -5.067 | -0.37 |
| Left rib 7 | Male | -1.442 | -1.895 | 0.453 |
| Left rib 8 | Female | -5.982 | -5.567 | -0.415 |
| Left rib 8 | Male | -1.661 | -2.183 | 0.523 |
| Left rib 9 | Female | -6.02 | -5.517 | -0.503 |
| Left rib 9 | Male | -1.536 | -2.184 | 0.648 |
| Right rib 1 | Female | -4.589 | -4.228 | -0.361 |
| Right rib 1 | Male | -1.847 | -2.12 | 0.273 |
| Right rib 10 | Female | -5.938 | -5.485 | -0.453 |
| Right rib 10 | Male | -1.451 | -2.092 | 0.641 |
| Right rib 11 | Female | -6.674 | -6.125 | -0.55 |
| Right rib 11 | Male | -1.946 | -2.68 | 0.734 |
| Right rib 12 | Female | -6.13 | -5.722 | -0.408 |
| Right rib 12 | Male | -1.838 | -2.377 | 0.539 |
| Right rib 2 | Female | -4.516 | -4.121 | -0.395 |
| Right rib 2 | Male | -1.064 | -1.595 | 0.531 |
| Right rib 3 | Female | -5.371 | -5.019 | -0.351 |
| Right rib 3 | Male | -1.827 | -2.204 | 0.377 |
| Right rib 4 | Female | -5.431 | -5.008 | -0.424 |
| Right rib 4 | Male | -1.756 | -2.087 | 0.331 |
| Right rib 5 | Female | -4.961 | -4.517 | -0.444 |
| Right rib 5 | Male | -1.412 | -1.727 | 0.315 |
| Right rib 6 | Female | -5.104 | -4.675 | -0.429 |
| Right rib 6 | Male | -1.388 | -1.782 | 0.394 |
| Right rib 7 | Female | -5.29 | -4.873 | -0.417 |
| Right rib 7 | Male | -1.431 | -1.873 | 0.442 |
| Right rib 8 | Female | -5.553 | -5.243 | -0.311 |
| Right rib 8 | Male | -1.548 | -2.064 | 0.516 |
| Right rib 9 | Female | -5.615 | -5.198 | -0.417 |
| Right rib 9 | Male | -1.438 | -2.031 | 0.594 |
| C1 | Female | -4.625 | -4.168 | -0.457 |
| C1 | Male | -0.302 | -1.028 | 0.726 |
| C2 | Female | -5.51 | -5.138 | -0.372 |
| C2 | Male | -1.174 | -1.731 | 0.557 |
| C3 | Female | -4.803 | -4.237 | -0.566 |
| C3 | Male | -0.388 | -1.179 | 0.791 |
| C4 | Female | -4.872 | -4.446 | -0.426 |
| C4 | Male | -0.118 | -0.874 | 0.756 |
| C5 | Female | -4.764 | -4.281 | -0.482 |
| C5 | Male | -0.133 | -0.816 | 0.683 |
| C6 | Female | -4.258 | -3.799 | -0.458 |
| C6 | Male | -0.019 | -0.709 | 0.69 |
| C7 | Female | -3.653 | -3.21 | -0.443 |
| C7 | Male | 0.064 | -0.573 | 0.638 |
| L1 | Female | -3.498 | -3.211 | -0.287 |
| L1 | Male | -0.916 | -1.326 | 0.41 |
| L2 | Female | -3.532 | -3.227 | -0.304 |
| L2 | Male | -0.707 | -1.269 | 0.561 |
| L3 | Female | -3.695 | -3.391 | -0.304 |
| L3 | Male | -0.742 | -1.315 | 0.573 |
| L4 | Female | -3.711 | -3.322 | -0.389 |
| L4 | Male | -0.633 | -1.169 | 0.536 |
| L5 | Female | -3.694 | -3.329 | -0.366 |
| L5 | Male | -0.439 | -1.041 | 0.602 |
| S1 | Female | -4.339 | -4.089 | -0.25 |
| S1 | Male | -1.344 | -1.838 | 0.494 |
| T1 | Female | -3.772 | -3.465 | -0.307 |
| T1 | Male | -0.649 | -1.162 | 0.513 |
| T10 | Female | -3.724 | -3.388 | -0.336 |
| T10 | Male | -0.73 | -1.243 | 0.513 |
| T11 | Female | -3.733 | -3.421 | -0.312 |
| T11 | Male | -0.903 | -1.356 | 0.453 |
| T12 | Female | -3.615 | -3.311 | -0.303 |
| T12 | Male | -1.009 | -1.436 | 0.427 |
| T2 | Female | -3.479 | -3.154 | -0.326 |
| T2 | Male | -0.592 | -1.058 | 0.466 |
| T3 | Female | -3.608 | -3.33 | -0.278 |
| T3 | Male | -0.712 | -1.135 | 0.423 |
| T4 | Female | -3.699 | -3.452 | -0.247 |
| T4 | Male | -0.758 | -1.23 | 0.472 |
| T5 | Female | -3.612 | -3.317 | -0.296 |
| T5 | Male | -0.702 | -1.138 | 0.437 |
| T6 | Female | -3.526 | -3.182 | -0.345 |
| T6 | Male | -0.68 | -1.145 | 0.464 |
| T7 | Female | -3.47 | -3.178 | -0.292 |
| T7 | Male | -0.64 | -1.078 | 0.438 |
| T8 | Female | -3.429 | -3.152 | -0.277 |
| T8 | Male | -0.606 | -1.035 | 0.429 |
| T9 | Female | -3.626 | -3.302 | -0.325 |
| T9 | Male | -0.626 | -1.121 | 0.495 |

Primary rule: adjacency, truncation and a mild robust artifact rule. Legacy rule: 5th–95th percentile trimming of attenuation and volume within each 1-year age stratum. Differences are primary minus legacy.

**Supplementary Table 7B: Summary of the sensitivity analyses**

| **Analysis** | **Value** |
| --- | --- |
| QC rule: \|change\| in fitted HU at 50 (max across bone x sex) | 11.98 |
| QC rule: median \|change\| in fitted HU at 50 | 3.77 |
| QC rule: max \|change\| in 45-60 y decline rate (HU/yr) | 0.791 |
| Center: range of age/sex-adjusted L1 deviation (HU) | 13.3 |
| Center: median range of adjusted deviation across the 49 sites (HU) | 21.6 |
| Center: largest range of adjusted deviation across the 49 sites (HU) | 39 |
| Leave-one-institution-out: max \|delta\| fitted HU at 50 | 6.17 |
| Leave-one-institution-out: max \|delta\| 45-60 y decline rate | 0.765 |
| BMI: median HU per BMI unit | 1.223 |
| BMI: max \|change\| in 45-60 y decline rate after adjustment | 0.233 |
| HU vs volume: median age-adjusted r | -0.013 |

**Supplementary Table 8: Attenuation by contributing center and skeletal site, adjusted for age and sex**

| **Site** | **Region** | **Center** | **n** | **Crude mean attenuation (HU)** | **Mean age (y)** | **Women (%)** | **Deviation from the cohort mean, adjusted (HU)** |
| --- | --- | --- | --- | --- | --- | --- | --- |
| C1 | Cervical vertebrae | Center 1 | 9172 | 428.2 | 64 | 52.3 | -9.9 |
| C1 | Cervical vertebrae | Center 2 | 3698 | 436.5 | 63.6 | 43.5 | -5.3 |
| C1 | Cervical vertebrae | Center 3 | 2877 | 436 | 65.1 | 46.1 | -2.8 |
| C1 | Cervical vertebrae | Center 4 | 1346 | 444.2 | 66.7 | 43.2 | 5.7 |
| C1 | Cervical vertebrae | Center 5 | 2798 | 422.2 | 65.6 | 57.2 | -11.4 |
| C1 | Cervical vertebrae | Center 6 | 2889 | 453.5 | 66.1 | 57.3 | 21.5 |
| C1 | Cervical vertebrae | Center 7 | 2359 | 420.1 | 65.8 | 52.8 | -14.7 |
| C1 | Cervical vertebrae | Center 8 | 1024 | 447.5 | 64.9 | 47.3 | 10.2 |
| C1 | Cervical vertebrae | Center 9 | 132 | 454.1 | 61.9 | 37.1 | 6.6 |
| C2 | Cervical vertebrae | Center 1 | 9113 | 389.9 | 63.9 | 52.5 | -4.8 |
| C2 | Cervical vertebrae | Center 2 | 3715 | 388.5 | 63.6 | 43.5 | -9 |
| C2 | Cervical vertebrae | Center 3 | 2851 | 385.8 | 65.4 | 46.1 | -7.4 |
| C2 | Cervical vertebrae | Center 4 | 1457 | 397.6 | 66.7 | 43.5 | 5.8 |
| C2 | Cervical vertebrae | Center 5 | 2755 | 379.7 | 65.7 | 57.2 | -9.3 |
| C2 | Cervical vertebrae | Center 6 | 2883 | 400.4 | 66.1 | 56.8 | 13.1 |
| C2 | Cervical vertebrae | Center 7 | 2349 | 380.4 | 65.8 | 53.6 | -9.2 |
| C2 | Cervical vertebrae | Center 8 | 1040 | 397.7 | 65.2 | 46.2 | 5.7 |
| C2 | Cervical vertebrae | Center 9 | 125 | 417.7 | 62.5 | 37.6 | 15.1 |
| C3 | Cervical vertebrae | Center 1 | 9189 | 386.2 | 64 | 52.4 | -3.5 |
| C3 | Cervical vertebrae | Center 2 | 4183 | 387.1 | 64.1 | 43.5 | -5.6 |
| C3 | Cervical vertebrae | Center 3 | 3132 | 383.7 | 65.6 | 45.5 | -5.8 |
| C3 | Cervical vertebrae | Center 4 | 2015 | 398.5 | 67 | 49.1 | 11.8 |
| C3 | Cervical vertebrae | Center 5 | 2815 | 377.5 | 65.8 | 57.3 | -7.5 |
| C3 | Cervical vertebrae | Center 6 | 3240 | 400.8 | 66.9 | 55.2 | 17.4 |
| C3 | Cervical vertebrae | Center 7 | 2427 | 377.8 | 66.1 | 52.7 | -8.3 |
| C3 | Cervical vertebrae | Center 8 | 1103 | 391.6 | 65.2 | 46.1 | 3.1 |
| C3 | Cervical vertebrae | Center 9 | 134 | 395.9 | 62.4 | 37.3 | -1.8 |
| C4 | Cervical vertebrae | Center 1 | 9293 | 403.9 | 64 | 52.3 | -6.8 |
| C4 | Cervical vertebrae | Center 2 | 4935 | 409.1 | 64.6 | 43.4 | -4.1 |
| C4 | Cervical vertebrae | Center 3 | 3569 | 408.8 | 66 | 44.3 | -2 |
| C4 | Cervical vertebrae | Center 4 | 2479 | 418.4 | 67.3 | 49.4 | 10.7 |
| C4 | Cervical vertebrae | Center 5 | 2901 | 398.2 | 65.9 | 57 | -8.1 |
| C4 | Cervical vertebrae | Center 6 | 3746 | 429.5 | 67.6 | 52 | 24.6 |
| C4 | Cervical vertebrae | Center 7 | 2661 | 394.5 | 66.2 | 51.3 | -13.3 |
| C4 | Cervical vertebrae | Center 8 | 1148 | 420.4 | 65.5 | 45.8 | 11 |
| C4 | Cervical vertebrae | Center 9 | 140 | 404.5 | 63.4 | 37.9 | -12 |
| C5 | Cervical vertebrae | Center 1 | 9755 | 388.6 | 64.2 | 51.8 | -6.4 |
| C5 | Cervical vertebrae | Center 2 | 6546 | 390.2 | 65.2 | 43.5 | -5.5 |
| C5 | Cervical vertebrae | Center 3 | 4453 | 392.2 | 66.5 | 43 | -1.8 |
| C5 | Cervical vertebrae | Center 4 | 3664 | 401.7 | 68.8 | 47 | 11.7 |
| C5 | Cervical vertebrae | Center 5 | 3296 | 381.6 | 66.2 | 55.9 | -9.2 |
| C5 | Cervical vertebrae | Center 6 | 4533 | 413.1 | 68.3 | 49.3 | 24.1 |
| C5 | Cervical vertebrae | Center 7 | 3061 | 378.4 | 66.4 | 49.9 | -13.6 |
| C5 | Cervical vertebrae | Center 8 | 1444 | 402.3 | 66.4 | 47.2 | 10.3 |
| C5 | Cervical vertebrae | Center 9 | 153 | 388.6 | 64.1 | 38.6 | -9.7 |
| C6 | Cervical vertebrae | Center 1 | 11322 | 362.3 | 64.7 | 49.2 | -3.4 |
| C6 | Cervical vertebrae | Center 2 | 8715 | 363.6 | 65 | 44.9 | -2.4 |
| C6 | Cervical vertebrae | Center 3 | 5915 | 362.2 | 66.7 | 43.8 | -1.9 |
| C6 | Cervical vertebrae | Center 4 | 5896 | 375.1 | 68.9 | 46.4 | 14.3 |
| C6 | Cervical vertebrae | Center 5 | 4614 | 356.1 | 66.7 | 53.2 | -5.9 |
| C6 | Cervical vertebrae | Center 6 | 5443 | 380.6 | 68.2 | 47.7 | 19.8 |
| C6 | Cervical vertebrae | Center 7 | 3509 | 350.7 | 66.6 | 49.2 | -12.1 |
| C6 | Cervical vertebrae | Center 8 | 2081 | 371 | 66.6 | 45.7 | 7.6 |
| C6 | Cervical vertebrae | Center 9 | 182 | 353.3 | 63.9 | 39 | -16 |
| C7 | Cervical vertebrae | Center 1 | 13174 | 322.1 | 65.3 | 47.5 | -1 |
| C7 | Cervical vertebrae | Center 2 | 9359 | 323.2 | 65.6 | 45.7 | 0.1 |
| C7 | Cervical vertebrae | Center 3 | 6808 | 318.2 | 67 | 43.9 | -3.8 |
| C7 | Cervical vertebrae | Center 4 | 6594 | 332.2 | 69.2 | 46.5 | 13.5 |
| C7 | Cervical vertebrae | Center 5 | 5622 | 317.7 | 66.9 | 52.8 | -2.3 |
| C7 | Cervical vertebrae | Center 6 | 5528 | 335.3 | 68.5 | 47.4 | 16.4 |
| C7 | Cervical vertebrae | Center 7 | 3636 | 307.4 | 66.9 | 49.1 | -13.3 |
| C7 | Cervical vertebrae | Center 8 | 2383 | 326 | 66.8 | 45.5 | 4.6 |
| C7 | Cervical vertebrae | Center 9 | 204 | 310.4 | 65.4 | 40.7 | -14.2 |
| T1 | Thoracic vertebrae | Center 1 | 13645 | 281.1 | 65.5 | 47.3 | 0.4 |
| T1 | Thoracic vertebrae | Center 2 | 9383 | 282.9 | 65.7 | 45.8 | 2.4 |
| T1 | Thoracic vertebrae | Center 3 | 6991 | 275 | 67 | 44.2 | -3.9 |
| T1 | Thoracic vertebrae | Center 4 | 6554 | 286.8 | 69.2 | 46.6 | 11.2 |
| T1 | Thoracic vertebrae | Center 5 | 5764 | 279.5 | 66.8 | 52.3 | 1.7 |
| T1 | Thoracic vertebrae | Center 6 | 5460 | 283.6 | 68.6 | 47.9 | 8.1 |
| T1 | Thoracic vertebrae | Center 7 | 3647 | 266.5 | 67 | 49.3 | -11.4 |
| T1 | Thoracic vertebrae | Center 8 | 2388 | 279.4 | 66.8 | 45.6 | 0.9 |
| T1 | Thoracic vertebrae | Center 9 | 208 | 271.9 | 65.9 | 40.4 | -9.4 |
| T2 | Thoracic vertebrae | Center 1 | 13633 | 261.9 | 65.5 | 47.2 | -1.3 |
| T2 | Thoracic vertebrae | Center 2 | 9351 | 263.9 | 65.6 | 46 | 0.7 |
| T2 | Thoracic vertebrae | Center 3 | 7018 | 257.3 | 67 | 44.2 | -4.4 |
| T2 | Thoracic vertebrae | Center 4 | 6562 | 267.2 | 69.2 | 46.6 | 8.7 |
| T2 | Thoracic vertebrae | Center 5 | 5737 | 260.6 | 66.8 | 52.3 | -0 |
| T2 | Thoracic vertebrae | Center 6 | 5458 | 262 | 68.6 | 47.9 | 3.5 |
| T2 | Thoracic vertebrae | Center 7 | 3650 | 252.2 | 67 | 49.1 | -8.6 |
| T2 | Thoracic vertebrae | Center 8 | 2385 | 259.4 | 66.8 | 45.4 | -1.9 |
| T2 | Thoracic vertebrae | Center 9 | 207 | 267.1 | 65.9 | 39.6 | 3.3 |
| T3 | Thoracic vertebrae | Center 1 | 13596 | 260.1 | 65.5 | 47 | -0.2 |
| T3 | Thoracic vertebrae | Center 2 | 9355 | 260.8 | 65.6 | 45.9 | 0.4 |
| T3 | Thoracic vertebrae | Center 3 | 7021 | 255.1 | 66.9 | 44.2 | -3.8 |
| T3 | Thoracic vertebrae | Center 4 | 6558 | 264.4 | 69.2 | 46.6 | 9.2 |
| T3 | Thoracic vertebrae | Center 5 | 5780 | 255.9 | 66.7 | 52.4 | -1.5 |
| T3 | Thoracic vertebrae | Center 6 | 5473 | 258.1 | 68.4 | 48 | 2.7 |
| T3 | Thoracic vertebrae | Center 7 | 3654 | 251.1 | 66.9 | 49.5 | -6.6 |
| T3 | Thoracic vertebrae | Center 8 | 2402 | 255.5 | 66.7 | 45.4 | -3 |
| T3 | Thoracic vertebrae | Center 9 | 212 | 265.3 | 65.1 | 38.7 | 2.8 |
| T4 | Thoracic vertebrae | Center 1 | 13554 | 256.2 | 65.5 | 47 | 0.1 |
| T4 | Thoracic vertebrae | Center 2 | 9349 | 256.4 | 65.7 | 46 | 0.5 |
| T4 | Thoracic vertebrae | Center 3 | 7065 | 251.9 | 66.9 | 44 | -2.8 |
| T4 | Thoracic vertebrae | Center 4 | 6531 | 259.6 | 69.2 | 46.7 | 8.7 |
| T4 | Thoracic vertebrae | Center 5 | 5801 | 251.8 | 66.8 | 52.4 | -1.1 |
| T4 | Thoracic vertebrae | Center 6 | 5497 | 253.2 | 68.4 | 48.1 | 2.1 |
| T4 | Thoracic vertebrae | Center 7 | 3643 | 247 | 66.9 | 49.2 | -6.3 |
| T4 | Thoracic vertebrae | Center 8 | 2402 | 250.3 | 66.7 | 45.6 | -3.9 |
| T4 | Thoracic vertebrae | Center 9 | 212 | 260.6 | 65.4 | 40.1 | 2.6 |
| T5 | Thoracic vertebrae | Center 1 | 13610 | 252.2 | 65.5 | 47.1 | -0.5 |
| T5 | Thoracic vertebrae | Center 2 | 9395 | 252.7 | 65.6 | 45.8 | -0.1 |
| T5 | Thoracic vertebrae | Center 3 | 7095 | 249.3 | 66.9 | 44.2 | -2.2 |
| T5 | Thoracic vertebrae | Center 4 | 6546 | 255.7 | 69.2 | 46.8 | 7.8 |
| T5 | Thoracic vertebrae | Center 5 | 5788 | 249.1 | 66.8 | 52.4 | -0.7 |
| T5 | Thoracic vertebrae | Center 6 | 5462 | 250.3 | 68.4 | 48.2 | 2.3 |
| T5 | Thoracic vertebrae | Center 7 | 3677 | 242.9 | 66.9 | 49.3 | -7.2 |
| T5 | Thoracic vertebrae | Center 8 | 2387 | 246.9 | 66.8 | 45.2 | -4.1 |
| T5 | Thoracic vertebrae | Center 9 | 221 | 260.2 | 65 | 39.4 | 4.8 |
| T6 | Thoracic vertebrae | Center 1 | 13825 | 251.4 | 65.3 | 47 | -0.7 |
| T6 | Thoracic vertebrae | Center 2 | 9780 | 253.1 | 65.3 | 46.1 | 0.7 |
| T6 | Thoracic vertebrae | Center 3 | 7308 | 248.8 | 66.7 | 44.2 | -2.2 |
| T6 | Thoracic vertebrae | Center 4 | 6632 | 255.2 | 69 | 46.7 | 7.9 |
| T6 | Thoracic vertebrae | Center 5 | 5853 | 248.6 | 66.7 | 52.3 | -0.4 |
| T6 | Thoracic vertebrae | Center 6 | 5542 | 250 | 68.3 | 47.9 | 2.5 |
| T6 | Thoracic vertebrae | Center 7 | 3711 | 242.4 | 66.7 | 49.3 | -7.2 |
| T6 | Thoracic vertebrae | Center 8 | 2415 | 246.2 | 66.6 | 45.4 | -4.2 |
| T6 | Thoracic vertebrae | Center 9 | 215 | 257.5 | 65.2 | 40 | 3.5 |
| T7 | Thoracic vertebrae | Center 1 | 14501 | 248 | 65 | 46.7 | -0.9 |
| T7 | Thoracic vertebrae | Center 2 | 10865 | 250.7 | 64.8 | 45.9 | 1.4 |
| T7 | Thoracic vertebrae | Center 3 | 8250 | 246.2 | 66.1 | 44.7 | -1.8 |
| T7 | Thoracic vertebrae | Center 4 | 6878 | 251.6 | 68.8 | 46.7 | 7.4 |
| T7 | Thoracic vertebrae | Center 5 | 6190 | 246.3 | 66.4 | 52.5 | 0.6 |
| T7 | Thoracic vertebrae | Center 6 | 5863 | 247.5 | 68 | 47.7 | 3.1 |
| T7 | Thoracic vertebrae | Center 7 | 3828 | 238.4 | 66.4 | 49.4 | -8 |
| T7 | Thoracic vertebrae | Center 8 | 2546 | 244.1 | 66 | 45.7 | -3.4 |
| T7 | Thoracic vertebrae | Center 9 | 233 | 253.8 | 63.9 | 39.5 | 1.6 |
| T8 | Thoracic vertebrae | Center 1 | 15657 | 245.1 | 64.7 | 46.3 | -0.9 |
| T8 | Thoracic vertebrae | Center 2 | 12303 | 247.5 | 64.5 | 45.8 | 1.1 |
| T8 | Thoracic vertebrae | Center 3 | 9716 | 242.6 | 65.8 | 45.1 | -2.5 |
| T8 | Thoracic vertebrae | Center 4 | 7306 | 248.3 | 68.4 | 47.2 | 6.9 |
| T8 | Thoracic vertebrae | Center 5 | 6666 | 243.1 | 66.4 | 52.6 | 0.7 |
| T8 | Thoracic vertebrae | Center 6 | 6252 | 245.1 | 67.8 | 47.8 | 3.7 |
| T8 | Thoracic vertebrae | Center 7 | 3931 | 234.8 | 66.1 | 49.3 | -8.8 |
| T8 | Thoracic vertebrae | Center 8 | 2702 | 241.2 | 65.9 | 45.4 | -3.2 |
| T8 | Thoracic vertebrae | Center 9 | 248 | 252 | 64 | 38.3 | 3 |
| T9 | Thoracic vertebrae | Center 1 | 17253 | 249.6 | 64.4 | 46.2 | -0.4 |
| T9 | Thoracic vertebrae | Center 2 | 13350 | 250.8 | 64.6 | 46.5 | 1 |
| T9 | Thoracic vertebrae | Center 3 | 11106 | 245.9 | 65.7 | 46.2 | -2.7 |
| T9 | Thoracic vertebrae | Center 4 | 7773 | 253.3 | 68 | 47.7 | 8 |
| T9 | Thoracic vertebrae | Center 5 | 7062 | 247.1 | 66.4 | 52.6 | 1.2 |
| T9 | Thoracic vertebrae | Center 6 | 6573 | 248.5 | 67.8 | 47.6 | 3.7 |
| T9 | Thoracic vertebrae | Center 7 | 4020 | 238.5 | 66 | 49.6 | -8.7 |
| T9 | Thoracic vertebrae | Center 8 | 2868 | 245 | 65.9 | 45.4 | -2.9 |
| T9 | Thoracic vertebrae | Center 9 | 259 | 252.7 | 64.4 | 39.4 | 0.9 |
| T10 | Thoracic vertebrae | Center 1 | 18710 | 247.5 | 64.2 | 46.5 | 0.4 |
| T10 | Thoracic vertebrae | Center 2 | 13617 | 246.7 | 65.1 | 46.7 | 0.8 |
| T10 | Thoracic vertebrae | Center 3 | 11834 | 242.5 | 65.9 | 47.2 | -2.4 |
| T10 | Thoracic vertebrae | Center 4 | 8101 | 250.9 | 67.9 | 48.1 | 8.7 |
| T10 | Thoracic vertebrae | Center 5 | 7240 | 243.8 | 66.7 | 53 | 1.4 |
| T10 | Thoracic vertebrae | Center 6 | 6726 | 244.4 | 67.9 | 47.5 | 3 |
| T10 | Thoracic vertebrae | Center 7 | 4024 | 235.2 | 66 | 49.5 | -8.8 |
| T10 | Thoracic vertebrae | Center 8 | 2965 | 240.8 | 66.3 | 45.3 | -3.4 |
| T10 | Thoracic vertebrae | Center 9 | 257 | 247.7 | 65.1 | 40.5 | 0.3 |
| T11 | Thoracic vertebrae | Center 1 | 19283 | 235.7 | 64 | 46.8 | 0.6 |
| T11 | Thoracic vertebrae | Center 2 | 13557 | 234.6 | 65 | 46.7 | 0.8 |
| T11 | Thoracic vertebrae | Center 3 | 12062 | 230.7 | 65.8 | 47.3 | -2.2 |
| T11 | Thoracic vertebrae | Center 4 | 8165 | 238.6 | 67.8 | 48.2 | 8.4 |
| T11 | Thoracic vertebrae | Center 5 | 7250 | 231.5 | 66.6 | 53.1 | 1.2 |
| T11 | Thoracic vertebrae | Center 6 | 6836 | 232.5 | 67.6 | 47.6 | 2.9 |
| T11 | Thoracic vertebrae | Center 7 | 3993 | 223.9 | 65.9 | 49.8 | -8 |
| T11 | Thoracic vertebrae | Center 8 | 3052 | 228.7 | 66.1 | 45.2 | -3.4 |
| T11 | Thoracic vertebrae | Center 9 | 268 | 234.5 | 65.2 | 39.6 | -0.4 |
| T12 | Thoracic vertebrae | Center 1 | 19228 | 219.3 | 64 | 46.8 | -0.5 |
| T12 | Thoracic vertebrae | Center 2 | 13541 | 219.4 | 64.9 | 46.7 | 0.7 |
| T12 | Thoracic vertebrae | Center 3 | 12050 | 215.9 | 65.8 | 47.3 | -1.8 |
| T12 | Thoracic vertebrae | Center 4 | 8162 | 222 | 67.8 | 48 | 6.6 |
| T12 | Thoracic vertebrae | Center 5 | 7223 | 216.3 | 66.6 | 52.8 | 1 |
| T12 | Thoracic vertebrae | Center 6 | 6885 | 218.6 | 67.5 | 47.5 | 4.2 |
| T12 | Thoracic vertebrae | Center 7 | 3985 | 209.1 | 65.9 | 50.1 | -7.7 |
| T12 | Thoracic vertebrae | Center 8 | 3033 | 213.8 | 66.2 | 45.3 | -2.8 |
| T12 | Thoracic vertebrae | Center 9 | 264 | 219.2 | 65.5 | 39.8 | 0.2 |
| L1 | Lumbar vertebrae | Center 1 | 19197 | 222.1 | 64.1 | 46.9 | -1 |
| L1 | Lumbar vertebrae | Center 2 | 13577 | 223.3 | 65 | 46.6 | 0.9 |
| L1 | Lumbar vertebrae | Center 3 | 12015 | 220.1 | 65.8 | 47.3 | -1.3 |
| L1 | Lumbar vertebrae | Center 4 | 8210 | 225.7 | 67.9 | 48.1 | 6.3 |
| L1 | Lumbar vertebrae | Center 5 | 7220 | 220.3 | 66.7 | 52.9 | 1.2 |
| L1 | Lumbar vertebrae | Center 6 | 6860 | 223.9 | 67.7 | 47.4 | 5.7 |
| L1 | Lumbar vertebrae | Center 7 | 4007 | 213.5 | 65.9 | 49.9 | -7 |
| L1 | Lumbar vertebrae | Center 8 | 3036 | 218.4 | 66.1 | 44.9 | -2.3 |
| L1 | Lumbar vertebrae | Center 9 | 262 | 219.7 | 65.7 | 39.3 | -2.5 |
| L2 | Lumbar vertebrae | Center 1 | 19196 | 235.8 | 64.3 | 46.9 | -0.8 |
| L2 | Lumbar vertebrae | Center 2 | 13587 | 236.6 | 65.2 | 46.7 | 0.5 |
| L2 | Lumbar vertebrae | Center 3 | 12023 | 233.3 | 66 | 47.3 | -1.7 |
| L2 | Lumbar vertebrae | Center 4 | 8230 | 240.4 | 68 | 48.2 | 7.3 |
| L2 | Lumbar vertebrae | Center 5 | 7243 | 233.1 | 66.8 | 52.9 | 0.4 |
| L2 | Lumbar vertebrae | Center 6 | 6848 | 238.8 | 67.9 | 47.5 | 6.8 |
| L2 | Lumbar vertebrae | Center 7 | 4002 | 226.9 | 66 | 49.8 | -7.5 |
| L2 | Lumbar vertebrae | Center 8 | 3020 | 231.6 | 66.4 | 45 | -2.7 |
| L2 | Lumbar vertebrae | Center 9 | 259 | 233.4 | 65.9 | 40.2 | -2.3 |
| L3 | Lumbar vertebrae | Center 1 | 19181 | 241.3 | 64.2 | 46.8 | -0.3 |
| L3 | Lumbar vertebrae | Center 2 | 13553 | 241.6 | 65.2 | 46.6 | 0.7 |
| L3 | Lumbar vertebrae | Center 3 | 12070 | 239 | 65.9 | 47.2 | -0.9 |
| L3 | Lumbar vertebrae | Center 4 | 8201 | 245.5 | 67.9 | 48.2 | 7.8 |
| L3 | Lumbar vertebrae | Center 5 | 7220 | 237.2 | 66.8 | 52.9 | 0.1 |
| L3 | Lumbar vertebrae | Center 6 | 6888 | 243.7 | 67.8 | 47.6 | 7.2 |
| L3 | Lumbar vertebrae | Center 7 | 3993 | 232 | 65.9 | 49.9 | -7 |
| L3 | Lumbar vertebrae | Center 8 | 3022 | 236.9 | 66.4 | 45.2 | -2.1 |
| L3 | Lumbar vertebrae | Center 9 | 261 | 236.5 | 65.2 | 40.2 | -5.4 |
| L4 | Lumbar vertebrae | Center 1 | 19201 | 248.4 | 64.2 | 46.8 | -1 |
| L4 | Lumbar vertebrae | Center 2 | 13595 | 248.9 | 65.2 | 46.6 | 0.2 |
| L4 | Lumbar vertebrae | Center 3 | 12027 | 246.6 | 66 | 47.4 | -1 |
| L4 | Lumbar vertebrae | Center 4 | 8199 | 253.7 | 67.9 | 48.2 | 8 |
| L4 | Lumbar vertebrae | Center 5 | 7233 | 244.5 | 66.8 | 53 | -0.6 |
| L4 | Lumbar vertebrae | Center 6 | 6839 | 252.3 | 67.9 | 47.5 | 7.7 |
| L4 | Lumbar vertebrae | Center 7 | 3986 | 241.1 | 65.9 | 49.7 | -5.8 |
| L4 | Lumbar vertebrae | Center 8 | 3041 | 245 | 66.5 | 45.3 | -2 |
| L4 | Lumbar vertebrae | Center 9 | 266 | 243.4 | 65.6 | 40.6 | -5.7 |
| L5 | Lumbar vertebrae | Center 1 | 19282 | 250.8 | 64.2 | 46.8 | -1.1 |
| L5 | Lumbar vertebrae | Center 2 | 13578 | 251.5 | 65.2 | 46.6 | 0.3 |
| L5 | Lumbar vertebrae | Center 3 | 12031 | 249.1 | 66 | 47.3 | -1 |
| L5 | Lumbar vertebrae | Center 4 | 8221 | 255.9 | 68 | 48.2 | 8 |
| L5 | Lumbar vertebrae | Center 5 | 7200 | 247.1 | 66.8 | 52.8 | -0.4 |
| L5 | Lumbar vertebrae | Center 6 | 6831 | 254.8 | 67.9 | 47.7 | 7.7 |
| L5 | Lumbar vertebrae | Center 7 | 3989 | 243.4 | 66 | 49.6 | -5.8 |
| L5 | Lumbar vertebrae | Center 8 | 3011 | 248.6 | 66.5 | 45.2 | -0.8 |
| L5 | Lumbar vertebrae | Center 9 | 267 | 245.1 | 65.5 | 40.4 | -6.8 |
| S1 | Sacrum | Center 1 | 19226 | 252.3 | 64 | 46.8 | 2.6 |
| S1 | Sacrum | Center 2 | 13608 | 249.7 | 64.9 | 46.5 | 2 |
| S1 | Sacrum | Center 3 | 12031 | 246.7 | 65.8 | 47.3 | 0.4 |
| S1 | Sacrum | Center 4 | 8181 | 249.3 | 67.8 | 48.3 | 7.1 |
| S1 | Sacrum | Center 5 | 7197 | 243 | 66.6 | 52.9 | -0.4 |
| S1 | Sacrum | Center 6 | 6924 | 244.4 | 67.7 | 47.6 | 2.4 |
| S1 | Sacrum | Center 7 | 4001 | 241.5 | 65.8 | 49.7 | -4 |
| S1 | Sacrum | Center 8 | 3037 | 243.5 | 66.1 | 45.8 | -1.9 |
| S1 | Sacrum | Center 9 | 262 | 241.1 | 65.1 | 40.1 | -8.1 |
| Left rib 1 | Left ribs | Center 1 | 13665 | 296.5 | 65.3 | 47.1 | 1.2 |
| Left rib 1 | Left ribs | Center 2 | 9285 | 295.9 | 65.5 | 45.8 | 1.1 |
| Left rib 1 | Left ribs | Center 3 | 6939 | 287.9 | 66.9 | 44.1 | -4.1 |
| Left rib 1 | Left ribs | Center 4 | 6590 | 297.5 | 69 | 46.7 | 10.6 |
| Left rib 1 | Left ribs | Center 5 | 5760 | 281.7 | 66.6 | 52.4 | -9.5 |
| Left rib 1 | Left ribs | Center 6 | 5600 | 299.4 | 68.2 | 48 | 11.7 |
| Left rib 1 | Left ribs | Center 7 | 3647 | 281.9 | 66.8 | 49.6 | -9 |
| Left rib 1 | Left ribs | Center 8 | 2399 | 290.7 | 66.7 | 45.6 | -1 |
| Left rib 1 | Left ribs | Center 9 | 215 | 296.6 | 65 | 40 | -1.1 |
| Left rib 2 | Left ribs | Center 1 | 13541 | 243.6 | 64.9 | 47.2 | 3.8 |
| Left rib 2 | Left ribs | Center 2 | 9328 | 242.8 | 64.7 | 45.7 | 1.3 |
| Left rib 2 | Left ribs | Center 3 | 7002 | 232 | 66.4 | 44.2 | -6.3 |
| Left rib 2 | Left ribs | Center 4 | 6585 | 246 | 68.7 | 46.8 | 11.9 |
| Left rib 2 | Left ribs | Center 5 | 5726 | 230.7 | 66.2 | 52.3 | -6.4 |
| Left rib 2 | Left ribs | Center 6 | 5713 | 240.3 | 67.9 | 48.1 | 6.4 |
| Left rib 2 | Left ribs | Center 7 | 3666 | 227.9 | 66.5 | 49.2 | -9.1 |
| Left rib 2 | Left ribs | Center 8 | 2413 | 234.7 | 66.1 | 45.3 | -3.1 |
| Left rib 2 | Left ribs | Center 9 | 214 | 243.4 | 64.1 | 38.8 | 1.4 |
| Left rib 3 | Left ribs | Center 1 | 13882 | 233.8 | 65 | 47.4 | 4.2 |
| Left rib 3 | Left ribs | Center 2 | 9652 | 231.4 | 64.7 | 46.1 | -0.5 |
| Left rib 3 | Left ribs | Center 3 | 7207 | 222.6 | 66.4 | 44.6 | -5.1 |
| Left rib 3 | Left ribs | Center 4 | 6723 | 235.2 | 68.7 | 46.8 | 12.1 |
| Left rib 3 | Left ribs | Center 5 | 5916 | 221 | 66.3 | 52.7 | -6 |
| Left rib 3 | Left ribs | Center 6 | 5736 | 228.5 | 67.9 | 48.2 | 5.7 |
| Left rib 3 | Left ribs | Center 7 | 3735 | 218.3 | 66.4 | 49.6 | -8.7 |
| Left rib 3 | Left ribs | Center 8 | 2465 | 222.5 | 66.1 | 45.8 | -4.9 |
| Left rib 3 | Left ribs | Center 9 | 217 | 235 | 64.1 | 38.7 | 3.3 |
| Left rib 4 | Left ribs | Center 1 | 14651 | 239.9 | 65 | 47.4 | 3.8 |
| Left rib 4 | Left ribs | Center 2 | 11739 | 234.9 | 64.4 | 47.2 | -4.3 |
| Left rib 4 | Left ribs | Center 3 | 9200 | 227.9 | 65.8 | 46.8 | -8.1 |
| Left rib 4 | Left ribs | Center 4 | 7028 | 243.2 | 68.7 | 47.1 | 13 |
| Left rib 4 | Left ribs | Center 5 | 6296 | 228.4 | 66.5 | 53.1 | -4.8 |
| Left rib 4 | Left ribs | Center 6 | 6003 | 239.6 | 67.9 | 48.3 | 10 |
| Left rib 4 | Left ribs | Center 7 | 3879 | 226.8 | 66.2 | 49.5 | -7.1 |
| Left rib 4 | Left ribs | Center 8 | 2603 | 228.1 | 66 | 45.8 | -6.3 |
| Left rib 4 | Left ribs | Center 9 | 237 | 242.5 | 64.1 | 38.4 | 3.9 |
| Left rib 5 | Left ribs | Center 1 | 16824 | 239.8 | 64.4 | 47.3 | 1.4 |
| Left rib 5 | Left ribs | Center 2 | 13658 | 234 | 64.4 | 46.7 | -5.9 |
| Left rib 5 | Left ribs | Center 3 | 11572 | 225.9 | 65.4 | 47.5 | -11.8 |
| Left rib 5 | Left ribs | Center 4 | 7735 | 243.1 | 68 | 48 | 9.6 |
| Left rib 5 | Left ribs | Center 5 | 6995 | 230.7 | 66.4 | 53.1 | -4 |
| Left rib 5 | Left ribs | Center 6 | 6538 | 246.2 | 67.7 | 48.3 | 14.5 |
| Left rib 5 | Left ribs | Center 7 | 4020 | 231.3 | 65.9 | 49.6 | -4.5 |
| Left rib 5 | Left ribs | Center 8 | 2857 | 231.9 | 65.8 | 46 | -3.9 |
| Left rib 5 | Left ribs | Center 9 | 262 | 244 | 64.4 | 39.7 | 4.6 |
| Left rib 6 | Left ribs | Center 1 | 18510 | 248.5 | 64 | 46.9 | -3.9 |
| Left rib 6 | Left ribs | Center 2 | 13982 | 247.6 | 64.4 | 46.6 | -5 |
| Left rib 6 | Left ribs | Center 3 | 12091 | 238.3 | 65.5 | 47.4 | -12.3 |
| Left rib 6 | Left ribs | Center 4 | 8125 | 253.6 | 67.8 | 48.2 | 6.6 |
| Left rib 6 | Left ribs | Center 5 | 7328 | 242.7 | 66.4 | 53 | -5.4 |
| Left rib 6 | Left ribs | Center 6 | 6859 | 264.5 | 67.5 | 47.8 | 19.2 |
| Left rib 6 | Left ribs | Center 7 | 4098 | 246 | 65.7 | 49.9 | -3.5 |
| Left rib 6 | Left ribs | Center 8 | 2998 | 246.4 | 65.8 | 45.5 | -2.4 |
| Left rib 6 | Left ribs | Center 9 | 272 | 259.6 | 64.1 | 40.1 | 6.7 |
| Left rib 7 | Left ribs | Center 1 | 18708 | 259.6 | 63.9 | 46.9 | -4.1 |
| Left rib 7 | Left ribs | Center 2 | 13943 | 261.1 | 64.4 | 46.5 | -2.6 |
| Left rib 7 | Left ribs | Center 3 | 12001 | 253.7 | 65.5 | 47.2 | -7.7 |
| Left rib 7 | Left ribs | Center 4 | 8182 | 263.6 | 67.7 | 48.2 | 5.8 |
| Left rib 7 | Left ribs | Center 5 | 7373 | 253.1 | 66.4 | 53 | -6 |
| Left rib 7 | Left ribs | Center 6 | 6870 | 276.6 | 67.5 | 47.8 | 21 |
| Left rib 7 | Left ribs | Center 7 | 4112 | 252.3 | 65.7 | 49.8 | -8.1 |
| Left rib 7 | Left ribs | Center 8 | 3007 | 256.7 | 65.8 | 45.5 | -2.7 |
| Left rib 7 | Left ribs | Center 9 | 271 | 267.5 | 64.2 | 39.5 | 4.4 |
| Left rib 8 | Left ribs | Center 1 | 18757 | 272.4 | 63.9 | 46.9 | -3.8 |
| Left rib 8 | Left ribs | Center 2 | 13927 | 274.9 | 64.4 | 46.5 | -1.4 |
| Left rib 8 | Left ribs | Center 3 | 11960 | 270.3 | 65.5 | 47.3 | -3 |
| Left rib 8 | Left ribs | Center 4 | 8204 | 273.8 | 67.7 | 48.2 | 4.7 |
| Left rib 8 | Left ribs | Center 5 | 7374 | 264.7 | 66.3 | 52.9 | -5.9 |
| Left rib 8 | Left ribs | Center 6 | 6875 | 287.1 | 67.5 | 47.8 | 20.4 |
| Left rib 8 | Left ribs | Center 7 | 4109 | 262.5 | 65.7 | 49.7 | -9.7 |
| Left rib 8 | Left ribs | Center 8 | 3026 | 266.1 | 65.8 | 45.7 | -5.2 |
| Left rib 8 | Left ribs | Center 9 | 269 | 279.8 | 64.2 | 39.4 | 3.9 |
| Left rib 9 | Left ribs | Center 1 | 18905 | 287.9 | 63.9 | 46.8 | -3 |
| Left rib 9 | Left ribs | Center 2 | 13842 | 288.1 | 64.5 | 46.6 | -2.4 |
| Left rib 9 | Left ribs | Center 3 | 11932 | 285.6 | 65.5 | 47.3 | -2.1 |
| Left rib 9 | Left ribs | Center 4 | 8233 | 287.4 | 67.7 | 48.3 | 4.1 |
| Left rib 9 | Left ribs | Center 5 | 7364 | 277.6 | 66.4 | 53.1 | -6.7 |
| Left rib 9 | Left ribs | Center 6 | 6862 | 303.9 | 67.6 | 47.7 | 22.8 |
| Left rib 9 | Left ribs | Center 7 | 4115 | 274.5 | 65.7 | 49.6 | -12.1 |
| Left rib 9 | Left ribs | Center 8 | 3035 | 281.2 | 65.9 | 45.5 | -4.9 |
| Left rib 9 | Left ribs | Center 9 | 268 | 295.5 | 64.2 | 39.9 | 4.4 |
| Left rib 10 | Left ribs | Center 1 | 19339 | 317.3 | 63.8 | 46.7 | -2 |
| Left rib 10 | Left ribs | Center 2 | 13672 | 316.3 | 64.6 | 46.8 | -1.9 |
| Left rib 10 | Left ribs | Center 3 | 12029 | 313.8 | 65.6 | 47.3 | -1.9 |
| Left rib 10 | Left ribs | Center 4 | 8216 | 316.5 | 67.6 | 48.5 | 4.6 |
| Left rib 10 | Left ribs | Center 5 | 7311 | 304.9 | 66.5 | 52.9 | -7.6 |
| Left rib 10 | Left ribs | Center 6 | 6738 | 331.8 | 67.5 | 47.8 | 22.7 |
| Left rib 10 | Left ribs | Center 7 | 4058 | 303.3 | 65.7 | 49.7 | -11.5 |
| Left rib 10 | Left ribs | Center 8 | 3006 | 306.9 | 66 | 45 | -6.7 |
| Left rib 10 | Left ribs | Center 9 | 266 | 324.1 | 64.1 | 38.3 | 4.3 |
| Left rib 11 | Left ribs | Center 1 | 19341 | 323.6 | 63.8 | 46.7 | 0.6 |
| Left rib 11 | Left ribs | Center 2 | 13597 | 321.4 | 64.6 | 46.5 | -0.2 |
| Left rib 11 | Left ribs | Center 3 | 12070 | 318.4 | 65.5 | 47.5 | -0.3 |
| Left rib 11 | Left ribs | Center 4 | 8231 | 320.3 | 67.5 | 48.5 | 6.3 |
| Left rib 11 | Left ribs | Center 5 | 7241 | 309.3 | 66.4 | 53 | -5.5 |
| Left rib 11 | Left ribs | Center 6 | 6814 | 329.3 | 67.4 | 48 | 17.8 |
| Left rib 11 | Left ribs | Center 7 | 4028 | 304 | 65.7 | 49.8 | -13.3 |
| Left rib 11 | Left ribs | Center 8 | 3062 | 309.4 | 65.9 | 45.2 | -7.3 |
| Left rib 11 | Left ribs | Center 9 | 261 | 324.3 | 64.5 | 37.9 | 1.9 |
| Left rib 12 | Left ribs | Center 1 | 18734 | 316.2 | 63.7 | 46.3 | 0.7 |
| Left rib 12 | Left ribs | Center 2 | 13443 | 318.8 | 64.4 | 46 | 4 |
| Left rib 12 | Left ribs | Center 3 | 11807 | 314.1 | 65.4 | 46.7 | 2.1 |
| Left rib 12 | Left ribs | Center 4 | 8041 | 316.1 | 67.5 | 47.7 | 8.4 |
| Left rib 12 | Left ribs | Center 5 | 7064 | 304 | 66.4 | 52.4 | -4.6 |
| Left rib 12 | Left ribs | Center 6 | 6748 | 319.4 | 67.3 | 47.3 | 14.1 |
| Left rib 12 | Left ribs | Center 7 | 3910 | 299.2 | 65.6 | 49.1 | -11.4 |
| Left rib 12 | Left ribs | Center 8 | 3003 | 302.2 | 65.8 | 44.8 | -7.1 |
| Left rib 12 | Left ribs | Center 9 | 257 | 306.6 | 64.7 | 38.9 | -6.3 |
| Right rib 1 | Right ribs | Center 1 | 13593 | 297.4 | 65.3 | 46.9 | 3.3 |
| Right rib 1 | Right ribs | Center 2 | 9305 | 294.8 | 65.5 | 46 | 1 |
| Right rib 1 | Right ribs | Center 3 | 6944 | 286.8 | 67 | 44 | -3.9 |
| Right rib 1 | Right ribs | Center 4 | 6546 | 296.9 | 69.1 | 46.8 | 11.3 |
| Right rib 1 | Right ribs | Center 5 | 5785 | 281.5 | 66.7 | 52.3 | -8.4 |
| Right rib 1 | Right ribs | Center 6 | 5635 | 297.4 | 68.2 | 48 | 10.9 |
| Right rib 1 | Right ribs | Center 7 | 3630 | 281.2 | 66.8 | 49.6 | -8.7 |
| Right rib 1 | Right ribs | Center 8 | 2407 | 288.8 | 66.8 | 45.9 | -1.5 |
| Right rib 1 | Right ribs | Center 9 | 213 | 293.9 | 64.6 | 39.9 | -4 |
| Right rib 2 | Right ribs | Center 1 | 13503 | 244.7 | 64.9 | 47.1 | 3.8 |
| Right rib 2 | Right ribs | Center 2 | 9344 | 243.7 | 64.7 | 45.8 | 0.9 |
| Right rib 2 | Right ribs | Center 3 | 7011 | 233 | 66.4 | 44.3 | -6.3 |
| Right rib 2 | Right ribs | Center 4 | 6569 | 246.9 | 68.7 | 46.7 | 11.7 |
| Right rib 2 | Right ribs | Center 5 | 5788 | 232.1 | 66.3 | 52.3 | -6.1 |
| Right rib 2 | Right ribs | Center 6 | 5710 | 241.4 | 68 | 48.2 | 6.5 |
| Right rib 2 | Right ribs | Center 7 | 3640 | 229.8 | 66.4 | 49.3 | -8.5 |
| Right rib 2 | Right ribs | Center 8 | 2438 | 235.9 | 66.2 | 45.8 | -2.9 |
| Right rib 2 | Right ribs | Center 9 | 210 | 243.8 | 64.3 | 39.5 | 0.9 |
| Right rib 3 | Right ribs | Center 1 | 13879 | 231.3 | 64.9 | 47.3 | 3.2 |
| Right rib 3 | Right ribs | Center 2 | 9609 | 229.8 | 64.7 | 46.2 | -0.7 |
| Right rib 3 | Right ribs | Center 3 | 7196 | 221.1 | 66.4 | 44.5 | -5.1 |
| Right rib 3 | Right ribs | Center 4 | 6723 | 232.8 | 68.8 | 46.9 | 11.2 |
| Right rib 3 | Right ribs | Center 5 | 5924 | 220.6 | 66.4 | 52.7 | -5 |
| Right rib 3 | Right ribs | Center 6 | 5716 | 226.7 | 67.9 | 48.3 | 5.3 |
| Right rib 3 | Right ribs | Center 7 | 3731 | 216.6 | 66.4 | 49.6 | -8.9 |
| Right rib 3 | Right ribs | Center 8 | 2465 | 221.3 | 66.1 | 45.8 | -4.7 |
| Right rib 3 | Right ribs | Center 9 | 218 | 234.5 | 64.3 | 38.5 | 4.7 |
| Right rib 4 | Right ribs | Center 1 | 14587 | 241.3 | 65 | 47.4 | 3.5 |
| Right rib 4 | Right ribs | Center 2 | 11695 | 235.7 | 64.4 | 47.2 | -5.5 |
| Right rib 4 | Right ribs | Center 3 | 9042 | 228.7 | 65.8 | 46.7 | -9.1 |
| Right rib 4 | Right ribs | Center 4 | 7021 | 244.8 | 68.6 | 47 | 12.5 |
| Right rib 4 | Right ribs | Center 5 | 6270 | 232.7 | 66.5 | 53 | -2.6 |
| Right rib 4 | Right ribs | Center 6 | 5987 | 242.4 | 67.9 | 48.3 | 10.8 |
| Right rib 4 | Right ribs | Center 7 | 3865 | 229 | 66.3 | 49.5 | -6.7 |
| Right rib 4 | Right ribs | Center 8 | 2601 | 231.1 | 65.9 | 45.9 | -5.1 |
| Right rib 4 | Right ribs | Center 9 | 239 | 242.1 | 64 | 39.3 | 2.1 |
| Right rib 5 | Right ribs | Center 1 | 16624 | 242.2 | 64.5 | 47.3 | 1.1 |
| Right rib 5 | Right ribs | Center 2 | 13614 | 234.7 | 64.3 | 46.8 | -7.9 |
| Right rib 5 | Right ribs | Center 3 | 11472 | 225.4 | 65.5 | 47.5 | -15 |
| Right rib 5 | Right ribs | Center 4 | 7694 | 247.2 | 68 | 48 | 10.5 |
| Right rib 5 | Right ribs | Center 5 | 6968 | 235.9 | 66.4 | 53.1 | -2.1 |
| Right rib 5 | Right ribs | Center 6 | 6503 | 251.1 | 67.7 | 48.3 | 16.2 |
| Right rib 5 | Right ribs | Center 7 | 4004 | 235.3 | 65.9 | 49.6 | -3.3 |
| Right rib 5 | Right ribs | Center 8 | 2847 | 236.3 | 65.8 | 46 | -2.5 |
| Right rib 5 | Right ribs | Center 9 | 260 | 245.3 | 64 | 38.8 | 2.9 |
| Right rib 6 | Right ribs | Center 1 | 18466 | 252.1 | 64 | 46.9 | -2.8 |
| Right rib 6 | Right ribs | Center 2 | 14016 | 249.8 | 64.4 | 46.5 | -5.4 |
| Right rib 6 | Right ribs | Center 3 | 12114 | 239.5 | 65.5 | 47.4 | -13.5 |
| Right rib 6 | Right ribs | Center 4 | 8112 | 258.4 | 67.7 | 48.1 | 8.8 |
| Right rib 6 | Right ribs | Center 5 | 7318 | 247.8 | 66.4 | 53 | -3 |
| Right rib 6 | Right ribs | Center 6 | 6839 | 266.2 | 67.4 | 47.9 | 18.4 |
| Right rib 6 | Right ribs | Center 7 | 4086 | 247.3 | 65.7 | 49.7 | -4.6 |
| Right rib 6 | Right ribs | Center 8 | 3003 | 248.9 | 65.8 | 45.5 | -2.4 |
| Right rib 6 | Right ribs | Center 9 | 269 | 259.3 | 64.3 | 40.5 | 4.4 |
| Right rib 7 | Right ribs | Center 1 | 18692 | 263.2 | 63.9 | 46.9 | -3.6 |
| Right rib 7 | Right ribs | Center 2 | 13956 | 263.9 | 64.4 | 46.5 | -3 |
| Right rib 7 | Right ribs | Center 3 | 12014 | 256.8 | 65.4 | 47.3 | -7.8 |
| Right rib 7 | Right ribs | Center 4 | 8192 | 265.9 | 67.7 | 48.3 | 5 |
| Right rib 7 | Right ribs | Center 5 | 7362 | 258.2 | 66.4 | 53 | -4.2 |
| Right rib 7 | Right ribs | Center 6 | 6859 | 279 | 67.5 | 47.8 | 20.2 |
| Right rib 7 | Right ribs | Center 7 | 4099 | 255.9 | 65.7 | 49.8 | -7.8 |
| Right rib 7 | Right ribs | Center 8 | 3010 | 259 | 65.8 | 45.4 | -3.6 |
| Right rib 7 | Right ribs | Center 9 | 272 | 270.9 | 64.3 | 39.7 | 4.7 |
| Right rib 8 | Right ribs | Center 1 | 18751 | 275 | 64 | 46.8 | -3.4 |
| Right rib 8 | Right ribs | Center 2 | 13927 | 276.7 | 64.4 | 46.5 | -1.8 |
| Right rib 8 | Right ribs | Center 3 | 11915 | 272.9 | 65.5 | 47.2 | -2.9 |
| Right rib 8 | Right ribs | Center 4 | 8224 | 275.6 | 67.6 | 48.3 | 3.7 |
| Right rib 8 | Right ribs | Center 5 | 7378 | 268.8 | 66.4 | 52.9 | -4.6 |
| Right rib 8 | Right ribs | Center 6 | 6862 | 291.3 | 67.5 | 47.8 | 21.7 |
| Right rib 8 | Right ribs | Center 7 | 4113 | 263.6 | 65.7 | 49.7 | -11.2 |
| Right rib 8 | Right ribs | Center 8 | 3025 | 268.5 | 65.8 | 45.5 | -5.4 |
| Right rib 8 | Right ribs | Center 9 | 273 | 281.9 | 64.1 | 39.6 | 3.9 |
| Right rib 9 | Right ribs | Center 1 | 18880 | 290.1 | 63.9 | 46.8 | -2.4 |
| Right rib 9 | Right ribs | Center 2 | 13877 | 290.1 | 64.5 | 46.5 | -2.3 |
| Right rib 9 | Right ribs | Center 3 | 11918 | 287.6 | 65.6 | 47.2 | -2.1 |
| Right rib 9 | Right ribs | Center 4 | 8236 | 289.5 | 67.7 | 48.3 | 3.5 |
| Right rib 9 | Right ribs | Center 5 | 7369 | 282.9 | 66.4 | 53 | -4.3 |
| Right rib 9 | Right ribs | Center 6 | 6856 | 307.6 | 67.5 | 47.8 | 23.8 |
| Right rib 9 | Right ribs | Center 7 | 4104 | 276.1 | 65.7 | 49.7 | -12.7 |
| Right rib 9 | Right ribs | Center 8 | 3025 | 282.6 | 65.9 | 45.5 | -5.6 |
| Right rib 9 | Right ribs | Center 9 | 269 | 294.8 | 64.1 | 39 | 2 |
| Right rib 10 | Right ribs | Center 1 | 19257 | 321.9 | 63.9 | 46.6 | -2 |
| Right rib 10 | Right ribs | Center 2 | 13663 | 321.1 | 64.7 | 46.7 | -1.9 |
| Right rib 10 | Right ribs | Center 3 | 12019 | 319.8 | 65.6 | 47.4 | -1 |
| Right rib 10 | Right ribs | Center 4 | 8246 | 321.6 | 67.7 | 48.4 | 4.3 |
| Right rib 10 | Right ribs | Center 5 | 7335 | 312.4 | 66.5 | 53 | -5.7 |
| Right rib 10 | Right ribs | Center 6 | 6742 | 339.6 | 67.6 | 47.9 | 24.9 |
| Right rib 10 | Right ribs | Center 7 | 4063 | 305.8 | 65.8 | 49.5 | -14.1 |
| Right rib 10 | Right ribs | Center 8 | 3027 | 312.1 | 65.9 | 45.2 | -6.7 |
| Right rib 10 | Right ribs | Center 9 | 267 | 325.2 | 64.6 | 38.6 | 2.3 |
| Right rib 11 | Right ribs | Center 1 | 19361 | 326.5 | 63.8 | 46.7 | -1 |
| Right rib 11 | Right ribs | Center 2 | 13600 | 325.7 | 64.5 | 46.5 | -0.8 |
| Right rib 11 | Right ribs | Center 3 | 12042 | 323.1 | 65.5 | 47.5 | -0.5 |
| Right rib 11 | Right ribs | Center 4 | 8255 | 325.6 | 67.5 | 48.4 | 6.2 |
| Right rib 11 | Right ribs | Center 5 | 7245 | 313.3 | 66.4 | 52.9 | -6.4 |
| Right rib 11 | Right ribs | Center 6 | 6817 | 337.3 | 67.4 | 48 | 20.6 |
| Right rib 11 | Right ribs | Center 7 | 4040 | 306.9 | 65.6 | 49.8 | -15.6 |
| Right rib 11 | Right ribs | Center 8 | 3058 | 314.6 | 65.8 | 45.2 | -7.3 |
| Right rib 11 | Right ribs | Center 9 | 263 | 331.6 | 64.4 | 39.2 | 4.8 |
| Right rib 12 | Right ribs | Center 1 | 18911 | 316.9 | 63.6 | 46.6 | -0.1 |
| Right rib 12 | Right ribs | Center 2 | 13528 | 320.2 | 64.4 | 46.2 | 3.9 |
| Right rib 12 | Right ribs | Center 3 | 11884 | 316.9 | 65.3 | 47.1 | 2.9 |
| Right rib 12 | Right ribs | Center 4 | 8075 | 318.5 | 67.5 | 48.2 | 8.3 |
| Right rib 12 | Right ribs | Center 5 | 7147 | 305 | 66.3 | 52.9 | -5.6 |
| Right rib 12 | Right ribs | Center 6 | 6770 | 324.5 | 67.3 | 47.3 | 17.1 |
| Right rib 12 | Right ribs | Center 7 | 3933 | 298.4 | 65.6 | 49.7 | -14.1 |
| Right rib 12 | Right ribs | Center 8 | 3021 | 303.7 | 65.9 | 45 | -7.3 |
| Right rib 12 | Right ribs | Center 9 | 266 | 310 | 64.4 | 38.7 | -5.1 |

Centers are pseudonymized and ordered by decreasing number of patients. Deviations come from a model with a spline in age, a term for sex and center indicators, and are centered to sum to zero, so they are not confounded by differences in case mix. Their spread per site is summarized in Supplementary Table 14.

**Supplementary Table 9: Leave-one-center-out refits**

| **Site** | **Sex** | **Center omitted** | **n after omission** | **Attenuation at 50 y, omitting center (HU)** | **Attenuation at 50 y, all centers (HU)** | **Difference at 50 y (HU)** | **Change 45–60 y, omitting center (HU/y)** | **Change 45–60 y, all centers (HU/y)** | **Difference in change 45–60 y (HU/y)** |
| --- | --- | --- | --- | --- | --- | --- | --- | --- | --- |
| C1 | Male | Center 1 | 8570 | 450.5 | 447.4 | 3.13 | -0.274 | -0.302 | 0.028 |
| C1 | Male | Center 2 | 10853 | 448 | 447.4 | 0.62 | -0.327 | -0.302 | -0.025 |
| C1 | Male | Center 3 | 11389 | 447.2 | 447.4 | -0.14 | -0.239 | -0.302 | 0.063 |
| C1 | Male | Center 4 | 12177 | 446.9 | 447.4 | -0.43 | -0.302 | -0.302 | -0 |
| C1 | Male | Center 5 | 11744 | 447.6 | 447.4 | 0.27 | -0.24 | -0.302 | 0.062 |
| C1 | Male | Center 6 | 11708 | 445.2 | 447.4 | -2.14 | -0.416 | -0.302 | -0.115 |
| C1 | Male | Center 7 | 11828 | 448.2 | 447.4 | 0.87 | -0.327 | -0.302 | -0.025 |
| C1 | Male | Center 8 | 12401 | 446.3 | 447.4 | -1.02 | -0.302 | -0.302 | 0 |
| C1 | Male | Center 9 | 12858 | 447.5 | 447.4 | 0.11 | -0.307 | -0.302 | -0.005 |
| C1 | Female | Center 1 | 8553 | 471.8 | 470 | 1.83 | -4.532 | -4.625 | 0.093 |
| C1 | Female | Center 2 | 11744 | 470.3 | 470 | 0.35 | -4.619 | -4.625 | 0.006 |
| C1 | Female | Center 3 | 12029 | 467.4 | 470 | -2.54 | -4.485 | -4.625 | 0.14 |
| C1 | Female | Center 4 | 12772 | 469.9 | 470 | -0.1 | -4.643 | -4.625 | -0.018 |
| C1 | Female | Center 5 | 11753 | 471.1 | 470 | 1.18 | -4.605 | -4.625 | 0.019 |
| C1 | Female | Center 6 | 11698 | 468.1 | 470 | -1.88 | -4.865 | -4.625 | -0.24 |
| C1 | Female | Center 7 | 12108 | 472.5 | 470 | 2.51 | -4.725 | -4.625 | -0.101 |
| C1 | Female | Center 8 | 12870 | 469.5 | 470 | -0.48 | -4.568 | -4.625 | 0.057 |
| C1 | Female | Center 9 | 13305 | 469.8 | 470 | -0.22 | -4.611 | -4.625 | 0.014 |
| C2 | Male | Center 1 | 8610 | 414.5 | 414.5 | 0.08 | -1.195 | -1.174 | -0.022 |
| C2 | Male | Center 2 | 10840 | 416.4 | 414.5 | 1.97 | -1.247 | -1.174 | -0.074 |
| C2 | Male | Center 3 | 11402 | 414.8 | 414.5 | 0.38 | -1.062 | -1.174 | 0.112 |
| C2 | Male | Center 4 | 12116 | 414.1 | 414.5 | -0.32 | -1.209 | -1.174 | -0.035 |
| C2 | Male | Center 5 | 11761 | 414.5 | 414.5 | 0.06 | -1.092 | -1.174 | 0.082 |
| C2 | Male | Center 6 | 11693 | 412.7 | 414.5 | -1.73 | -1.219 | -1.174 | -0.046 |
| C2 | Male | Center 7 | 11850 | 415 | 414.5 | 0.55 | -1.211 | -1.174 | -0.037 |
| C2 | Male | Center 8 | 12379 | 413.8 | 414.5 | -0.66 | -1.171 | -1.174 | 0.002 |
| C2 | Male | Center 9 | 12861 | 414.5 | 414.5 | 0.03 | -1.181 | -1.174 | -0.007 |
| C2 | Female | Center 1 | 8565 | 442.1 | 442.3 | -0.25 | -5.649 | -5.51 | -0.139 |
| C2 | Female | Center 2 | 11733 | 443.1 | 442.3 | 0.77 | -5.501 | -5.51 | 0.009 |
| C2 | Female | Center 3 | 12035 | 440.4 | 442.3 | -1.93 | -5.393 | -5.51 | 0.117 |
| C2 | Female | Center 4 | 12715 | 442.3 | 442.3 | -0 | -5.527 | -5.51 | -0.017 |
| C2 | Female | Center 5 | 11772 | 443.4 | 442.3 | 1.03 | -5.508 | -5.51 | 0.002 |
| C2 | Female | Center 6 | 11712 | 441 | 442.3 | -1.33 | -5.604 | -5.51 | -0.094 |
| C2 | Female | Center 7 | 12089 | 445.1 | 442.3 | 2.73 | -5.658 | -5.51 | -0.148 |
| C2 | Female | Center 8 | 12869 | 442 | 442.3 | -0.36 | -5.489 | -5.51 | 0.021 |
| C2 | Female | Center 9 | 13302 | 442 | 442.3 | -0.34 | -5.49 | -5.51 | 0.02 |
| C3 | Male | Center 1 | 9575 | 402.7 | 401.5 | 1.22 | -0.425 | -0.388 | -0.038 |
| C3 | Male | Center 2 | 11582 | 402.7 | 401.5 | 1.23 | -0.435 | -0.388 | -0.047 |
| C3 | Male | Center 3 | 12239 | 401.7 | 401.5 | 0.25 | -0.269 | -0.388 | 0.118 |
| C3 | Male | Center 4 | 12920 | 400.6 | 401.5 | -0.9 | -0.377 | -0.388 | 0.011 |
| C3 | Male | Center 5 | 12742 | 401.8 | 401.5 | 0.36 | -0.327 | -0.388 | 0.061 |
| C3 | Male | Center 6 | 12495 | 399.8 | 401.5 | -1.65 | -0.5 | -0.388 | -0.112 |
| C3 | Male | Center 7 | 12796 | 401.8 | 401.5 | 0.32 | -0.412 | -0.388 | -0.024 |
| C3 | Male | Center 8 | 13350 | 401 | 401.5 | -0.44 | -0.376 | -0.388 | 0.012 |
| C3 | Male | Center 9 | 13861 | 401.7 | 401.5 | 0.26 | -0.399 | -0.388 | -0.011 |
| C3 | Female | Center 1 | 9474 | 427.3 | 428.7 | -1.41 | -4.636 | -4.803 | 0.167 |
| C3 | Female | Center 2 | 12473 | 429.1 | 428.7 | 0.43 | -4.76 | -4.803 | 0.043 |
| C3 | Female | Center 3 | 12867 | 427.9 | 428.7 | -0.77 | -4.714 | -4.803 | 0.089 |
| C3 | Female | Center 4 | 13303 | 427.9 | 428.7 | -0.77 | -4.838 | -4.803 | -0.035 |
| C3 | Female | Center 5 | 12681 | 429.9 | 428.7 | 1.27 | -4.825 | -4.803 | -0.022 |
| C3 | Female | Center 6 | 12503 | 427.5 | 428.7 | -1.16 | -4.866 | -4.803 | -0.063 |
| C3 | Female | Center 7 | 13015 | 430.5 | 428.7 | 1.85 | -4.873 | -4.803 | -0.07 |
| C3 | Female | Center 8 | 13785 | 428.6 | 428.7 | -0.06 | -4.785 | -4.803 | 0.018 |
| C3 | Female | Center 9 | 14243 | 428.5 | 428.7 | -0.19 | -4.796 | -4.803 | 0.007 |
| C4 | Male | Center 1 | 11086 | 420.5 | 416.7 | 3.74 | -0.148 | -0.118 | -0.03 |
| C4 | Male | Center 2 | 12722 | 417.4 | 416.7 | 0.63 | -0.16 | -0.118 | -0.042 |
| C4 | Male | Center 3 | 13530 | 416.4 | 416.7 | -0.34 | -0.043 | -0.118 | 0.075 |
| C4 | Male | Center 4 | 14262 | 415.5 | 416.7 | -1.2 | -0.095 | -0.118 | 0.023 |
| C4 | Male | Center 5 | 14271 | 417.4 | 416.7 | 0.64 | -0.067 | -0.118 | 0.051 |
| C4 | Male | Center 6 | 13719 | 414.5 | 416.7 | -2.29 | -0.226 | -0.118 | -0.108 |
| C4 | Male | Center 7 | 14221 | 417.4 | 416.7 | 0.68 | -0.122 | -0.118 | -0.004 |
| C4 | Male | Center 8 | 14895 | 416 | 416.7 | -0.77 | -0.119 | -0.118 | -0.001 |
| C4 | Male | Center 9 | 15430 | 417 | 416.7 | 0.22 | -0.12 | -0.118 | -0.002 |
| C4 | Female | Center 1 | 10493 | 448.8 | 447.9 | 0.91 | -5.013 | -4.872 | -0.142 |
| C4 | Female | Center 2 | 13215 | 448.6 | 447.9 | 0.73 | -4.868 | -4.872 | 0.004 |
| C4 | Female | Center 3 | 13773 | 446.4 | 447.9 | -1.5 | -4.763 | -4.872 | 0.109 |
| C4 | Female | Center 4 | 14131 | 447.5 | 447.9 | -0.41 | -4.98 | -4.872 | -0.108 |
| C4 | Female | Center 5 | 13700 | 449.5 | 447.9 | 1.65 | -4.889 | -4.872 | -0.017 |
| C4 | Female | Center 6 | 13407 | 445.4 | 447.9 | -2.42 | -4.937 | -4.872 | -0.065 |
| C4 | Female | Center 7 | 13990 | 450.4 | 447.9 | 2.53 | -4.93 | -4.872 | -0.058 |
| C4 | Female | Center 8 | 14829 | 447.5 | 447.9 | -0.38 | -4.856 | -4.872 | 0.015 |
| C4 | Female | Center 9 | 15302 | 447.7 | 447.9 | -0.12 | -4.867 | -4.872 | 0.005 |
| C5 | Male | Center 1 | 14319 | 402.1 | 399.4 | 2.75 | -0.104 | -0.133 | 0.029 |
| C5 | Male | Center 2 | 15324 | 400 | 399.4 | 0.64 | -0.1 | -0.133 | 0.033 |
| C5 | Male | Center 3 | 16486 | 399.5 | 399.4 | 0.11 | -0.087 | -0.133 | 0.046 |
| C5 | Male | Center 4 | 17083 | 398.1 | 399.4 | -1.22 | -0.129 | -0.133 | 0.004 |
| C5 | Male | Center 5 | 17570 | 400.2 | 399.4 | 0.82 | -0.126 | -0.133 | 0.007 |
| C5 | Male | Center 6 | 16727 | 396.8 | 399.4 | -2.53 | -0.24 | -0.133 | -0.107 |
| C5 | Male | Center 7 | 17491 | 400.3 | 399.4 | 0.94 | -0.159 | -0.133 | -0.026 |
| C5 | Male | Center 8 | 18262 | 398.5 | 399.4 | -0.81 | -0.124 | -0.133 | 0.009 |
| C5 | Male | Center 9 | 18930 | 399.6 | 399.4 | 0.23 | -0.138 | -0.133 | -0.005 |
| C5 | Female | Center 1 | 12831 | 437.1 | 436.5 | 0.61 | -4.782 | -4.764 | -0.018 |
| C5 | Female | Center 2 | 15035 | 436.9 | 436.5 | 0.45 | -4.546 | -4.764 | 0.217 |
| C5 | Female | Center 3 | 15966 | 434.5 | 436.5 | -2.02 | -4.449 | -4.764 | 0.315 |
| C5 | Female | Center 4 | 16158 | 434.7 | 436.5 | -1.74 | -4.674 | -4.764 | 0.089 |
| C5 | Female | Center 5 | 16039 | 438 | 436.5 | 1.5 | -4.769 | -4.764 | -0.005 |
| C5 | Female | Center 6 | 15645 | 433.6 | 436.5 | -2.89 | -4.841 | -4.764 | -0.077 |
| C5 | Female | Center 7 | 16353 | 438.7 | 436.5 | 2.18 | -4.792 | -4.764 | -0.028 |
| C5 | Female | Center 8 | 17199 | 436 | 436.5 | -0.48 | -4.741 | -4.764 | 0.022 |
| C5 | Female | Center 9 | 17822 | 436.4 | 436.5 | -0.04 | -4.762 | -4.764 | 0.002 |
| C6 | Male | Center 1 | 19319 | 372.5 | 370.7 | 1.78 | 0.009 | -0.019 | 0.029 |
| C6 | Male | Center 2 | 20268 | 371.3 | 370.7 | 0.59 | -0.008 | -0.019 | 0.011 |
| C6 | Male | Center 3 | 21749 | 371.2 | 370.7 | 0.46 | 0.004 | -0.019 | 0.023 |
| C6 | Male | Center 4 | 21915 | 369.1 | 370.7 | -1.63 | -0.009 | -0.019 | 0.011 |
| C6 | Male | Center 5 | 22915 | 371.3 | 370.7 | 0.57 | -0.007 | -0.019 | 0.013 |
| C6 | Male | Center 6 | 22229 | 368.7 | 370.7 | -2.02 | -0.068 | -0.019 | -0.049 |
| C6 | Male | Center 7 | 23291 | 371.6 | 370.7 | 0.84 | -0.032 | -0.019 | -0.012 |
| C6 | Male | Center 8 | 23943 | 370.2 | 370.7 | -0.5 | -0.021 | -0.019 | -0.002 |
| C6 | Male | Center 9 | 24963 | 371.1 | 370.7 | 0.38 | -0.037 | -0.019 | -0.017 |
| C6 | Female | Center 1 | 17036 | 407.1 | 407.4 | -0.3 | -4.311 | -4.258 | -0.054 |
| C6 | Female | Center 2 | 18694 | 409.1 | 407.4 | 1.73 | -4.241 | -4.258 | 0.017 |
| C6 | Female | Center 3 | 20013 | 406.6 | 407.4 | -0.72 | -4.148 | -4.258 | 0.109 |
| C6 | Female | Center 4 | 19866 | 405.5 | 407.4 | -1.82 | -4.321 | -4.258 | -0.063 |
| C6 | Female | Center 5 | 20148 | 408.7 | 407.4 | 1.39 | -4.277 | -4.258 | -0.02 |
| C6 | Female | Center 6 | 20005 | 405 | 407.4 | -2.31 | -4.306 | -4.258 | -0.048 |
| C6 | Female | Center 7 | 20877 | 409 | 407.4 | 1.64 | -4.292 | -4.258 | -0.034 |
| C6 | Female | Center 8 | 21653 | 407.1 | 407.4 | -0.28 | -4.237 | -4.258 | 0.02 |
| C6 | Female | Center 9 | 22532 | 407.4 | 407.4 | 0.03 | -4.257 | -4.258 | 0 |
| C7 | Male | Center 1 | 21257 | 331.7 | 330.5 | 1.13 | 0.071 | 0.064 | 0.007 |
| C7 | Male | Center 2 | 23092 | 330.8 | 330.5 | 0.22 | 0.053 | 0.064 | -0.012 |
| C7 | Male | Center 3 | 24361 | 331.4 | 330.5 | 0.88 | 0.071 | 0.064 | 0.007 |
| C7 | Male | Center 4 | 24652 | 328.9 | 330.5 | -1.65 | 0.076 | 0.064 | 0.012 |
| C7 | Male | Center 5 | 25523 | 330.6 | 330.5 | 0.08 | 0.094 | 0.064 | 0.029 |
| C7 | Male | Center 6 | 25269 | 329.2 | 330.5 | -1.35 | 0.022 | 0.064 | -0.043 |
| C7 | Male | Center 7 | 26328 | 331.6 | 330.5 | 1.06 | 0.071 | 0.064 | 0.006 |
| C7 | Male | Center 8 | 26878 | 330.3 | 330.5 | -0.24 | 0.055 | 0.064 | -0.01 |
| C7 | Male | Center 9 | 28056 | 330.8 | 330.5 | 0.24 | 0.056 | 0.064 | -0.008 |
| C7 | Female | Center 1 | 18877 | 357.5 | 358.4 | -0.88 | -3.685 | -3.653 | -0.031 |
| C7 | Female | Center 2 | 20857 | 359.3 | 358.4 | 0.94 | -3.615 | -3.653 | 0.038 |
| C7 | Female | Center 3 | 22139 | 358.4 | 358.4 | -0.03 | -3.607 | -3.653 | 0.046 |
| C7 | Female | Center 4 | 22062 | 356.5 | 358.4 | -1.88 | -3.654 | -3.653 | -0.001 |
| C7 | Female | Center 5 | 22163 | 359.3 | 358.4 | 0.89 | -3.671 | -3.653 | -0.017 |
| C7 | Female | Center 6 | 22511 | 357.7 | 358.4 | -0.71 | -3.721 | -3.653 | -0.067 |
| C7 | Female | Center 7 | 23344 | 359.7 | 358.4 | 1.31 | -3.675 | -3.653 | -0.022 |
| C7 | Female | Center 8 | 24047 | 358.5 | 358.4 | 0.05 | -3.65 | -3.653 | 0.004 |
| C7 | Female | Center 9 | 25048 | 358.5 | 358.4 | 0.12 | -3.657 | -3.653 | -0.003 |
| T1 | Male | Center 1 | 21353 | 295.4 | 294.3 | 1.03 | -0.669 | -0.649 | -0.019 |
| T1 | Male | Center 2 | 23460 | 293.7 | 294.3 | -0.57 | -0.664 | -0.649 | -0.015 |
| T1 | Male | Center 3 | 24645 | 295.4 | 294.3 | 1.09 | -0.654 | -0.649 | -0.005 |
| T1 | Male | Center 4 | 25052 | 292.9 | 294.3 | -1.42 | -0.636 | -0.649 | 0.013 |
| T1 | Male | Center 5 | 25799 | 293.9 | 294.3 | -0.38 | -0.606 | -0.649 | 0.043 |
| T1 | Male | Center 6 | 25707 | 293.7 | 294.3 | -0.61 | -0.654 | -0.649 | -0.004 |
| T1 | Male | Center 7 | 26700 | 295.2 | 294.3 | 0.9 | -0.649 | -0.649 | -0 |
| T1 | Male | Center 8 | 27251 | 294.3 | 294.3 | 0 | -0.659 | -0.649 | -0.01 |
| T1 | Male | Center 9 | 28425 | 294.5 | 294.3 | 0.19 | -0.658 | -0.649 | -0.008 |
| T1 | Female | Center 1 | 19042 | 315.4 | 320.5 | -5.1 | -3.339 | -3.772 | 0.433 |
| T1 | Female | Center 2 | 21197 | 321.2 | 320.5 | 0.64 | -3.765 | -3.772 | 0.007 |
| T1 | Female | Center 3 | 22404 | 320.7 | 320.5 | 0.11 | -3.75 | -3.772 | 0.022 |
| T1 | Female | Center 4 | 22434 | 319.4 | 320.5 | -1.15 | -3.83 | -3.772 | -0.059 |
| T1 | Female | Center 5 | 22477 | 320.8 | 320.5 | 0.22 | -3.791 | -3.772 | -0.019 |
| T1 | Female | Center 6 | 22873 | 320.6 | 320.5 | 0.04 | -3.8 | -3.772 | -0.028 |
| T1 | Female | Center 7 | 23693 | 321.8 | 320.5 | 1.24 | -3.791 | -3.772 | -0.019 |
| T1 | Female | Center 8 | 24401 | 320.7 | 320.5 | 0.16 | -3.762 | -3.772 | 0.01 |
| T1 | Female | Center 9 | 25407 | 320.7 | 320.5 | 0.12 | -3.777 | -3.772 | -0.005 |
| T2 | Male | Center 1 | 21338 | 273.8 | 273.3 | 0.49 | -0.592 | -0.592 | -0 |
| T2 | Male | Center 2 | 23479 | 272.9 | 273.3 | -0.34 | -0.613 | -0.592 | -0.021 |
| T2 | Male | Center 3 | 24613 | 274.2 | 273.3 | 0.97 | -0.582 | -0.592 | 0.01 |
| T2 | Male | Center 4 | 25029 | 272.4 | 273.3 | -0.92 | -0.59 | -0.592 | 0.002 |
| T2 | Male | Center 5 | 25795 | 273.1 | 273.3 | -0.18 | -0.574 | -0.592 | 0.018 |
| T2 | Male | Center 6 | 25691 | 272.8 | 273.3 | -0.44 | -0.592 | -0.592 | -0 |
| T2 | Male | Center 7 | 26675 | 273.8 | 273.3 | 0.51 | -0.592 | -0.592 | 0 |
| T2 | Male | Center 8 | 27229 | 273.4 | 273.3 | 0.08 | -0.601 | -0.592 | -0.009 |
| T2 | Male | Center 9 | 28407 | 273.3 | 273.3 | -0.02 | -0.596 | -0.592 | -0.004 |
| T2 | Female | Center 1 | 19030 | 296.9 | 298.1 | -1.27 | -3.449 | -3.479 | 0.031 |
| T2 | Female | Center 2 | 21171 | 298.7 | 298.1 | 0.55 | -3.426 | -3.479 | 0.054 |
| T2 | Female | Center 3 | 22370 | 298.1 | 298.1 | -0.01 | -3.417 | -3.479 | 0.062 |
| T2 | Female | Center 4 | 22410 | 296.5 | 298.1 | -1.59 | -3.53 | -3.479 | -0.051 |
| T2 | Female | Center 5 | 22469 | 298.4 | 298.1 | 0.3 | -3.471 | -3.479 | 0.008 |
| T2 | Female | Center 6 | 22852 | 298.4 | 298.1 | 0.32 | -3.509 | -3.479 | -0.03 |
| T2 | Female | Center 7 | 23676 | 298.7 | 298.1 | 0.56 | -3.496 | -3.479 | -0.016 |
| T2 | Female | Center 8 | 24387 | 298.2 | 298.1 | 0.05 | -3.469 | -3.479 | 0.01 |
| T2 | Female | Center 9 | 25387 | 298.1 | 298.1 | -0 | -3.476 | -3.479 | 0.003 |
| T3 | Male | Center 1 | 21361 | 275.2 | 275.1 | 0.11 | -0.721 | -0.712 | -0.009 |
| T3 | Male | Center 2 | 23508 | 274.9 | 275.1 | -0.17 | -0.724 | -0.712 | -0.012 |
| T3 | Male | Center 3 | 24653 | 276.1 | 275.1 | 0.99 | -0.709 | -0.712 | 0.003 |
| T3 | Male | Center 4 | 25067 | 274.2 | 275.1 | -0.92 | -0.7 | -0.712 | 0.012 |
| T3 | Male | Center 5 | 25815 | 275.1 | 275.1 | -0.01 | -0.698 | -0.712 | 0.014 |
| T3 | Male | Center 6 | 25724 | 274.7 | 275.1 | -0.44 | -0.717 | -0.712 | -0.005 |
| T3 | Male | Center 7 | 26721 | 275.5 | 275.1 | 0.39 | -0.707 | -0.712 | 0.005 |
| T3 | Male | Center 8 | 27257 | 275.3 | 275.1 | 0.15 | -0.722 | -0.712 | -0.01 |
| T3 | Male | Center 9 | 28438 | 275.1 | 275.1 | -0.01 | -0.717 | -0.712 | -0.005 |
| T3 | Female | Center 1 | 19094 | 293.3 | 294.6 | -1.29 | -3.575 | -3.608 | 0.032 |
| T3 | Female | Center 2 | 21188 | 295.3 | 294.6 | 0.74 | -3.526 | -3.608 | 0.082 |
| T3 | Female | Center 3 | 22377 | 294.8 | 294.6 | 0.25 | -3.553 | -3.608 | 0.055 |
| T3 | Female | Center 4 | 22426 | 293 | 294.6 | -1.62 | -3.637 | -3.608 | -0.029 |
| T3 | Female | Center 5 | 22456 | 294.9 | 294.6 | 0.34 | -3.638 | -3.608 | -0.03 |
| T3 | Female | Center 6 | 22854 | 295.6 | 294.6 | 0.97 | -3.641 | -3.608 | -0.033 |
| T3 | Female | Center 7 | 23676 | 295.3 | 294.6 | 0.67 | -3.609 | -3.608 | -0.001 |
| T3 | Female | Center 8 | 24392 | 294.8 | 294.6 | 0.16 | -3.603 | -3.608 | 0.005 |
| T3 | Female | Center 9 | 25401 | 294.6 | 294.6 | -0.01 | -3.607 | -3.608 | 0.001 |
| T4 | Male | Center 1 | 21385 | 273.2 | 273 | 0.2 | -0.765 | -0.758 | -0.007 |
| T4 | Male | Center 2 | 23521 | 272.8 | 273 | -0.21 | -0.777 | -0.758 | -0.019 |
| T4 | Male | Center 3 | 24616 | 273.8 | 273 | 0.79 | -0.747 | -0.758 | 0.011 |
| T4 | Male | Center 4 | 25085 | 272.1 | 273 | -0.94 | -0.739 | -0.758 | 0.019 |
| T4 | Male | Center 5 | 25809 | 272.9 | 273 | -0.08 | -0.739 | -0.758 | 0.019 |
| T4 | Male | Center 6 | 25715 | 272.7 | 273 | -0.3 | -0.766 | -0.758 | -0.008 |
| T4 | Male | Center 7 | 26717 | 273.4 | 273 | 0.4 | -0.765 | -0.758 | -0.007 |
| T4 | Male | Center 8 | 27262 | 273.2 | 273 | 0.2 | -0.766 | -0.758 | -0.008 |
| T4 | Male | Center 9 | 28442 | 273 | 273 | 0.01 | -0.763 | -0.758 | -0.004 |
| T4 | Female | Center 1 | 19115 | 288.2 | 289.6 | -1.42 | -3.647 | -3.699 | 0.052 |
| T4 | Female | Center 2 | 21184 | 290.4 | 289.6 | 0.78 | -3.65 | -3.699 | 0.049 |
| T4 | Female | Center 3 | 22373 | 289.6 | 289.6 | 0.01 | -3.652 | -3.699 | 0.047 |
| T4 | Female | Center 4 | 22438 | 288.2 | 289.6 | -1.41 | -3.745 | -3.699 | -0.046 |
| T4 | Female | Center 5 | 22444 | 289.9 | 289.6 | 0.28 | -3.718 | -3.699 | -0.019 |
| T4 | Female | Center 6 | 22842 | 290.6 | 289.6 | 1.02 | -3.742 | -3.699 | -0.043 |
| T4 | Female | Center 7 | 23694 | 290.4 | 289.6 | 0.77 | -3.7 | -3.699 | -0.001 |
| T4 | Female | Center 8 | 24390 | 290.1 | 289.6 | 0.48 | -3.681 | -3.699 | 0.018 |
| T4 | Female | Center 9 | 25400 | 289.9 | 289.6 | 0.27 | -3.69 | -3.699 | 0.009 |
| T5 | Male | Center 1 | 21429 | 268.9 | 268.5 | 0.39 | -0.698 | -0.702 | 0.004 |
| T5 | Male | Center 2 | 23539 | 268.2 | 268.5 | -0.26 | -0.699 | -0.702 | 0.002 |
| T5 | Male | Center 3 | 24677 | 269.3 | 268.5 | 0.77 | -0.701 | -0.702 | 0.001 |
| T5 | Male | Center 4 | 25150 | 267.7 | 268.5 | -0.82 | -0.693 | -0.702 | 0.009 |
| T5 | Male | Center 5 | 25879 | 268.3 | 268.5 | -0.19 | -0.689 | -0.702 | 0.013 |
| T5 | Male | Center 6 | 25808 | 268.1 | 268.5 | -0.37 | -0.71 | -0.702 | -0.008 |
| T5 | Male | Center 7 | 26770 | 269 | 268.5 | 0.48 | -0.71 | -0.702 | -0.008 |
| T5 | Male | Center 8 | 27327 | 268.6 | 268.5 | 0.13 | -0.708 | -0.702 | -0.007 |
| T5 | Male | Center 9 | 28501 | 268.5 | 268.5 | 0.01 | -0.707 | -0.702 | -0.006 |
| T5 | Female | Center 1 | 19142 | 283 | 284.4 | -1.41 | -3.555 | -3.612 | 0.058 |
| T5 | Female | Center 2 | 21247 | 284.8 | 284.4 | 0.42 | -3.577 | -3.612 | 0.036 |
| T5 | Female | Center 3 | 22409 | 284.1 | 284.4 | -0.31 | -3.575 | -3.612 | 0.038 |
| T5 | Female | Center 4 | 22485 | 282.9 | 284.4 | -1.52 | -3.67 | -3.612 | -0.058 |
| T5 | Female | Center 5 | 22514 | 284.3 | 284.4 | -0.09 | -3.651 | -3.612 | -0.039 |
| T5 | Female | Center 6 | 22911 | 285.1 | 284.4 | 0.71 | -3.673 | -3.612 | -0.06 |
| T5 | Female | Center 7 | 23734 | 284.8 | 284.4 | 0.44 | -3.624 | -3.612 | -0.012 |
| T5 | Female | Center 8 | 24467 | 284.5 | 284.4 | 0.14 | -3.592 | -3.612 | 0.02 |
| T5 | Female | Center 9 | 25459 | 284.3 | 284.4 | -0.06 | -3.611 | -3.612 | 0.001 |
| T6 | Male | Center 1 | 21902 | 268.9 | 268.3 | 0.64 | -0.692 | -0.68 | -0.012 |
| T6 | Male | Center 2 | 23949 | 267.8 | 268.3 | -0.42 | -0.691 | -0.68 | -0.01 |
| T6 | Male | Center 3 | 25149 | 269 | 268.3 | 0.79 | -0.674 | -0.68 | 0.006 |
| T6 | Male | Center 4 | 25687 | 267.4 | 268.3 | -0.9 | -0.657 | -0.68 | 0.023 |
| T6 | Male | Center 5 | 26431 | 268.1 | 268.3 | -0.11 | -0.671 | -0.68 | 0.009 |
| T6 | Male | Center 6 | 26335 | 267.9 | 268.3 | -0.39 | -0.69 | -0.68 | -0.009 |
| T6 | Male | Center 7 | 27345 | 268.7 | 268.3 | 0.41 | -0.684 | -0.68 | -0.004 |
| T6 | Male | Center 8 | 27906 | 268.4 | 268.3 | 0.12 | -0.682 | -0.68 | -0.002 |
| T6 | Male | Center 9 | 29096 | 268.3 | 268.3 | 0.02 | -0.686 | -0.68 | -0.005 |
| T6 | Female | Center 1 | 19554 | 279.8 | 281.2 | -1.37 | -3.456 | -3.526 | 0.07 |
| T6 | Female | Center 2 | 21552 | 282 | 281.2 | 0.79 | -3.462 | -3.526 | 0.064 |
| T6 | Female | Center 3 | 22824 | 281 | 281.2 | -0.15 | -3.515 | -3.526 | 0.011 |
| T6 | Female | Center 4 | 22962 | 280 | 281.2 | -1.24 | -3.558 | -3.526 | -0.032 |
| T6 | Female | Center 5 | 22997 | 281.7 | 281.2 | 0.5 | -3.508 | -3.526 | 0.018 |
| T6 | Female | Center 6 | 23404 | 282.3 | 281.2 | 1.15 | -3.58 | -3.526 | -0.054 |
| T6 | Female | Center 7 | 24225 | 282 | 281.2 | 0.81 | -3.541 | -3.526 | -0.014 |
| T6 | Female | Center 8 | 24960 | 281.3 | 281.2 | 0.12 | -3.507 | -3.526 | 0.02 |
| T6 | Female | Center 9 | 25970 | 281.1 | 281.2 | -0.05 | -3.523 | -3.526 | 0.003 |
| T7 | Male | Center 1 | 23569 | 265.2 | 264.4 | 0.82 | -0.641 | -0.64 | -0.001 |
| T7 | Male | Center 2 | 25427 | 263.8 | 264.4 | -0.65 | -0.637 | -0.64 | 0.003 |
| T7 | Male | Center 3 | 26736 | 265.2 | 264.4 | 0.78 | -0.646 | -0.64 | -0.006 |
| T7 | Male | Center 4 | 27634 | 263.7 | 264.4 | -0.72 | -0.626 | -0.64 | 0.014 |
| T7 | Male | Center 5 | 28359 | 264.3 | 264.4 | -0.14 | -0.626 | -0.64 | 0.014 |
| T7 | Male | Center 6 | 28236 | 263.9 | 264.4 | -0.46 | -0.661 | -0.64 | -0.021 |
| T7 | Male | Center 7 | 29362 | 264.8 | 264.4 | 0.44 | -0.64 | -0.64 | -0 |
| T7 | Male | Center 8 | 29918 | 264.5 | 264.4 | 0.05 | -0.639 | -0.64 | 0.001 |
| T7 | Male | Center 9 | 31159 | 264.4 | 264.4 | 0.02 | -0.647 | -0.64 | -0.007 |
| T7 | Female | Center 1 | 21084 | 274.3 | 275.4 | -1.12 | -3.418 | -3.47 | 0.052 |
| T7 | Female | Center 2 | 22862 | 275.9 | 275.4 | 0.43 | -3.44 | -3.47 | 0.031 |
| T7 | Female | Center 3 | 24168 | 275.2 | 275.4 | -0.23 | -3.429 | -3.47 | 0.041 |
| T7 | Female | Center 4 | 24642 | 273.9 | 275.4 | -1.56 | -3.501 | -3.47 | -0.031 |
| T7 | Female | Center 5 | 24605 | 275.7 | 275.4 | 0.32 | -3.489 | -3.47 | -0.018 |
| T7 | Female | Center 6 | 25055 | 276.1 | 275.4 | 0.65 | -3.515 | -3.47 | -0.045 |
| T7 | Female | Center 7 | 25964 | 276.2 | 275.4 | 0.78 | -3.472 | -3.47 | -0.002 |
| T7 | Female | Center 8 | 26690 | 275.5 | 275.4 | 0.09 | -3.45 | -3.47 | 0.021 |
| T7 | Female | Center 9 | 27762 | 275.3 | 275.4 | -0.08 | -3.468 | -3.47 | 0.002 |
| T8 | Male | Center 1 | 25911 | 261.5 | 260.8 | 0.61 | -0.585 | -0.606 | 0.021 |
| T8 | Male | Center 2 | 27654 | 260.5 | 260.8 | -0.36 | -0.619 | -0.606 | -0.013 |
| T8 | Male | Center 3 | 28986 | 261.6 | 260.8 | 0.8 | -0.602 | -0.606 | 0.004 |
| T8 | Male | Center 4 | 30468 | 260.2 | 260.8 | -0.68 | -0.603 | -0.606 | 0.003 |
| T8 | Male | Center 5 | 31162 | 260.7 | 260.8 | -0.13 | -0.596 | -0.606 | 0.01 |
| T8 | Male | Center 6 | 31061 | 260.2 | 260.8 | -0.6 | -0.622 | -0.606 | -0.016 |
| T8 | Male | Center 7 | 32331 | 261.3 | 260.8 | 0.5 | -0.602 | -0.606 | 0.004 |
| T8 | Male | Center 8 | 32848 | 260.9 | 260.8 | 0.05 | -0.611 | -0.606 | -0.005 |
| T8 | Male | Center 9 | 34171 | 260.9 | 260.8 | 0.01 | -0.613 | -0.606 | -0.007 |
| T8 | Female | Center 1 | 23213 | 268.9 | 269.7 | -0.85 | -3.355 | -3.429 | 0.074 |
| T8 | Female | Center 2 | 24824 | 270.5 | 269.7 | 0.75 | -3.414 | -3.429 | 0.014 |
| T8 | Female | Center 3 | 26079 | 269.9 | 269.7 | 0.19 | -3.384 | -3.429 | 0.044 |
| T8 | Female | Center 4 | 27007 | 268.6 | 269.7 | -1.18 | -3.471 | -3.429 | -0.043 |
| T8 | Female | Center 5 | 26953 | 270.4 | 269.7 | 0.67 | -3.458 | -3.429 | -0.029 |
| T8 | Female | Center 6 | 27468 | 270.7 | 269.7 | 0.94 | -3.483 | -3.429 | -0.055 |
| T8 | Female | Center 7 | 28519 | 270.5 | 269.7 | 0.74 | -3.43 | -3.429 | -0.002 |
| T8 | Female | Center 8 | 29231 | 269.9 | 269.7 | 0.13 | -3.422 | -3.429 | 0.006 |
| T8 | Female | Center 9 | 30362 | 269.7 | 269.7 | -0.08 | -3.427 | -3.429 | 0.002 |
| T9 | Male | Center 1 | 27723 | 264.1 | 263.7 | 0.41 | -0.596 | -0.626 | 0.03 |
| T9 | Male | Center 2 | 29871 | 263.5 | 263.7 | -0.21 | -0.634 | -0.626 | -0.008 |
| T9 | Male | Center 3 | 31040 | 264.7 | 263.7 | 0.99 | -0.643 | -0.626 | -0.018 |
| T9 | Male | Center 4 | 32945 | 263.1 | 263.7 | -0.65 | -0.621 | -0.626 | 0.004 |
| T9 | Male | Center 5 | 33665 | 263.6 | 263.7 | -0.14 | -0.625 | -0.626 | 0.001 |
| T9 | Male | Center 6 | 33568 | 263 | 263.7 | -0.74 | -0.631 | -0.626 | -0.006 |
| T9 | Male | Center 7 | 34988 | 264.2 | 263.7 | 0.43 | -0.618 | -0.626 | 0.008 |
| T9 | Male | Center 8 | 35448 | 263.8 | 263.7 | 0.11 | -0.63 | -0.626 | -0.004 |
| T9 | Male | Center 9 | 36856 | 263.7 | 263.7 | 0 | -0.631 | -0.626 | -0.005 |
| T9 | Female | Center 1 | 25288 | 273.4 | 274.7 | -1.35 | -3.543 | -3.626 | 0.083 |
| T9 | Female | Center 2 | 27043 | 275.4 | 274.7 | 0.66 | -3.613 | -3.626 | 0.014 |
| T9 | Female | Center 3 | 28118 | 275.1 | 274.7 | 0.42 | -3.614 | -3.626 | 0.012 |
| T9 | Female | Center 4 | 29546 | 273.7 | 274.7 | -1.05 | -3.667 | -3.626 | -0.041 |
| T9 | Female | Center 5 | 29537 | 275.4 | 274.7 | 0.71 | -3.676 | -3.626 | -0.05 |
| T9 | Female | Center 6 | 30123 | 275.4 | 274.7 | 0.71 | -3.652 | -3.626 | -0.026 |
| T9 | Female | Center 7 | 31256 | 275.5 | 274.7 | 0.78 | -3.639 | -3.626 | -0.013 |
| T9 | Female | Center 8 | 31948 | 274.8 | 274.7 | 0.1 | -3.614 | -3.626 | 0.013 |
| T9 | Female | Center 9 | 33149 | 274.7 | 274.7 | -0.07 | -3.624 | -3.626 | 0.002 |
| T10 | Male | Center 1 | 28453 | 260.2 | 259.7 | 0.45 | -0.705 | -0.73 | 0.025 |
| T10 | Male | Center 2 | 31209 | 259.5 | 259.7 | -0.19 | -0.746 | -0.73 | -0.017 |
| T10 | Male | Center 3 | 32212 | 260.5 | 259.7 | 0.73 | -0.733 | -0.73 | -0.003 |
| T10 | Male | Center 4 | 34262 | 259.1 | 259.7 | -0.67 | -0.727 | -0.73 | 0.003 |
| T10 | Male | Center 5 | 35057 | 259.6 | 259.7 | -0.17 | -0.727 | -0.73 | 0.003 |
| T10 | Male | Center 6 | 34929 | 259.1 | 259.7 | -0.66 | -0.728 | -0.73 | 0.002 |
| T10 | Male | Center 7 | 36432 | 260.3 | 259.7 | 0.54 | -0.733 | -0.73 | -0.004 |
| T10 | Male | Center 8 | 36840 | 259.8 | 259.7 | 0.12 | -0.731 | -0.73 | -0.001 |
| T10 | Male | Center 9 | 38310 | 259.7 | 259.7 | 0.01 | -0.735 | -0.73 | -0.005 |
| T10 | Female | Center 1 | 26311 | 272.5 | 273.5 | -1.07 | -3.631 | -3.724 | 0.093 |
| T10 | Female | Center 2 | 28648 | 274.2 | 273.5 | 0.67 | -3.74 | -3.724 | -0.016 |
| T10 | Female | Center 3 | 29428 | 274.1 | 273.5 | 0.52 | -3.714 | -3.724 | 0.009 |
| T10 | Female | Center 4 | 31111 | 272.4 | 273.5 | -1.12 | -3.751 | -3.724 | -0.027 |
| T10 | Female | Center 5 | 31177 | 273.6 | 273.5 | 0.03 | -3.776 | -3.724 | -0.052 |
| T10 | Female | Center 6 | 31819 | 274.1 | 273.5 | 0.53 | -3.729 | -3.724 | -0.006 |
| T10 | Female | Center 7 | 33018 | 274.3 | 273.5 | 0.72 | -3.741 | -3.724 | -0.017 |
| T10 | Female | Center 8 | 33669 | 273.7 | 273.5 | 0.14 | -3.702 | -3.724 | 0.022 |
| T10 | Female | Center 9 | 34907 | 273.5 | 273.5 | -0.08 | -3.721 | -3.724 | 0.003 |
| T11 | Male | Center 1 | 28646 | 248 | 247.4 | 0.56 | -0.89 | -0.903 | 0.013 |
| T11 | Male | Center 2 | 31675 | 247 | 247.4 | -0.44 | -0.899 | -0.903 | 0.004 |
| T11 | Male | Center 3 | 32547 | 247.9 | 247.4 | 0.5 | -0.897 | -0.903 | 0.006 |
| T11 | Male | Center 4 | 34678 | 246.8 | 247.4 | -0.58 | -0.912 | -0.903 | -0.009 |
| T11 | Male | Center 5 | 35506 | 247.4 | 247.4 | -0.04 | -0.907 | -0.903 | -0.004 |
| T11 | Male | Center 6 | 35323 | 246.8 | 247.4 | -0.55 | -0.896 | -0.903 | 0.007 |
| T11 | Male | Center 7 | 36901 | 247.9 | 247.4 | 0.46 | -0.903 | -0.903 | 0 |
| T11 | Male | Center 8 | 37235 | 247.6 | 247.4 | 0.2 | -0.911 | -0.903 | -0.008 |
| T11 | Male | Center 9 | 38745 | 247.4 | 247.4 | 0.04 | -0.907 | -0.903 | -0.004 |
| T11 | Female | Center 1 | 26537 | 261.3 | 262.4 | -1.17 | -3.611 | -3.733 | 0.122 |
| T11 | Female | Center 2 | 29234 | 263.2 | 262.4 | 0.72 | -3.74 | -3.733 | -0.007 |
| T11 | Female | Center 3 | 29857 | 262.9 | 262.4 | 0.51 | -3.736 | -3.733 | -0.004 |
| T11 | Female | Center 4 | 31623 | 261.2 | 262.4 | -1.19 | -3.757 | -3.733 | -0.025 |
| T11 | Female | Center 5 | 31710 | 263.2 | 262.4 | 0.75 | -3.791 | -3.733 | -0.058 |
| T11 | Female | Center 6 | 32307 | 262.9 | 262.4 | 0.47 | -3.734 | -3.733 | -0.001 |
| T11 | Female | Center 7 | 33572 | 263 | 262.4 | 0.59 | -3.743 | -3.733 | -0.01 |
| T11 | Female | Center 8 | 34179 | 262.6 | 262.4 | 0.17 | -3.72 | -3.733 | 0.013 |
| T11 | Female | Center 9 | 35453 | 262.4 | 262.4 | -0.07 | -3.73 | -3.733 | 0.002 |
| T12 | Male | Center 1 | 28640 | 231.9 | 231 | 0.91 | -1 | -1.009 | 0.01 |
| T12 | Male | Center 2 | 31661 | 230.6 | 231 | -0.38 | -1.001 | -1.009 | 0.008 |
| T12 | Male | Center 3 | 32523 | 231.3 | 231 | 0.28 | -1.012 | -1.009 | -0.003 |
| T12 | Male | Center 4 | 34632 | 230.4 | 231 | -0.58 | -1.01 | -1.009 | -0.001 |
| T12 | Male | Center 5 | 35470 | 231 | 231 | -0.03 | -1.016 | -1.009 | -0.007 |
| T12 | Male | Center 6 | 35263 | 230.3 | 231 | -0.64 | -1.003 | -1.009 | 0.006 |
| T12 | Male | Center 7 | 36886 | 231.4 | 231 | 0.43 | -1.01 | -1.009 | -0.001 |
| T12 | Male | Center 8 | 37216 | 231.2 | 231 | 0.18 | -1.014 | -1.009 | -0.005 |
| T12 | Male | Center 9 | 38717 | 231 | 231 | 0.04 | -1.013 | -1.009 | -0.004 |
| T12 | Female | Center 1 | 26503 | 245.6 | 246.1 | -0.56 | -3.525 | -3.615 | 0.089 |
| T12 | Female | Center 2 | 29169 | 246.8 | 246.1 | 0.61 | -3.626 | -3.615 | -0.012 |
| T12 | Female | Center 3 | 29798 | 246.3 | 246.1 | 0.18 | -3.602 | -3.615 | 0.012 |
| T12 | Female | Center 4 | 31577 | 246 | 246.1 | -0.12 | -3.664 | -3.615 | -0.049 |
| T12 | Female | Center 5 | 31678 | 246.9 | 246.1 | 0.76 | -3.677 | -3.615 | -0.062 |
| T12 | Female | Center 6 | 32223 | 247 | 246.1 | 0.85 | -3.632 | -3.615 | -0.017 |
| T12 | Female | Center 7 | 33500 | 246.7 | 246.1 | 0.55 | -3.62 | -3.615 | -0.006 |
| T12 | Female | Center 8 | 34122 | 246.2 | 246.1 | 0.04 | -3.594 | -3.615 | 0.02 |
| T12 | Female | Center 9 | 35390 | 246.1 | 246.1 | -0.03 | -3.614 | -3.615 | 0 |
| L1 | Male | Center 1 | 28687 | 233.2 | 232 | 1.17 | -0.901 | -0.916 | 0.015 |
| L1 | Male | Center 2 | 31633 | 231.6 | 232 | -0.47 | -0.891 | -0.916 | 0.025 |
| L1 | Male | Center 3 | 32555 | 232.2 | 232 | 0.16 | -0.908 | -0.916 | 0.008 |
| L1 | Male | Center 4 | 34629 | 231.4 | 232 | -0.64 | -0.915 | -0.916 | 0 |
| L1 | Male | Center 5 | 35484 | 232.1 | 232 | 0.03 | -0.933 | -0.916 | -0.018 |
| L1 | Male | Center 6 | 35280 | 231.4 | 232 | -0.62 | -0.924 | -0.916 | -0.008 |
| L1 | Male | Center 7 | 36879 | 232.4 | 232 | 0.38 | -0.914 | -0.916 | 0.002 |
| L1 | Male | Center 8 | 37214 | 232.2 | 232 | 0.16 | -0.922 | -0.916 | -0.006 |
| L1 | Male | Center 9 | 38727 | 232.1 | 232 | 0.08 | -0.922 | -0.916 | -0.006 |
| L1 | Female | Center 1 | 26500 | 246.6 | 246.8 | -0.17 | -3.42 | -3.498 | 0.078 |
| L1 | Female | Center 2 | 29174 | 247.3 | 246.8 | 0.52 | -3.504 | -3.498 | -0.006 |
| L1 | Female | Center 3 | 29814 | 246.9 | 246.8 | 0.1 | -3.485 | -3.498 | 0.013 |
| L1 | Female | Center 4 | 31545 | 246.1 | 246.8 | -0.73 | -3.526 | -3.498 | -0.028 |
| L1 | Female | Center 5 | 31680 | 246.9 | 246.8 | 0.16 | -3.549 | -3.498 | -0.051 |
| L1 | Female | Center 6 | 32244 | 246.7 | 246.8 | -0.12 | -3.485 | -3.498 | 0.013 |
| L1 | Female | Center 7 | 33498 | 247.3 | 246.8 | 0.5 | -3.503 | -3.498 | -0.005 |
| L1 | Female | Center 8 | 34134 | 246.8 | 246.8 | 0.05 | -3.484 | -3.498 | 0.014 |
| L1 | Female | Center 9 | 35395 | 246.8 | 246.8 | -0.01 | -3.499 | -3.498 | -0.001 |
| L2 | Male | Center 1 | 28684 | 244.8 | 243.5 | 1.23 | -0.674 | -0.707 | 0.034 |
| L2 | Male | Center 2 | 31635 | 243.1 | 243.5 | -0.46 | -0.684 | -0.707 | 0.024 |
| L2 | Male | Center 3 | 32543 | 243.8 | 243.5 | 0.3 | -0.7 | -0.707 | 0.008 |
| L2 | Male | Center 4 | 34619 | 242.9 | 243.5 | -0.62 | -0.71 | -0.707 | -0.003 |
| L2 | Male | Center 5 | 35467 | 243.6 | 243.5 | 0.03 | -0.726 | -0.707 | -0.018 |
| L2 | Male | Center 6 | 35283 | 242.8 | 243.5 | -0.75 | -0.72 | -0.707 | -0.012 |
| L2 | Male | Center 7 | 36870 | 243.9 | 243.5 | 0.38 | -0.706 | -0.707 | 0.001 |
| L2 | Male | Center 8 | 37221 | 243.6 | 243.5 | 0.09 | -0.713 | -0.707 | -0.005 |
| L2 | Male | Center 9 | 38726 | 243.6 | 243.5 | 0.08 | -0.715 | -0.707 | -0.008 |
| L2 | Female | Center 1 | 26528 | 258.5 | 258.8 | -0.29 | -3.455 | -3.532 | 0.076 |
| L2 | Female | Center 2 | 29186 | 259.5 | 258.8 | 0.74 | -3.538 | -3.532 | -0.006 |
| L2 | Female | Center 3 | 29842 | 259 | 258.8 | 0.25 | -3.519 | -3.532 | 0.013 |
| L2 | Female | Center 4 | 31559 | 258 | 258.8 | -0.77 | -3.588 | -3.532 | -0.056 |
| L2 | Female | Center 5 | 31698 | 259.2 | 258.8 | 0.36 | -3.595 | -3.532 | -0.064 |
| L2 | Female | Center 6 | 32277 | 258.5 | 258.8 | -0.25 | -3.509 | -3.532 | 0.022 |
| L2 | Female | Center 7 | 33536 | 259.3 | 258.8 | 0.46 | -3.529 | -3.532 | 0.003 |
| L2 | Female | Center 8 | 34167 | 258.9 | 258.8 | 0.08 | -3.512 | -3.532 | 0.02 |
| L2 | Female | Center 9 | 35423 | 258.8 | 258.8 | -0.02 | -3.532 | -3.532 | -0.001 |
| L3 | Male | Center 1 | 28684 | 251.7 | 250.7 | 0.99 | -0.705 | -0.742 | 0.037 |
| L3 | Male | Center 2 | 31637 | 250.5 | 250.7 | -0.22 | -0.724 | -0.742 | 0.018 |
| L3 | Male | Center 3 | 32513 | 251 | 250.7 | 0.3 | -0.727 | -0.742 | 0.015 |
| L3 | Male | Center 4 | 34632 | 250 | 250.7 | -0.7 | -0.749 | -0.742 | -0.007 |
| L3 | Male | Center 5 | 35478 | 250.8 | 250.7 | 0.12 | -0.761 | -0.742 | -0.018 |
| L3 | Male | Center 6 | 35274 | 249.8 | 250.7 | -0.87 | -0.749 | -0.742 | -0.006 |
| L3 | Male | Center 7 | 36879 | 251.2 | 250.7 | 0.45 | -0.747 | -0.742 | -0.005 |
| L3 | Male | Center 8 | 37226 | 250.8 | 250.7 | 0.06 | -0.746 | -0.742 | -0.003 |
| L3 | Male | Center 9 | 38725 | 250.8 | 250.7 | 0.13 | -0.751 | -0.742 | -0.009 |
| L3 | Female | Center 1 | 26524 | 264.7 | 265.1 | -0.41 | -3.648 | -3.695 | 0.047 |
| L3 | Female | Center 2 | 29199 | 266.6 | 265.1 | 1.46 | -3.695 | -3.695 | 0 |
| L3 | Female | Center 3 | 29806 | 266.3 | 265.1 | 1.15 | -3.694 | -3.695 | 0.002 |
| L3 | Female | Center 4 | 31556 | 265.3 | 265.1 | 0.17 | -3.768 | -3.695 | -0.073 |
| L3 | Female | Center 5 | 31691 | 266.3 | 265.1 | 1.18 | -3.75 | -3.695 | -0.055 |
| L3 | Female | Center 6 | 32227 | 265.8 | 265.1 | 0.61 | -3.686 | -3.695 | 0.009 |
| L3 | Female | Center 7 | 33517 | 266.6 | 265.1 | 1.5 | -3.713 | -3.695 | -0.018 |
| L3 | Female | Center 8 | 34141 | 266.1 | 265.1 | 0.91 | -3.683 | -3.695 | 0.013 |
| L3 | Female | Center 9 | 35403 | 266 | 265.1 | 0.89 | -3.705 | -3.695 | -0.01 |
| L4 | Male | Center 1 | 28659 | 256.1 | 254.8 | 1.3 | -0.632 | -0.633 | 0.001 |
| L4 | Male | Center 2 | 31617 | 254.8 | 254.8 | -0.03 | -0.677 | -0.633 | -0.044 |
| L4 | Male | Center 3 | 32548 | 256.4 | 254.8 | 1.57 | -0.5 | -0.633 | 0.133 |
| L4 | Male | Center 4 | 34629 | 254 | 254.8 | -0.78 | -0.623 | -0.633 | 0.01 |
| L4 | Male | Center 5 | 35477 | 254.9 | 254.8 | 0.13 | -0.641 | -0.633 | -0.008 |
| L4 | Male | Center 6 | 35291 | 253.9 | 254.8 | -0.87 | -0.641 | -0.633 | -0.009 |
| L4 | Male | Center 7 | 36875 | 255.1 | 254.8 | 0.29 | -0.633 | -0.633 | -0.001 |
| L4 | Male | Center 8 | 37215 | 254.8 | 254.8 | 0.03 | -0.633 | -0.633 | -0 |
| L4 | Male | Center 9 | 38721 | 254.9 | 254.8 | 0.1 | -0.64 | -0.633 | -0.008 |
| L4 | Female | Center 1 | 26527 | 271.1 | 271.4 | -0.35 | -3.655 | -3.711 | 0.056 |
| L4 | Female | Center 2 | 29175 | 272.1 | 271.4 | 0.64 | -3.701 | -3.711 | 0.01 |
| L4 | Female | Center 3 | 29812 | 271.7 | 271.4 | 0.29 | -3.706 | -3.711 | 0.005 |
| L4 | Female | Center 4 | 31559 | 271.5 | 271.4 | 0.08 | -3.785 | -3.711 | -0.074 |
| L4 | Female | Center 5 | 31677 | 271.7 | 271.4 | 0.33 | -3.755 | -3.711 | -0.045 |
| L4 | Female | Center 6 | 32257 | 271.3 | 271.4 | -0.1 | -3.698 | -3.711 | 0.013 |
| L4 | Female | Center 7 | 33526 | 271.9 | 271.4 | 0.48 | -3.709 | -3.711 | 0.001 |
| L4 | Female | Center 8 | 34131 | 271.5 | 271.4 | 0.05 | -3.694 | -3.711 | 0.017 |
| L4 | Female | Center 9 | 35400 | 271.4 | 271.4 | 0.01 | -3.712 | -3.711 | -0.001 |
| L5 | Male | Center 1 | 28644 | 258.6 | 257.5 | 1.17 | -0.427 | -0.439 | 0.012 |
| L5 | Male | Center 2 | 31646 | 257.4 | 257.5 | -0.08 | -0.505 | -0.439 | -0.066 |
| L5 | Male | Center 3 | 32566 | 259.2 | 257.5 | 1.76 | -0.284 | -0.439 | 0.155 |
| L5 | Male | Center 4 | 34640 | 256.8 | 257.5 | -0.62 | -0.436 | -0.439 | 0.003 |
| L5 | Male | Center 5 | 35504 | 257.4 | 257.5 | -0.04 | -0.454 | -0.439 | -0.015 |
| L5 | Male | Center 6 | 35330 | 256.8 | 257.5 | -0.64 | -0.442 | -0.439 | -0.003 |
| L5 | Male | Center 7 | 36891 | 257.7 | 257.5 | 0.25 | -0.445 | -0.439 | -0.006 |
| L5 | Male | Center 8 | 37252 | 257.5 | 257.5 | 0.07 | -0.435 | -0.439 | 0.004 |
| L5 | Male | Center 9 | 38743 | 257.5 | 257.5 | 0.07 | -0.444 | -0.439 | -0.005 |
| L5 | Female | Center 1 | 26484 | 272.6 | 273.9 | -1.35 | -3.627 | -3.694 | 0.067 |
| L5 | Female | Center 2 | 29186 | 274.7 | 273.9 | 0.83 | -3.728 | -3.694 | -0.034 |
| L5 | Female | Center 3 | 29813 | 274.1 | 273.9 | 0.23 | -3.669 | -3.694 | 0.025 |
| L5 | Female | Center 4 | 31549 | 273.1 | 273.9 | -0.83 | -3.757 | -3.694 | -0.063 |
| L5 | Female | Center 5 | 31706 | 274.2 | 273.9 | 0.32 | -3.735 | -3.694 | -0.04 |
| L5 | Female | Center 6 | 32249 | 273.8 | 273.9 | -0.16 | -3.698 | -3.694 | -0.003 |
| L5 | Female | Center 7 | 33530 | 274.4 | 273.9 | 0.45 | -3.69 | -3.694 | 0.005 |
| L5 | Female | Center 8 | 34147 | 273.9 | 273.9 | 0.01 | -3.681 | -3.694 | 0.014 |
| L5 | Female | Center 9 | 35400 | 273.9 | 273.9 | -0.02 | -3.693 | -3.694 | 0.002 |
| S1 | Male | Center 1 | 28685 | 272 | 272.3 | -0.26 | -1.325 | -1.344 | 0.019 |
| S1 | Male | Center 2 | 31633 | 272.2 | 272.3 | -0.13 | -1.354 | -1.344 | -0.01 |
| S1 | Male | Center 3 | 32563 | 272.8 | 272.3 | 0.49 | -1.341 | -1.344 | 0.003 |
| S1 | Male | Center 4 | 34676 | 271.8 | 272.3 | -0.46 | -1.349 | -1.344 | -0.005 |
| S1 | Male | Center 5 | 35520 | 272.4 | 272.3 | 0.08 | -1.345 | -1.344 | -0.001 |
| S1 | Male | Center 6 | 35278 | 271.8 | 272.3 | -0.53 | -1.321 | -1.344 | 0.023 |
| S1 | Male | Center 7 | 36896 | 272.7 | 272.3 | 0.37 | -1.35 | -1.344 | -0.006 |
| S1 | Male | Center 8 | 37262 | 272.5 | 272.3 | 0.23 | -1.349 | -1.344 | -0.005 |
| S1 | Male | Center 9 | 38751 | 272.5 | 272.3 | 0.17 | -1.352 | -1.344 | -0.008 |
| S1 | Female | Center 1 | 26556 | 290.1 | 292.5 | -2.43 | -4.213 | -4.339 | 0.126 |
| S1 | Female | Center 2 | 29226 | 293 | 292.5 | 0.49 | -4.337 | -4.339 | 0.003 |
| S1 | Female | Center 3 | 29873 | 292.7 | 292.5 | 0.22 | -4.344 | -4.339 | -0.005 |
| S1 | Female | Center 4 | 31610 | 292 | 292.5 | -0.52 | -4.392 | -4.339 | -0.052 |
| S1 | Female | Center 5 | 31750 | 292.4 | 292.5 | -0.08 | -4.38 | -4.339 | -0.04 |
| S1 | Female | Center 6 | 32265 | 292.8 | 292.5 | 0.3 | -4.391 | -4.339 | -0.052 |
| S1 | Female | Center 7 | 33570 | 293 | 292.5 | 0.47 | -4.335 | -4.339 | 0.004 |
| S1 | Female | Center 8 | 34168 | 292.6 | 292.5 | 0.14 | -4.317 | -4.339 | 0.022 |
| S1 | Female | Center 9 | 35454 | 292.5 | 292.5 | 0.01 | -4.339 | -4.339 | 0 |
| Left rib 1 | Male | Center 1 | 21344 | 328.7 | 328.1 | 0.52 | -1.896 | -1.847 | -0.049 |
| Left rib 1 | Male | Center 2 | 23547 | 328.1 | 328.1 | 0.01 | -1.878 | -1.847 | -0.031 |
| Left rib 1 | Male | Center 3 | 24703 | 329 | 328.1 | 0.86 | -1.805 | -1.847 | 0.042 |
| Left rib 1 | Male | Center 4 | 25068 | 326.9 | 328.1 | -1.22 | -1.832 | -1.847 | 0.015 |
| Left rib 1 | Male | Center 5 | 25840 | 328.7 | 328.1 | 0.54 | -1.823 | -1.847 | 0.024 |
| Left rib 1 | Male | Center 6 | 25665 | 326.8 | 328.1 | -1.37 | -1.816 | -1.847 | 0.031 |
| Left rib 1 | Male | Center 7 | 26741 | 328.8 | 328.1 | 0.67 | -1.879 | -1.847 | -0.032 |
| Left rib 1 | Male | Center 8 | 27274 | 328.2 | 328.1 | 0.06 | -1.853 | -1.847 | -0.006 |
| Left rib 1 | Male | Center 9 | 28450 | 328.3 | 328.1 | 0.16 | -1.856 | -1.847 | -0.009 |
| Left rib 1 | Female | Center 1 | 19091 | 346.1 | 346.9 | -0.8 | -4.518 | -4.528 | 0.011 |
| Left rib 1 | Female | Center 2 | 21268 | 347.3 | 346.9 | 0.42 | -4.52 | -4.528 | 0.008 |
| Left rib 1 | Female | Center 3 | 22458 | 346.8 | 346.9 | -0.13 | -4.481 | -4.528 | 0.047 |
| Left rib 1 | Female | Center 4 | 22442 | 345.8 | 346.9 | -1.12 | -4.549 | -4.528 | -0.02 |
| Left rib 1 | Female | Center 5 | 22500 | 349.2 | 346.9 | 2.32 | -4.587 | -4.528 | -0.058 |
| Left rib 1 | Female | Center 6 | 22835 | 344.7 | 346.9 | -2.24 | -4.506 | -4.528 | 0.023 |
| Left rib 1 | Female | Center 7 | 23712 | 348.3 | 346.9 | 1.4 | -4.564 | -4.528 | -0.035 |
| Left rib 1 | Female | Center 8 | 24427 | 346.9 | 346.9 | 0.03 | -4.502 | -4.528 | 0.026 |
| Left rib 1 | Female | Center 9 | 25435 | 346.9 | 346.9 | -0.04 | -4.526 | -4.528 | 0.002 |
| Left rib 2 | Male | Center 1 | 21483 | 244.2 | 245.4 | -1.2 | -1.048 | -1.072 | 0.024 |
| Left rib 2 | Male | Center 2 | 23569 | 245 | 245.4 | -0.46 | -1.23 | -1.072 | -0.158 |
| Left rib 2 | Male | Center 3 | 24722 | 246.9 | 245.4 | 1.52 | -1.058 | -1.072 | 0.014 |
| Left rib 2 | Male | Center 4 | 25131 | 244.4 | 245.4 | -1 | -1.072 | -1.072 | 0 |
| Left rib 2 | Male | Center 5 | 25897 | 246 | 245.4 | 0.55 | -1.051 | -1.072 | 0.022 |
| Left rib 2 | Male | Center 6 | 25667 | 245 | 245.4 | -0.39 | -1.067 | -1.072 | 0.005 |
| Left rib 2 | Male | Center 7 | 26768 | 245.8 | 245.4 | 0.4 | -1.085 | -1.072 | -0.012 |
| Left rib 2 | Male | Center 8 | 27311 | 245.6 | 245.4 | 0.17 | -1.078 | -1.072 | -0.006 |
| Left rib 2 | Male | Center 9 | 28500 | 245.5 | 245.4 | 0.04 | -1.07 | -1.072 | 0.002 |
| Left rib 2 | Female | Center 1 | 19164 | 278.9 | 284 | -5.08 | -3.985 | -4.378 | 0.393 |
| Left rib 2 | Female | Center 2 | 21291 | 284.5 | 284 | 0.48 | -4.257 | -4.378 | 0.121 |
| Left rib 2 | Female | Center 3 | 22464 | 281.6 | 284 | -2.4 | -3.999 | -4.378 | 0.379 |
| Left rib 2 | Female | Center 4 | 22472 | 282.1 | 284 | -1.89 | -4.429 | -4.378 | -0.051 |
| Left rib 2 | Female | Center 5 | 22565 | 286 | 284 | 1.95 | -4.308 | -4.378 | 0.07 |
| Left rib 2 | Female | Center 6 | 22808 | 281.8 | 284 | -2.19 | -4.03 | -4.378 | 0.348 |
| Left rib 2 | Female | Center 7 | 23754 | 285.1 | 284 | 1.13 | -4.409 | -4.378 | -0.031 |
| Left rib 2 | Female | Center 8 | 24464 | 281.5 | 284 | -2.49 | -4.005 | -4.378 | 0.373 |
| Left rib 2 | Female | Center 9 | 25474 | 284 | 284 | -0.04 | -4.377 | -4.378 | 0.001 |
| Left rib 3 | Male | Center 1 | 21899 | 232.6 | 234.4 | -1.77 | -1.752 | -1.797 | 0.046 |
| Left rib 3 | Male | Center 2 | 23993 | 234.5 | 234.4 | 0.15 | -1.839 | -1.797 | -0.042 |
| Left rib 3 | Male | Center 3 | 25206 | 235.9 | 234.4 | 1.55 | -1.804 | -1.797 | -0.007 |
| Left rib 3 | Male | Center 4 | 25626 | 233.1 | 234.4 | -1.32 | -1.786 | -1.797 | 0.011 |
| Left rib 3 | Male | Center 5 | 26401 | 234.9 | 234.4 | 0.56 | -1.791 | -1.797 | 0.007 |
| Left rib 3 | Male | Center 6 | 26228 | 234.1 | 234.4 | -0.33 | -1.782 | -1.797 | 0.016 |
| Left rib 3 | Male | Center 7 | 27317 | 234.9 | 234.4 | 0.52 | -1.813 | -1.797 | -0.016 |
| Left rib 3 | Male | Center 8 | 27863 | 234.6 | 234.4 | 0.25 | -1.805 | -1.797 | -0.007 |
| Left rib 3 | Male | Center 9 | 29067 | 234.4 | 234.4 | 0.04 | -1.799 | -1.797 | -0.002 |
| Left rib 3 | Female | Center 1 | 19752 | 280.2 | 284 | -3.86 | -5.3 | -5.362 | 0.062 |
| Left rib 3 | Female | Center 2 | 21888 | 284.3 | 284 | 0.25 | -5.177 | -5.362 | 0.186 |
| Left rib 3 | Female | Center 3 | 23120 | 283.9 | 284 | -0.13 | -5.34 | -5.362 | 0.023 |
| Left rib 3 | Female | Center 4 | 23184 | 282.6 | 284 | -1.46 | -5.421 | -5.362 | -0.058 |
| Left rib 3 | Female | Center 5 | 23216 | 285.8 | 284 | 1.78 | -5.397 | -5.362 | -0.034 |
| Left rib 3 | Female | Center 6 | 23569 | 284.7 | 284 | 0.62 | -5.396 | -5.362 | -0.034 |
| Left rib 3 | Female | Center 7 | 24481 | 285.2 | 284 | 1.13 | -5.404 | -5.362 | -0.041 |
| Left rib 3 | Female | Center 8 | 25205 | 284.3 | 284 | 0.23 | -5.35 | -5.362 | 0.012 |
| Left rib 3 | Female | Center 9 | 26249 | 284 | 284 | -0.08 | -5.358 | -5.362 | 0.005 |
| Left rib 4 | Male | Center 1 | 24383 | 238.6 | 240.4 | -1.83 | -1.623 | -1.708 | 0.084 |
| Left rib 4 | Male | Center 2 | 25897 | 241.4 | 240.4 | 0.99 | -1.769 | -1.708 | -0.062 |
| Left rib 4 | Male | Center 3 | 27199 | 242.3 | 240.4 | 1.93 | -1.75 | -1.708 | -0.042 |
| Left rib 4 | Male | Center 4 | 28375 | 239.1 | 240.4 | -1.28 | -1.687 | -1.708 | 0.02 |
| Left rib 4 | Male | Center 5 | 29140 | 240.7 | 240.4 | 0.32 | -1.695 | -1.708 | 0.013 |
| Left rib 4 | Male | Center 6 | 28990 | 239.6 | 240.4 | -0.79 | -1.706 | -1.708 | 0.002 |
| Left rib 4 | Male | Center 7 | 30137 | 240.7 | 240.4 | 0.35 | -1.72 | -1.708 | -0.012 |
| Left rib 4 | Male | Center 8 | 30683 | 240.6 | 240.4 | 0.19 | -1.708 | -1.708 | -0 |
| Left rib 4 | Male | Center 9 | 31948 | 240.4 | 240.4 | -0.01 | -1.708 | -1.708 | -0 |
| Left rib 4 | Female | Center 1 | 22602 | 282.1 | 286.1 | -4.05 | -5.369 | -5.461 | 0.093 |
| Left rib 4 | Female | Center 2 | 24000 | 289.6 | 286.1 | 3.45 | -5.507 | -5.461 | -0.046 |
| Left rib 4 | Female | Center 3 | 25237 | 288.5 | 286.1 | 2.41 | -5.496 | -5.461 | -0.035 |
| Left rib 4 | Female | Center 4 | 26233 | 283.9 | 286.1 | -2.25 | -5.49 | -5.461 | -0.028 |
| Left rib 4 | Female | Center 5 | 26200 | 286.9 | 286.1 | 0.72 | -5.477 | -5.461 | -0.016 |
| Left rib 4 | Female | Center 6 | 26643 | 286 | 286.1 | -0.09 | -5.464 | -5.461 | -0.003 |
| Left rib 4 | Female | Center 7 | 27620 | 287 | 286.1 | 0.86 | -5.481 | -5.461 | -0.02 |
| Left rib 4 | Female | Center 8 | 28350 | 286.3 | 286.1 | 0.16 | -5.425 | -5.461 | 0.036 |
| Left rib 4 | Female | Center 9 | 29451 | 286 | 286.1 | -0.16 | -5.452 | -5.461 | 0.009 |
| Left rib 5 | Male | Center 1 | 27768 | 236.1 | 237.3 | -1.18 | -1.438 | -1.495 | 0.057 |
| Left rib 5 | Male | Center 2 | 29353 | 238.2 | 237.3 | 0.92 | -1.497 | -1.495 | -0.003 |
| Left rib 5 | Male | Center 3 | 30555 | 239.3 | 237.3 | 2.05 | -1.523 | -1.495 | -0.028 |
| Left rib 5 | Male | Center 4 | 32611 | 236.2 | 237.3 | -1.03 | -1.485 | -1.495 | 0.01 |
| Left rib 5 | Male | Center 5 | 33350 | 237.5 | 237.3 | 0.2 | -1.481 | -1.495 | 0.013 |
| Left rib 5 | Male | Center 6 | 33249 | 236.2 | 237.3 | -1.05 | -1.523 | -1.495 | -0.028 |
| Left rib 5 | Male | Center 7 | 34607 | 237.4 | 237.3 | 0.13 | -1.498 | -1.495 | -0.003 |
| Left rib 5 | Male | Center 8 | 35089 | 237.4 | 237.3 | 0.12 | -1.506 | -1.495 | -0.011 |
| Left rib 5 | Male | Center 9 | 36474 | 237.2 | 237.3 | -0.02 | -1.495 | -1.495 | -0 |
| Left rib 5 | Female | Center 1 | 25869 | 281.7 | 285.6 | -3.89 | -5.217 | -5.224 | 0.008 |
| Left rib 5 | Female | Center 2 | 27450 | 288 | 285.6 | 2.4 | -5.219 | -5.224 | 0.006 |
| Left rib 5 | Female | Center 3 | 28334 | 288.4 | 285.6 | 2.8 | -5.262 | -5.224 | -0.038 |
| Left rib 5 | Female | Center 4 | 30115 | 283.7 | 285.6 | -1.94 | -5.261 | -5.224 | -0.037 |
| Left rib 5 | Female | Center 5 | 30116 | 285.3 | 285.6 | -0.3 | -5.195 | -5.224 | 0.029 |
| Left rib 5 | Female | Center 6 | 30674 | 284.6 | 285.6 | -1.07 | -5.233 | -5.224 | -0.009 |
| Left rib 5 | Female | Center 7 | 31834 | 286 | 285.6 | 0.42 | -5.238 | -5.224 | -0.014 |
| Left rib 5 | Female | Center 8 | 32515 | 285.6 | 285.6 | -0.05 | -5.169 | -5.224 | 0.055 |
| Left rib 5 | Female | Center 9 | 33725 | 285.5 | 285.6 | -0.15 | -5.22 | -5.224 | 0.004 |
| Left rib 6 | Male | Center 1 | 28919 | 249.3 | 248.4 | 0.81 | -1.329 | -1.332 | 0.003 |
| Left rib 6 | Male | Center 2 | 31276 | 248.6 | 248.4 | 0.13 | -1.322 | -1.332 | 0.009 |
| Left rib 6 | Male | Center 3 | 32384 | 250 | 248.4 | 1.52 | -1.325 | -1.332 | 0.007 |
| Left rib 6 | Male | Center 4 | 34539 | 247.5 | 248.4 | -0.96 | -1.301 | -1.332 | 0.031 |
| Left rib 6 | Male | Center 5 | 35308 | 248.6 | 248.4 | 0.18 | -1.308 | -1.332 | 0.024 |
| Left rib 6 | Male | Center 6 | 35171 | 247.3 | 248.4 | -1.18 | -1.38 | -1.332 | -0.048 |
| Left rib 6 | Male | Center 7 | 36695 | 248.4 | 248.4 | -0.04 | -1.337 | -1.332 | -0.006 |
| Left rib 6 | Male | Center 8 | 37114 | 248.5 | 248.4 | 0.05 | -1.349 | -1.332 | -0.017 |
| Left rib 6 | Male | Center 9 | 38586 | 248.4 | 248.4 | -0.07 | -1.329 | -1.332 | 0.002 |
| Left rib 6 | Female | Center 1 | 26834 | 298.6 | 299.7 | -1.09 | -5.031 | -5.001 | -0.03 |
| Left rib 6 | Female | Center 2 | 29005 | 300.8 | 299.7 | 1.12 | -4.95 | -5.001 | 0.051 |
| Left rib 6 | Female | Center 3 | 29788 | 302.4 | 299.7 | 2.72 | -4.921 | -5.001 | 0.08 |
| Left rib 6 | Female | Center 4 | 31599 | 298 | 299.7 | -1.7 | -5.033 | -5.001 | -0.032 |
| Left rib 6 | Female | Center 5 | 31627 | 299.3 | 299.7 | -0.32 | -4.967 | -5.001 | 0.033 |
| Left rib 6 | Female | Center 6 | 32233 | 297.8 | 299.7 | -1.82 | -5.044 | -5.001 | -0.043 |
| Left rib 6 | Female | Center 7 | 33470 | 300.3 | 299.7 | 0.66 | -5.058 | -5.001 | -0.057 |
| Left rib 6 | Female | Center 8 | 34151 | 299.5 | 299.7 | -0.14 | -4.962 | -5.001 | 0.039 |
| Left rib 6 | Female | Center 9 | 35405 | 299.6 | 299.7 | -0.11 | -5.001 | -5.001 | 0 |
| Left rib 7 | Male | Center 1 | 28957 | 260.5 | 259.3 | 1.21 | -1.431 | -1.442 | 0.011 |
| Left rib 7 | Male | Center 2 | 31422 | 259.3 | 259.3 | 0.04 | -1.441 | -1.442 | 0.001 |
| Left rib 7 | Male | Center 3 | 32549 | 260.2 | 259.3 | 0.91 | -1.415 | -1.442 | 0.027 |
| Left rib 7 | Male | Center 4 | 34642 | 258.5 | 259.3 | -0.74 | -1.424 | -1.442 | 0.017 |
| Left rib 7 | Male | Center 5 | 35416 | 259.6 | 259.3 | 0.37 | -1.434 | -1.442 | 0.008 |
| Left rib 7 | Male | Center 6 | 35295 | 257.7 | 259.3 | -1.56 | -1.487 | -1.442 | -0.045 |
| Left rib 7 | Male | Center 7 | 36819 | 259.5 | 259.3 | 0.2 | -1.449 | -1.442 | -0.008 |
| Left rib 7 | Male | Center 8 | 37245 | 259.3 | 259.3 | 0.05 | -1.459 | -1.442 | -0.017 |
| Left rib 7 | Male | Center 9 | 38719 | 259.2 | 259.3 | -0.06 | -1.442 | -1.442 | -0 |
| Left rib 7 | Female | Center 1 | 26802 | 316.7 | 317.2 | -0.57 | -5.421 | -5.436 | 0.015 |
| Left rib 7 | Female | Center 2 | 29102 | 317.5 | 317.2 | 0.28 | -5.408 | -5.436 | 0.028 |
| Left rib 7 | Female | Center 3 | 29917 | 318.3 | 317.2 | 1.1 | -5.377 | -5.436 | 0.06 |
| Left rib 7 | Female | Center 4 | 31643 | 316 | 317.2 | -1.19 | -5.47 | -5.436 | -0.033 |
| Left rib 7 | Female | Center 5 | 31678 | 317.6 | 317.2 | 0.34 | -5.433 | -5.436 | 0.004 |
| Left rib 7 | Female | Center 6 | 32302 | 315.3 | 317.2 | -1.95 | -5.473 | -5.436 | -0.037 |
| Left rib 7 | Female | Center 7 | 33536 | 318 | 317.2 | 0.81 | -5.47 | -5.436 | -0.034 |
| Left rib 7 | Female | Center 8 | 34215 | 317.2 | 317.2 | -0.04 | -5.408 | -5.436 | 0.028 |
| Left rib 7 | Female | Center 9 | 35477 | 317.1 | 317.2 | -0.12 | -5.43 | -5.436 | 0.006 |
| Left rib 8 | Male | Center 1 | 28950 | 278.8 | 277.6 | 1.18 | -1.623 | -1.661 | 0.038 |
| Left rib 8 | Male | Center 2 | 31455 | 277.6 | 277.6 | 0.07 | -1.67 | -1.661 | -0.01 |
| Left rib 8 | Male | Center 3 | 32604 | 278 | 277.6 | 0.41 | -1.636 | -1.661 | 0.024 |
| Left rib 8 | Male | Center 4 | 34663 | 277.2 | 277.6 | -0.33 | -1.661 | -1.661 | -0.001 |
| Left rib 8 | Male | Center 5 | 35436 | 277.9 | 277.6 | 0.33 | -1.645 | -1.661 | 0.015 |
| Left rib 8 | Male | Center 6 | 35320 | 275.9 | 277.6 | -1.68 | -1.699 | -1.661 | -0.038 |
| Left rib 8 | Male | Center 7 | 36845 | 277.9 | 277.6 | 0.28 | -1.669 | -1.661 | -0.008 |
| Left rib 8 | Male | Center 8 | 37267 | 277.7 | 277.6 | 0.13 | -1.679 | -1.661 | -0.019 |
| Left rib 8 | Male | Center 9 | 38748 | 277.5 | 277.6 | -0.05 | -1.664 | -1.661 | -0.003 |
| Left rib 8 | Female | Center 1 | 26794 | 332.2 | 332.2 | -0 | -5.925 | -5.982 | 0.056 |
| Left rib 8 | Female | Center 2 | 29119 | 331.9 | 332.2 | -0.27 | -5.937 | -5.982 | 0.044 |
| Left rib 8 | Female | Center 3 | 29937 | 332.4 | 332.2 | 0.15 | -5.958 | -5.982 | 0.023 |
| Left rib 8 | Female | Center 4 | 31634 | 331.6 | 332.2 | -0.63 | -6.056 | -5.982 | -0.074 |
| Left rib 8 | Female | Center 5 | 31691 | 333.1 | 332.2 | 0.86 | -5.992 | -5.982 | -0.011 |
| Left rib 8 | Female | Center 6 | 32306 | 330.7 | 332.2 | -1.55 | -6.009 | -5.982 | -0.027 |
| Left rib 8 | Female | Center 7 | 33547 | 333 | 332.2 | 0.82 | -6.008 | -5.982 | -0.026 |
| Left rib 8 | Female | Center 8 | 34208 | 332.4 | 332.2 | 0.2 | -5.957 | -5.982 | 0.025 |
| Left rib 8 | Female | Center 9 | 35484 | 332.1 | 332.2 | -0.14 | -5.972 | -5.982 | 0.009 |
| Left rib 9 | Male | Center 1 | 28876 | 298.2 | 297.6 | 0.66 | -1.502 | -1.536 | 0.034 |
| Left rib 9 | Male | Center 2 | 31544 | 298.1 | 297.6 | 0.57 | -1.556 | -1.536 | -0.02 |
| Left rib 9 | Male | Center 3 | 32649 | 298 | 297.6 | 0.45 | -1.509 | -1.536 | 0.027 |
| Left rib 9 | Male | Center 4 | 34678 | 297.4 | 297.6 | -0.17 | -1.53 | -1.536 | 0.006 |
| Left rib 9 | Male | Center 5 | 35481 | 297.8 | 297.6 | 0.25 | -1.526 | -1.536 | 0.01 |
| Left rib 9 | Male | Center 6 | 35345 | 295.6 | 297.6 | -1.96 | -1.585 | -1.536 | -0.049 |
| Left rib 9 | Male | Center 7 | 36865 | 297.9 | 297.6 | 0.36 | -1.525 | -1.536 | 0.011 |
| Left rib 9 | Male | Center 8 | 37282 | 297.7 | 297.6 | 0.15 | -1.554 | -1.536 | -0.018 |
| Left rib 9 | Male | Center 9 | 38776 | 297.5 | 297.6 | -0.03 | -1.541 | -1.536 | -0.005 |
| Left rib 9 | Female | Center 1 | 26775 | 342.9 | 343.5 | -0.54 | -5.861 | -6.02 | 0.16 |
| Left rib 9 | Female | Center 2 | 29170 | 343.3 | 343.5 | -0.2 | -5.978 | -6.02 | 0.042 |
| Left rib 9 | Female | Center 3 | 29975 | 343.8 | 343.5 | 0.3 | -6.057 | -6.02 | -0.037 |
| Left rib 9 | Female | Center 4 | 31645 | 343 | 343.5 | -0.46 | -6.12 | -6.02 | -0.1 |
| Left rib 9 | Female | Center 5 | 31711 | 344.4 | 343.5 | 0.96 | -6.044 | -6.02 | -0.024 |
| Left rib 9 | Female | Center 6 | 32349 | 341.5 | 343.5 | -1.97 | -6.03 | -6.02 | -0.01 |
| Left rib 9 | Female | Center 7 | 33576 | 344.2 | 343.5 | 0.75 | -6.041 | -6.02 | -0.021 |
| Left rib 9 | Female | Center 8 | 34239 | 343.5 | 343.5 | 0.04 | -5.977 | -6.02 | 0.044 |
| Left rib 9 | Female | Center 9 | 35512 | 343.3 | 343.5 | -0.17 | -6.011 | -6.02 | 0.009 |
| Left rib 10 | Male | Center 1 | 28669 | 323.7 | 323.2 | 0.52 | -1.549 | -1.596 | 0.047 |
| Left rib 10 | Male | Center 2 | 31703 | 323.6 | 323.2 | 0.44 | -1.612 | -1.596 | -0.016 |
| Left rib 10 | Male | Center 3 | 32637 | 323.7 | 323.2 | 0.55 | -1.579 | -1.596 | 0.017 |
| Left rib 10 | Male | Center 4 | 34746 | 323 | 323.2 | -0.17 | -1.589 | -1.596 | 0.007 |
| Left rib 10 | Male | Center 5 | 35539 | 323.6 | 323.2 | 0.42 | -1.593 | -1.596 | 0.003 |
| Left rib 10 | Male | Center 6 | 35459 | 321.1 | 323.2 | -2.1 | -1.628 | -1.596 | -0.032 |
| Left rib 10 | Male | Center 7 | 36937 | 323.6 | 323.2 | 0.44 | -1.597 | -1.596 | -0.001 |
| Left rib 10 | Male | Center 8 | 37327 | 323.3 | 323.2 | 0.18 | -1.615 | -1.596 | -0.019 |
| Left rib 10 | Male | Center 9 | 38815 | 323.1 | 323.2 | -0.04 | -1.602 | -1.596 | -0.006 |
| Left rib 10 | Female | Center 1 | 26627 | 375.6 | 376.3 | -0.7 | -6.119 | -6.241 | 0.122 |
| Left rib 10 | Female | Center 2 | 29260 | 376.2 | 376.3 | -0.09 | -6.222 | -6.241 | 0.019 |
| Left rib 10 | Female | Center 3 | 29969 | 375.9 | 376.3 | -0.47 | -6.194 | -6.241 | 0.047 |
| Left rib 10 | Female | Center 4 | 31673 | 375.6 | 376.3 | -0.7 | -6.32 | -6.241 | -0.078 |
| Left rib 10 | Female | Center 5 | 31785 | 377.7 | 376.3 | 1.4 | -6.276 | -6.241 | -0.035 |
| Left rib 10 | Female | Center 6 | 32438 | 374.9 | 376.3 | -1.43 | -6.276 | -6.241 | -0.034 |
| Left rib 10 | Female | Center 7 | 33640 | 377.3 | 376.3 | 0.93 | -6.277 | -6.241 | -0.036 |
| Left rib 10 | Female | Center 8 | 34302 | 376.7 | 376.3 | 0.35 | -6.212 | -6.241 | 0.029 |
| Left rib 10 | Female | Center 9 | 35554 | 376.2 | 376.3 | -0.15 | -6.239 | -6.241 | 0.003 |
| Left rib 11 | Male | Center 1 | 28660 | 335.9 | 336.1 | -0.26 | -2.042 | -2.075 | 0.033 |
| Left rib 11 | Male | Center 2 | 31704 | 336.9 | 336.1 | 0.77 | -2.076 | -2.075 | -0.001 |
| Left rib 11 | Male | Center 3 | 32632 | 336.3 | 336.1 | 0.14 | -2.038 | -2.075 | 0.038 |
| Left rib 11 | Male | Center 4 | 34732 | 336 | 336.1 | -0.11 | -2.08 | -2.075 | -0.005 |
| Left rib 11 | Male | Center 5 | 35573 | 336.5 | 336.1 | 0.41 | -2.063 | -2.075 | 0.012 |
| Left rib 11 | Male | Center 6 | 35427 | 334.3 | 336.1 | -1.85 | -2.114 | -2.075 | -0.039 |
| Left rib 11 | Male | Center 7 | 36950 | 336.9 | 336.1 | 0.77 | -2.089 | -2.075 | -0.014 |
| Left rib 11 | Male | Center 8 | 37295 | 336.3 | 336.1 | 0.17 | -2.09 | -2.075 | -0.015 |
| Left rib 11 | Male | Center 9 | 38811 | 336.1 | 336.1 | 0.02 | -2.081 | -2.075 | -0.006 |
| Left rib 11 | Female | Center 1 | 26644 | 381.1 | 383.5 | -2.41 | -6.75 | -6.913 | 0.163 |
| Left rib 11 | Female | Center 2 | 29344 | 383.4 | 383.5 | -0.04 | -6.909 | -6.913 | 0.003 |
| Left rib 11 | Female | Center 3 | 29943 | 383.1 | 383.5 | -0.39 | -6.895 | -6.913 | 0.017 |
| Left rib 11 | Female | Center 4 | 31682 | 382.9 | 383.5 | -0.59 | -7.003 | -6.913 | -0.09 |
| Left rib 11 | Female | Center 5 | 31831 | 384.8 | 383.5 | 1.28 | -6.947 | -6.913 | -0.035 |
| Left rib 11 | Female | Center 6 | 32404 | 383 | 383.5 | -0.53 | -6.914 | -6.913 | -0.002 |
| Left rib 11 | Female | Center 7 | 33667 | 384.6 | 383.5 | 1.09 | -6.946 | -6.913 | -0.033 |
| Left rib 11 | Female | Center 8 | 34288 | 384 | 383.5 | 0.52 | -6.904 | -6.913 | 0.009 |
| Left rib 11 | Female | Center 9 | 35573 | 383.3 | 383.5 | -0.16 | -6.901 | -6.913 | 0.012 |
| Left rib 12 | Male | Center 1 | 28483 | 322.8 | 322.7 | 0.06 | -2.012 | -2.033 | 0.02 |
| Left rib 12 | Male | Center 2 | 31279 | 322.7 | 322.7 | -0.02 | -2.068 | -2.033 | -0.035 |
| Left rib 12 | Male | Center 3 | 32250 | 322.8 | 322.7 | 0.06 | -2.017 | -2.033 | 0.016 |
| Left rib 12 | Male | Center 4 | 34337 | 322.5 | 322.7 | -0.21 | -2.037 | -2.033 | -0.004 |
| Left rib 12 | Male | Center 5 | 35183 | 322.9 | 322.7 | 0.19 | -1.995 | -2.033 | 0.038 |
| Left rib 12 | Male | Center 6 | 34984 | 321.7 | 322.7 | -1.09 | -2.05 | -2.033 | -0.018 |
| Left rib 12 | Male | Center 7 | 36552 | 323.3 | 322.7 | 0.61 | -2.045 | -2.033 | -0.013 |
| Left rib 12 | Male | Center 8 | 36883 | 323.1 | 322.7 | 0.32 | -2.038 | -2.033 | -0.005 |
| Left rib 12 | Male | Center 9 | 38385 | 322.9 | 322.7 | 0.12 | -2.04 | -2.033 | -0.007 |
| Left rib 12 | Female | Center 1 | 25790 | 374.5 | 379.3 | -4.76 | -5.66 | -6.38 | 0.72 |
| Left rib 12 | Female | Center 2 | 28285 | 378.6 | 379.3 | -0.66 | -6.371 | -6.38 | 0.009 |
| Left rib 12 | Female | Center 3 | 28950 | 378.8 | 379.3 | -0.43 | -6.4 | -6.38 | -0.02 |
| Left rib 12 | Female | Center 4 | 30629 | 378.7 | 379.3 | -0.61 | -6.444 | -6.38 | -0.064 |
| Left rib 12 | Female | Center 5 | 30760 | 380.9 | 379.3 | 1.63 | -6.45 | -6.38 | -0.07 |
| Left rib 12 | Female | Center 6 | 31275 | 378.5 | 379.3 | -0.75 | -6.321 | -6.38 | 0.059 |
| Left rib 12 | Female | Center 7 | 32545 | 380.3 | 379.3 | 1.07 | -6.416 | -6.38 | -0.036 |
| Left rib 12 | Female | Center 8 | 33121 | 379.7 | 379.3 | 0.4 | -6.37 | -6.38 | 0.01 |
| Left rib 12 | Female | Center 9 | 34365 | 379.2 | 379.3 | -0.06 | -6.378 | -6.38 | 0.002 |
| Right rib 1 | Male | Center 1 | 21350 | 328.1 | 328.3 | -0.23 | -1.885 | -1.847 | -0.038 |
| Right rib 1 | Male | Center 2 | 23534 | 328.7 | 328.3 | 0.4 | -1.872 | -1.847 | -0.025 |
| Right rib 1 | Male | Center 3 | 24674 | 329.1 | 328.3 | 0.83 | -1.816 | -1.847 | 0.031 |
| Right rib 1 | Male | Center 4 | 25081 | 327.1 | 328.3 | -1.13 | -1.833 | -1.847 | 0.014 |
| Right rib 1 | Male | Center 5 | 25801 | 328.9 | 328.3 | 0.64 | -1.842 | -1.847 | 0.005 |
| Right rib 1 | Male | Center 6 | 25630 | 326.9 | 328.3 | -1.37 | -1.826 | -1.847 | 0.021 |
| Right rib 1 | Male | Center 7 | 26732 | 329 | 328.3 | 0.69 | -1.862 | -1.847 | -0.015 |
| Right rib 1 | Male | Center 8 | 27260 | 328.3 | 328.3 | 0.07 | -1.844 | -1.847 | 0.002 |
| Right rib 1 | Male | Center 9 | 28434 | 328.5 | 328.3 | 0.2 | -1.857 | -1.847 | -0.01 |
| Right rib 1 | Female | Center 1 | 19115 | 344.9 | 346.6 | -1.72 | -4.627 | -4.589 | -0.038 |
| Right rib 1 | Female | Center 2 | 21219 | 347.7 | 346.6 | 1.05 | -4.609 | -4.589 | -0.02 |
| Right rib 1 | Female | Center 3 | 22440 | 346.3 | 346.6 | -0.3 | -4.556 | -4.589 | 0.033 |
| Right rib 1 | Female | Center 4 | 22431 | 345.7 | 346.6 | -0.97 | -4.629 | -4.589 | -0.04 |
| Right rib 1 | Female | Center 5 | 22472 | 348.8 | 346.6 | 2.11 | -4.628 | -4.589 | -0.039 |
| Right rib 1 | Female | Center 6 | 22793 | 344.9 | 346.6 | -1.74 | -4.535 | -4.589 | 0.054 |
| Right rib 1 | Female | Center 7 | 23696 | 347.8 | 346.6 | 1.2 | -4.609 | -4.589 | -0.02 |
| Right rib 1 | Female | Center 8 | 24391 | 346.7 | 346.6 | 0.02 | -4.563 | -4.589 | 0.026 |
| Right rib 1 | Female | Center 9 | 25411 | 346.7 | 346.6 | 0.01 | -4.59 | -4.589 | -0.001 |
| Right rib 2 | Male | Center 1 | 21479 | 245.4 | 246.3 | -0.91 | -1.047 | -1.064 | 0.016 |
| Right rib 2 | Male | Center 2 | 23560 | 245.7 | 246.3 | -0.57 | -1.218 | -1.064 | -0.154 |
| Right rib 2 | Male | Center 3 | 24719 | 247.6 | 246.3 | 1.33 | -1.038 | -1.064 | 0.025 |
| Right rib 2 | Male | Center 4 | 25119 | 245.2 | 246.3 | -1.1 | -1.057 | -1.064 | 0.007 |
| Right rib 2 | Male | Center 5 | 25863 | 246.8 | 246.3 | 0.51 | -1.04 | -1.064 | 0.023 |
| Right rib 2 | Male | Center 6 | 25665 | 246 | 246.3 | -0.29 | -1.051 | -1.064 | 0.013 |
| Right rib 2 | Male | Center 7 | 26776 | 246.6 | 246.3 | 0.38 | -1.084 | -1.064 | -0.021 |
| Right rib 2 | Male | Center 8 | 27300 | 246.4 | 246.3 | 0.15 | -1.069 | -1.064 | -0.005 |
| Right rib 2 | Male | Center 9 | 28495 | 246.4 | 246.3 | 0.13 | -1.065 | -1.064 | -0.001 |
| Right rib 2 | Female | Center 1 | 19231 | 280.9 | 287 | -6.17 | -4.067 | -4.516 | 0.45 |
| Right rib 2 | Female | Center 2 | 21309 | 287.2 | 287 | 0.15 | -4.374 | -4.516 | 0.142 |
| Right rib 2 | Female | Center 3 | 22483 | 283.8 | 287 | -3.24 | -4.101 | -4.516 | 0.415 |
| Right rib 2 | Female | Center 4 | 22525 | 285.7 | 287 | -1.33 | -4.586 | -4.516 | -0.07 |
| Right rib 2 | Female | Center 5 | 22562 | 288.4 | 287 | 1.36 | -4.425 | -4.516 | 0.091 |
| Right rib 2 | Female | Center 6 | 22838 | 287.2 | 287 | 0.16 | -4.543 | -4.516 | -0.026 |
| Right rib 2 | Female | Center 7 | 23797 | 288.3 | 287 | 1.28 | -4.552 | -4.516 | -0.035 |
| Right rib 2 | Female | Center 8 | 24475 | 287.3 | 287 | 0.21 | -4.501 | -4.516 | 0.015 |
| Right rib 2 | Female | Center 9 | 25508 | 287 | 287 | -0.04 | -4.513 | -4.516 | 0.003 |
| Right rib 3 | Male | Center 1 | 21847 | 230 | 231 | -1.03 | -1.793 | -1.827 | 0.034 |
| Right rib 3 | Male | Center 2 | 23992 | 231.2 | 231 | 0.18 | -1.874 | -1.827 | -0.047 |
| Right rib 3 | Male | Center 3 | 25172 | 232.5 | 231 | 1.43 | -1.836 | -1.827 | -0.008 |
| Right rib 3 | Male | Center 4 | 25593 | 229.8 | 231 | -1.25 | -1.81 | -1.827 | 0.017 |
| Right rib 3 | Male | Center 5 | 26359 | 231.4 | 231 | 0.34 | -1.812 | -1.827 | 0.016 |
| Right rib 3 | Male | Center 6 | 26209 | 230.6 | 231 | -0.39 | -1.817 | -1.827 | 0.011 |
| Right rib 3 | Male | Center 7 | 27282 | 231.4 | 231 | 0.39 | -1.844 | -1.827 | -0.016 |
| Right rib 3 | Male | Center 8 | 27828 | 231.2 | 231 | 0.17 | -1.834 | -1.827 | -0.007 |
| Right rib 3 | Male | Center 9 | 29030 | 231 | 231 | -0 | -1.827 | -1.827 | 0.001 |
| Right rib 3 | Female | Center 1 | 19735 | 278.4 | 283.9 | -5.46 | -4.812 | -5.371 | 0.558 |
| Right rib 3 | Female | Center 2 | 21860 | 284.3 | 283.9 | 0.47 | -5.22 | -5.371 | 0.151 |
| Right rib 3 | Female | Center 3 | 23093 | 283.6 | 283.9 | -0.21 | -5.329 | -5.371 | 0.041 |
| Right rib 3 | Female | Center 4 | 23145 | 282.4 | 283.9 | -1.42 | -5.428 | -5.371 | -0.058 |
| Right rib 3 | Female | Center 5 | 23178 | 285 | 283.9 | 1.19 | -5.28 | -5.371 | 0.091 |
| Right rib 3 | Female | Center 6 | 23536 | 284.5 | 283.9 | 0.66 | -5.413 | -5.371 | -0.043 |
| Right rib 3 | Female | Center 7 | 24448 | 285 | 283.9 | 1.16 | -5.405 | -5.371 | -0.035 |
| Right rib 3 | Female | Center 8 | 25168 | 284.1 | 283.9 | 0.28 | -5.36 | -5.371 | 0.01 |
| Right rib 3 | Female | Center 9 | 26213 | 283.8 | 283.9 | -0.09 | -5.363 | -5.371 | 0.007 |
| Right rib 4 | Male | Center 1 | 24260 | 236.7 | 238.4 | -1.7 | -1.66 | -1.756 | 0.096 |
| Right rib 4 | Male | Center 2 | 25760 | 239.7 | 238.4 | 1.24 | -1.833 | -1.756 | -0.077 |
| Right rib 4 | Male | Center 3 | 27120 | 240.3 | 238.4 | 1.92 | -1.792 | -1.756 | -0.036 |
| Right rib 4 | Male | Center 4 | 28217 | 237.2 | 238.4 | -1.19 | -1.738 | -1.756 | 0.018 |
| Right rib 4 | Male | Center 5 | 28987 | 238.6 | 238.4 | 0.16 | -1.746 | -1.756 | 0.01 |
| Right rib 4 | Male | Center 6 | 28840 | 237.6 | 238.4 | -0.8 | -1.758 | -1.756 | -0.002 |
| Right rib 4 | Male | Center 7 | 29985 | 238.6 | 238.4 | 0.22 | -1.77 | -1.756 | -0.014 |
| Right rib 4 | Male | Center 8 | 30528 | 238.5 | 238.4 | 0.07 | -1.755 | -1.756 | 0.001 |
| Right rib 4 | Male | Center 9 | 31791 | 238.5 | 238.4 | 0.03 | -1.758 | -1.756 | -0.002 |
| Right rib 4 | Female | Center 1 | 22460 | 285.7 | 291 | -5.24 | -5.32 | -5.431 | 0.111 |
| Right rib 4 | Female | Center 2 | 23852 | 293.5 | 291 | 2.47 | -5.492 | -5.431 | -0.061 |
| Right rib 4 | Female | Center 3 | 25145 | 292.7 | 291 | 1.76 | -5.443 | -5.431 | -0.012 |
| Right rib 4 | Female | Center 4 | 26069 | 289.2 | 291 | -1.81 | -5.469 | -5.431 | -0.037 |
| Right rib 4 | Female | Center 5 | 26050 | 291.4 | 291 | 0.38 | -5.428 | -5.431 | 0.004 |
| Right rib 4 | Female | Center 6 | 26480 | 290.9 | 291 | -0.08 | -5.457 | -5.431 | -0.026 |
| Right rib 4 | Female | Center 7 | 27457 | 291.9 | 291 | 0.9 | -5.459 | -5.431 | -0.027 |
| Right rib 4 | Female | Center 8 | 28178 | 291 | 291 | 0.07 | -5.392 | -5.431 | 0.04 |
| Right rib 4 | Female | Center 9 | 29277 | 290.9 | 291 | -0.12 | -5.425 | -5.431 | 0.006 |
| Right rib 5 | Male | Center 1 | 27623 | 235.5 | 236.8 | -1.31 | -1.327 | -1.412 | 0.085 |
| Right rib 5 | Male | Center 2 | 29140 | 238 | 236.8 | 1.16 | -1.418 | -1.412 | -0.006 |
| Right rib 5 | Male | Center 3 | 30358 | 239.1 | 236.8 | 2.26 | -1.436 | -1.412 | -0.024 |
| Right rib 5 | Male | Center 4 | 32384 | 235.6 | 236.8 | -1.17 | -1.393 | -1.412 | 0.019 |
| Right rib 5 | Male | Center 5 | 33115 | 237 | 236.8 | 0.16 | -1.414 | -1.412 | -0.002 |
| Right rib 5 | Male | Center 6 | 33023 | 235.7 | 236.8 | -1.08 | -1.441 | -1.412 | -0.029 |
| Right rib 5 | Male | Center 7 | 34369 | 236.9 | 236.8 | 0.13 | -1.423 | -1.412 | -0.011 |
| Right rib 5 | Male | Center 8 | 34849 | 236.8 | 236.8 | 0.01 | -1.427 | -1.412 | -0.015 |
| Right rib 5 | Male | Center 9 | 36227 | 236.8 | 236.8 | 0 | -1.414 | -1.412 | -0.002 |
| Right rib 5 | Female | Center 1 | 25739 | 285 | 288 | -3.06 | -4.904 | -4.961 | 0.057 |
| Right rib 5 | Female | Center 2 | 27232 | 290.9 | 288 | 2.93 | -4.976 | -4.961 | -0.015 |
| Right rib 5 | Female | Center 3 | 28156 | 292.6 | 288 | 4.56 | -4.952 | -4.961 | 0.009 |
| Right rib 5 | Female | Center 4 | 29908 | 285.9 | 288 | -2.13 | -4.995 | -4.961 | -0.034 |
| Right rib 5 | Female | Center 5 | 29903 | 287.3 | 288 | -0.69 | -4.93 | -4.961 | 0.031 |
| Right rib 5 | Female | Center 6 | 30460 | 287 | 288 | -1.03 | -4.976 | -4.961 | -0.015 |
| Right rib 5 | Female | Center 7 | 31613 | 288.5 | 288 | 0.54 | -5 | -4.961 | -0.04 |
| Right rib 5 | Female | Center 8 | 32290 | 288 | 288 | -0.02 | -4.917 | -4.961 | 0.043 |
| Right rib 5 | Female | Center 9 | 33499 | 287.9 | 288 | -0.13 | -4.959 | -4.961 | 0.002 |
| Right rib 6 | Male | Center 1 | 28924 | 249.7 | 249.3 | 0.41 | -1.357 | -1.388 | 0.031 |
| Right rib 6 | Male | Center 2 | 31240 | 249.5 | 249.3 | 0.15 | -1.385 | -1.388 | 0.003 |
| Right rib 6 | Male | Center 3 | 32359 | 251.2 | 249.3 | 1.9 | -1.372 | -1.388 | 0.016 |
| Right rib 6 | Male | Center 4 | 34527 | 248.2 | 249.3 | -1.09 | -1.364 | -1.388 | 0.024 |
| Right rib 6 | Male | Center 5 | 35298 | 249.4 | 249.3 | 0.13 | -1.382 | -1.388 | 0.006 |
| Right rib 6 | Male | Center 6 | 35173 | 248.2 | 249.3 | -1.13 | -1.438 | -1.388 | -0.05 |
| Right rib 6 | Male | Center 7 | 36680 | 249.3 | 249.3 | 0.03 | -1.391 | -1.388 | -0.003 |
| Right rib 6 | Male | Center 8 | 37097 | 249.3 | 249.3 | 0.03 | -1.404 | -1.388 | -0.016 |
| Right rib 6 | Male | Center 9 | 38574 | 249.3 | 249.3 | -0.04 | -1.389 | -1.388 | -0.001 |
| Right rib 6 | Female | Center 1 | 26833 | 306.2 | 306.3 | -0.09 | -5.087 | -5.104 | 0.017 |
| Right rib 6 | Female | Center 2 | 28967 | 307.1 | 306.3 | 0.79 | -5.063 | -5.104 | 0.042 |
| Right rib 6 | Female | Center 3 | 29750 | 309.2 | 306.3 | 2.84 | -5.04 | -5.104 | 0.064 |
| Right rib 6 | Female | Center 4 | 31584 | 304.6 | 306.3 | -1.78 | -5.156 | -5.104 | -0.052 |
| Right rib 6 | Female | Center 5 | 31607 | 305.8 | 306.3 | -0.58 | -5.082 | -5.104 | 0.022 |
| Right rib 6 | Female | Center 6 | 32211 | 304.9 | 306.3 | -1.48 | -5.15 | -5.104 | -0.046 |
| Right rib 6 | Female | Center 7 | 33457 | 307.1 | 306.3 | 0.76 | -5.155 | -5.104 | -0.051 |
| Right rib 6 | Female | Center 8 | 34123 | 306.3 | 306.3 | -0.06 | -5.066 | -5.104 | 0.038 |
| Right rib 6 | Female | Center 9 | 35380 | 306.2 | 306.3 | -0.17 | -5.096 | -5.104 | 0.008 |
| Right rib 7 | Male | Center 1 | 28946 | 263.3 | 262.2 | 1.13 | -1.424 | -1.431 | 0.006 |
| Right rib 7 | Male | Center 2 | 31404 | 262.1 | 262.2 | -0.09 | -1.412 | -1.431 | 0.019 |
| Right rib 7 | Male | Center 3 | 32541 | 263.3 | 262.2 | 1.1 | -1.41 | -1.431 | 0.02 |
| Right rib 7 | Male | Center 4 | 34638 | 261.5 | 262.2 | -0.69 | -1.419 | -1.431 | 0.012 |
| Right rib 7 | Male | Center 5 | 35416 | 262.4 | 262.2 | 0.23 | -1.425 | -1.431 | 0.005 |
| Right rib 7 | Male | Center 6 | 35296 | 260.6 | 262.2 | -1.53 | -1.463 | -1.431 | -0.032 |
| Right rib 7 | Male | Center 7 | 36817 | 262.4 | 262.2 | 0.24 | -1.442 | -1.431 | -0.012 |
| Right rib 7 | Male | Center 8 | 37231 | 262.2 | 262.2 | 0.06 | -1.449 | -1.431 | -0.018 |
| Right rib 7 | Male | Center 9 | 38711 | 262.1 | 262.2 | -0.06 | -1.433 | -1.431 | -0.002 |
| Right rib 7 | Female | Center 1 | 26818 | 322.9 | 322 | 0.82 | -5.289 | -5.29 | 0.001 |
| Right rib 7 | Female | Center 2 | 29096 | 322.2 | 322 | 0.16 | -5.235 | -5.29 | 0.055 |
| Right rib 7 | Female | Center 3 | 29901 | 323.3 | 322 | 1.25 | -5.232 | -5.29 | 0.058 |
| Right rib 7 | Female | Center 4 | 31626 | 320.9 | 322 | -1.19 | -5.323 | -5.29 | -0.033 |
| Right rib 7 | Female | Center 5 | 31678 | 322 | 322 | -0.02 | -5.274 | -5.29 | 0.015 |
| Right rib 7 | Female | Center 6 | 32301 | 320.4 | 322 | -1.61 | -5.341 | -5.29 | -0.052 |
| Right rib 7 | Female | Center 7 | 33540 | 322.7 | 322 | 0.7 | -5.32 | -5.29 | -0.03 |
| Right rib 7 | Female | Center 8 | 34215 | 322.1 | 322 | 0.06 | -5.251 | -5.29 | 0.038 |
| Right rib 7 | Female | Center 9 | 35473 | 321.9 | 322 | -0.14 | -5.282 | -5.29 | 0.008 |
| Right rib 8 | Male | Center 1 | 28925 | 278.2 | 277.2 | 1 | -1.517 | -1.548 | 0.031 |
| Right rib 8 | Male | Center 2 | 31450 | 277.3 | 277.2 | 0.12 | -1.566 | -1.548 | -0.018 |
| Right rib 8 | Male | Center 3 | 32603 | 277.7 | 277.2 | 0.5 | -1.53 | -1.548 | 0.018 |
| Right rib 8 | Male | Center 4 | 34642 | 276.9 | 277.2 | -0.27 | -1.55 | -1.548 | -0.002 |
| Right rib 8 | Male | Center 5 | 35423 | 277.5 | 277.2 | 0.26 | -1.533 | -1.548 | 0.015 |
| Right rib 8 | Male | Center 6 | 35313 | 275.5 | 277.2 | -1.67 | -1.581 | -1.548 | -0.032 |
| Right rib 8 | Male | Center 7 | 36827 | 277.5 | 277.2 | 0.29 | -1.548 | -1.548 | 0.001 |
| Right rib 8 | Male | Center 8 | 37247 | 277.3 | 277.2 | 0.12 | -1.568 | -1.548 | -0.019 |
| Right rib 8 | Male | Center 9 | 38730 | 277.2 | 277.2 | -0.03 | -1.551 | -1.548 | -0.003 |
| Right rib 8 | Female | Center 1 | 26792 | 335.5 | 334.8 | 0.67 | -5.457 | -5.553 | 0.096 |
| Right rib 8 | Female | Center 2 | 29091 | 334.8 | 334.8 | -0.02 | -5.501 | -5.553 | 0.053 |
| Right rib 8 | Female | Center 3 | 29950 | 334.8 | 334.8 | 0.05 | -5.516 | -5.553 | 0.037 |
| Right rib 8 | Female | Center 4 | 31602 | 334.1 | 334.8 | -0.68 | -5.628 | -5.553 | -0.075 |
| Right rib 8 | Female | Center 5 | 31667 | 335.6 | 334.8 | 0.77 | -5.586 | -5.553 | -0.032 |
| Right rib 8 | Female | Center 6 | 32293 | 333.1 | 334.8 | -1.73 | -5.584 | -5.553 | -0.031 |
| Right rib 8 | Female | Center 7 | 33528 | 335.6 | 334.8 | 0.82 | -5.586 | -5.553 | -0.033 |
| Right rib 8 | Female | Center 8 | 34196 | 335 | 334.8 | 0.16 | -5.521 | -5.553 | 0.033 |
| Right rib 8 | Female | Center 9 | 35465 | 334.7 | 334.8 | -0.13 | -5.545 | -5.553 | 0.008 |
| Right rib 9 | Male | Center 1 | 28888 | 294.6 | 294.1 | 0.42 | -1.381 | -1.438 | 0.056 |
| Right rib 9 | Male | Center 2 | 31507 | 294.9 | 294.1 | 0.72 | -1.478 | -1.438 | -0.04 |
| Right rib 9 | Male | Center 3 | 32627 | 294.6 | 294.1 | 0.43 | -1.402 | -1.438 | 0.035 |
| Right rib 9 | Male | Center 4 | 34668 | 294.1 | 294.1 | -0.09 | -1.444 | -1.438 | -0.006 |
| Right rib 9 | Male | Center 5 | 35463 | 294.3 | 294.1 | 0.13 | -1.414 | -1.438 | 0.023 |
| Right rib 9 | Male | Center 6 | 35349 | 292.1 | 294.1 | -2 | -1.478 | -1.438 | -0.04 |
| Right rib 9 | Male | Center 7 | 36860 | 294.5 | 294.1 | 0.4 | -1.432 | -1.438 | 0.006 |
| Right rib 9 | Male | Center 8 | 37277 | 294.4 | 294.1 | 0.24 | -1.465 | -1.438 | -0.028 |
| Right rib 9 | Male | Center 9 | 38761 | 294.1 | 294.1 | -0.01 | -1.443 | -1.438 | -0.006 |
| Right rib 9 | Female | Center 1 | 26766 | 344.4 | 344 | 0.34 | -5.465 | -5.615 | 0.15 |
| Right rib 9 | Female | Center 2 | 29150 | 344.2 | 344 | 0.18 | -5.591 | -5.615 | 0.024 |
| Right rib 9 | Female | Center 3 | 29989 | 344.4 | 344 | 0.43 | -5.561 | -5.615 | 0.053 |
| Right rib 9 | Female | Center 4 | 31630 | 343.6 | 344 | -0.37 | -5.713 | -5.615 | -0.098 |
| Right rib 9 | Female | Center 5 | 31702 | 345 | 344 | 1 | -5.55 | -5.615 | 0.065 |
| Right rib 9 | Female | Center 6 | 32329 | 342.6 | 344 | -1.46 | -5.545 | -5.615 | 0.069 |
| Right rib 9 | Female | Center 7 | 33570 | 345 | 344 | 0.94 | -5.645 | -5.615 | -0.031 |
| Right rib 9 | Female | Center 8 | 34232 | 344.6 | 344 | 0.56 | -5.516 | -5.615 | 0.098 |
| Right rib 9 | Female | Center 9 | 35504 | 343.9 | 344 | -0.13 | -5.607 | -5.615 | 0.007 |
| Right rib 10 | Male | Center 1 | 28677 | 323.7 | 323.2 | 0.5 | -1.36 | -1.451 | 0.091 |
| Right rib 10 | Male | Center 2 | 31674 | 323.5 | 323.2 | 0.29 | -1.466 | -1.451 | -0.015 |
| Right rib 10 | Male | Center 3 | 32630 | 323.7 | 323.2 | 0.47 | -1.441 | -1.451 | 0.01 |
| Right rib 10 | Male | Center 4 | 34700 | 323.3 | 323.2 | 0.09 | -1.474 | -1.451 | -0.023 |
| Right rib 10 | Male | Center 5 | 35505 | 323.6 | 323.2 | 0.37 | -1.449 | -1.451 | 0.002 |
| Right rib 10 | Male | Center 6 | 35441 | 321 | 323.2 | -2.21 | -1.479 | -1.451 | -0.028 |
| Right rib 10 | Male | Center 7 | 36901 | 323.7 | 323.2 | 0.5 | -1.453 | -1.451 | -0.002 |
| Right rib 10 | Male | Center 8 | 37293 | 323.4 | 323.2 | 0.21 | -1.47 | -1.451 | -0.019 |
| Right rib 10 | Male | Center 9 | 38787 | 323.2 | 323.2 | -0.01 | -1.456 | -1.451 | -0.005 |
| Right rib 10 | Female | Center 1 | 26685 | 379.4 | 379.3 | 0.13 | -5.792 | -5.938 | 0.146 |
| Right rib 10 | Female | Center 2 | 29282 | 379.1 | 379.3 | -0.13 | -5.899 | -5.938 | 0.039 |
| Right rib 10 | Female | Center 3 | 29970 | 379.3 | 379.3 | 0.02 | -5.853 | -5.938 | 0.085 |
| Right rib 10 | Female | Center 4 | 31673 | 378.8 | 379.3 | -0.42 | -6.019 | -5.938 | -0.081 |
| Right rib 10 | Female | Center 5 | 31779 | 380.9 | 379.3 | 1.63 | -5.9 | -5.938 | 0.038 |
| Right rib 10 | Female | Center 6 | 32436 | 377.4 | 379.3 | -1.82 | -5.94 | -5.938 | -0.002 |
| Right rib 10 | Female | Center 7 | 33655 | 380.2 | 379.3 | 1 | -5.971 | -5.938 | -0.034 |
| Right rib 10 | Female | Center 8 | 34299 | 379.6 | 379.3 | 0.33 | -5.924 | -5.938 | 0.013 |
| Right rib 10 | Female | Center 9 | 35565 | 379.1 | 379.3 | -0.14 | -5.931 | -5.938 | 0.007 |
| Right rib 11 | Male | Center 1 | 28675 | 337 | 336.5 | 0.42 | -1.891 | -1.946 | 0.055 |
| Right rib 11 | Male | Center 2 | 31717 | 337 | 336.5 | 0.43 | -1.979 | -1.946 | -0.033 |
| Right rib 11 | Male | Center 3 | 32662 | 336.7 | 336.5 | 0.19 | -1.915 | -1.946 | 0.031 |
| Right rib 11 | Male | Center 4 | 34729 | 336.5 | 336.5 | -0.08 | -1.954 | -1.946 | -0.008 |
| Right rib 11 | Male | Center 5 | 35580 | 336.9 | 336.5 | 0.34 | -1.926 | -1.946 | 0.02 |
| Right rib 11 | Male | Center 6 | 35446 | 334.5 | 336.5 | -2.04 | -1.975 | -1.946 | -0.029 |
| Right rib 11 | Male | Center 7 | 36962 | 337.3 | 336.5 | 0.75 | -1.946 | -1.946 | -0 |
| Right rib 11 | Male | Center 8 | 37312 | 336.7 | 336.5 | 0.15 | -1.971 | -1.946 | -0.025 |
| Right rib 11 | Male | Center 9 | 38829 | 336.6 | 336.5 | 0.03 | -1.951 | -1.946 | -0.005 |
| Right rib 11 | Female | Center 1 | 26645 | 383.6 | 384.7 | -1.1 | -6.473 | -6.674 | 0.202 |
| Right rib 11 | Female | Center 2 | 29364 | 384.9 | 384.7 | 0.18 | -6.684 | -6.674 | -0.01 |
| Right rib 11 | Female | Center 3 | 29977 | 384.3 | 384.7 | -0.36 | -6.691 | -6.674 | -0.016 |
| Right rib 11 | Female | Center 4 | 31697 | 384 | 384.7 | -0.75 | -6.741 | -6.674 | -0.067 |
| Right rib 11 | Female | Center 5 | 31856 | 386 | 384.7 | 1.29 | -6.701 | -6.674 | -0.027 |
| Right rib 11 | Female | Center 6 | 32418 | 383.8 | 384.7 | -0.88 | -6.679 | -6.674 | -0.004 |
| Right rib 11 | Female | Center 7 | 33679 | 386 | 384.7 | 1.25 | -6.701 | -6.674 | -0.027 |
| Right rib 11 | Female | Center 8 | 34311 | 385.2 | 384.7 | 0.52 | -6.679 | -6.674 | -0.005 |
| Right rib 11 | Female | Center 9 | 35589 | 384.5 | 384.7 | -0.18 | -6.664 | -6.674 | 0.01 |
| Right rib 12 | Male | Center 1 | 28472 | 320.6 | 319.9 | 0.71 | -1.797 | -1.838 | 0.041 |
| Right rib 12 | Male | Center 2 | 31290 | 319.6 | 319.9 | -0.36 | -1.845 | -1.838 | -0.007 |
| Right rib 12 | Male | Center 3 | 32280 | 319.5 | 319.9 | -0.39 | -1.834 | -1.838 | 0.003 |
| Right rib 12 | Male | Center 4 | 34379 | 319.7 | 319.9 | -0.22 | -1.841 | -1.838 | -0.003 |
| Right rib 12 | Male | Center 5 | 35197 | 320.3 | 319.9 | 0.35 | -1.808 | -1.838 | 0.029 |
| Right rib 12 | Male | Center 6 | 34996 | 318.7 | 319.9 | -1.18 | -1.848 | -1.838 | -0.01 |
| Right rib 12 | Male | Center 7 | 36584 | 320.6 | 319.9 | 0.7 | -1.856 | -1.838 | -0.018 |
| Right rib 12 | Male | Center 8 | 36899 | 320.3 | 319.9 | 0.39 | -1.863 | -1.838 | -0.025 |
| Right rib 12 | Male | Center 9 | 38399 | 320 | 319.9 | 0.08 | -1.845 | -1.838 | -0.007 |
| Right rib 12 | Female | Center 1 | 26152 | 370.4 | 375.2 | -4.73 | -5.366 | -6.13 | 0.765 |
| Right rib 12 | Female | Center 2 | 28717 | 374.6 | 375.2 | -0.58 | -6.084 | -6.13 | 0.047 |
| Right rib 12 | Female | Center 3 | 29371 | 374.1 | 375.2 | -1.13 | -6.164 | -6.13 | -0.034 |
| Right rib 12 | Female | Center 4 | 31081 | 374.9 | 375.2 | -0.32 | -6.187 | -6.13 | -0.057 |
| Right rib 12 | Female | Center 5 | 31191 | 377 | 375.2 | 1.84 | -6.206 | -6.13 | -0.076 |
| Right rib 12 | Female | Center 6 | 31769 | 374.4 | 375.2 | -0.82 | -6.021 | -6.13 | 0.11 |
| Right rib 12 | Female | Center 7 | 33018 | 376.4 | 375.2 | 1.27 | -6.159 | -6.13 | -0.029 |
| Right rib 12 | Female | Center 8 | 33615 | 375.7 | 375.2 | 0.54 | -6.126 | -6.13 | 0.004 |
| Right rib 12 | Female | Center 9 | 34870 | 375.2 | 375.2 | 0 | -6.135 | -6.13 | -0.005 |

Each trajectory was refitted with one center omitted. Differences are the refitted value minus the value from all centers.

**Supplementary Table 10: Body-mass-index-adjusted models**

| **Site** | **Sex** | **Patients with a body-mass index, n** | **Attenuation per unit body-mass index, HU (95% CI)** | **Change 45–60 years, unadjusted, HU/year** | **Change 45–60 years, adjusted for body-mass index, HU/year** |
| --- | --- | --- | --- | --- | --- |
| C1 | Male | 8027 | 3.10 (2.65 to 3.56) | -0.478 | -0.303 |
| C1 | Female | 8060 | 1.84 (1.46 to 2.21) | -3.986 | -3.938 |
| C2 | Male | 8041 | 4.18 (3.74 to 4.63) | -1.373 | -1.14 |
| C2 | Female | 8069 | 2.12 (1.76 to 2.48) | -5.219 | -5.166 |
| C3 | Male | 8772 | 3.58 (3.13 to 4.02) | -0.511 | -0.307 |
| C3 | Female | 8755 | 1.03 (0.69 to 1.37) | -4.645 | -4.617 |
| C4 | Male | 9928 | 2.84 (2.42 to 3.26) | -0.201 | -0.043 |
| C4 | Female | 9550 | 0.42 (0.09 to 0.75) | -4.14 | -4.128 |
| C5 | Male | 12556 | 1.53 (1.17 to 1.89) | -0.156 | -0.081 |
| C5 | Female | 11395 | -0.18 (-0.47 to 0.11) | -4.003 | -4.008 |
| C6 | Male | 16893 | 1.16 (0.88 to 1.45) | -0.033 | 0.019 |
| C6 | Female | 14764 | -0.29 (-0.52 to -0.05) | -3.644 | -3.652 |
| C7 | Male | 18753 | 1.75 (1.50 to 1.99) | 0.038 | 0.113 |
| C7 | Female | 16264 | 0.23 (0.04 to 0.43) | -3.172 | -3.166 |
| T1 | Male | 18853 | 2.69 (2.48 to 2.89) | -0.69 | -0.572 |
| T1 | Female | 16387 | 1.12 (0.95 to 1.28) | -3.301 | -3.272 |
| T2 | Male | 18834 | 2.85 (2.65 to 3.05) | -0.607 | -0.481 |
| T2 | Female | 16377 | 1.22 (1.06 to 1.38) | -2.994 | -2.961 |
| T3 | Male | 18839 | 3.06 (2.85 to 3.26) | -0.711 | -0.578 |
| T3 | Female | 16389 | 1.44 (1.27 to 1.61) | -3.106 | -3.074 |
| T4 | Male | 18860 | 3.07 (2.86 to 3.27) | -0.737 | -0.604 |
| T4 | Female | 16427 | 1.49 (1.32 to 1.66) | -3.543 | -3.519 |
| T5 | Male | 18899 | 2.99 (2.79 to 3.19) | -0.66 | -0.532 |
| T5 | Female | 16437 | 1.40 (1.24 to 1.56) | -3.109 | -3.072 |
| T6 | Male | 19301 | 2.84 (2.64 to 3.03) | -0.67 | -0.537 |
| T6 | Female | 16778 | 1.30 (1.14 to 1.45) | -2.998 | -2.96 |
| T7 | Male | 20757 | 2.82 (2.64 to 3.01) | -0.65 | -0.53 |
| T7 | Female | 18054 | 1.43 (1.28 to 1.57) | -3.347 | -3.302 |
| T8 | Male | 22889 | 2.93 (2.75 to 3.10) | -0.586 | -0.462 |
| T8 | Female | 19860 | 1.63 (1.49 to 1.77) | -2.935 | -2.889 |
| T9 | Male | 24646 | 2.93 (2.75 to 3.10) | -0.606 | -0.496 |
| T9 | Female | 21562 | 1.63 (1.50 to 1.77) | -3.507 | -3.474 |
| T10 | Male | 25498 | 2.51 (2.34 to 2.68) | -0.684 | -0.587 |
| T10 | Female | 22509 | 1.27 (1.13 to 1.40) | -3.574 | -3.548 |
| T11 | Male | 25696 | 2.07 (1.91 to 2.23) | -0.845 | -0.776 |
| T11 | Female | 22724 | 0.93 (0.80 to 1.07) | -3.575 | -3.557 |
| T12 | Male | 25686 | 1.90 (1.74 to 2.05) | -0.975 | -0.91 |
| T12 | Female | 22666 | 0.84 (0.72 to 0.97) | -3.509 | -3.491 |
| L1 | Male | 25730 | 2.11 (1.96 to 2.26) | -0.87 | -0.79 |
| L1 | Female | 22686 | 0.96 (0.84 to 1.08) | -3.427 | -3.409 |
| L2 | Male | 25766 | 1.95 (1.79 to 2.11) | -0.633 | -0.559 |
| L2 | Female | 22693 | 0.85 (0.72 to 0.98) | -3.455 | -3.434 |
| L3 | Male | 25764 | 1.92 (1.75 to 2.08) | -0.675 | -0.603 |
| L3 | Female | 22699 | 0.74 (0.61 to 0.87) | -3.623 | -3.605 |
| L4 | Male | 25765 | 2.05 (1.87 to 2.22) | -0.598 | -0.533 |
| L4 | Female | 22697 | 0.77 (0.63 to 0.91) | -3.622 | -3.603 |
| L5 | Male | 25748 | 2.48 (2.30 to 2.67) | -0.217 | -0.127 |
| L5 | Female | 22683 | 1.14 (0.99 to 1.28) | -3.145 | -3.116 |
| S1 | Male | 25734 | 2.97 (2.79 to 3.16) | -1.276 | -1.176 |
| S1 | Female | 22719 | 1.36 (1.21 to 1.52) | -4.006 | -3.982 |
| Left rib 1 | Male | 18884 | 0.41 (0.17 to 0.65) | -1.912 | -1.894 |
| Left rib 1 | Female | 16432 | -1.10 (-1.28 to -0.91) | -4.397 | -4.427 |
| Left rib 2 | Male | 18923 | 1.23 (1.01 to 1.44) | -1.235 | -1.186 |
| Left rib 2 | Female | 16491 | -0.61 (-0.79 to -0.43) | -3.997 | -4.01 |
| Left rib 3 | Male | 19257 | 0.49 (0.28 to 0.69) | -1.815 | -1.797 |
| Left rib 3 | Female | 16948 | -1.07 (-1.26 to -0.88) | -4.805 | -4.826 |
| Left rib 4 | Male | 21421 | 0.39 (0.17 to 0.60) | -1.733 | -1.72 |
| Left rib 4 | Female | 19250 | -1.18 (-1.38 to -0.98) | -5.296 | -5.323 |
| Left rib 5 | Male | 24466 | 0.38 (0.17 to 0.59) | -1.516 | -1.504 |
| Left rib 5 | Female | 21912 | -1.13 (-1.32 to -0.93) | -5.191 | -5.213 |
| Left rib 6 | Male | 25745 | 0.37 (0.16 to 0.59) | -1.395 | -1.382 |
| Left rib 6 | Female | 22847 | -1.00 (-1.20 to -0.80) | -5.046 | -5.064 |
| Left rib 7 | Male | 25813 | 0.79 (0.58 to 1.00) | -1.432 | -1.406 |
| Left rib 7 | Female | 22860 | -0.75 (-0.95 to -0.55) | -5.463 | -5.475 |
| Left rib 8 | Male | 25841 | 2.27 (2.05 to 2.50) | -1.585 | -1.509 |
| Left rib 8 | Female | 22876 | 0.81 (0.60 to 1.01) | -5.937 | -5.925 |
| Left rib 9 | Male | 25826 | 3.47 (3.23 to 3.71) | -1.437 | -1.321 |
| Left rib 9 | Female | 22870 | 1.76 (1.55 to 1.97) | -5.867 | -5.839 |
| Left rib 10 | Male | 25718 | 3.55 (3.30 to 3.79) | -1.509 | -1.383 |
| Left rib 10 | Female | 22775 | 1.06 (0.85 to 1.27) | -6.034 | -6.014 |
| Left rib 11 | Male | 25698 | 2.38 (2.14 to 2.62) | -1.974 | -1.889 |
| Left rib 11 | Female | 22763 | -0.01 (-0.22 to 0.20) | -6.683 | -6.684 |
| Left rib 12 | Male | 25477 | 1.53 (1.28 to 1.77) | -1.982 | -1.939 |
| Left rib 12 | Female | 21981 | -0.74 (-0.97 to -0.51) | -5.705 | -5.716 |
| Right rib 1 | Male | 18884 | 0.51 (0.27 to 0.75) | -1.919 | -1.896 |
| Right rib 1 | Female | 16449 | -0.98 (-1.16 to -0.80) | -4.436 | -4.463 |
| Right rib 2 | Male | 18931 | 1.27 (1.05 to 1.48) | -1.242 | -1.19 |
| Right rib 2 | Female | 16527 | -0.66 (-0.84 to -0.48) | -4.083 | -4.096 |
| Right rib 3 | Male | 19218 | 0.46 (0.26 to 0.67) | -1.857 | -1.839 |
| Right rib 3 | Female | 16926 | -1.09 (-1.28 to -0.90) | -4.822 | -4.843 |
| Right rib 4 | Male | 21307 | 0.47 (0.26 to 0.69) | -1.758 | -1.741 |
| Right rib 4 | Female | 19163 | -1.30 (-1.50 to -1.09) | -5.304 | -5.335 |
| Right rib 5 | Male | 24361 | 0.33 (0.12 to 0.54) | -1.431 | -1.421 |
| Right rib 5 | Female | 21821 | -1.19 (-1.40 to -0.99) | -4.9 | -4.921 |
| Right rib 6 | Male | 25755 | 0.47 (0.26 to 0.69) | -1.44 | -1.424 |
| Right rib 6 | Female | 22844 | -0.84 (-1.04 to -0.64) | -5.069 | -5.083 |
| Right rib 7 | Male | 25837 | 0.89 (0.68 to 1.10) | -1.451 | -1.42 |
| Right rib 7 | Female | 22890 | -0.43 (-0.64 to -0.23) | -5.328 | -5.335 |
| Right rib 8 | Male | 25866 | 2.42 (2.21 to 2.65) | -1.492 | -1.411 |
| Right rib 8 | Female | 22897 | 0.97 (0.76 to 1.18) | -5.493 | -5.475 |
| Right rib 9 | Male | 25831 | 3.53 (3.30 to 3.77) | -1.364 | -1.243 |
| Right rib 9 | Female | 22893 | 1.83 (1.62 to 2.05) | -5.485 | -5.451 |
| Right rib 10 | Male | 25727 | 3.56 (3.32 to 3.81) | -1.345 | -1.214 |
| Right rib 10 | Female | 22805 | 1.15 (0.94 to 1.36) | -5.618 | -5.596 |
| Right rib 11 | Male | 25682 | 2.37 (2.13 to 2.61) | -1.817 | -1.732 |
| Right rib 11 | Female | 22744 | -0.12 (-0.34 to 0.09) | -6.337 | -6.34 |
| Right rib 12 | Male | 25481 | 1.42 (1.17 to 1.66) | -1.785 | -1.743 |
| Right rib 12 | Female | 22328 | -0.72 (-0.95 to -0.50) | -5.47 | -5.483 |

Restricted to patients with height and weight recorded. Body-mass index was centered within each site and sex, so the adjusted rate of change is directly comparable with the unadjusted one.

**Supplementary Table 11: Dependence of mean attenuation on segmented volume**

| **Site** | **Sex** | **n** | **Age-adjusted correlation of attenuation with volume (95% CI)** |
| --- | --- | --- | --- |
| C1 | Male | 12941 | -0.107 (-0.124 to -0.090) |
| C1 | Female | 13354 | -0.103 (-0.119 to -0.086) |
| C2 | Male | 12939 | -0.079 (-0.096 to -0.062) |
| C2 | Female | 13349 | -0.107 (-0.124 to -0.090) |
| C3 | Male | 13945 | 0.070 (0.053 to 0.086) |
| C3 | Female | 14293 | 0.045 (0.029 to 0.061) |
| C4 | Male | 15517 | 0.122 (0.106 to 0.137) |
| C4 | Female | 15355 | 0.068 (0.053 to 0.084) |
| C5 | Male | 19024 | 0.082 (0.067 to 0.096) |
| C5 | Female | 17881 | 0.032 (0.017 to 0.046) |
| C6 | Male | 25074 | 0.082 (0.069 to 0.094) |
| C6 | Female | 22603 | 0.031 (0.018 to 0.044) |
| C7 | Male | 28177 | 0.023 (0.012 to 0.035) |
| C7 | Female | 25131 | -0.028 (-0.040 to -0.015) |
| T1 | Male | 28549 | -0.081 (-0.093 to -0.070) |
| T1 | Female | 25491 | -0.105 (-0.117 to -0.092) |
| T2 | Male | 28532 | -0.049 (-0.060 to -0.037) |
| T2 | Female | 25469 | -0.088 (-0.100 to -0.076) |
| T3 | Male | 28568 | -0.034 (-0.046 to -0.023) |
| T3 | Female | 25483 | -0.091 (-0.103 to -0.078) |
| T4 | Male | 28569 | -0.048 (-0.059 to -0.036) |
| T4 | Female | 25485 | -0.127 (-0.139 to -0.114) |
| T5 | Male | 28635 | -0.024 (-0.035 to -0.012) |
| T5 | Female | 25546 | -0.103 (-0.115 to -0.091) |
| T6 | Male | 29225 | -0.024 (-0.036 to -0.013) |
| T6 | Female | 26056 | -0.081 (-0.093 to -0.069) |
| T7 | Male | 31300 | -0.051 (-0.062 to -0.040) |
| T7 | Female | 27854 | -0.116 (-0.127 to -0.104) |
| T8 | Male | 34324 | -0.034 (-0.044 to -0.023) |
| T8 | Female | 30457 | -0.093 (-0.105 to -0.082) |
| T9 | Male | 37013 | -0.022 (-0.032 to -0.011) |
| T9 | Female | 33251 | -0.091 (-0.102 to -0.081) |
| T10 | Male | 38463 | -0.047 (-0.057 to -0.037) |
| T10 | Female | 35011 | -0.107 (-0.118 to -0.097) |
| T11 | Male | 38907 | -0.064 (-0.074 to -0.054) |
| T11 | Female | 35559 | -0.142 (-0.152 to -0.131) |
| T12 | Male | 38876 | -0.054 (-0.063 to -0.044) |
| T12 | Female | 35495 | -0.134 (-0.144 to -0.124) |
| L1 | Male | 38886 | -0.000 (-0.010 to 0.010) |
| L1 | Female | 35498 | -0.108 (-0.118 to -0.098) |
| L2 | Male | 38881 | 0.029 (0.020 to 0.039) |
| L2 | Female | 35527 | -0.071 (-0.081 to -0.061) |
| L3 | Male | 38881 | 0.013 (0.003 to 0.023) |
| L3 | Female | 35508 | -0.059 (-0.069 to -0.049) |
| L4 | Male | 38879 | 0.040 (0.030 to 0.049) |
| L4 | Female | 35508 | -0.036 (-0.046 to -0.025) |
| L5 | Male | 38902 | 0.011 (0.001 to 0.021) |
| L5 | Female | 35508 | -0.059 (-0.070 to -0.049) |
| S1 | Male | 38908 | -0.062 (-0.071 to -0.052) |
| S1 | Female | 35559 | -0.111 (-0.121 to -0.101) |
| Left rib 1 | Male | 28579 | 0.304 (0.294 to 0.315) |
| Left rib 1 | Female | 25521 | 0.186 (0.174 to 0.198) |
| Left rib 2 | Male | 28631 | 0.121 (0.110 to 0.133) |
| Left rib 2 | Female | 25557 | 0.070 (0.058 to 0.082) |
| Left rib 3 | Male | 29200 | 0.099 (0.088 to 0.111) |
| Left rib 3 | Female | 26333 | 0.020 (0.008 to 0.032) |
| Left rib 4 | Male | 32094 | 0.083 (0.072 to 0.094) |
| Left rib 4 | Female | 29542 | 0.143 (0.132 to 0.154) |
| Left rib 5 | Male | 36632 | 0.198 (0.188 to 0.208) |
| Left rib 5 | Female | 33829 | 0.310 (0.301 to 0.320) |
| Left rib 6 | Male | 38749 | 0.252 (0.243 to 0.262) |
| Left rib 6 | Female | 35514 | 0.337 (0.327 to 0.346) |
| Left rib 7 | Male | 38883 | 0.107 (0.098 to 0.117) |
| Left rib 7 | Female | 35584 | 0.117 (0.106 to 0.127) |
| Left rib 8 | Male | 38911 | -0.006 (-0.016 to 0.004) |
| Left rib 8 | Female | 35590 | -0.017 (-0.027 to -0.007) |
| Left rib 9 | Male | 38937 | -0.028 (-0.038 to -0.018) |
| Left rib 9 | Female | 35619 | -0.056 (-0.066 to -0.045) |
| Left rib 10 | Male | 38979 | -0.051 (-0.061 to -0.041) |
| Left rib 10 | Female | 35656 | -0.126 (-0.137 to -0.116) |
| Left rib 11 | Male | 38973 | -0.043 (-0.053 to -0.033) |
| Left rib 11 | Female | 35672 | -0.084 (-0.094 to -0.073) |
| Left rib 12 | Male | 38542 | 0.109 (0.099 to 0.118) |
| Left rib 12 | Female | 34465 | 0.098 (0.088 to 0.109) |
| Right rib 1 | Male | 28562 | 0.285 (0.274 to 0.296) |
| Right rib 1 | Female | 25496 | 0.190 (0.178 to 0.202) |
| Right rib 2 | Male | 28622 | 0.106 (0.095 to 0.118) |
| Right rib 2 | Female | 25591 | 0.065 (0.052 to 0.077) |
| Right rib 3 | Male | 29164 | 0.057 (0.046 to 0.069) |
| Right rib 3 | Female | 26297 | -0.006 (-0.018 to 0.006) |
| Right rib 4 | Male | 31936 | 0.111 (0.100 to 0.122) |
| Right rib 4 | Female | 29371 | 0.168 (0.157 to 0.179) |
| Right rib 5 | Male | 36386 | 0.229 (0.219 to 0.239) |
| Right rib 5 | Female | 33600 | 0.375 (0.366 to 0.385) |
| Right rib 6 | Male | 38734 | 0.266 (0.257 to 0.275) |
| Right rib 6 | Female | 35489 | 0.348 (0.339 to 0.357) |
| Right rib 7 | Male | 38875 | 0.133 (0.123 to 0.143) |
| Right rib 7 | Female | 35581 | 0.143 (0.132 to 0.153) |
| Right rib 8 | Male | 38895 | 0.006 (-0.004 to 0.016) |
| Right rib 8 | Female | 35573 | -0.009 (-0.019 to 0.002) |
| Right rib 9 | Male | 38925 | -0.049 (-0.059 to -0.040) |
| Right rib 9 | Female | 35609 | -0.085 (-0.095 to -0.075) |
| Right rib 10 | Male | 38951 | -0.060 (-0.070 to -0.050) |
| Right rib 10 | Female | 35668 | -0.117 (-0.127 to -0.106) |
| Right rib 11 | Male | 38989 | -0.038 (-0.048 to -0.028) |
| Right rib 11 | Female | 35692 | -0.077 (-0.088 to -0.067) |
| Right rib 12 | Male | 38562 | 0.077 (0.067 to 0.087) |
| Right rib 12 | Female | 34973 | 0.063 (0.053 to 0.074) |

Pearson correlation between attenuation and segmented volume after removing a spline in age from both. Values near zero indicate that quality control removed the volume dependence introduced by partially covered structures.

**Supplementary Table 12: Association with L1 and surrogacy, all sites (Correlation with L1)**

| **Site** | **Sex** | **Region** | **n** | **Correlation with L1, unadjusted** | **Unadjusted, lower 95% limit** | **Unadjusted, upper 95% limit** | **Unadjusted, p** | **Unadjusted, p (FDR)** | **Correlation with L1, age-adjusted** | **Age-adjusted, lower 95% limit** | **Age-adjusted, upper 95% limit** | **Age-adjusted, p** | **Age-adjusted, p (FDR)** |
| --- | --- | --- | --- | --- | --- | --- | --- | --- | --- | --- | --- | --- | --- |
| C1 | Female | Cervical vertebrae | 12745 | 0.7056 | 0.6967 | 0.7142 | <0.0001 | <0.0001 | 0.598 | 0.5867 | 0.609 | <0.0001 | <0.0001 |
| C2 | Female | Cervical vertebrae | 12817 | 0.7738 | 0.7668 | 0.7807 | <0.0001 | <0.0001 | 0.6657 | 0.656 | 0.6753 | <0.0001 | <0.0001 |
| C3 | Female | Cervical vertebrae | 13708 | 0.7556 | 0.7483 | 0.7627 | <0.0001 | <0.0001 | 0.6561 | 0.6465 | 0.6656 | <0.0001 | <0.0001 |
| C4 | Female | Cervical vertebrae | 14720 | 0.7474 | 0.7402 | 0.7545 | <0.0001 | <0.0001 | 0.655 | 0.6457 | 0.6642 | <0.0001 | <0.0001 |
| C5 | Female | Cervical vertebrae | 17139 | 0.7439 | 0.7371 | 0.7505 | <0.0001 | <0.0001 | 0.6483 | 0.6395 | 0.6569 | <0.0001 | <0.0001 |
| C6 | Female | Cervical vertebrae | 21607 | 0.7564 | 0.7506 | 0.762 | <0.0001 | <0.0001 | 0.6644 | 0.6568 | 0.6717 | <0.0001 | <0.0001 |
| C7 | Female | Cervical vertebrae | 24178 | 0.7735 | 0.7683 | 0.7785 | <0.0001 | <0.0001 | 0.6894 | 0.6827 | 0.6959 | <0.0001 | <0.0001 |
| T1 | Female | Thoracic vertebrae | 24597 | 0.8201 | 0.816 | 0.8242 | <0.0001 | <0.0001 | 0.7386 | 0.7328 | 0.7442 | <0.0001 | <0.0001 |
| T2 | Female | Thoracic vertebrae | 24621 | 0.8326 | 0.8287 | 0.8364 | <0.0001 | <0.0001 | 0.758 | 0.7526 | 0.7633 | <0.0001 | <0.0001 |
| T3 | Female | Thoracic vertebrae | 24623 | 0.8345 | 0.8307 | 0.8383 | <0.0001 | <0.0001 | 0.759 | 0.7536 | 0.7642 | <0.0001 | <0.0001 |
| T4 | Female | Thoracic vertebrae | 24615 | 0.8352 | 0.8314 | 0.8389 | <0.0001 | <0.0001 | 0.7586 | 0.7533 | 0.7639 | <0.0001 | <0.0001 |
| T5 | Female | Thoracic vertebrae | 24721 | 0.8374 | 0.8336 | 0.841 | <0.0001 | <0.0001 | 0.7605 | 0.7552 | 0.7658 | <0.0001 | <0.0001 |
| T6 | Female | Thoracic vertebrae | 25034 | 0.8377 | 0.8339 | 0.8413 | <0.0001 | <0.0001 | 0.7622 | 0.7569 | 0.7673 | <0.0001 | <0.0001 |
| T7 | Female | Thoracic vertebrae | 26509 | 0.8378 | 0.8341 | 0.8413 | <0.0001 | <0.0001 | 0.7628 | 0.7577 | 0.7678 | <0.0001 | <0.0001 |
| T8 | Female | Thoracic vertebrae | 28900 | 0.8455 | 0.8422 | 0.8488 | <0.0001 | <0.0001 | 0.7724 | 0.7677 | 0.777 | <0.0001 | <0.0001 |
| T9 | Female | Thoracic vertebrae | 31583 | 0.8593 | 0.8564 | 0.8622 | <0.0001 | <0.0001 | 0.7895 | 0.7853 | 0.7936 | <0.0001 | <0.0001 |
| T10 | Female | Thoracic vertebrae | 33715 | 0.8748 | 0.8723 | 0.8773 | <0.0001 | <0.0001 | 0.8089 | 0.8052 | 0.8126 | <0.0001 | <0.0001 |
| T11 | Female | Thoracic vertebrae | 34403 | 0.8811 | 0.8787 | 0.8835 | <0.0001 | <0.0001 | 0.8172 | 0.8136 | 0.8207 | <0.0001 | <0.0001 |
| T12 | Female | Thoracic vertebrae | 34530 | 0.9002 | 0.8982 | 0.9022 | <0.0001 | <0.0001 | 0.8445 | 0.8414 | 0.8475 | <0.0001 | <0.0001 |
| L2 | Female | Lumbar vertebrae | 34673 | 0.9 | 0.898 | 0.902 | <0.0001 | <0.0001 | 0.8485 | 0.8456 | 0.8515 | <0.0001 | <0.0001 |
| L3 | Female | Lumbar vertebrae | 34386 | 0.8694 | 0.8668 | 0.872 | <0.0001 | <0.0001 | 0.7975 | 0.7936 | 0.8013 | <0.0001 | <0.0001 |
| L4 | Female | Lumbar vertebrae | 34305 | 0.8262 | 0.8228 | 0.8295 | <0.0001 | <0.0001 | 0.7387 | 0.7338 | 0.7434 | <0.0001 | <0.0001 |
| L5 | Female | Lumbar vertebrae | 34344 | 0.795 | 0.791 | 0.7988 | <0.0001 | <0.0001 | 0.702 | 0.6966 | 0.7073 | <0.0001 | <0.0001 |
| S1 | Female | Sacrum | 34062 | 0.8076 | 0.8039 | 0.8112 | <0.0001 | <0.0001 | 0.7089 | 0.7036 | 0.7141 | <0.0001 | <0.0001 |
| Left rib 1 | Female | Left ribs | 24409 | 0.6702 | 0.6633 | 0.6771 | <0.0001 | <0.0001 | 0.4972 | 0.4877 | 0.5066 | <0.0001 | <0.0001 |
| Left rib 2 | Female | Left ribs | 24393 | 0.7404 | 0.7347 | 0.746 | <0.0001 | <0.0001 | 0.5923 | 0.5841 | 0.6004 | <0.0001 | <0.0001 |
| Left rib 3 | Female | Left ribs | 25060 | 0.7347 | 0.7289 | 0.7403 | <0.0001 | <0.0001 | 0.5748 | 0.5664 | 0.583 | <0.0001 | <0.0001 |
| Left rib 4 | Female | Left ribs | 27959 | 0.6963 | 0.6902 | 0.7023 | <0.0001 | <0.0001 | 0.5266 | 0.5181 | 0.5351 | <0.0001 | <0.0001 |
| Left rib 5 | Female | Left ribs | 31989 | 0.6484 | 0.642 | 0.6547 | <0.0001 | <0.0001 | 0.4735 | 0.465 | 0.482 | <0.0001 | <0.0001 |
| Left rib 6 | Female | Left ribs | 33607 | 0.6364 | 0.63 | 0.6427 | <0.0001 | <0.0001 | 0.4647 | 0.4563 | 0.473 | <0.0001 | <0.0001 |
| Left rib 7 | Female | Left ribs | 33712 | 0.6806 | 0.6748 | 0.6862 | <0.0001 | <0.0001 | 0.5093 | 0.5013 | 0.5171 | <0.0001 | <0.0001 |
| Left rib 8 | Female | Left ribs | 33724 | 0.717 | 0.7118 | 0.7221 | <0.0001 | <0.0001 | 0.5446 | 0.5371 | 0.5521 | <0.0001 | <0.0001 |
| Left rib 9 | Female | Left ribs | 33763 | 0.7267 | 0.7217 | 0.7317 | <0.0001 | <0.0001 | 0.5597 | 0.5524 | 0.567 | <0.0001 | <0.0001 |
| Left rib 10 | Female | Left ribs | 34020 | 0.7334 | 0.7285 | 0.7383 | <0.0001 | <0.0001 | 0.5661 | 0.5589 | 0.5733 | <0.0001 | <0.0001 |
| Left rib 11 | Female | Left ribs | 34023 | 0.7236 | 0.7185 | 0.7286 | <0.0001 | <0.0001 | 0.5391 | 0.5315 | 0.5466 | <0.0001 | <0.0001 |
| Left rib 12 | Female | Left ribs | 32899 | 0.677 | 0.6711 | 0.6828 | <0.0001 | <0.0001 | 0.4835 | 0.4752 | 0.4918 | <0.0001 | <0.0001 |
| Right rib 1 | Female | Right ribs | 24417 | 0.6842 | 0.6775 | 0.6908 | <0.0001 | <0.0001 | 0.513 | 0.5037 | 0.5221 | <0.0001 | <0.0001 |
| Right rib 2 | Female | Right ribs | 24438 | 0.7404 | 0.7347 | 0.746 | <0.0001 | <0.0001 | 0.5904 | 0.5822 | 0.5985 | <0.0001 | <0.0001 |
| Right rib 3 | Female | Right ribs | 25034 | 0.7336 | 0.7278 | 0.7392 | <0.0001 | <0.0001 | 0.5728 | 0.5644 | 0.5811 | <0.0001 | <0.0001 |
| Right rib 4 | Female | Right ribs | 27793 | 0.6878 | 0.6816 | 0.694 | <0.0001 | <0.0001 | 0.5165 | 0.5078 | 0.525 | <0.0001 | <0.0001 |
| Right rib 5 | Female | Right ribs | 31756 | 0.6256 | 0.6189 | 0.6323 | <0.0001 | <0.0001 | 0.455 | 0.4462 | 0.4636 | <0.0001 | <0.0001 |
| Right rib 6 | Female | Right ribs | 33578 | 0.6487 | 0.6425 | 0.6548 | <0.0001 | <0.0001 | 0.4784 | 0.4701 | 0.4866 | <0.0001 | <0.0001 |
| Right rib 7 | Female | Right ribs | 33704 | 0.6835 | 0.6778 | 0.6892 | <0.0001 | <0.0001 | 0.514 | 0.5061 | 0.5219 | <0.0001 | <0.0001 |
| Right rib 8 | Female | Right ribs | 33703 | 0.7113 | 0.706 | 0.7165 | <0.0001 | <0.0001 | 0.5437 | 0.5361 | 0.5512 | <0.0001 | <0.0001 |
| Right rib 9 | Female | Right ribs | 33773 | 0.7225 | 0.7174 | 0.7276 | <0.0001 | <0.0001 | 0.5629 | 0.5556 | 0.5701 | <0.0001 | <0.0001 |
| Right rib 10 | Female | Right ribs | 33999 | 0.7266 | 0.7216 | 0.7316 | <0.0001 | <0.0001 | 0.5639 | 0.5566 | 0.5711 | <0.0001 | <0.0001 |
| Right rib 11 | Female | Right ribs | 34056 | 0.7201 | 0.7149 | 0.7252 | <0.0001 | <0.0001 | 0.5372 | 0.5296 | 0.5447 | <0.0001 | <0.0001 |
| Right rib 12 | Female | Right ribs | 33384 | 0.6595 | 0.6534 | 0.6655 | <0.0001 | <0.0001 | 0.4685 | 0.4601 | 0.4769 | <0.0001 | <0.0001 |
| C1 | Male | Cervical vertebrae | 12331 | 0.6257 | 0.6149 | 0.6363 | <0.0001 | <0.0001 | 0.6372 | 0.6266 | 0.6476 | <0.0001 | <0.0001 |
| C2 | Male | Cervical vertebrae | 12424 | 0.7095 | 0.7007 | 0.7182 | <0.0001 | <0.0001 | 0.7049 | 0.6959 | 0.7136 | <0.0001 | <0.0001 |
| C3 | Male | Cervical vertebrae | 13383 | 0.6867 | 0.6776 | 0.6955 | <0.0001 | <0.0001 | 0.6923 | 0.6834 | 0.701 | <0.0001 | <0.0001 |
| C4 | Male | Cervical vertebrae | 14912 | 0.6719 | 0.663 | 0.6806 | <0.0001 | <0.0001 | 0.6826 | 0.674 | 0.6911 | <0.0001 | <0.0001 |
| C5 | Male | Cervical vertebrae | 18234 | 0.6731 | 0.6651 | 0.681 | <0.0001 | <0.0001 | 0.68 | 0.6721 | 0.6877 | <0.0001 | <0.0001 |
| C6 | Male | Cervical vertebrae | 23964 | 0.6794 | 0.6725 | 0.6861 | <0.0001 | <0.0001 | 0.6862 | 0.6794 | 0.6928 | <0.0001 | <0.0001 |
| C7 | Male | Cervical vertebrae | 27196 | 0.7056 | 0.6996 | 0.7116 | <0.0001 | <0.0001 | 0.7149 | 0.7091 | 0.7207 | <0.0001 | <0.0001 |
| T1 | Male | Thoracic vertebrae | 27683 | 0.769 | 0.7641 | 0.7738 | <0.0001 | <0.0001 | 0.7665 | 0.7616 | 0.7713 | <0.0001 | <0.0001 |
| T2 | Male | Thoracic vertebrae | 27707 | 0.7897 | 0.7852 | 0.794 | <0.0001 | <0.0001 | 0.7865 | 0.7819 | 0.7909 | <0.0001 | <0.0001 |
| T3 | Male | Thoracic vertebrae | 27753 | 0.7964 | 0.7921 | 0.8007 | <0.0001 | <0.0001 | 0.793 | 0.7886 | 0.7973 | <0.0001 | <0.0001 |
| T4 | Male | Thoracic vertebrae | 27764 | 0.8007 | 0.7965 | 0.8049 | <0.0001 | <0.0001 | 0.7973 | 0.793 | 0.8015 | <0.0001 | <0.0001 |
| T5 | Male | Thoracic vertebrae | 27813 | 0.8009 | 0.7967 | 0.8051 | <0.0001 | <0.0001 | 0.7975 | 0.7932 | 0.8018 | <0.0001 | <0.0001 |
| T6 | Male | Thoracic vertebrae | 28293 | 0.7996 | 0.7953 | 0.8037 | <0.0001 | <0.0001 | 0.7961 | 0.7918 | 0.8003 | <0.0001 | <0.0001 |
| T7 | Male | Thoracic vertebrae | 29856 | 0.8014 | 0.7973 | 0.8054 | <0.0001 | <0.0001 | 0.7977 | 0.7936 | 0.8018 | <0.0001 | <0.0001 |
| T8 | Male | Thoracic vertebrae | 32624 | 0.8055 | 0.8017 | 0.8093 | <0.0001 | <0.0001 | 0.8018 | 0.7979 | 0.8056 | <0.0001 | <0.0001 |
| T9 | Male | Thoracic vertebrae | 35315 | 0.8202 | 0.8167 | 0.8236 | <0.0001 | <0.0001 | 0.8161 | 0.8126 | 0.8196 | <0.0001 | <0.0001 |
| T10 | Male | Thoracic vertebrae | 37231 | 0.8338 | 0.8306 | 0.8368 | <0.0001 | <0.0001 | 0.8282 | 0.825 | 0.8313 | <0.0001 | <0.0001 |
| T11 | Male | Thoracic vertebrae | 37844 | 0.8453 | 0.8424 | 0.8481 | <0.0001 | <0.0001 | 0.838 | 0.835 | 0.841 | <0.0001 | <0.0001 |
| T12 | Male | Thoracic vertebrae | 38052 | 0.8849 | 0.8827 | 0.8871 | <0.0001 | <0.0001 | 0.8788 | 0.8765 | 0.8811 | <0.0001 | <0.0001 |
| L2 | Male | Lumbar vertebrae | 38069 | 0.8837 | 0.8815 | 0.8859 | <0.0001 | <0.0001 | 0.8799 | 0.8776 | 0.8822 | <0.0001 | <0.0001 |
| L3 | Male | Lumbar vertebrae | 37809 | 0.8323 | 0.8292 | 0.8354 | <0.0001 | <0.0001 | 0.8258 | 0.8226 | 0.829 | <0.0001 | <0.0001 |
| L4 | Male | Lumbar vertebrae | 37766 | 0.7765 | 0.7725 | 0.7805 | <0.0001 | <0.0001 | 0.7731 | 0.769 | 0.7771 | <0.0001 | <0.0001 |
| L5 | Male | Lumbar vertebrae | 37782 | 0.7369 | 0.7323 | 0.7415 | <0.0001 | <0.0001 | 0.7358 | 0.7312 | 0.7404 | <0.0001 | <0.0001 |
| S1 | Male | Sacrum | 37395 | 0.7591 | 0.7548 | 0.7634 | <0.0001 | <0.0001 | 0.7515 | 0.747 | 0.7559 | <0.0001 | <0.0001 |
| Left rib 1 | Male | Left ribs | 27448 | 0.5369 | 0.5284 | 0.5453 | <0.0001 | <0.0001 | 0.5254 | 0.5167 | 0.5339 | <0.0001 | <0.0001 |
| Left rib 2 | Male | Left ribs | 27478 | 0.644 | 0.637 | 0.6509 | <0.0001 | <0.0001 | 0.6238 | 0.6165 | 0.631 | <0.0001 | <0.0001 |
| Left rib 3 | Male | Left ribs | 27937 | 0.6484 | 0.6416 | 0.6551 | <0.0001 | <0.0001 | 0.6285 | 0.6213 | 0.6355 | <0.0001 | <0.0001 |
| Left rib 4 | Male | Left ribs | 30475 | 0.6286 | 0.6218 | 0.6354 | <0.0001 | <0.0001 | 0.6056 | 0.5985 | 0.6127 | <0.0001 | <0.0001 |
| Left rib 5 | Male | Left ribs | 34748 | 0.6039 | 0.5972 | 0.6105 | <0.0001 | <0.0001 | 0.5791 | 0.5721 | 0.586 | <0.0001 | <0.0001 |
| Left rib 6 | Male | Left ribs | 36798 | 0.5821 | 0.5753 | 0.5888 | <0.0001 | <0.0001 | 0.5577 | 0.5506 | 0.5647 | <0.0001 | <0.0001 |
| Left rib 7 | Male | Left ribs | 36949 | 0.6112 | 0.6048 | 0.6175 | <0.0001 | <0.0001 | 0.5857 | 0.5789 | 0.5923 | <0.0001 | <0.0001 |
| Left rib 8 | Male | Left ribs | 36985 | 0.6254 | 0.6191 | 0.6316 | <0.0001 | <0.0001 | 0.5996 | 0.593 | 0.6061 | <0.0001 | <0.0001 |
| Left rib 9 | Male | Left ribs | 37074 | 0.632 | 0.6259 | 0.6381 | <0.0001 | <0.0001 | 0.608 | 0.6015 | 0.6144 | <0.0001 | <0.0001 |
| Left rib 10 | Male | Left ribs | 37342 | 0.6415 | 0.6355 | 0.6475 | <0.0001 | <0.0001 | 0.6179 | 0.6116 | 0.6241 | <0.0001 | <0.0001 |
| Left rib 11 | Male | Left ribs | 37319 | 0.6345 | 0.6284 | 0.6405 | <0.0001 | <0.0001 | 0.6099 | 0.6035 | 0.6162 | <0.0001 | <0.0001 |
| Left rib 12 | Male | Left ribs | 36812 | 0.59 | 0.5833 | 0.5966 | <0.0001 | <0.0001 | 0.5595 | 0.5525 | 0.5665 | <0.0001 | <0.0001 |
| Right rib 1 | Male | Right ribs | 27468 | 0.55 | 0.5417 | 0.5582 | <0.0001 | <0.0001 | 0.5391 | 0.5306 | 0.5474 | <0.0001 | <0.0001 |
| Right rib 2 | Male | Right ribs | 27479 | 0.6489 | 0.642 | 0.6557 | <0.0001 | <0.0001 | 0.6289 | 0.6217 | 0.636 | <0.0001 | <0.0001 |
| Right rib 3 | Male | Right ribs | 27926 | 0.6463 | 0.6395 | 0.6531 | <0.0001 | <0.0001 | 0.6269 | 0.6197 | 0.6339 | <0.0001 | <0.0001 |
| Right rib 4 | Male | Right ribs | 30306 | 0.626 | 0.6191 | 0.6328 | <0.0001 | <0.0001 | 0.6024 | 0.5952 | 0.6095 | <0.0001 | <0.0001 |
| Right rib 5 | Male | Right ribs | 34525 | 0.5964 | 0.5896 | 0.6031 | <0.0001 | <0.0001 | 0.5727 | 0.5655 | 0.5797 | <0.0001 | <0.0001 |
| Right rib 6 | Male | Right ribs | 36765 | 0.5911 | 0.5844 | 0.5977 | <0.0001 | <0.0001 | 0.5666 | 0.5596 | 0.5735 | <0.0001 | <0.0001 |
| Right rib 7 | Male | Right ribs | 36940 | 0.6201 | 0.6137 | 0.6263 | <0.0001 | <0.0001 | 0.5945 | 0.5879 | 0.6011 | <0.0001 | <0.0001 |
| Right rib 8 | Male | Right ribs | 36970 | 0.631 | 0.6248 | 0.6371 | <0.0001 | <0.0001 | 0.6059 | 0.5994 | 0.6123 | <0.0001 | <0.0001 |
| Right rib 9 | Male | Right ribs | 37055 | 0.6399 | 0.6339 | 0.6459 | <0.0001 | <0.0001 | 0.6166 | 0.6103 | 0.6229 | <0.0001 | <0.0001 |
| Right rib 10 | Male | Right ribs | 37309 | 0.6415 | 0.6354 | 0.6474 | <0.0001 | <0.0001 | 0.6193 | 0.613 | 0.6255 | <0.0001 | <0.0001 |
| Right rib 11 | Male | Right ribs | 37379 | 0.6361 | 0.63 | 0.6421 | <0.0001 | <0.0001 | 0.6105 | 0.6041 | 0.6168 | <0.0001 | <0.0001 |
| Right rib 12 | Male | Right ribs | 36884 | 0.5851 | 0.5783 | 0.5918 | <0.0001 | <0.0001 | 0.5553 | 0.5482 | 0.5624 | <0.0001 | <0.0001 |

**Supplementary Table 12 (continued): Association with L1 and surrogacy, all sites (Conversion to the L1 scale)**

| **Site** | **Sex** | **Conversion to L1: intercept** | **Conversion to L1: slope** | **SD of L1 prediction error (HU)** | **Lower 95% limit of agreement (HU)** | **Upper 95% limit of agreement (HU)** |
| --- | --- | --- | --- | --- | --- | --- |
| C1 | Female | 12.7577 | 0.4899 | 35.5498 | -69.6776 | 69.6776 |
| C2 | Female | 26.3423 | 0.5038 | 31.6509 | -62.0358 | 62.0358 |
| C3 | Female | 16.9687 | 0.5355 | 32.9836 | -64.6478 | 64.6478 |
| C4 | Female | 10.8521 | 0.5213 | 33.6761 | -66.0051 | 66.0051 |
| C5 | Female | 11.3712 | 0.5366 | 33.881 | -66.4068 | 66.4068 |
| C6 | Female | -0.1312 | 0.6081 | 33.4346 | -65.5317 | 65.5317 |
| C7 | Female | -9.8159 | 0.7187 | 32.5648 | -63.827 | 63.827 |
| T1 | Female | -12.5073 | 0.8267 | 29.4192 | -57.6617 | 57.6617 |
| T2 | Female | -12.0625 | 0.8845 | 28.5909 | -56.0381 | 56.0381 |
| T3 | Female | -1.0157 | 0.8586 | 28.4413 | -55.7449 | 55.7449 |
| T4 | Female | 2.4279 | 0.8634 | 28.3241 | -55.5153 | 55.5153 |
| T5 | Female | -2.2611 | 0.8966 | 28.2663 | -55.402 | 55.402 |
| T6 | Female | -2.7683 | 0.9045 | 28.2912 | -55.4508 | 55.4508 |
| T7 | Female | -4.0889 | 0.9245 | 28.4192 | -55.7016 | 55.7016 |
| T8 | Female | -1.3714 | 0.9298 | 27.9396 | -54.7617 | 54.7617 |
| T9 | Female | -0.541 | 0.911 | 26.7635 | -52.4565 | 52.4565 |
| T10 | Female | 1.8909 | 0.9059 | 25.2756 | -49.5402 | 49.5402 |
| T11 | Female | 7.5271 | 0.9236 | 24.7111 | -48.4338 | 48.4338 |
| T12 | Female | 8.7515 | 0.9772 | 22.7118 | -44.5151 | 44.5151 |
| L2 | Female | -3.7965 | 0.961 | 22.7207 | -44.5326 | 44.5326 |
| L3 | Female | -1.5108 | 0.9373 | 25.85 | -50.666 | 50.666 |
| L4 | Female | 1.467 | 0.8998 | 29.6556 | -58.1249 | 58.1249 |
| L5 | Female | 13.4589 | 0.8464 | 32.0139 | -62.7472 | 62.7472 |
| S1 | Female | 39.9811 | 0.7302 | 30.6414 | -60.0572 | 60.0572 |
| Left rib 1 | Female | 38.6691 | 0.6162 | 38.2108 | -74.8932 | 74.8932 |
| Left rib 2 | Female | 52.9807 | 0.6829 | 34.7714 | -68.1519 | 68.1519 |
| Left rib 3 | Female | 73.0201 | 0.6069 | 34.845 | -68.2962 | 68.2962 |
| Left rib 4 | Female | 84.929 | 0.5479 | 37.0999 | -72.7158 | 72.7158 |
| Left rib 5 | Female | 98.0461 | 0.4957 | 39.5014 | -77.4228 | 77.4228 |
| Left rib 6 | Female | 98.032 | 0.4721 | 40.0682 | -78.5336 | 78.5336 |
| Left rib 7 | Female | 83.5648 | 0.4985 | 38.0865 | -74.6495 | 74.6495 |
| Left rib 8 | Female | 82.6005 | 0.4821 | 36.1129 | -70.7814 | 70.7814 |
| Left rib 9 | Female | 79.3122 | 0.4783 | 35.4943 | -69.5688 | 69.5688 |
| Left rib 10 | Female | 66.5263 | 0.4736 | 35.1006 | -68.7972 | 68.7972 |
| Left rib 11 | Female | 77.5697 | 0.4371 | 35.6103 | -69.7962 | 69.7962 |
| Left rib 12 | Female | 84.4853 | 0.4143 | 37.9289 | -74.3406 | 74.3406 |
| Right rib 1 | Female | 38.1613 | 0.6202 | 37.5254 | -73.5498 | 73.5498 |
| Right rib 2 | Female | 54.8683 | 0.6704 | 34.7527 | -68.1154 | 68.1154 |
| Right rib 3 | Female | 73.9338 | 0.6034 | 34.8868 | -68.3782 | 68.3782 |
| Right rib 4 | Female | 83.3008 | 0.5437 | 37.5732 | -73.6434 | 73.6434 |
| Right rib 5 | Female | 99.4104 | 0.4829 | 40.5131 | -79.4056 | 79.4056 |
| Right rib 6 | Female | 94.7318 | 0.476 | 39.5358 | -77.4901 | 77.4901 |
| Right rib 7 | Female | 80.7564 | 0.5016 | 37.9067 | -74.2972 | 74.2972 |
| Right rib 8 | Female | 75.5883 | 0.4987 | 36.4491 | -71.4402 | 71.4402 |
| Right rib 9 | Female | 70.5633 | 0.4991 | 35.8292 | -70.2253 | 70.2253 |
| Right rib 10 | Female | 58.1175 | 0.4886 | 35.5706 | -69.7183 | 69.7183 |
| Right rib 11 | Female | 71.9038 | 0.4485 | 35.8325 | -70.2316 | 70.2316 |
| Right rib 12 | Female | 84.1316 | 0.415 | 38.8439 | -76.134 | 76.134 |
| C1 | Male | 19.5844 | 0.4552 | 37.6514 | -73.7967 | 73.7967 |
| C2 | Male | 22.1175 | 0.5048 | 34.078 | -66.7929 | 66.7929 |
| C3 | Male | 30.1073 | 0.4849 | 35.2761 | -69.1411 | 69.1411 |
| C4 | Male | 26.1634 | 0.4704 | 36.2331 | -71.0169 | 71.0169 |
| C5 | Male | 19.4502 | 0.51 | 36.5821 | -71.7009 | 71.7009 |
| C6 | Male | 8.3677 | 0.5813 | 36.7163 | -71.964 | 71.964 |
| C7 | Male | -0.607 | 0.6787 | 35.5787 | -69.7343 | 69.7343 |
| T1 | Male | -15.0122 | 0.8403 | 32.2337 | -63.178 | 63.178 |
| T2 | Male | -12.4451 | 0.8911 | 30.8898 | -60.544 | 60.544 |
| T3 | Male | -2.1618 | 0.8555 | 30.4194 | -59.6219 | 59.6219 |
| T4 | Male | 0.2364 | 0.8558 | 30.0399 | -58.8782 | 58.8782 |
| T5 | Male | -3.1888 | 0.8793 | 30.0981 | -58.9922 | 58.9922 |
| T6 | Male | -2.946 | 0.8802 | 30.2415 | -59.2734 | 59.2734 |
| T7 | Male | -0.7652 | 0.8837 | 30.1083 | -59.0122 | 59.0122 |
| T8 | Male | 0.858 | 0.8891 | 29.8943 | -58.5928 | 58.5928 |
| T9 | Male | -0.2203 | 0.8821 | 28.7641 | -56.3776 | 56.3776 |
| T10 | Male | 0.4116 | 0.8933 | 27.6248 | -54.1446 | 54.1446 |
| T11 | Male | 3.9028 | 0.9285 | 26.7786 | -52.4861 | 52.4861 |
| T12 | Male | 4.0056 | 1.0007 | 23.2227 | -45.5166 | 45.5166 |
| L2 | Male | -1.7765 | 0.9444 | 23.2519 | -45.5738 | 45.5738 |
| L3 | Male | -2.11 | 0.921 | 27.8533 | -54.5925 | 54.5925 |
| L4 | Male | 6.7164 | 0.8536 | 31.8455 | -62.4171 | 62.4171 |
| L5 | Male | 28.7248 | 0.7569 | 34.0955 | -66.8273 | 66.8273 |
| S1 | Male | 32.2048 | 0.765 | 32.6956 | -64.0834 | 64.0834 |
| Left rib 1 | Male | 60.4105 | 0.5547 | 42.0787 | -82.4742 | 82.4742 |
| Left rib 2 | Male | 44.1845 | 0.7667 | 39.1914 | -76.8152 | 76.8152 |
| Left rib 3 | Male | 60.931 | 0.7465 | 38.8068 | -76.0613 | 76.0613 |
| Left rib 4 | Male | 73.9939 | 0.6626 | 39.5 | -77.42 | 77.42 |
| Left rib 5 | Male | 85.687 | 0.6107 | 40.3323 | -79.0514 | 79.0514 |
| Left rib 6 | Male | 90.4956 | 0.562 | 40.8781 | -80.1211 | 80.1211 |
| Left rib 7 | Male | 78.8374 | 0.5918 | 39.8281 | -78.0631 | 78.0631 |
| Left rib 8 | Male | 81.5182 | 0.5506 | 38.9996 | -76.4392 | 76.4392 |
| Left rib 9 | Male | 82.2748 | 0.5092 | 38.4398 | -75.342 | 75.342 |
| Left rib 10 | Male | 72.0078 | 0.497 | 37.9269 | -74.3368 | 74.3368 |
| Left rib 11 | Male | 72.754 | 0.4857 | 38.2161 | -74.9035 | 74.9035 |
| Left rib 12 | Male | 81.6289 | 0.4748 | 40.0351 | -78.4689 | 78.4689 |
| Right rib 1 | Male | 58.8653 | 0.56 | 41.6594 | -81.6525 | 81.6525 |
| Right rib 2 | Male | 45.271 | 0.7593 | 38.8288 | -76.1045 | 76.1045 |
| Right rib 3 | Male | 60.644 | 0.7592 | 39.0083 | -76.4562 | 76.4562 |
| Right rib 4 | Male | 71.6285 | 0.6765 | 39.6616 | -77.7367 | 77.7367 |
| Right rib 5 | Male | 81.8595 | 0.6236 | 40.7645 | -79.8984 | 79.8984 |
| Right rib 6 | Male | 87.8112 | 0.5706 | 40.5872 | -79.5508 | 79.5508 |
| Right rib 7 | Male | 75.5112 | 0.5989 | 39.494 | -77.4083 | 77.4083 |
| Right rib 8 | Male | 76.4124 | 0.5685 | 38.796 | -76.0401 | 76.0401 |
| Right rib 9 | Male | 76.0806 | 0.5315 | 38.1924 | -74.8572 | 74.8572 |
| Right rib 10 | Male | 64.7189 | 0.5143 | 38.0082 | -74.4961 | 74.4961 |
| Right rib 11 | Male | 65.8331 | 0.5003 | 38.2067 | -74.8851 | 74.8851 |
| Right rib 12 | Male | 81.1458 | 0.4709 | 40.1449 | -78.684 | 78.684 |

**Supplementary Table 12 (continued): Association with L1 and surrogacy, all sites (Discrimination of low-for-age L1)**

| **Site** | **Sex** | **AUC for low-for-age L1** | **AUC, lower 95% limit** | **AUC, upper 95% limit** | **Patients with low-for-age L1, n** | **Youden cut-off on this site's standardized score (SD)** | **Sensitivity at the cut-off** | **Specificity at the cut-off** |
| --- | --- | --- | --- | --- | --- | --- | --- | --- |
| C1 | Female | 0.8307 | 0.8198 | 0.8417 | 1311 | -0.6677 | 0.7208 | 0.7867 |
| C2 | Female | 0.8639 | 0.8544 | 0.8735 | 1333 | -0.4906 | 0.8455 | 0.7254 |
| C3 | Female | 0.8524 | 0.8426 | 0.8622 | 1400 | -0.4706 | 0.8414 | 0.7118 |
| C4 | Female | 0.8531 | 0.8436 | 0.8626 | 1495 | -0.5451 | 0.8094 | 0.7394 |
| C5 | Female | 0.8576 | 0.849 | 0.8662 | 1745 | -0.4895 | 0.8401 | 0.7224 |
| C6 | Female | 0.8681 | 0.8607 | 0.8755 | 2161 | -0.57 | 0.8269 | 0.756 |
| C7 | Female | 0.8786 | 0.8723 | 0.885 | 2475 | -0.4989 | 0.8606 | 0.7339 |
| T1 | Female | 0.9067 | 0.9014 | 0.9119 | 2550 | -0.602 | 0.8773 | 0.784 |
| T2 | Female | 0.9142 | 0.9091 | 0.9193 | 2553 | -0.672 | 0.8715 | 0.8151 |
| T3 | Female | 0.915 | 0.9099 | 0.92 | 2555 | -0.6483 | 0.8881 | 0.8061 |
| T4 | Female | 0.9167 | 0.9118 | 0.9216 | 2558 | -0.6061 | 0.8917 | 0.7907 |
| T5 | Female | 0.9187 | 0.9138 | 0.9235 | 2550 | -0.6546 | 0.8796 | 0.8079 |
| T6 | Female | 0.9195 | 0.9146 | 0.9245 | 2581 | -0.735 | 0.8543 | 0.8361 |
| T7 | Female | 0.9162 | 0.911 | 0.9214 | 2702 | -0.7019 | 0.8594 | 0.8239 |
| T8 | Female | 0.9196 | 0.9147 | 0.9246 | 2946 | -0.7057 | 0.8727 | 0.8249 |
| T9 | Female | 0.9278 | 0.9234 | 0.9321 | 3179 | -0.6858 | 0.8962 | 0.8174 |
| T10 | Female | 0.9359 | 0.932 | 0.9398 | 3384 | -0.7523 | 0.8913 | 0.8438 |
| T11 | Female | 0.9361 | 0.9318 | 0.9404 | 3431 | -0.7532 | 0.8948 | 0.8465 |
| T12 | Female | 0.9415 | 0.9365 | 0.9464 | 3451 | -0.8847 | 0.8786 | 0.897 |
| L2 | Female | 0.9521 | 0.9481 | 0.956 | 3492 | -0.8534 | 0.8952 | 0.8916 |
| L3 | Female | 0.9357 | 0.9312 | 0.9401 | 3451 | -0.8083 | 0.8713 | 0.8712 |
| L4 | Female | 0.9149 | 0.9099 | 0.92 | 3476 | -0.6874 | 0.8631 | 0.823 |
| L5 | Female | 0.8953 | 0.8897 | 0.9008 | 3481 | -0.6184 | 0.8509 | 0.7881 |
| S1 | Female | 0.9004 | 0.8955 | 0.9052 | 3462 | -0.6011 | 0.8616 | 0.7853 |
| Left rib 1 | Female | 0.7781 | 0.7686 | 0.7876 | 2397 | -0.5002 | 0.7 | 0.7184 |
| Left rib 2 | Female | 0.8298 | 0.8219 | 0.8377 | 2423 | -0.4265 | 0.8196 | 0.6965 |
| Left rib 3 | Female | 0.8141 | 0.8059 | 0.8223 | 2554 | -0.5146 | 0.7404 | 0.7335 |
| Left rib 4 | Female | 0.7895 | 0.7814 | 0.7977 | 2892 | -0.4937 | 0.712 | 0.7292 |
| Left rib 5 | Female | 0.7629 | 0.7551 | 0.7706 | 3290 | -0.1855 | 0.8082 | 0.6007 |
| Left rib 6 | Female | 0.7574 | 0.7499 | 0.765 | 3412 | -0.2304 | 0.7775 | 0.6184 |
| Left rib 7 | Female | 0.7785 | 0.7709 | 0.7861 | 3399 | -0.405 | 0.7337 | 0.6804 |
| Left rib 8 | Female | 0.7953 | 0.7879 | 0.8027 | 3391 | -0.4025 | 0.7656 | 0.6803 |
| Left rib 9 | Female | 0.8049 | 0.7976 | 0.8122 | 3370 | -0.5063 | 0.7401 | 0.7188 |
| Left rib 10 | Female | 0.8076 | 0.8003 | 0.8148 | 3383 | -0.3721 | 0.7928 | 0.6676 |
| Left rib 11 | Female | 0.7894 | 0.7817 | 0.7971 | 3380 | -0.4368 | 0.7456 | 0.6879 |
| Left rib 12 | Female | 0.761 | 0.7523 | 0.7696 | 3253 | -0.5344 | 0.6732 | 0.7272 |
| Right rib 1 | Female | 0.7854 | 0.7762 | 0.7946 | 2417 | -0.3809 | 0.7621 | 0.6674 |
| Right rib 2 | Female | 0.8292 | 0.8212 | 0.8371 | 2435 | -0.3938 | 0.8209 | 0.6838 |
| Right rib 3 | Female | 0.8128 | 0.8044 | 0.8212 | 2545 | -0.5016 | 0.7501 | 0.7276 |
| Right rib 4 | Female | 0.7835 | 0.7753 | 0.7917 | 2879 | -0.3928 | 0.7447 | 0.6861 |
| Right rib 5 | Female | 0.7532 | 0.7455 | 0.761 | 3261 | -0.265 | 0.7599 | 0.6416 |
| Right rib 6 | Female | 0.7675 | 0.7602 | 0.7749 | 3415 | -0.2814 | 0.7716 | 0.6414 |
| Right rib 7 | Female | 0.7812 | 0.7737 | 0.7887 | 3374 | -0.3435 | 0.7635 | 0.659 |
| Right rib 8 | Female | 0.7957 | 0.7883 | 0.8031 | 3379 | -0.4837 | 0.7254 | 0.7125 |
| Right rib 9 | Female | 0.8077 | 0.8005 | 0.8149 | 3380 | -0.4667 | 0.7562 | 0.7046 |
| Right rib 10 | Female | 0.8063 | 0.799 | 0.8136 | 3375 | -0.4022 | 0.7763 | 0.6796 |
| Right rib 11 | Female | 0.7886 | 0.7808 | 0.7964 | 3394 | -0.4214 | 0.7454 | 0.6842 |
| Right rib 12 | Female | 0.7572 | 0.7485 | 0.7659 | 3328 | -0.4925 | 0.6833 | 0.7049 |
| C1 | Male | 0.8396 | 0.829 | 0.8502 | 1310 | -0.5245 | 0.7885 | 0.7401 |
| C2 | Male | 0.8807 | 0.8724 | 0.889 | 1324 | -0.5975 | 0.8308 | 0.7719 |
| C3 | Male | 0.8719 | 0.8633 | 0.8806 | 1403 | -0.65 | 0.8004 | 0.7901 |
| C4 | Male | 0.8633 | 0.8548 | 0.8718 | 1579 | -0.5328 | 0.8201 | 0.7409 |
| C5 | Male | 0.8625 | 0.8545 | 0.8705 | 1910 | -0.6564 | 0.777 | 0.79 |
| C6 | Male | 0.8691 | 0.8624 | 0.8758 | 2421 | -0.5697 | 0.8265 | 0.7531 |
| C7 | Male | 0.8851 | 0.8793 | 0.8908 | 2765 | -0.5999 | 0.8409 | 0.7722 |
| T1 | Male | 0.9141 | 0.9095 | 0.9187 | 2813 | -0.644 | 0.8834 | 0.7994 |
| T2 | Male | 0.921 | 0.9165 | 0.9256 | 2824 | -0.7199 | 0.8718 | 0.8312 |
| T3 | Male | 0.9223 | 0.9179 | 0.9267 | 2828 | -0.6949 | 0.8815 | 0.8206 |
| T4 | Male | 0.9257 | 0.9215 | 0.93 | 2839 | -0.6587 | 0.9007 | 0.8062 |
| T5 | Male | 0.9283 | 0.9243 | 0.9324 | 2846 | -0.666 | 0.9009 | 0.8128 |
| T6 | Male | 0.9283 | 0.9242 | 0.9324 | 2868 | -0.6919 | 0.893 | 0.8192 |
| T7 | Male | 0.9271 | 0.9229 | 0.9312 | 2995 | -0.6771 | 0.8972 | 0.8129 |
| T8 | Male | 0.9289 | 0.9247 | 0.933 | 3233 | -0.732 | 0.8859 | 0.8348 |
| T9 | Male | 0.9379 | 0.9344 | 0.9415 | 3472 | -0.7457 | 0.9032 | 0.8412 |
| T10 | Male | 0.9429 | 0.9395 | 0.9462 | 3679 | -0.7839 | 0.9021 | 0.8549 |
| T11 | Male | 0.944 | 0.9406 | 0.9475 | 3765 | -0.7787 | 0.906 | 0.8569 |
| T12 | Male | 0.9546 | 0.951 | 0.9582 | 3780 | -0.8415 | 0.918 | 0.883 |
| L2 | Male | 0.9587 | 0.9557 | 0.9617 | 3818 | -0.8662 | 0.9078 | 0.8957 |
| L3 | Male | 0.9406 | 0.9368 | 0.9444 | 3797 | -0.8096 | 0.8891 | 0.8723 |
| L4 | Male | 0.9177 | 0.913 | 0.9224 | 3800 | -0.7249 | 0.8642 | 0.8358 |
| L5 | Male | 0.9033 | 0.8984 | 0.9082 | 3785 | -0.6309 | 0.8624 | 0.789 |
| S1 | Male | 0.9169 | 0.9127 | 0.921 | 3729 | -0.6653 | 0.885 | 0.8114 |
| Left rib 1 | Male | 0.7844 | 0.7756 | 0.7932 | 2650 | -0.4981 | 0.7245 | 0.7024 |
| Left rib 2 | Male | 0.8473 | 0.8403 | 0.8543 | 2676 | -0.5538 | 0.7986 | 0.7416 |
| Left rib 3 | Male | 0.8466 | 0.8398 | 0.8535 | 2769 | -0.4918 | 0.8183 | 0.7208 |
| Left rib 4 | Male | 0.8347 | 0.8279 | 0.8416 | 3084 | -0.5592 | 0.7643 | 0.7498 |
| Left rib 5 | Male | 0.8238 | 0.8173 | 0.8302 | 3490 | -0.4287 | 0.8032 | 0.696 |
| Left rib 6 | Male | 0.8116 | 0.8051 | 0.818 | 3642 | -0.4151 | 0.7935 | 0.6893 |
| Left rib 7 | Male | 0.83 | 0.8238 | 0.8362 | 3674 | -0.5304 | 0.7771 | 0.7307 |
| Left rib 8 | Male | 0.8358 | 0.8296 | 0.842 | 3673 | -0.5403 | 0.7819 | 0.7365 |
| Left rib 9 | Male | 0.8426 | 0.8365 | 0.8487 | 3648 | -0.4876 | 0.8188 | 0.7093 |
| Left rib 10 | Male | 0.8453 | 0.8392 | 0.8514 | 3648 | -0.5377 | 0.8013 | 0.7316 |
| Left rib 11 | Male | 0.8422 | 0.8361 | 0.8482 | 3693 | -0.5682 | 0.7812 | 0.7444 |
| Left rib 12 | Male | 0.7996 | 0.7923 | 0.8068 | 3664 | -0.5329 | 0.7325 | 0.7215 |
| Right rib 1 | Male | 0.7921 | 0.7835 | 0.8008 | 2645 | -0.4625 | 0.7543 | 0.6898 |
| Right rib 2 | Male | 0.847 | 0.84 | 0.8539 | 2681 | -0.477 | 0.8232 | 0.7088 |
| Right rib 3 | Male | 0.8455 | 0.8386 | 0.8524 | 2767 | -0.4932 | 0.8106 | 0.7208 |
| Right rib 4 | Male | 0.8362 | 0.8295 | 0.843 | 3075 | -0.5389 | 0.7821 | 0.74 |
| Right rib 5 | Male | 0.8196 | 0.8131 | 0.8261 | 3466 | -0.4453 | 0.7931 | 0.704 |
| Right rib 6 | Male | 0.8159 | 0.8095 | 0.8223 | 3671 | -0.3928 | 0.8039 | 0.6798 |
| Right rib 7 | Male | 0.8306 | 0.8245 | 0.8368 | 3677 | -0.5349 | 0.7729 | 0.7354 |
| Right rib 8 | Male | 0.8373 | 0.8312 | 0.8435 | 3659 | -0.4964 | 0.8005 | 0.7187 |
| Right rib 9 | Male | 0.8456 | 0.8394 | 0.8517 | 3639 | -0.5586 | 0.7983 | 0.7396 |
| Right rib 10 | Male | 0.8467 | 0.8406 | 0.8528 | 3651 | -0.5129 | 0.8118 | 0.7229 |
| Right rib 11 | Male | 0.8389 | 0.8327 | 0.8451 | 3695 | -0.5598 | 0.7821 | 0.7406 |
| Right rib 12 | Male | 0.7991 | 0.7918 | 0.8064 | 3673 | -0.5548 | 0.7234 | 0.7308 |

Low-for-age L1 is an L1 value below the 10th reference centile for the patient's age and sex. Discrimination uses each site's own age- and sex-standardized score, defined within each sex as the residual of that site's attenuation from a cubic spline in age divided by the SD of those residuals; the Youden cut-off is on that same score, in SD units, and a patient is classified as low when the score falls at or below it. Conversion is by Deming (errors-in-variables) regression with an assumed error-variance ratio of 1. p values are controlled across candidate sites within each sex by the Benjamini–Hochberg procedure.

**Supplementary Table 13: Agreement between left and right ribs (Association)**

| **Rib level** | **Sex** | **n** | **Mean, left (HU)** | **Mean, right (HU)** | **Correlation** | **Correlation, lower 95% limit** | **Correlation, upper 95% limit** | **ICC(A,1)** | **ICC, lower 95% limit** | **ICC, upper 95% limit** |
| --- | --- | --- | --- | --- | --- | --- | --- | --- | --- | --- |
| 1 | Male | 27870 | 297.49 | 297.629 | 0.771 | 0.766 | 0.776 | 0.771 | 0.764 | 0.777 |
| 1 | Female | 24851 | 288.536 | 287.526 | 0.881 | 0.878 | 0.883 | 0.88 | 0.877 | 0.884 |
| 2 | Male | 28028 | 237.45 | 238.434 | 0.938 | 0.937 | 0.94 | 0.938 | 0.936 | 0.94 |
| 2 | Female | 24989 | 241.461 | 243.205 | 0.963 | 0.962 | 0.964 | 0.963 | 0.961 | 0.964 |
| 3 | Male | 28799 | 221.172 | 217.938 | 0.955 | 0.954 | 0.956 | 0.954 | 0.952 | 0.955 |
| 3 | Female | 26042 | 237.183 | 237.191 | 0.965 | 0.964 | 0.965 | 0.965 | 0.963 | 0.966 |
| 4 | Male | 31658 | 229.636 | 228.448 | 0.938 | 0.936 | 0.939 | 0.937 | 0.935 | 0.939 |
| 4 | Female | 29063 | 241.787 | 246.543 | 0.95 | 0.949 | 0.951 | 0.948 | 0.946 | 0.95 |
| 5 | Male | 35979 | 230.873 | 231.998 | 0.926 | 0.924 | 0.927 | 0.925 | 0.924 | 0.927 |
| 5 | Female | 33174 | 242.737 | 246.057 | 0.938 | 0.937 | 0.939 | 0.937 | 0.935 | 0.94 |
| 6 | Male | 38192 | 242.374 | 243.354 | 0.93 | 0.929 | 0.931 | 0.93 | 0.928 | 0.932 |
| 6 | Female | 34976 | 255.786 | 260.613 | 0.947 | 0.946 | 0.948 | 0.945 | 0.944 | 0.947 |
| 7 | Male | 38343 | 249.913 | 252.523 | 0.933 | 0.931 | 0.934 | 0.932 | 0.93 | 0.934 |
| 7 | Female | 35054 | 271.69 | 275.645 | 0.946 | 0.945 | 0.947 | 0.945 | 0.944 | 0.946 |
| 8 | Male | 38420 | 263.483 | 264.254 | 0.938 | 0.936 | 0.939 | 0.937 | 0.936 | 0.939 |
| 8 | Female | 35061 | 282.712 | 287.399 | 0.949 | 0.947 | 0.95 | 0.947 | 0.945 | 0.948 |
| 9 | Male | 38434 | 282.988 | 282.794 | 0.928 | 0.927 | 0.93 | 0.928 | 0.926 | 0.93 |
| 9 | Female | 35123 | 291.276 | 296.712 | 0.944 | 0.943 | 0.945 | 0.941 | 0.939 | 0.943 |
| 10 | Male | 38228 | 309.849 | 313.648 | 0.914 | 0.912 | 0.916 | 0.913 | 0.911 | 0.915 |
| 10 | Female | 34924 | 320.788 | 328.157 | 0.933 | 0.931 | 0.934 | 0.929 | 0.927 | 0.931 |
| 11 | Male | 38077 | 315.816 | 320.248 | 0.898 | 0.897 | 0.9 | 0.897 | 0.894 | 0.899 |
| 11 | Female | 34711 | 322.857 | 327.095 | 0.92 | 0.918 | 0.921 | 0.918 | 0.916 | 0.92 |
| 12 | Male | 37365 | 304.959 | 308.584 | 0.817 | 0.814 | 0.821 | 0.817 | 0.812 | 0.821 |
| 12 | Female | 33409 | 324.638 | 324.825 | 0.837 | 0.834 | 0.84 | 0.837 | 0.833 | 0.841 |

**Supplementary Table 13 (continued): Agreement between left and right ribs (Agreement)**

| **Rib level** | **Sex** | **Mean difference, left − right (HU)** | **SD of the difference (HU)** | **Lower 95% limit of agreement (HU)** | **Upper 95% limit of agreement (HU)** |
| --- | --- | --- | --- | --- | --- |
| 1 | Male | -0.138 | 45.217 | -88.765 | 88.488 |
| 1 | Female | 1.01 | 34.091 | -65.809 | 67.829 |
| 2 | Male | -0.984 | 20.446 | -41.058 | 39.09 |
| 2 | Female | -1.744 | 18.595 | -38.19 | 34.702 |
| 3 | Male | 3.235 | 17.758 | -31.572 | 38.041 |
| 3 | Female | -0.007 | 19.885 | -38.982 | 38.967 |
| 4 | Male | 1.188 | 22.729 | -43.36 | 45.736 |
| 4 | Female | -4.757 | 25.325 | -54.394 | 44.881 |
| 5 | Male | -1.125 | 25.746 | -51.587 | 49.337 |
| 5 | Female | -3.32 | 29.435 | -61.013 | 54.373 |
| 6 | Male | -0.98 | 26.044 | -52.027 | 50.067 |
| 6 | Female | -4.827 | 28.071 | -59.846 | 50.192 |
| 7 | Male | -2.61 | 25.208 | -52.018 | 46.798 |
| 7 | Female | -3.955 | 27.958 | -58.753 | 50.842 |
| 8 | Male | -0.771 | 25.497 | -50.744 | 49.203 |
| 8 | Female | -4.686 | 28.58 | -60.703 | 51.33 |
| 9 | Male | 0.194 | 28.731 | -56.119 | 56.508 |
| 9 | Female | -5.436 | 30.079 | -64.391 | 53.519 |
| 10 | Male | -3.799 | 31.968 | -66.456 | 58.859 |
| 10 | Female | -7.369 | 33.094 | -72.234 | 57.497 |
| 11 | Male | -4.432 | 35.358 | -73.733 | 64.869 |
| 11 | Female | -4.238 | 38.659 | -80.01 | 71.535 |
| 12 | Male | -3.625 | 47.485 | -96.695 | 89.445 |
| 12 | Female | -0.187 | 55.616 | -109.195 | 108.821 |

ICC(A,1) is a two-way random-effects, absolute-agreement, single-measurement intraclass correlation with a bootstrap interval. Limits of agreement are Bland–Altman limits. A high correlation across a wide population distribution is compatible with substantial disagreement in an individual.

**Supplementary Table 14: Between-center spread of attenuation by skeletal site**

| **Site** | **Region** | **Range across centers (HU)** | **SD across centers (HU)** | **Lowest center deviation (HU)** | **Highest center deviation (HU)** |
| --- | --- | --- | --- | --- | --- |
| C1 | Cervical vertebrae | 36.2 | 11.17 | -14.7 | 21.5 |
| C2 | Cervical vertebrae | 24.4 | 9.4 | -9.3 | 15.1 |
| C3 | Cervical vertebrae | 25.7 | 8.55 | -8.3 | 17.4 |
| C4 | Cervical vertebrae | 37.9 | 11.99 | -13.3 | 24.6 |
| C5 | Cervical vertebrae | 37.7 | 11.84 | -13.6 | 24.1 |
| C6 | Cervical vertebrae | 35.8 | 11.12 | -16 | 19.8 |
| C7 | Cervical vertebrae | 30.6 | 9.84 | -14.2 | 16.4 |
| T1 | Thoracic vertebrae | 22.6 | 6.95 | -11.4 | 11.2 |
| T2 | Thoracic vertebrae | 17.3 | 4.69 | -8.6 | 8.7 |
| T3 | Thoracic vertebrae | 15.8 | 4.34 | -6.6 | 9.2 |
| T4 | Thoracic vertebrae | 15 | 4.1 | -6.3 | 8.7 |
| T5 | Thoracic vertebrae | 15 | 4.26 | -7.2 | 7.8 |
| T6 | Thoracic vertebrae | 15.1 | 4.17 | -7.2 | 7.9 |
| T7 | Thoracic vertebrae | 15.4 | 4.07 | -8 | 7.4 |
| T8 | Thoracic vertebrae | 15.7 | 4.3 | -8.8 | 6.9 |
| T9 | Thoracic vertebrae | 16.7 | 4.38 | -8.7 | 8 |
| T10 | Thoracic vertebrae | 17.5 | 4.5 | -8.8 | 8.7 |
| T11 | Thoracic vertebrae | 16.4 | 4.24 | -8 | 8.4 |
| T12 | Thoracic vertebrae | 14.3 | 3.85 | -7.7 | 6.6 |
| L1 | Lumbar vertebrae | 13.3 | 3.91 | -7 | 6.3 |
| L2 | Lumbar vertebrae | 14.8 | 4.38 | -7.5 | 7.3 |
| L3 | Lumbar vertebrae | 14.8 | 4.67 | -7 | 7.8 |
| L4 | Lumbar vertebrae | 13.8 | 4.66 | -5.8 | 8 |
| L5 | Lumbar vertebrae | 14.8 | 4.79 | -6.8 | 8 |
| S1 | Sacrum | 15.2 | 4.12 | -8.1 | 7.1 |
| Left rib 1 | Left ribs | 21.2 | 7.01 | -9.5 | 11.7 |
| Left rib 2 | Left ribs | 21 | 6.44 | -9.1 | 11.9 |
| Left rib 3 | Left ribs | 20.8 | 6.41 | -8.7 | 12.1 |
| Left rib 4 | Left ribs | 21.1 | 7.42 | -8.1 | 13 |
| Left rib 5 | Left ribs | 26.3 | 7.82 | -11.8 | 14.5 |
| Left rib 6 | Left ribs | 31.5 | 8.79 | -12.3 | 19.2 |
| Left rib 7 | Left ribs | 29.1 | 8.73 | -8.1 | 21 |
| Left rib 8 | Left ribs | 30.1 | 8.4 | -9.7 | 20.4 |
| Left rib 9 | Left ribs | 34.9 | 9.37 | -12.1 | 22.8 |
| Left rib 10 | Left ribs | 34.2 | 9.43 | -11.5 | 22.7 |
| Left rib 11 | Left ribs | 31.1 | 8.31 | -13.3 | 17.8 |
| Left rib 12 | Left ribs | 25.5 | 7.68 | -11.4 | 14.1 |
| Right rib 1 | Right ribs | 20 | 6.98 | -8.7 | 11.3 |
| Right rib 2 | Right ribs | 20.2 | 6.26 | -8.5 | 11.7 |
| Right rib 3 | Right ribs | 20.1 | 6.13 | -8.9 | 11.2 |
| Right rib 4 | Right ribs | 21.6 | 7.31 | -9.1 | 12.5 |
| Right rib 5 | Right ribs | 31.2 | 8.76 | -15 | 16.2 |
| Right rib 6 | Right ribs | 31.9 | 8.76 | -13.5 | 18.4 |
| Right rib 7 | Right ribs | 28 | 8.36 | -7.8 | 20.2 |
| Right rib 8 | Right ribs | 32.9 | 8.81 | -11.2 | 21.7 |
| Right rib 9 | Right ribs | 36.5 | 9.48 | -12.7 | 23.8 |
| Right rib 10 | Right ribs | 39 | 10.16 | -14.1 | 24.9 |
| Right rib 11 | Right ribs | 36.2 | 9.58 | -15.6 | 20.6 |
| Right rib 12 | Right ribs | 31.2 | 8.78 | -14.1 | 17.1 |

Summarizes the age- and sex-adjusted center deviations of Supplementary Table 8. A small range means the site's attenuation is reproducible across centers despite differences in scanner and reconstruction; a large range means it is not, and that values measured at one center should not be read against another center's reference without recalibration.
